# Migration of households from New York City and the Second Peak in Covid-19 cases in New Jersey, Connecticut and New York Counties

**DOI:** 10.1101/2021.03.29.21254583

**Authors:** Adam Schulman, Gyan Bhanot

## Abstract

The five boroughs of New York City (NYC) were early epicenters of the Covid-19 pandemic in the United States, with over 380,000 cases by May 31. High caseloads were also seen in nearby counties in New Jersey (NJ), Connecticut (CT) and New York (NY). The pandemic started in the area in March with an exponential rise in the number of daily cases, peaked in early April, briefly declined, and then, showed clear signs of a second peak in several counties. We will show that despite control measures such as lockdown and restriction of movement during the exponential rise in daily cases, there was a significant net migration of households from NYC boroughs to the neighboring counties in NJ, CT and NY State. We propose that the second peak in daily cases in these counties around NYC was due, in part, to the movement of people from NYC boroughs to these counties. We estimate the movement of people using “Change of Address” (CoA) data from the US Postal Service, provided under the “Freedom of Information Act” of 1967. To identify the timing of the second peak and the number of cases in it, we use a previously proposed SIR model, which accurately describes the early stages of the coronavirus pandemic in European countries. Subtracting the model fits from the data identified, we establish the timing and the number of cases, N_CS_, in the second peak. We then related the number of cases in the second peak to the county population density, P, and the excess Change of Address, E_CoA,_ into each county using the simple model 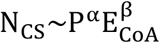 which fits the data very well with α = 0.68, *β* = 0.31 (R^2^ = 0.74, p = 1.3e-8). We also find that the time between the first and second peaks was proportional to the distance of the county seat from NY Penn Station, suggesting that this migration of households and disease was a directed flow and not a diffusion process. Our analysis provides a simple method to use change of address data to track the spread of an infectious agent, such as SARS-Cov-2, due to migrations away from epicenters during the initial stages of a pandemic.

## INTRODUCTION

Coronaviruses are large, enveloped, single-stranded RNA viruses. Although widespread in animals, they usually cause mild respiratory illnesses in humans [1–5]. In 2003, a new coronavirus emerged, which was named SARS-CoV (Severe Acute Respiratory Syndrome – Corona Virus) and which caused a life-threatening respiratory disease in humans, with a fatality rate of almost 10% [6,7]. Unfortunately, because its impact was felt only in a few countries and was quickly brought under control, the initial burst of interest in the development of treatment options and vaccines quickly waned. In late 2019 a second coronavirus, SARS-CoV-2, appeared in Wuhan, China, and has since caused a worldwide pandemic [8–13]. SARS-CoV-2 is the seventh known coronavirus to cause pathology in humans [1]. The associated respiratory illness, called COVID-19, ranges in severity from a symptomless infection [8], to common-cold like symptoms, to viral pneumonia, organ failure, neurological complications and death [9–11]. While the mortality in SARS-CoV-2 infections is significantly lower than in SARS-CoV [9–12], it has more favorable transmission characteristics, a higher reproduction number [13] and a long incubation period of 4-6 days when the patient may be asymptomatic but infective [14].

A large amount of consistent worldwide public data at varying granularity is available for the number of tests performed, confirmed cases, and deaths in different contexts, such as locations, comorbidities and complications [15,16]. These data provide important sources of information for the development and testing of models that can identify pandemic characteristics affecting viral dynamics, which can help guide policy by predicting the impact of various interventions [17]. It is well known that the count of confirmed cases seriously underestimates the actual number of infections [18,19]. This is because not everyone who is infected is symptomatic, tested and identified, and not everyone who dies from the disease is identified [20]. Even the number of reported deaths may be underestimated because of co-mortalities; i.e. COVID-19 increases susceptibility to other diseases and conditions [21]. Moreover, the virus can be transmitted by asymptomatic individuals – who comprise a substantial portion of the infected population [22] – militating against accurate estimates of transmission probabilities. Nonetheless, models can provide useful information [23].

The World Health Organization has identified contact tracing of infected individuals, followed by quarantining their contacts, as a key method to limit the spread of Covid-19 [24]. In places such as Singapore, South Korea, Thailand and China, where it has been effectively implemented, it has had an impressive impact [25,26]. However, for a variety of reasons, contact tracing was not implemented effectively everywhere [27,28]. In the United States, although strongly advocated by the CDC [29], contract tracing has been limited [30,31].

In this paper, we focus on a novel method of contact tracing which is available in the US to monitor Covid-19 infections, namely “Change of Addresses” or CoA data, which can be obtained from the United States Postal Service (USPS) under the 1967 “Freedom of Information Act”. We focus on CoA between New York City (NYC) boroughs of the Bronx, New York (Manhattan), Kings (Brooklyn), Queens and Richmond (Staten Island) and the surrounding counties in the tri-state area of New Jersey (NJ), Connecticut (CT) and New York (NY) State. Our working hypothesis is that the excess movement of households from NYC boroughs to these surrounding counties from March-June 2020 contributed to a second wave of Covid-19 infections in these counties, particularly in counties with large excess CoA and high population densities.

The first case of Covid-19 in the United States was confirmed on January 21, 2020 and the virus quickly spread to the tri-state area [32]. The first confirmed cases were registered in NY on March 1, in NJ on March 4 and in CT on March 8. Each of the tri-states responded by declaring a state of emergency (NY and NJ on March 7 and CT on March 10) and issuing a joint mandate that banned crowds of over 50 people, closed casinos, movie theatres and gyms, limited access to bars and restaurants to takeout service and set a night curfew. This was followed by each state ending in-person schooling, closing all non-essential businesses and issuing a “stay-at-home” order in an attempt at full-scale quarantine. All three states stayed in these lockdown conditions until a phased reopening began with Phase 1 (NY on June 8, NJ on June 10, CT on May 20) followed by several additional phases extending through June and July 2020 [33–35].

In spite of these mandates, we will show that from March 2020 onwards, there was a significant movement of households between NYC boroughs and the surrounding counties in NJ, CT and NY in the tri-state metropolitan area (map in Supplementary Figure 1). We find that that the county population densities and the CoA excess from NYC boroughs to these counties correlates strongly with the number of cases in the second peak in the counties. Finally, the timing between the first and second peak in cases in the counties was proportional to the distance of the county seat from Penn Station, NY by car, suggesting that the movement of disease was a directed process, related to the movement of households from NYC to these counties, and not diffusion.

## METHODS

### Data Sources

#### Change of Addresses (CoA) Data

Postal data was obtained from the United States Postal Service (USPS) under the Freedom of Information Act (FOIA) using the portal at pfiapal.usps.com. We requested information on “change of address for people moving between New York City and New Jersey/New York State/Connecticut between January 01, 2019 until the present day.” The request was received on January 4, 2021 and labeled “No. 2021-FPRO-00700”. It was routed to the National Customer Support Center on January 05, 2021 and fulfilled on January 08, 2021. The data provided by the USPS included monthly Change of Addresses (CoA) counts for households moving between NYC boroughs and counties in NJ, CT and NY State in both directions (Supplementary Table Ia and Ib). Monthly total CoA under 10 households in any county was excluded by USPS to avoid potential identification of specific households, which would violate USPS privacy policies.

#### Covid-19 Case data

A map of the tri-state area of New York (NY), New Jersey(NJ) and Connecticut (CT) (Supplementary Figure 1) shows that the boroughs of New York City (NYC) are surrounded by the counties of NJ and CT and nine counties in NY: Suffolk, Nassau, Westchester, Ulster, Dutchess, Orange, Sullivan, Rockland and Putnam. Data on Covid-19 Cases for the US was obtained from https://github.com/CSSEGISandData/COVID-19/tree/master/csse_covid_19_data/csse_covid_19_time_series and restricted to NJ Counties, CT Counties, NYC Boroughs of the Bronx, Kings (Brooklyn), New York City, Queens and Richmond and the nine NY Counties neighboring NYC named above (Supplementary Table IIa, IIb and IIc).

#### County Population and County Size

Population data for 2019 for each County in NJ, CT and NY was obtained from:

https://www.census.gov/data/tables/time-series/demo/popest/2010s-counties-total.html. Data on county size (land area) was obtained from the following:

NJ: https://www.indexmundi.com/facts/united-states/quick-facts/new-jersey/land-area#table

CT: https://www.indexmundi.com/facts/united-states/quick-facts/connecticut/land-area#table

NY: https://www.indexmundi.com/facts/united-states/quick-facts/new-york/land-area#table

Although not explicitly used in this paper, 2019 population data and land area for NYC Boroughs is available from: https://en.wikipedia.org/wiki/Boroughs_of_New_York_City.

## RESULTS

### Excess CoA from NYC Boroughs into NJ/CT/NY Counties

Figures 1a,b show the number of Changes of Addresses (CoA) for each month in 2019 and 2020 in both directions between all New York City (NYC) boroughs and all counties in New Jersey (NJ). Figures 1c,d and Figures 1e,f shows the same results for all counties in Connecticut (CT) and the nine counties in NY State named above. In each plot, the solid horizontal lines represent the average CoA for the twelve months in 2019 and the dashed horizontal lines represent one sigma limits for this average. Figure 1 clearly shows that there was a significant excess of households moving from NYC boroughs to surrounding counties in NJ/CT/NY from March 2020 onwards, compared to the same period in 2019.

**Figure 1:**
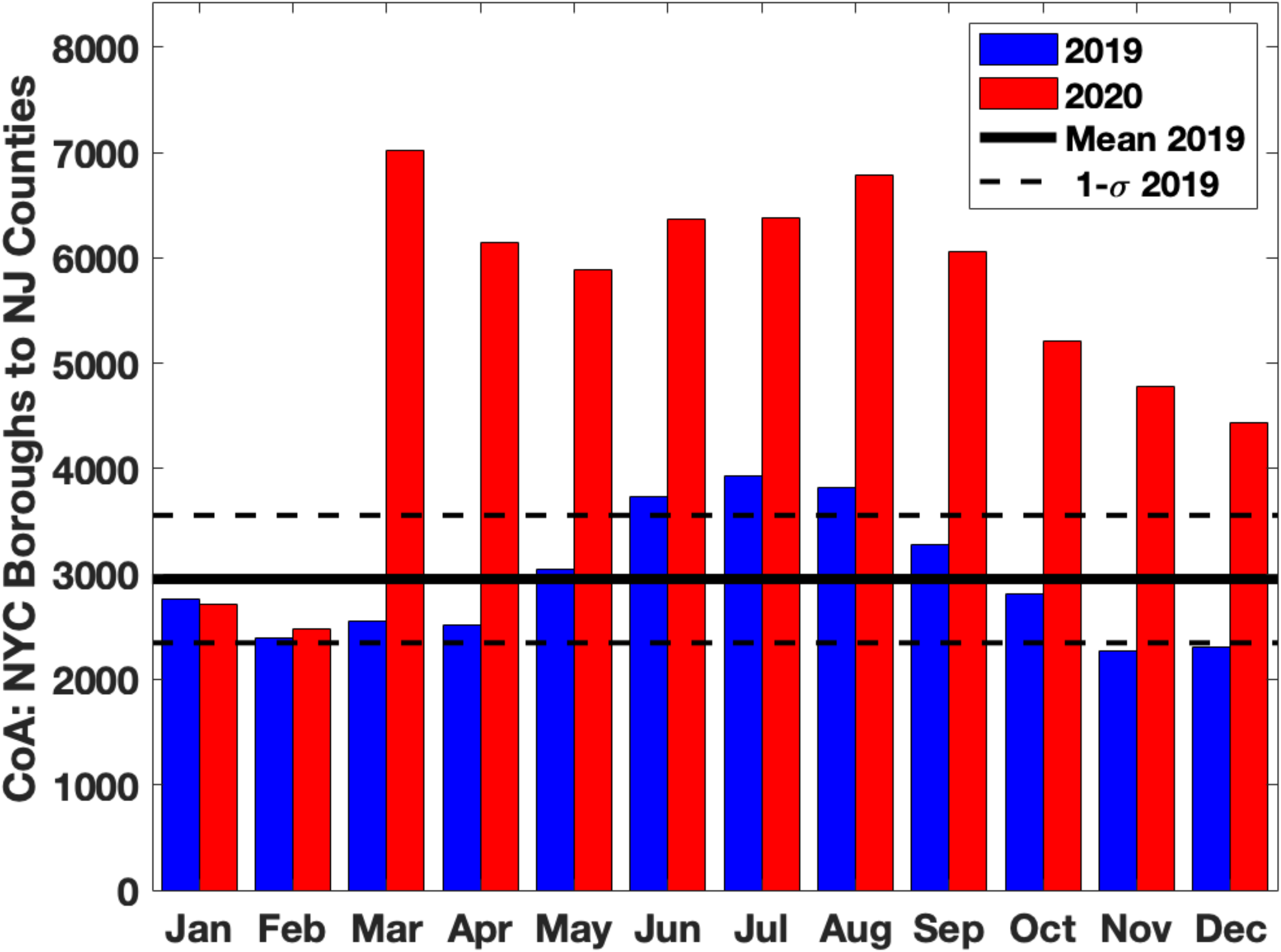
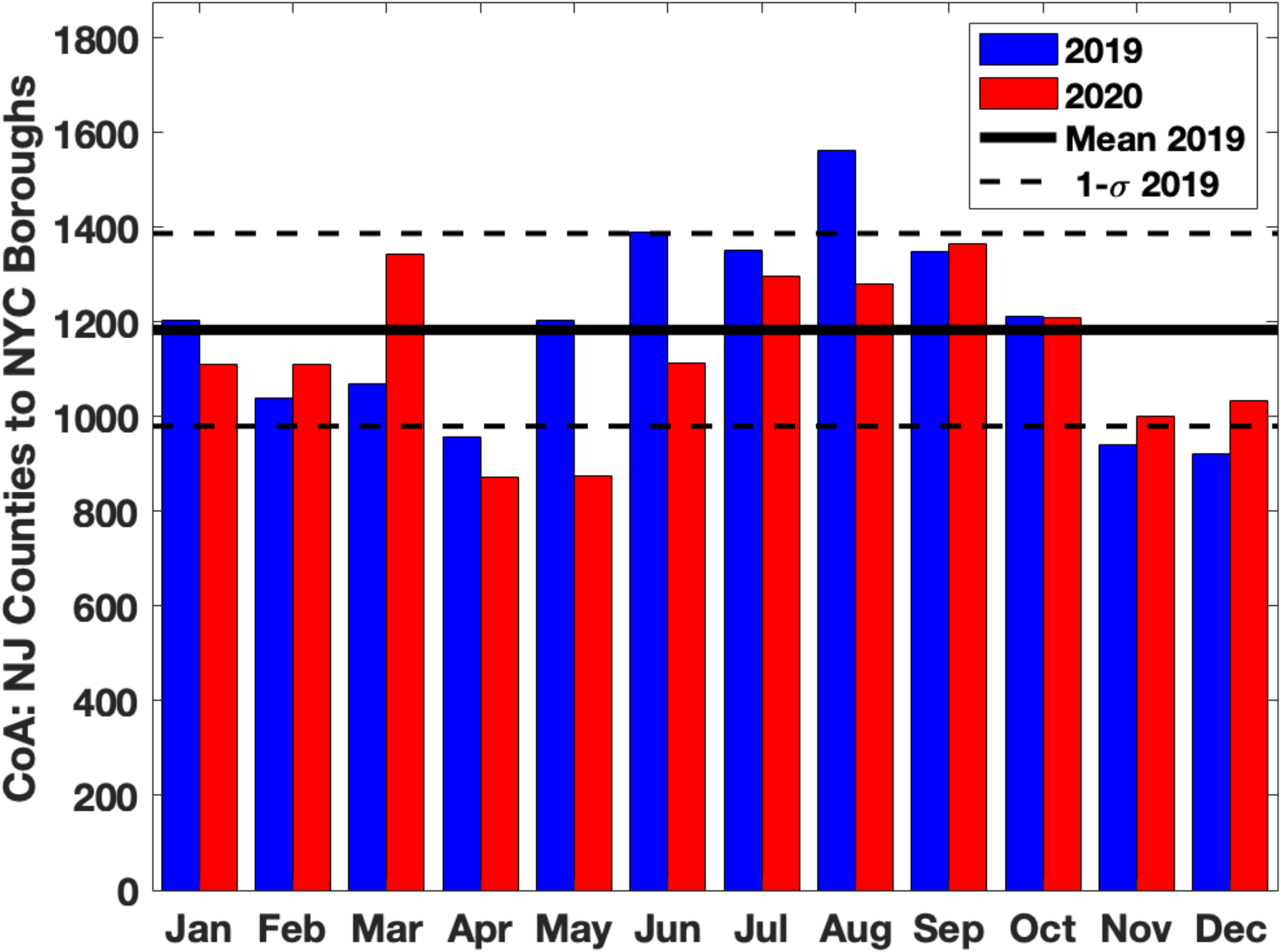
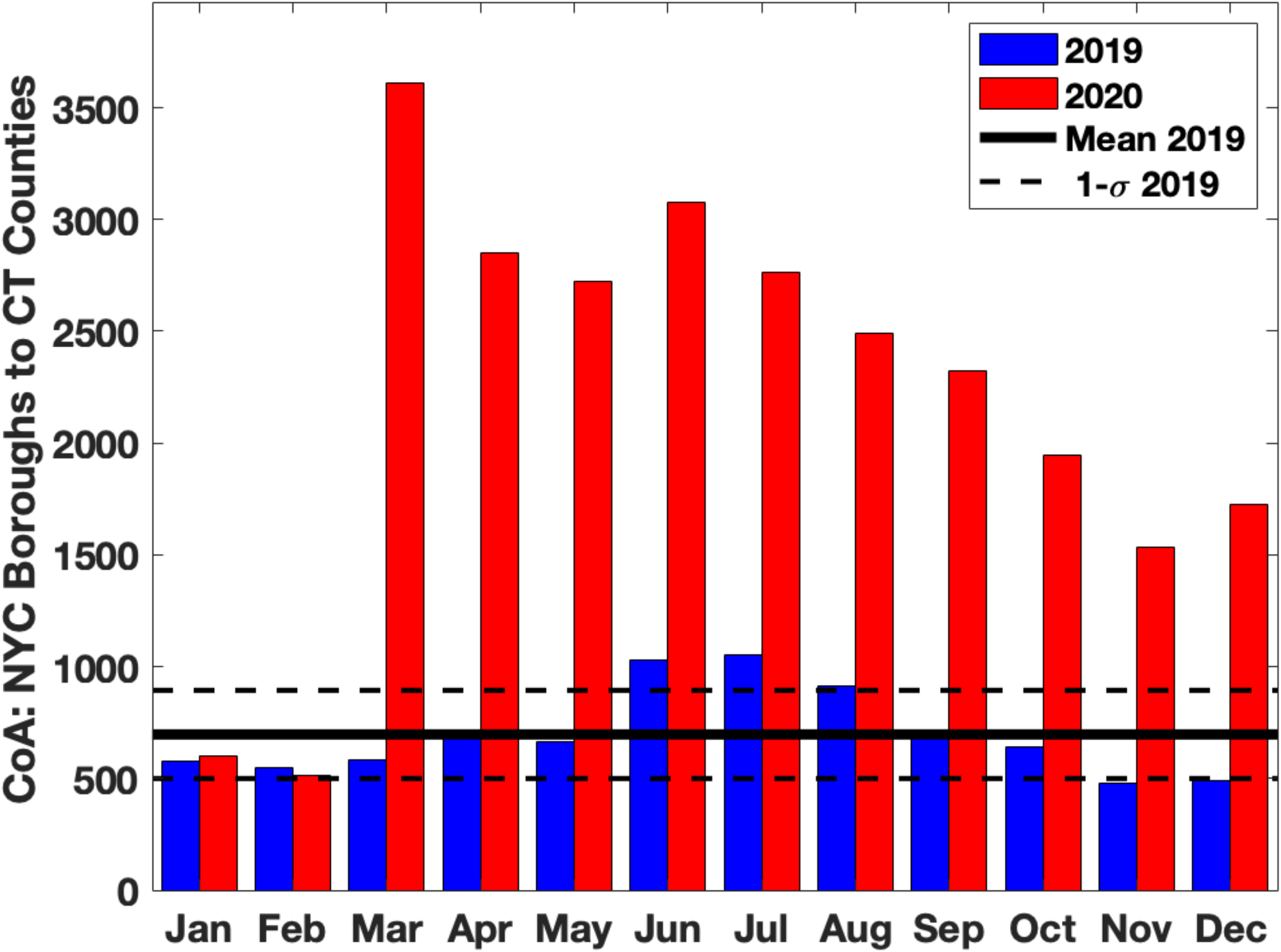
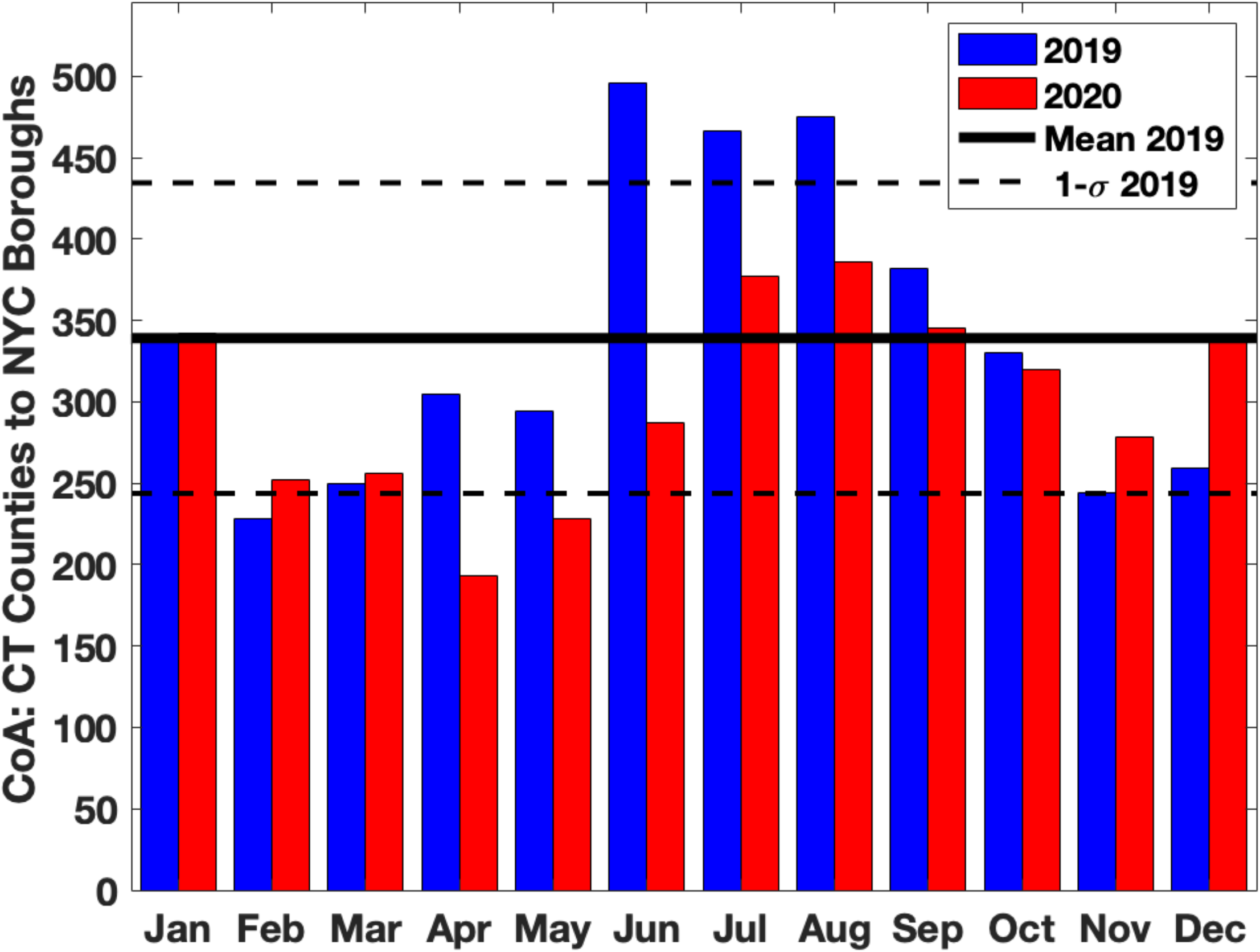
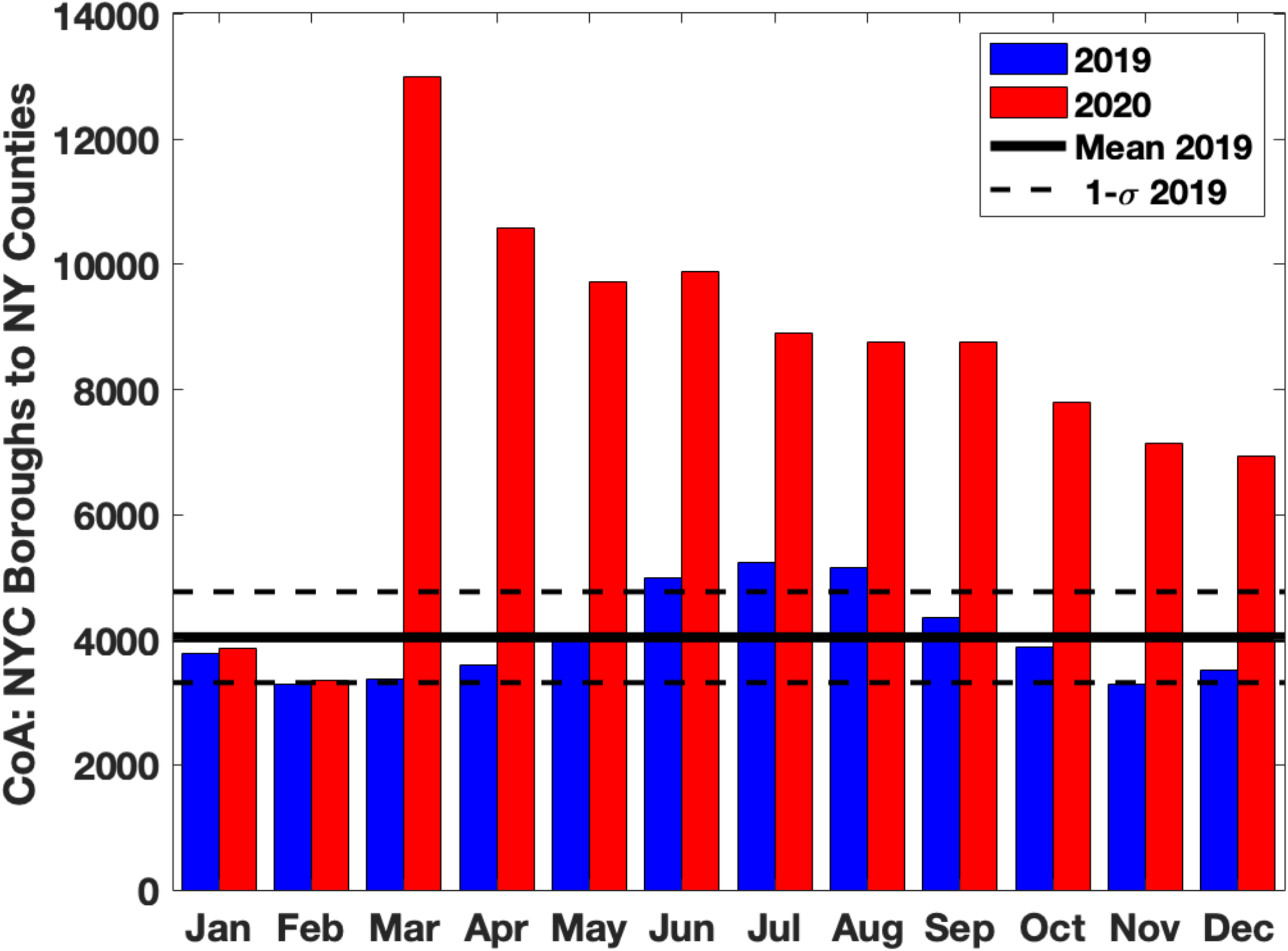
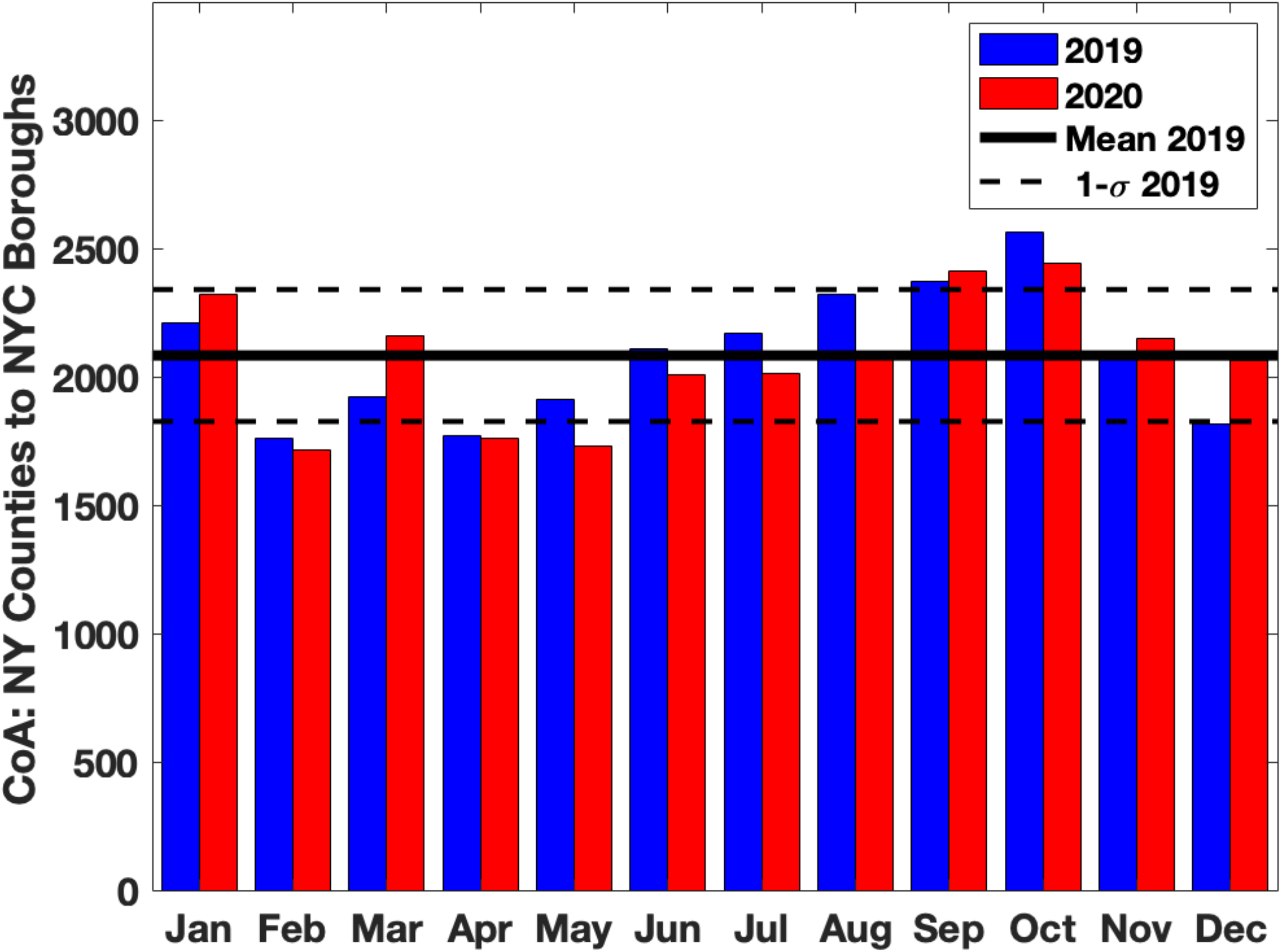
Monthly Change of Address (CoA) counts between New York City (NYC) boroughs and counties in New Jersey (NJ), Connecticut (CT) and nine New York (NY) counties near NYC in 2019 (blue bars) and 2020 (red bars). **Figures 1a, c, e** show movement from NYC boroughs to NJ, CT, NY counties respectively. **Figures 1b, d, f** show movement from NJ, CT, NY counties to NYC boroughs respectively. There is a clear excess movement of households from NYC boroughs to NJ, CT, NY counties in March-December of 2020 compared to the same months in 2019.

Figure 2a shows the CoA from NYC boroughs into each county in NJ from March-June in 2019 (blue) and 2020 (red). Figure 2b shows the CoA in the reverse direction, from each county in NJ to NYC boroughs. Similarly, Figure 2c and 2d show the CoA between NYC boroughs and each county in CT in both directions and Figure 2e and 2f shows CoA for the nine counties surrounding NYC in NY State. Finally, Figures 2g and 2h show the total CoA between each NYC borough and the counties in NJ, CT and NY. It is again clear from Figure 2a-h that there was a large excess CoA from NYC boroughs into NJ/CT/NY counties from March onwards 2020 compared to the same period in 2019.

**Figure 2:**
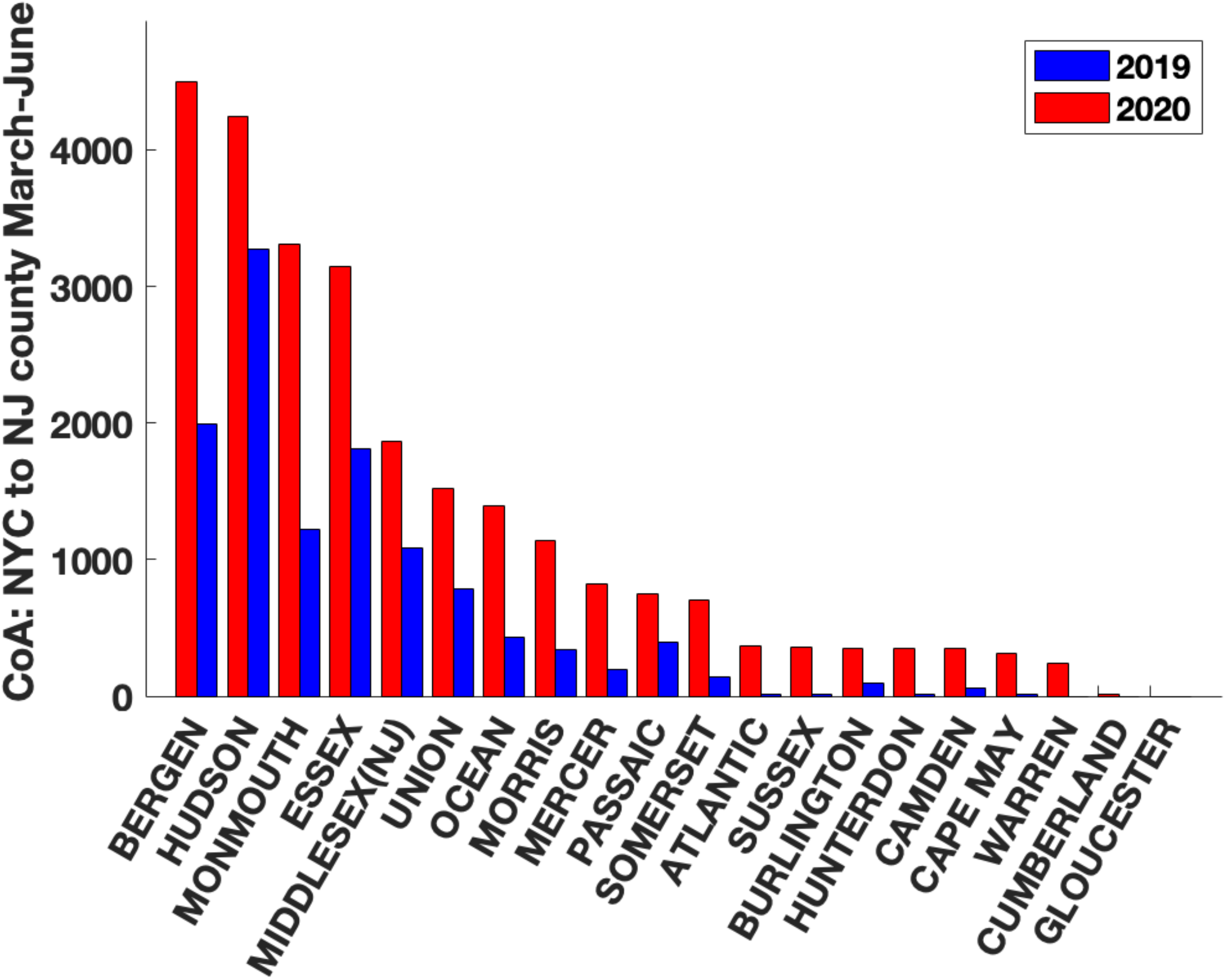
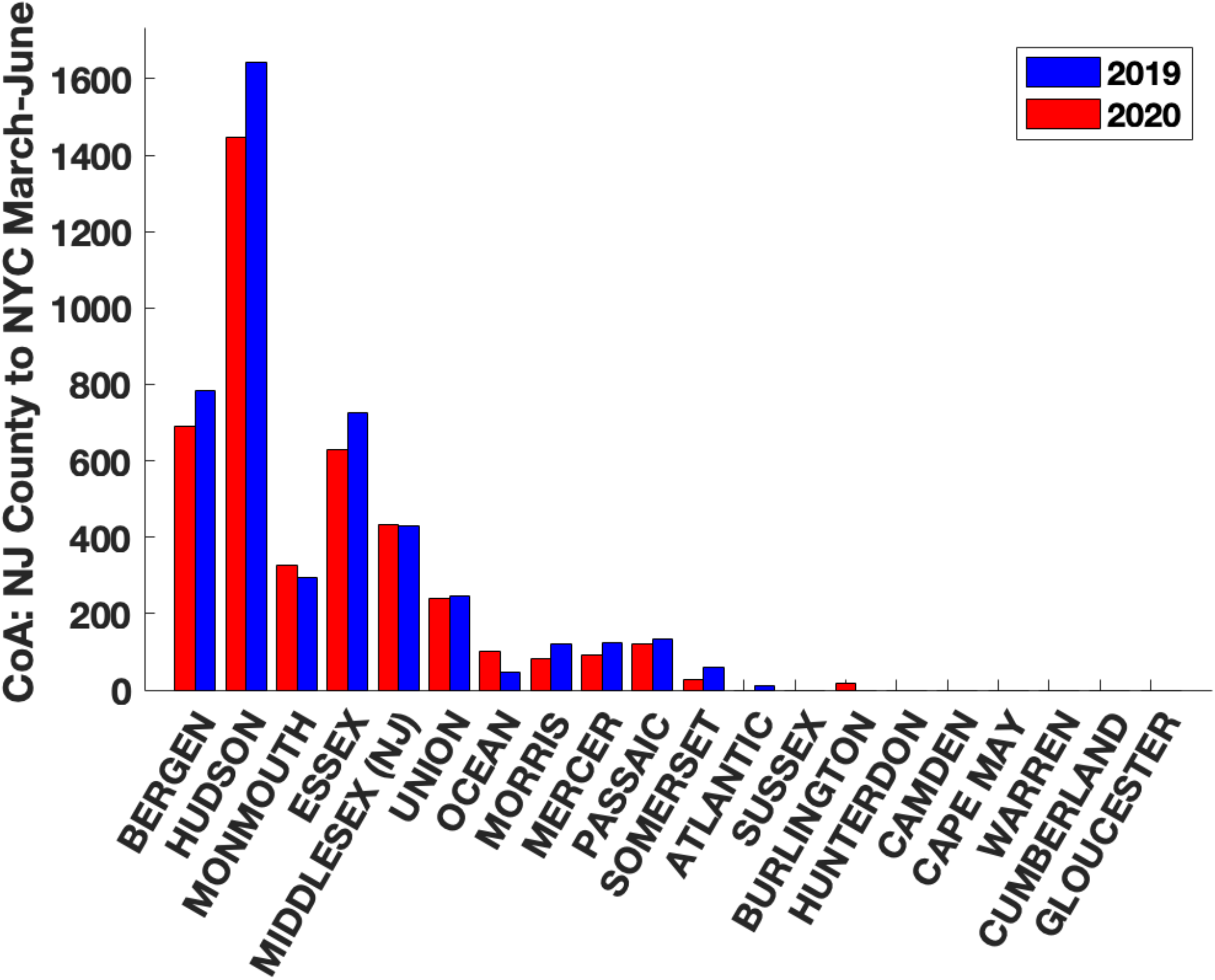
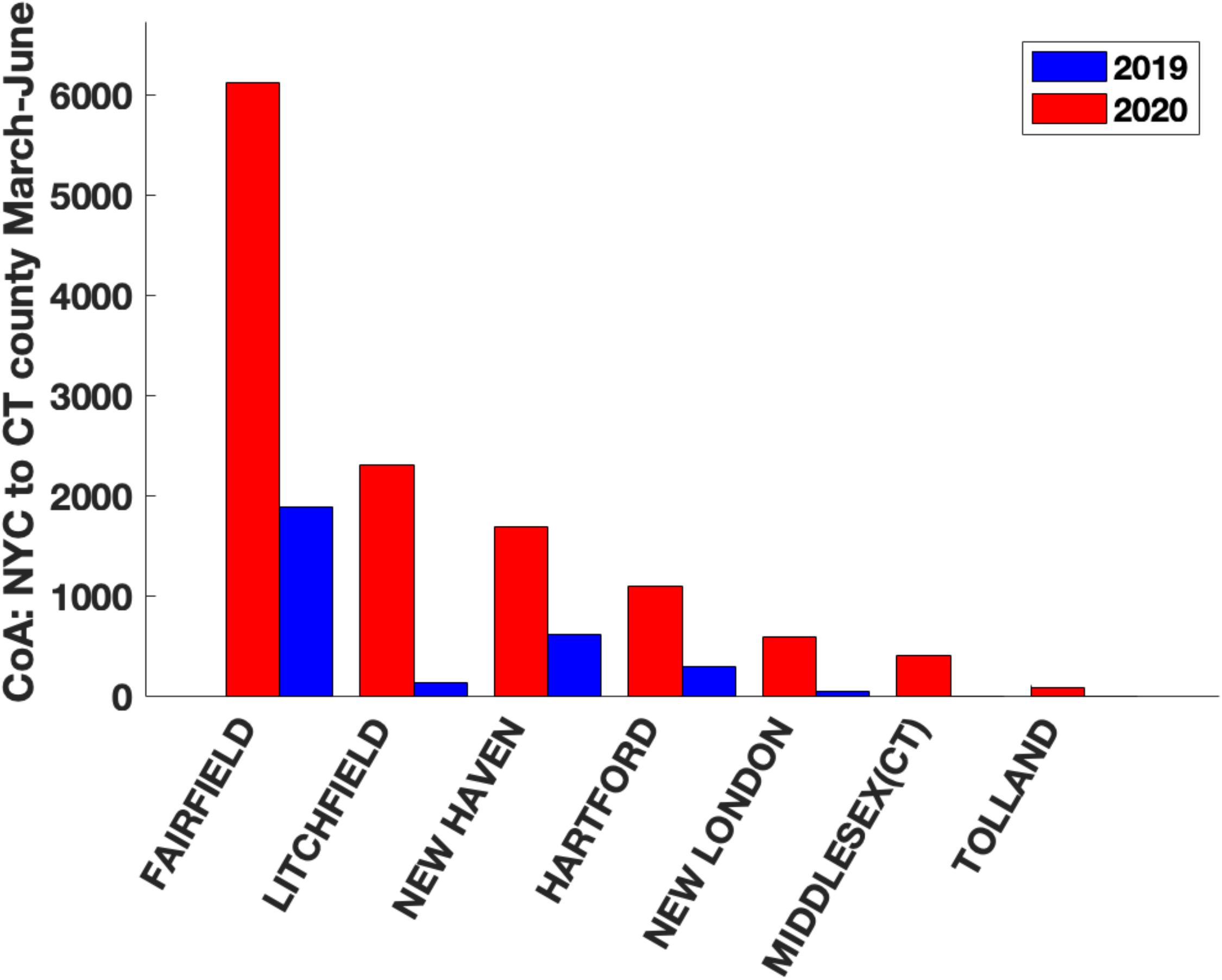
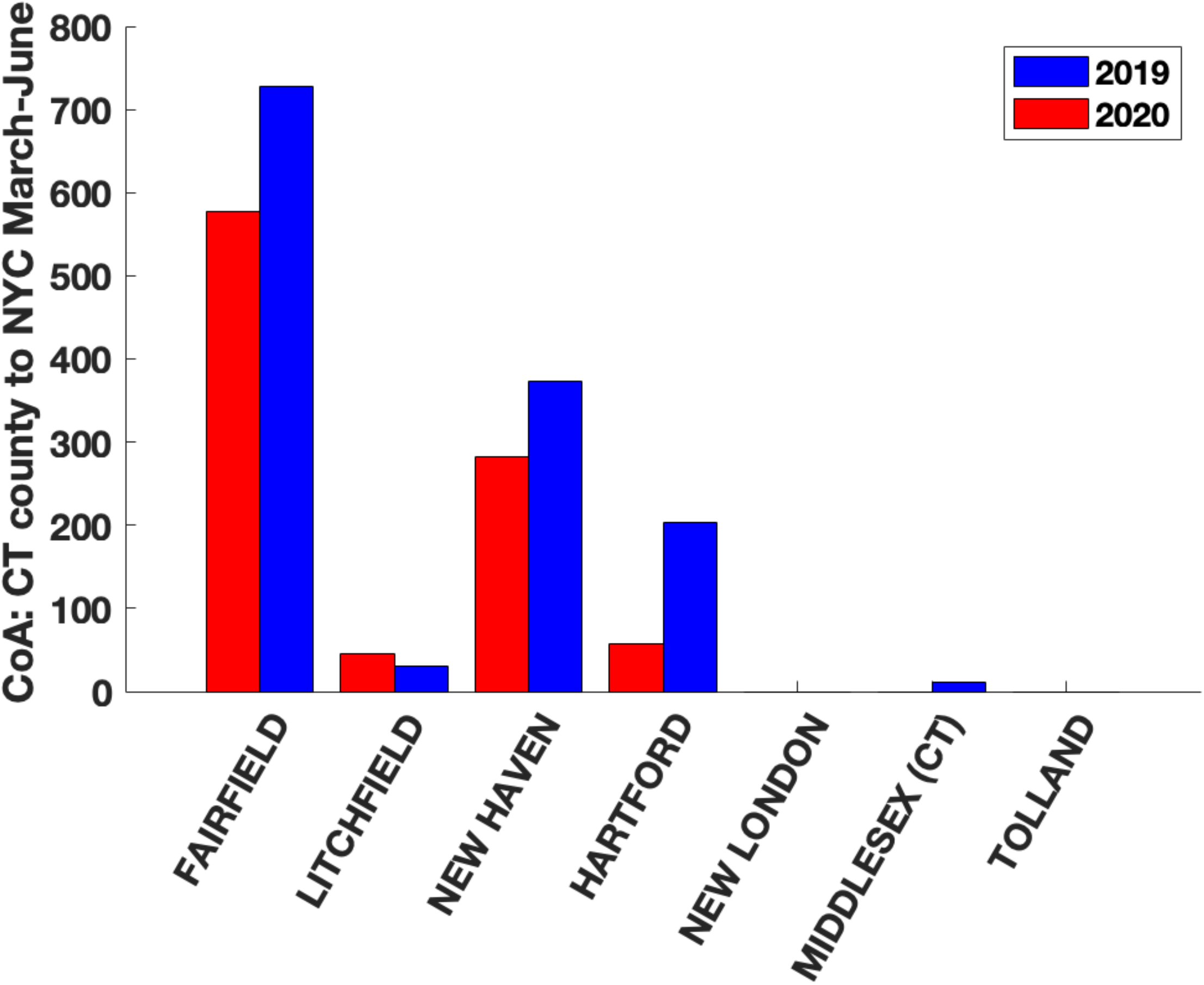
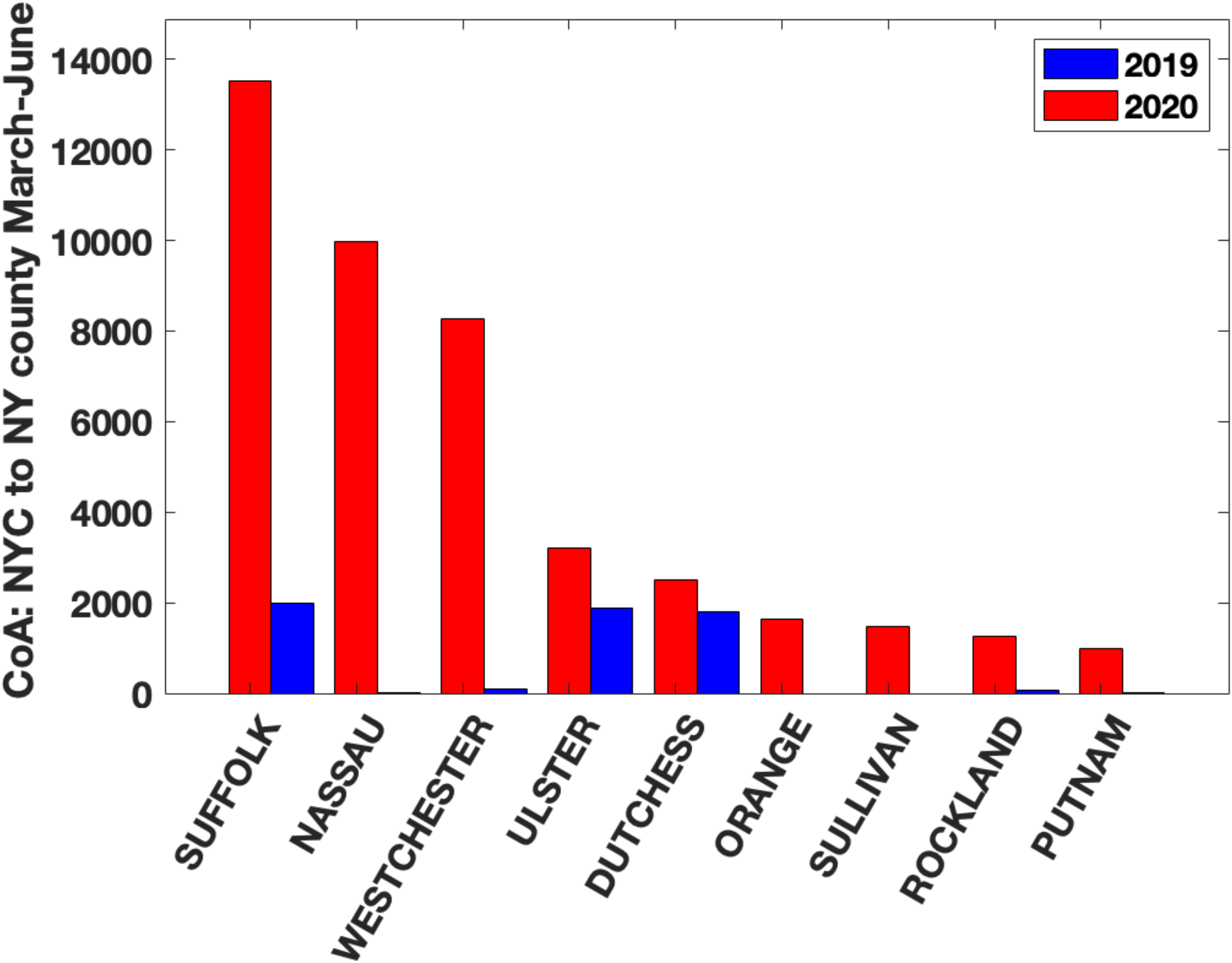
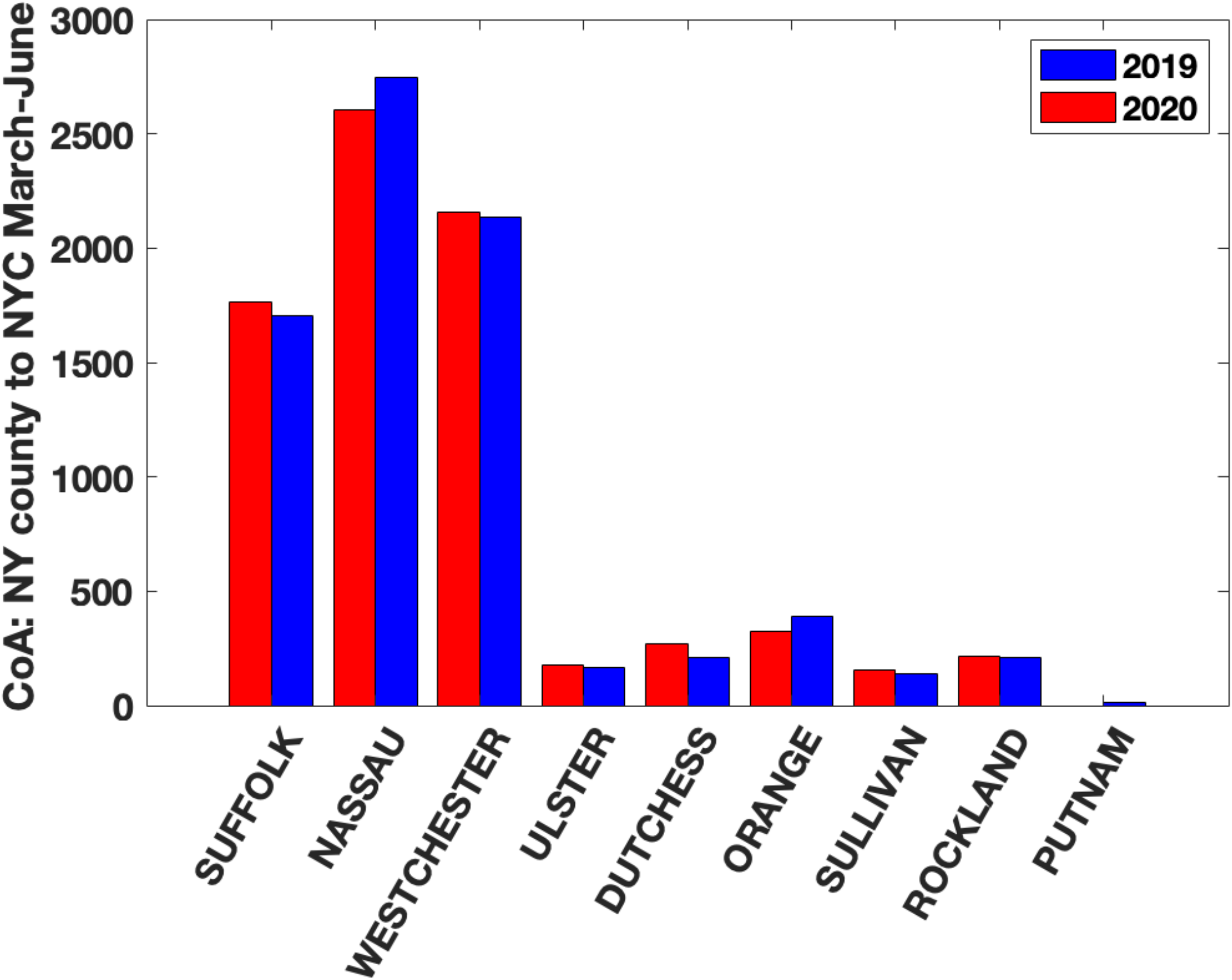
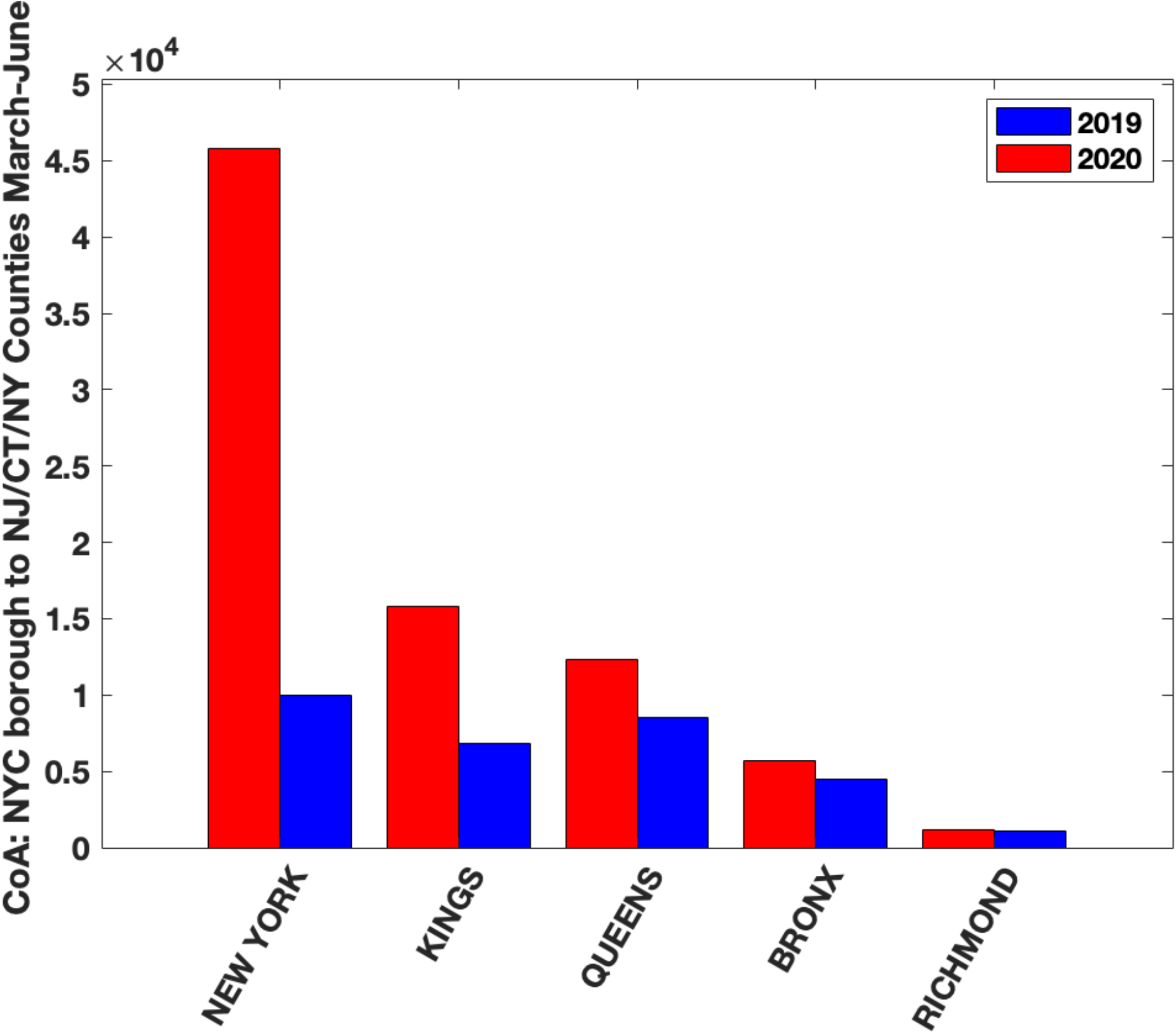
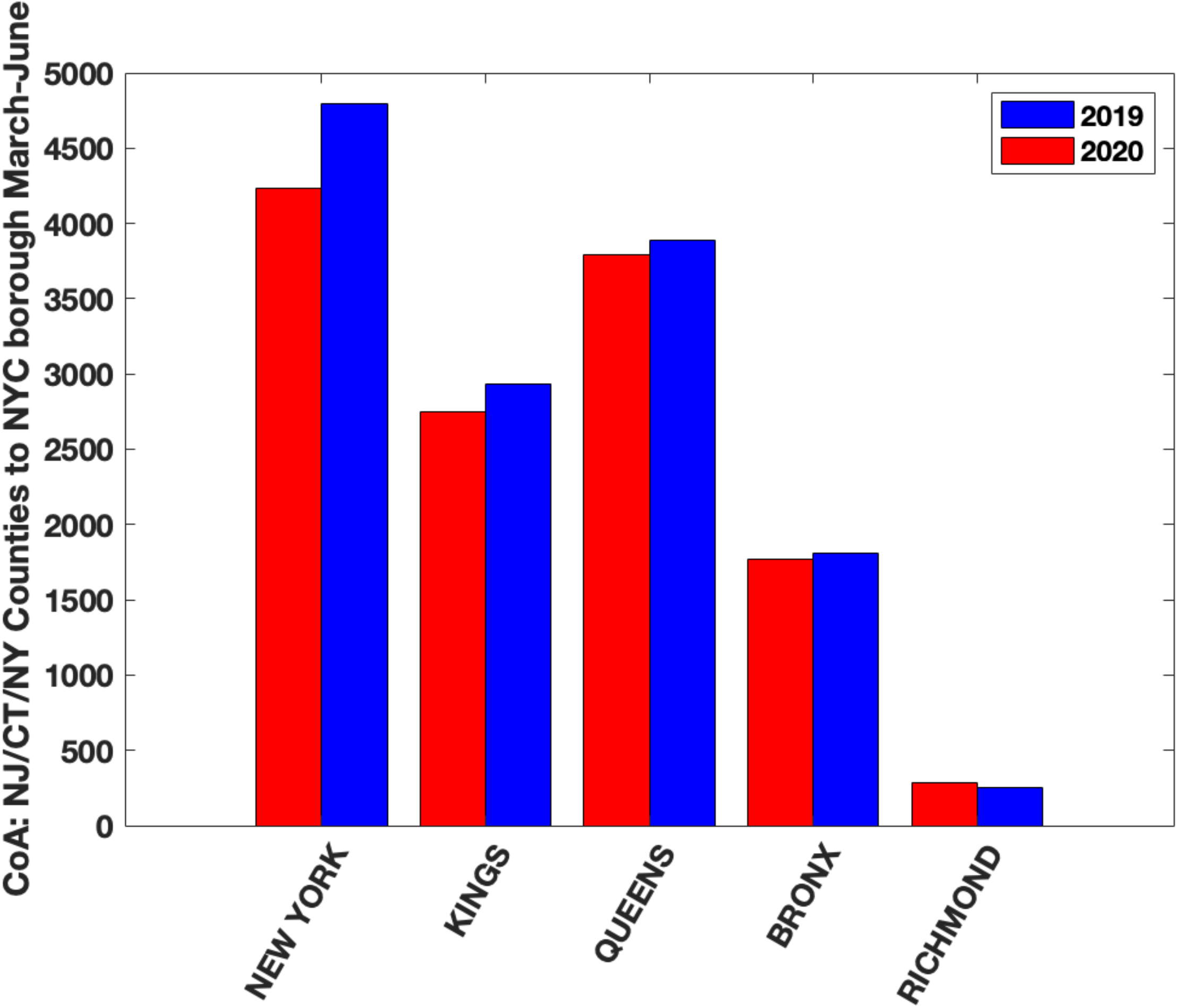
Cumulative change of address (CoA) counts between NYC boroughs and individual counties in NJ, CT and NY state from March-June in 2019 (blue bars) and 2020 (red bars). **Figures 2a, c, e** show household moves from NYC boroughs to the given county and **Figures 2b, d, f** show household moves from the given county to NYC boroughs. We see a clear excess CoA from NYC boroughs to many counties in March-December of 2020 in comparison to 2019. We note that the counties with the highest excess CoA are also the ones with the highest population densities (see Results). **Figure 2g** show household moves from NYC borough to these counties and **Figure 2h** shows household moves from the counties to each NYC borough. We note that the highest number of household moves were from New York Borough (Manhattan) to the counties.

Our hypothesis is that the movement of households from NYC boroughs to these counties was partly responsible for the second wave in Covid-19 cases in these counties. The effect of infective Covid-19 individuals moving into a county would be higher in counties with high population densities and high CoA into the county. The six NJ counties with the highest population densities are Hudson (14,557 people per square mile), Essex (5,409 people per square mile), Union (4,001 people per square mile), Bergen (4001 people per square mile), Passaic (2,718 people per square mile) and Middlesex (NJ) (2671 people per square mile); the three CT counties with the highest population densities are Fairfield (1,510 people per square mile), New Haven (1,414 people per square mile) and Hartford (1,213 people per square mile); and the six nearby counties in NY State with the highest population density are Nassau (8157 people per square mile), Westchester (2247 people per square mile), Rockland (1877 people per square mile), Suffolk (1619 people per square mile), Orange (474 people per square mile) and Putnam (426 people per square mile). Figure 3a-o show the results for CoA from NYC boroughs to each of these counties. Results for all CoA between NYC boroughs and NJ, CT, NY counties are in Supplementary Figures 2, 3, 4 and 5. It is clear from these that compared to the same months in 2019, from March-June 2020 and beyond, there was a significant excess in CoA *from* NYC boroughs to NJ, CT and NY counties.

**Figure 3:**
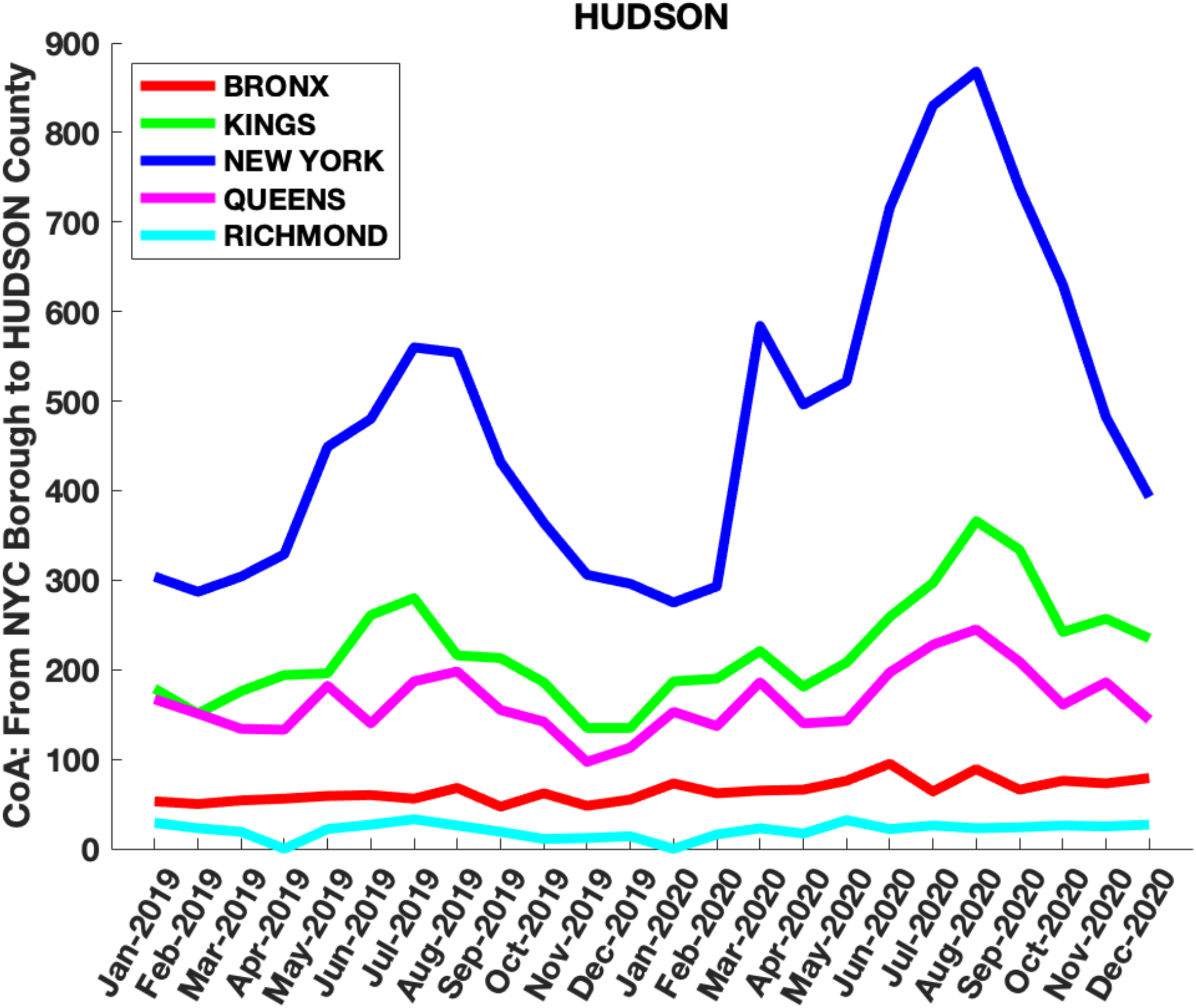
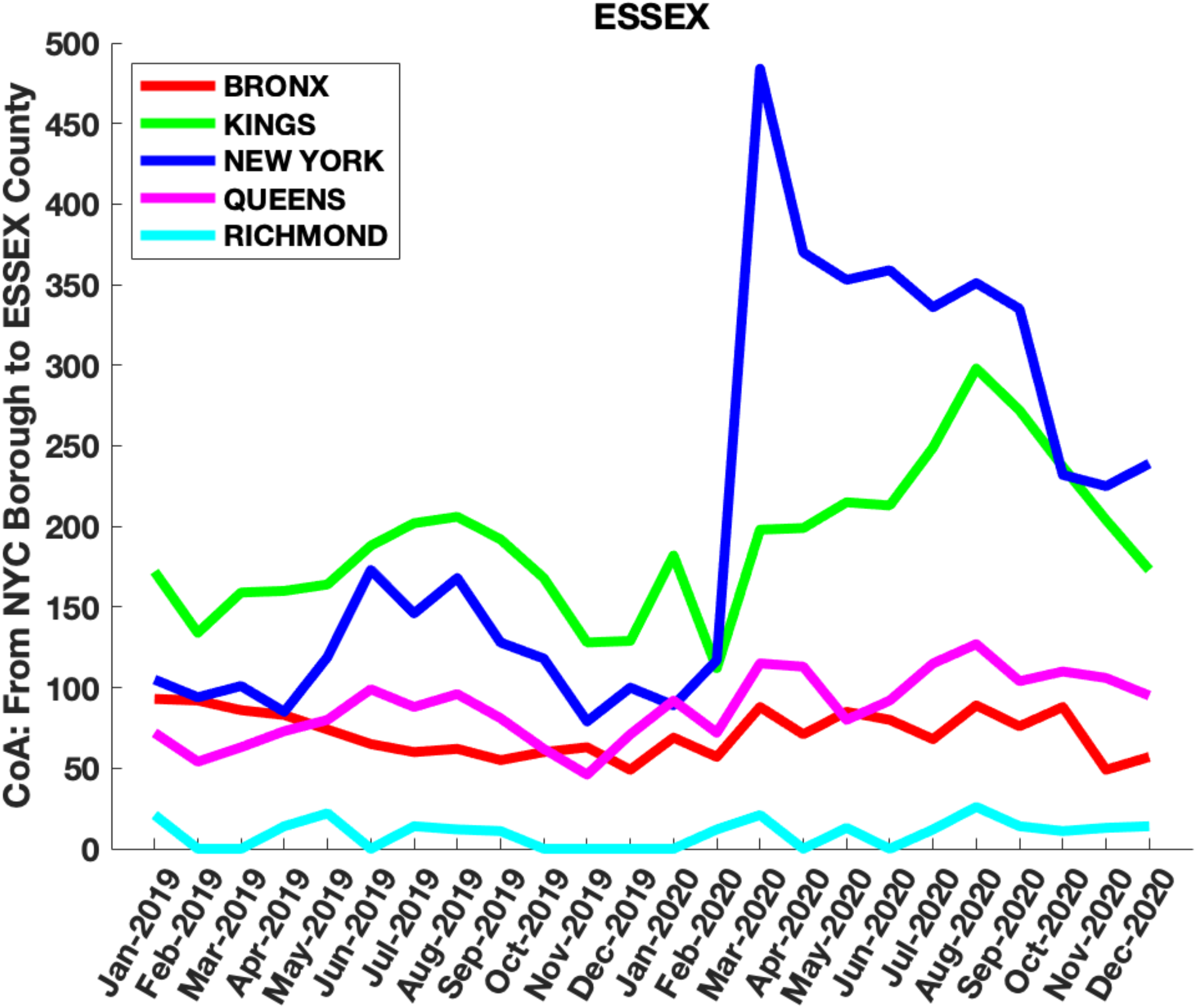
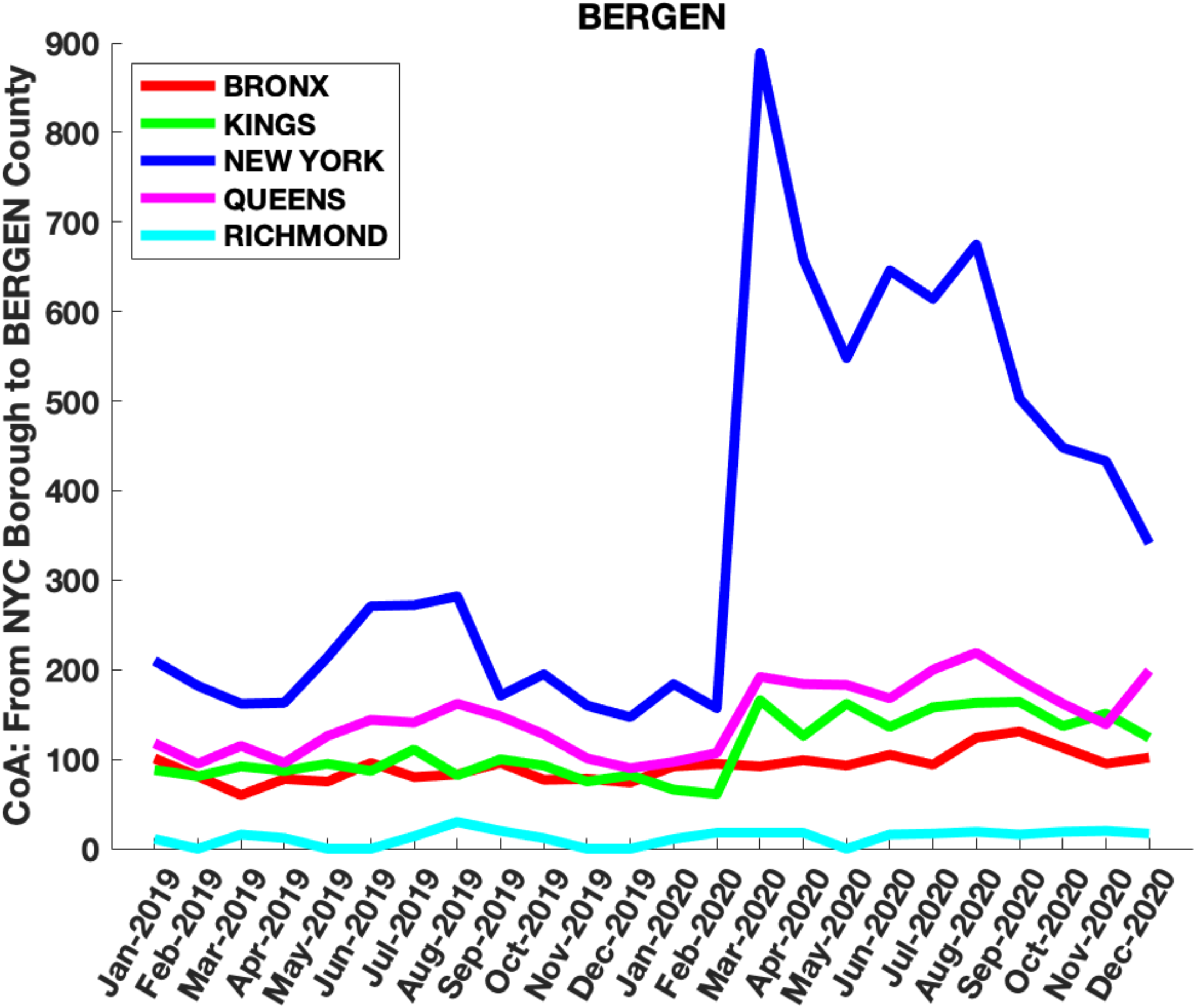
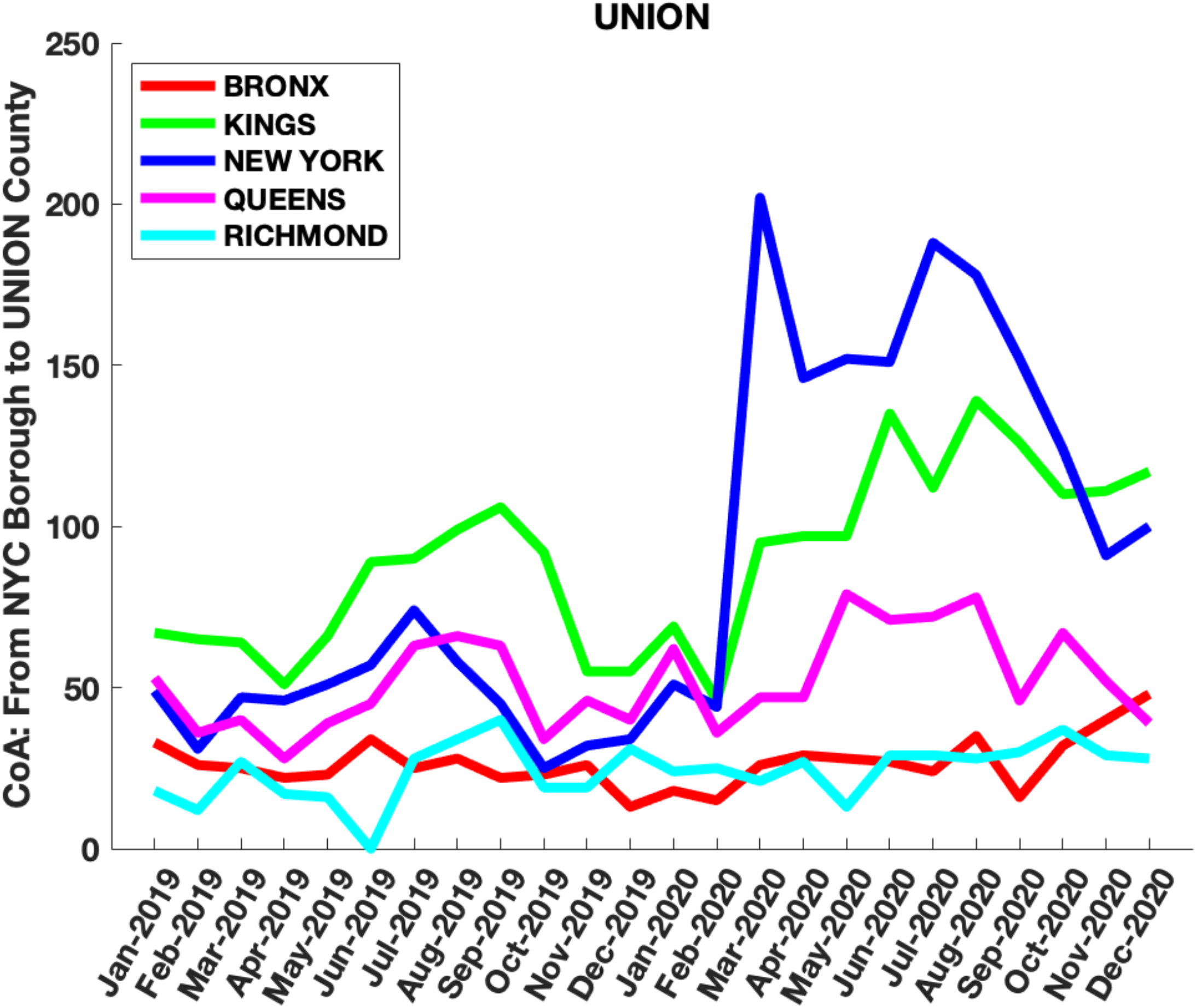
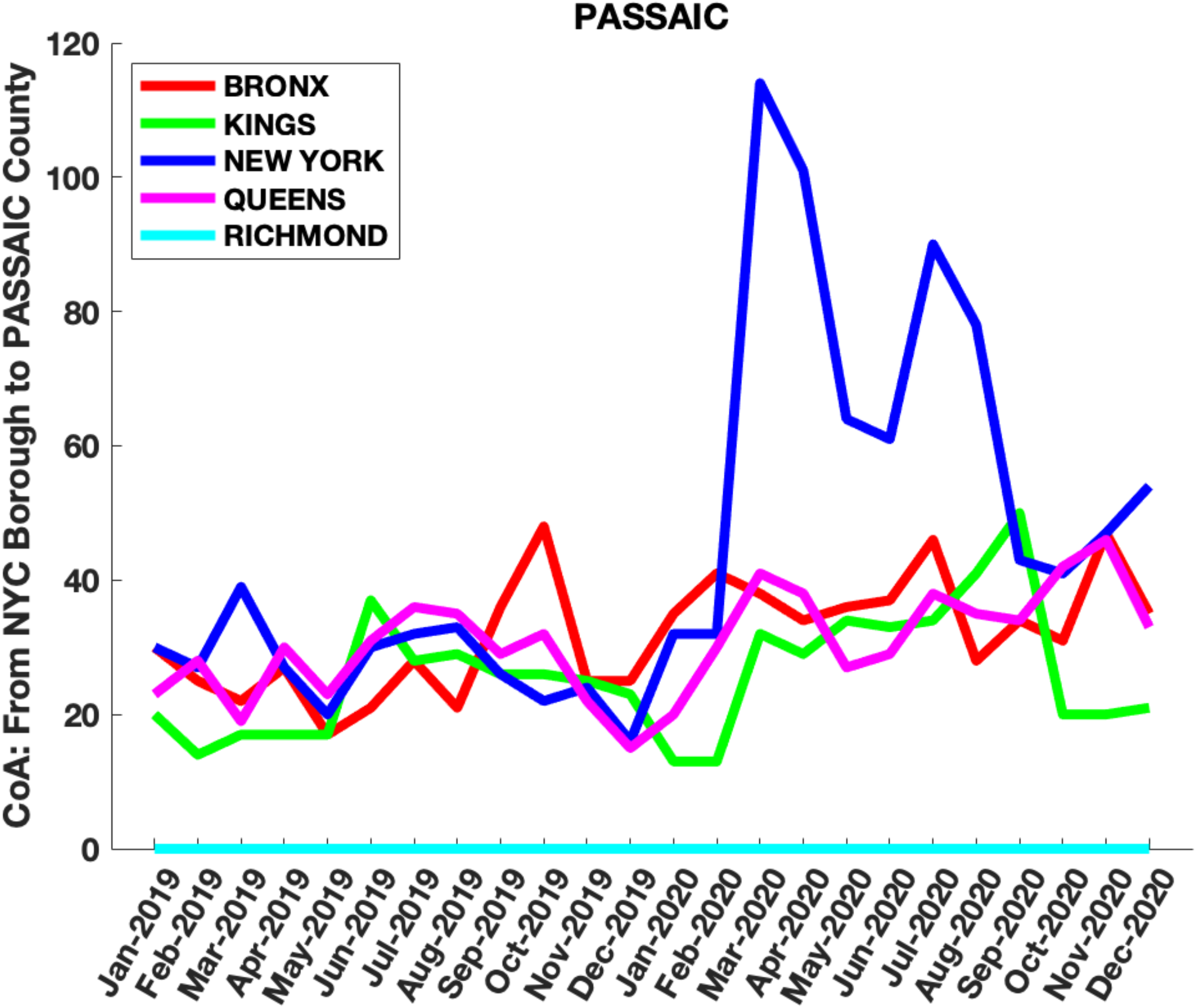
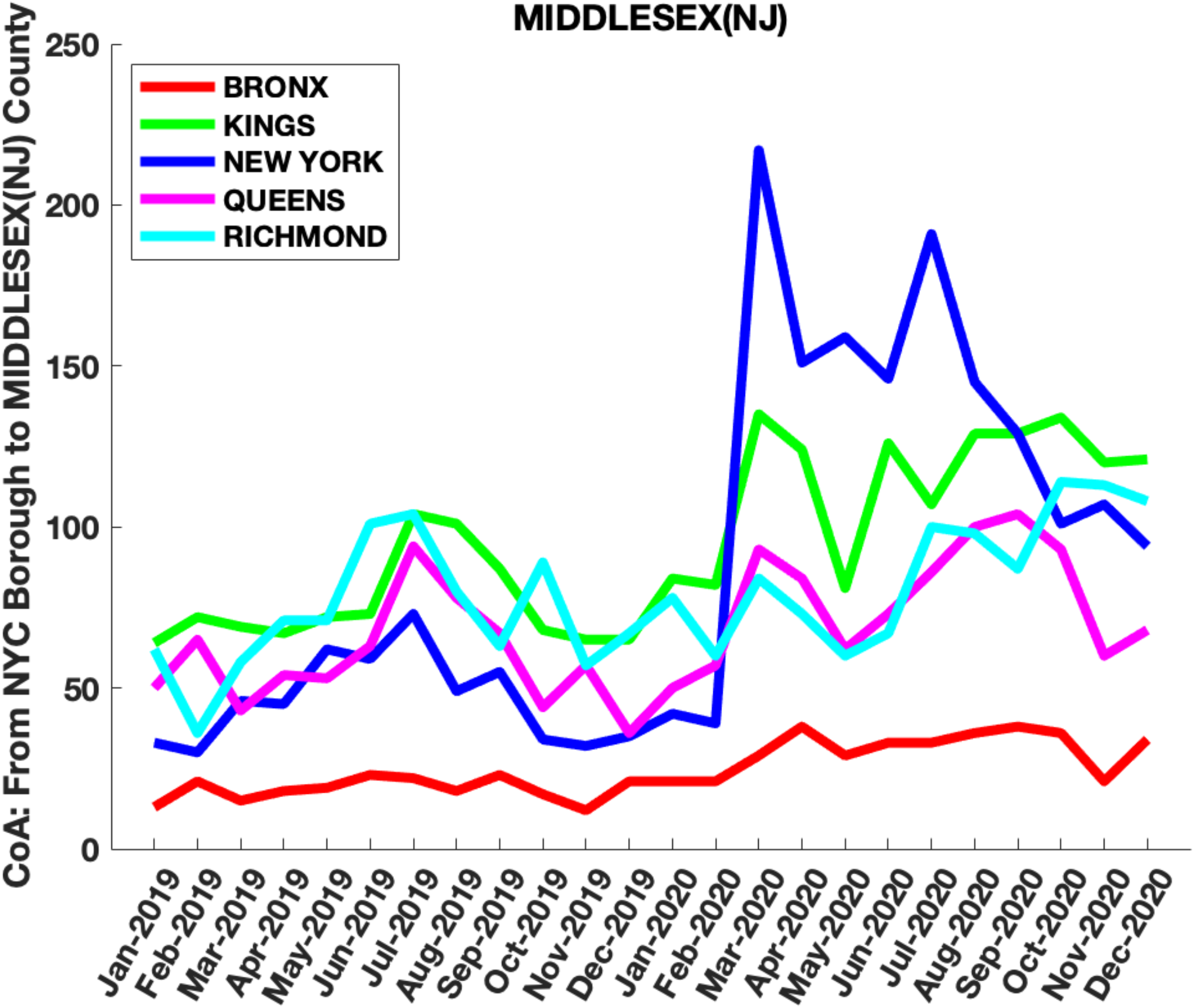
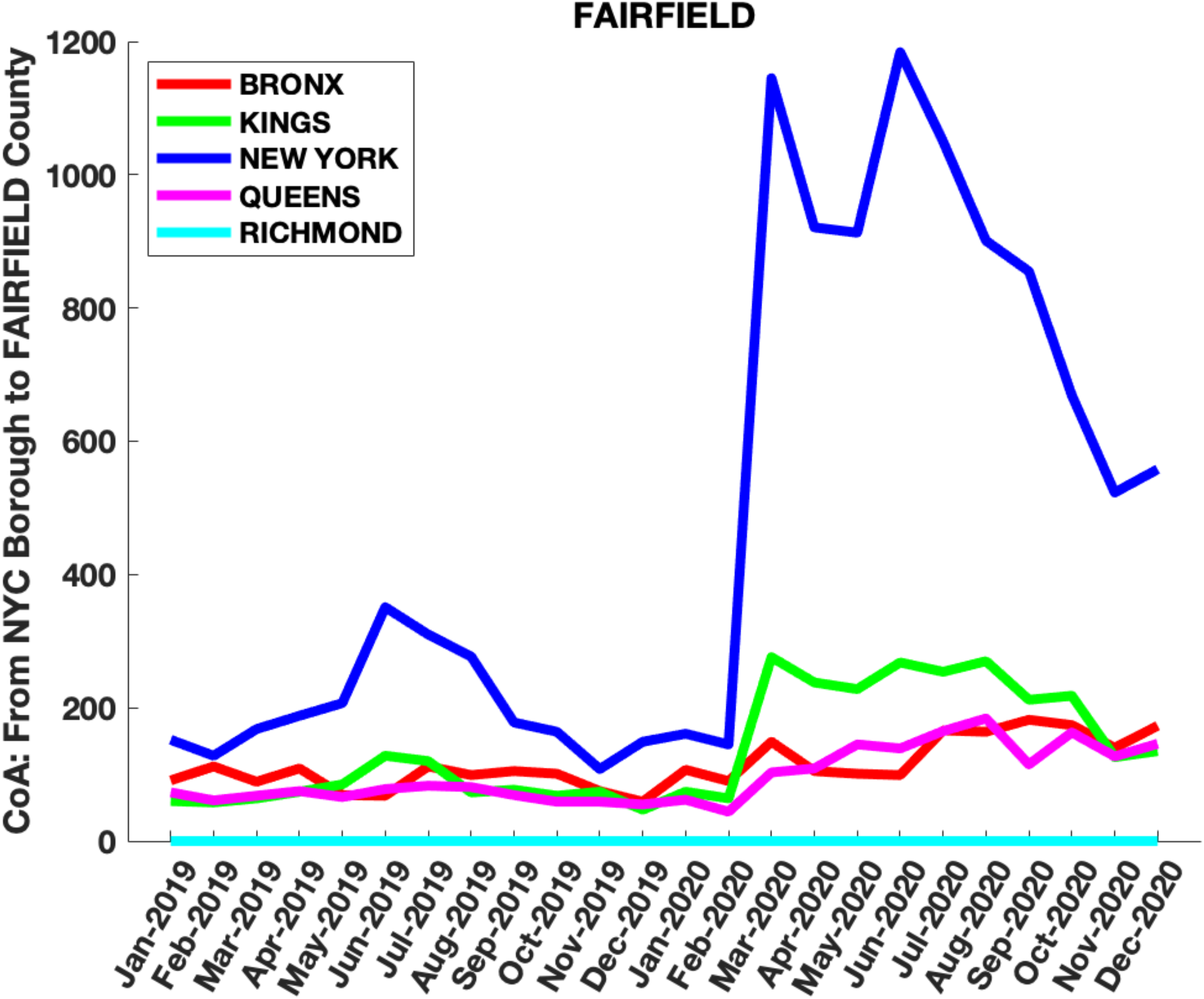
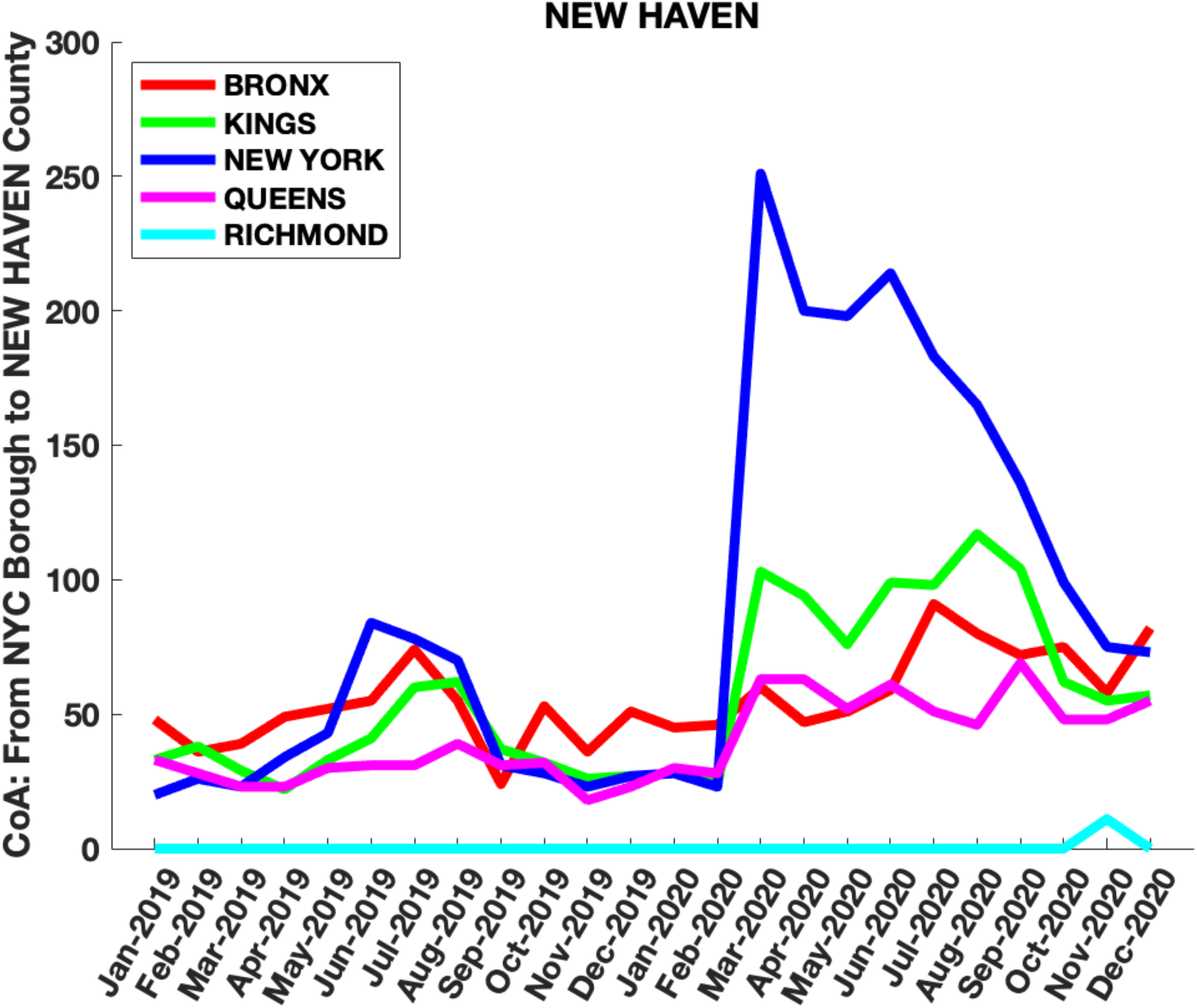
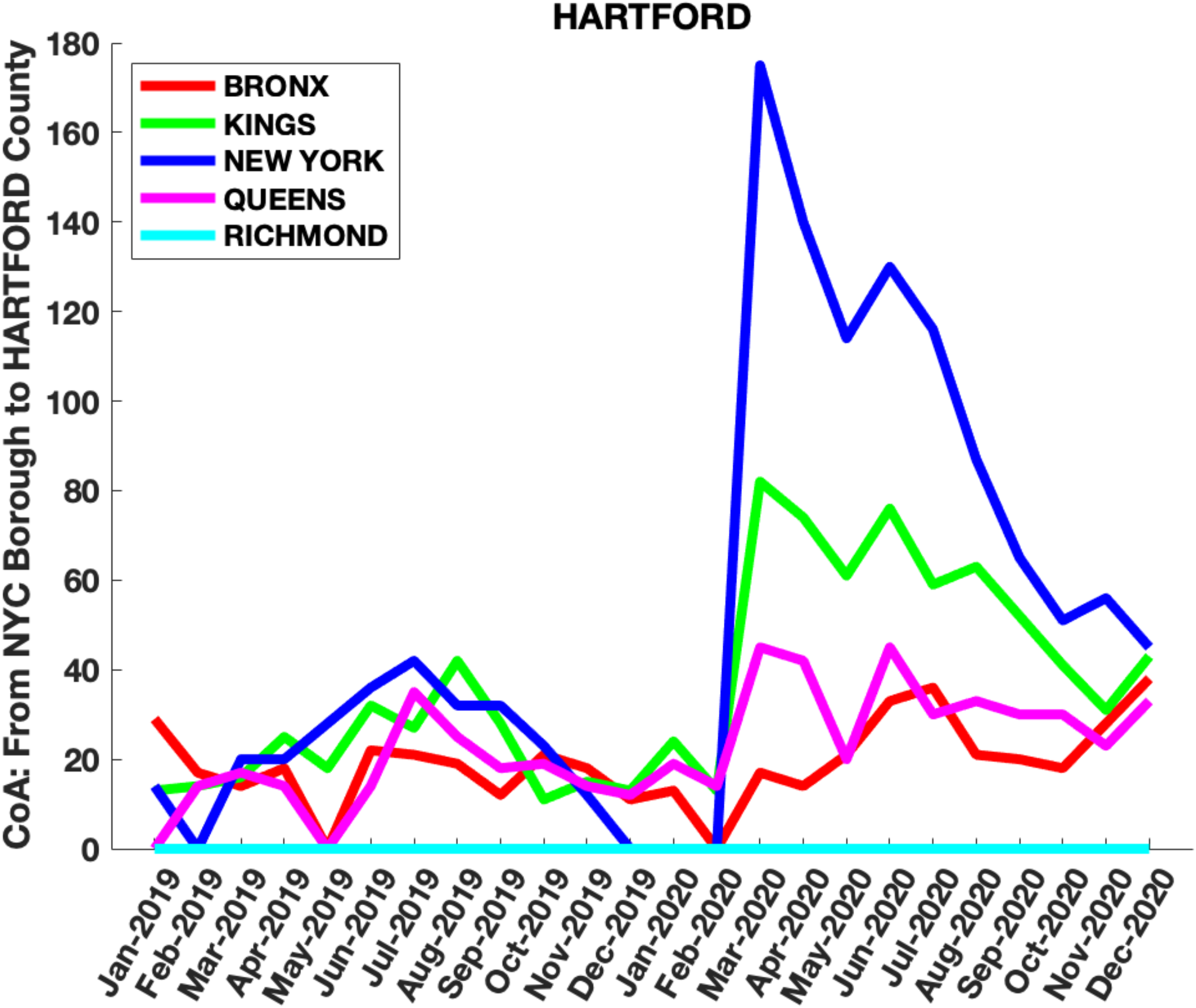
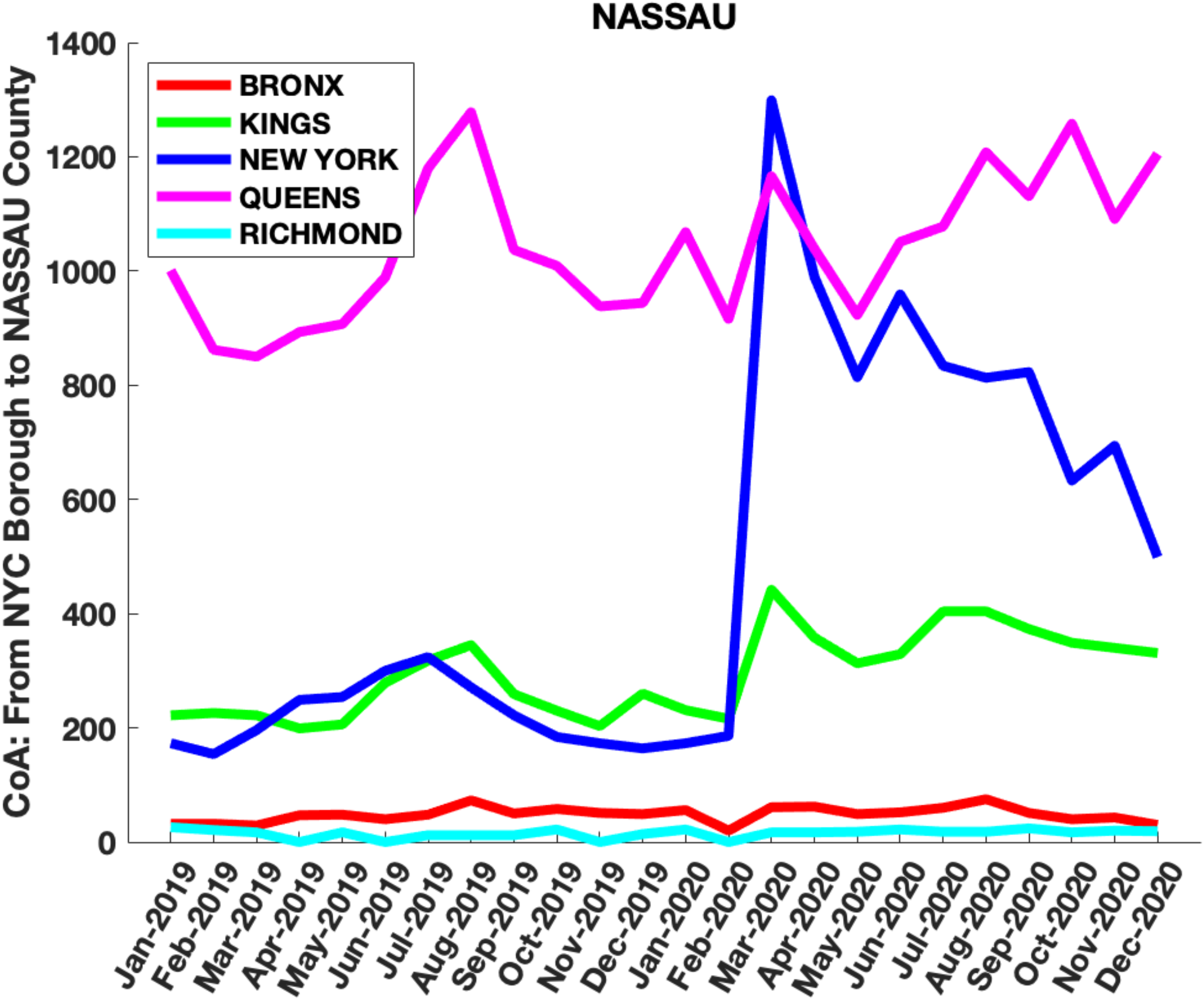
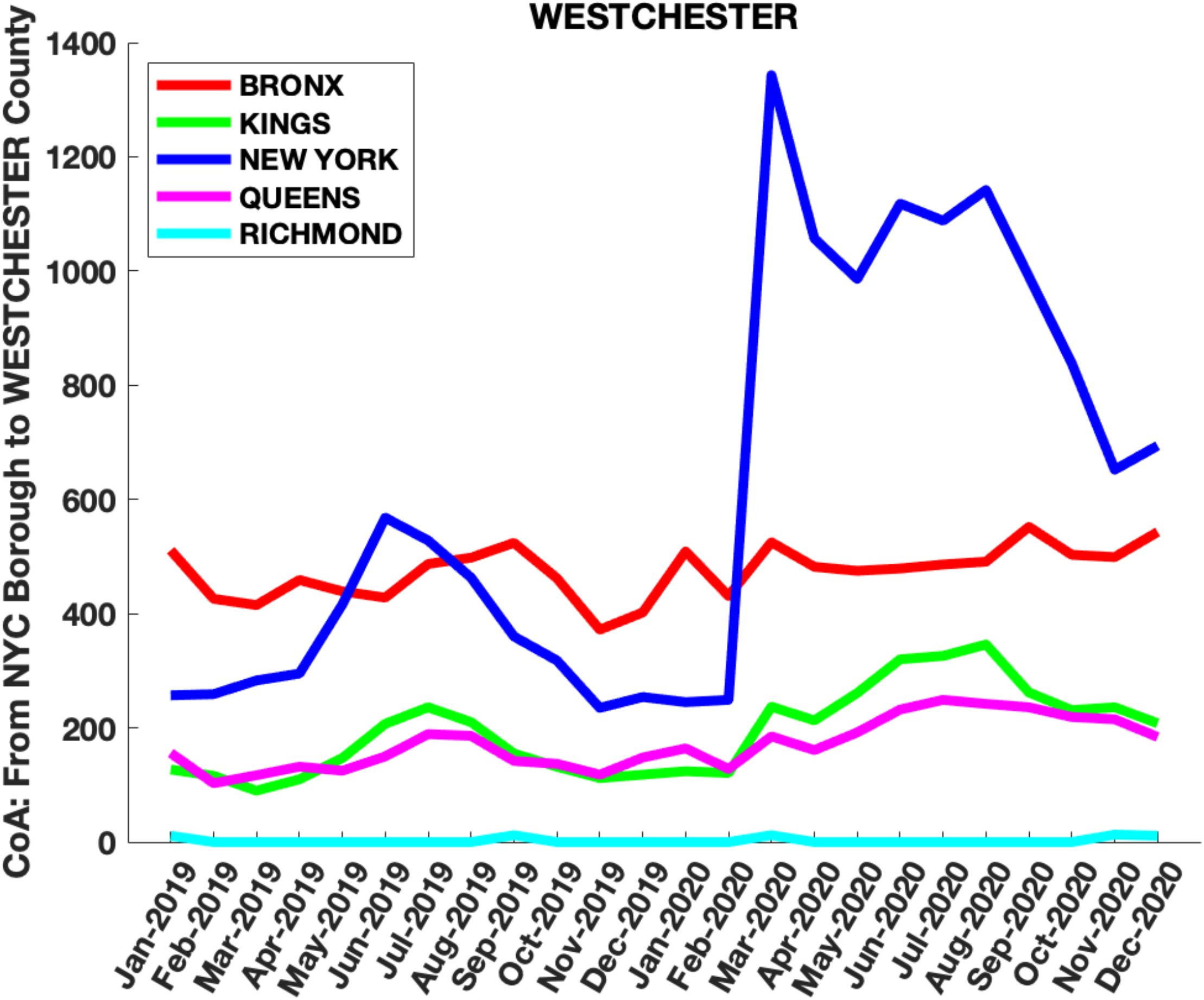
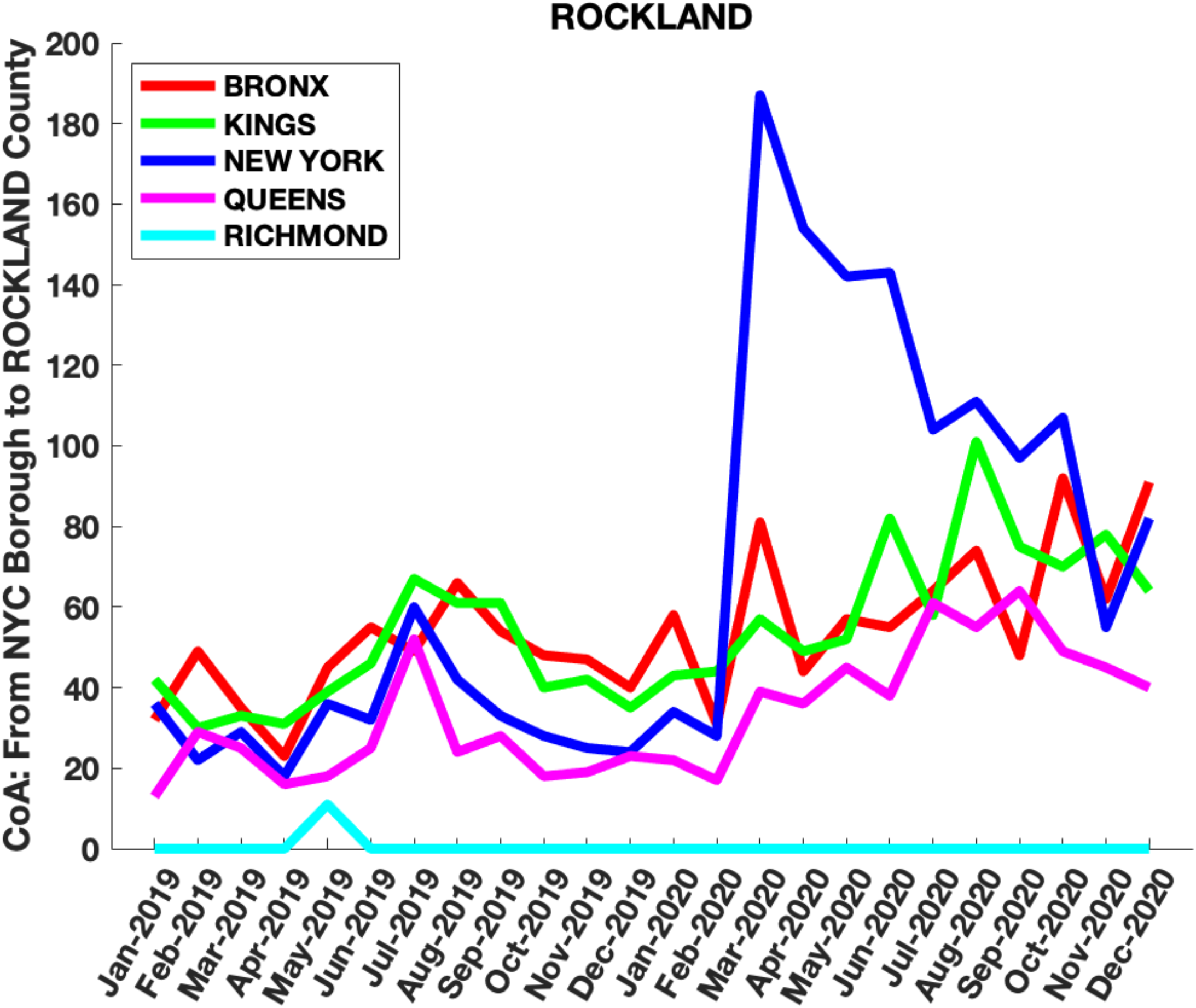
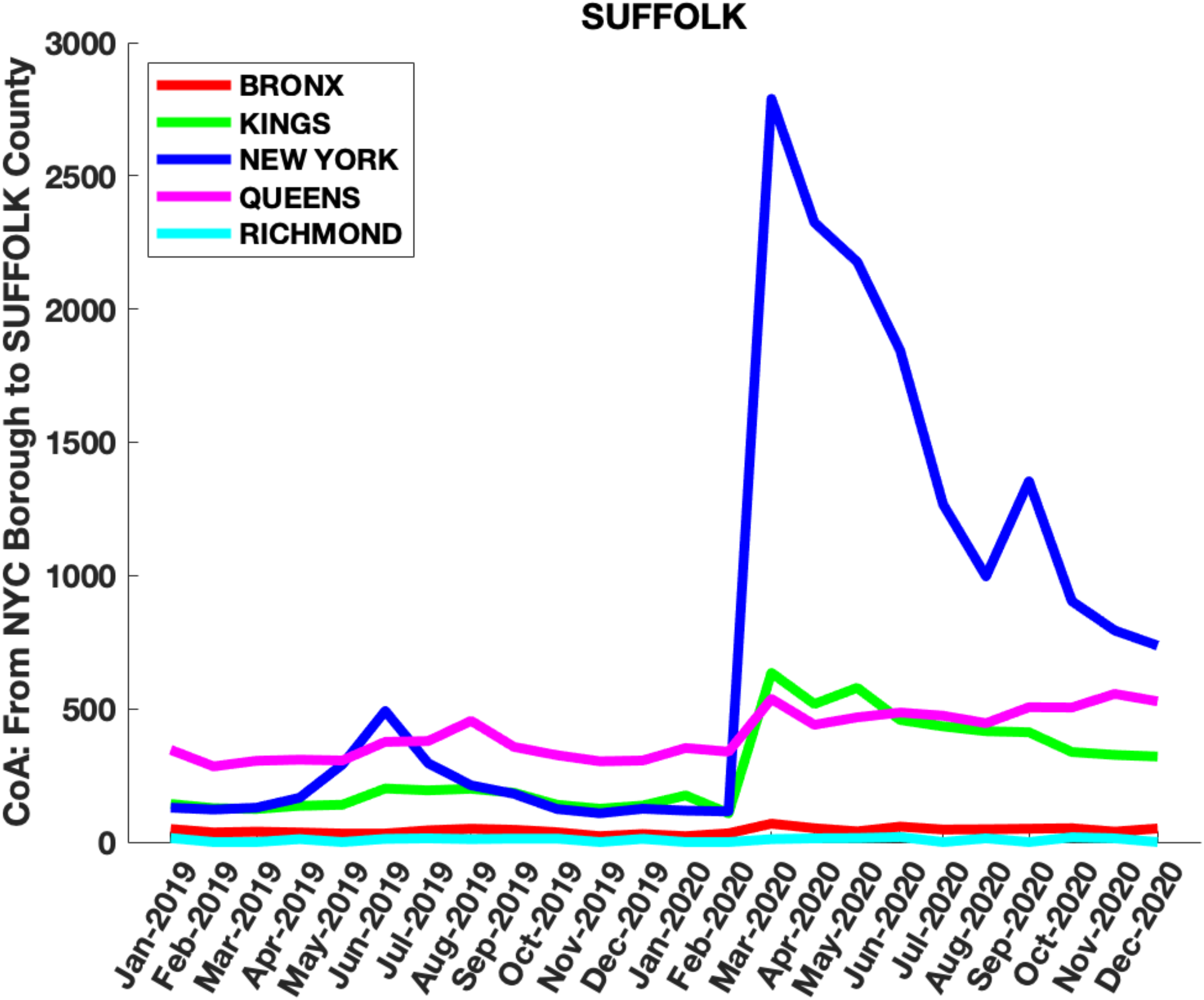
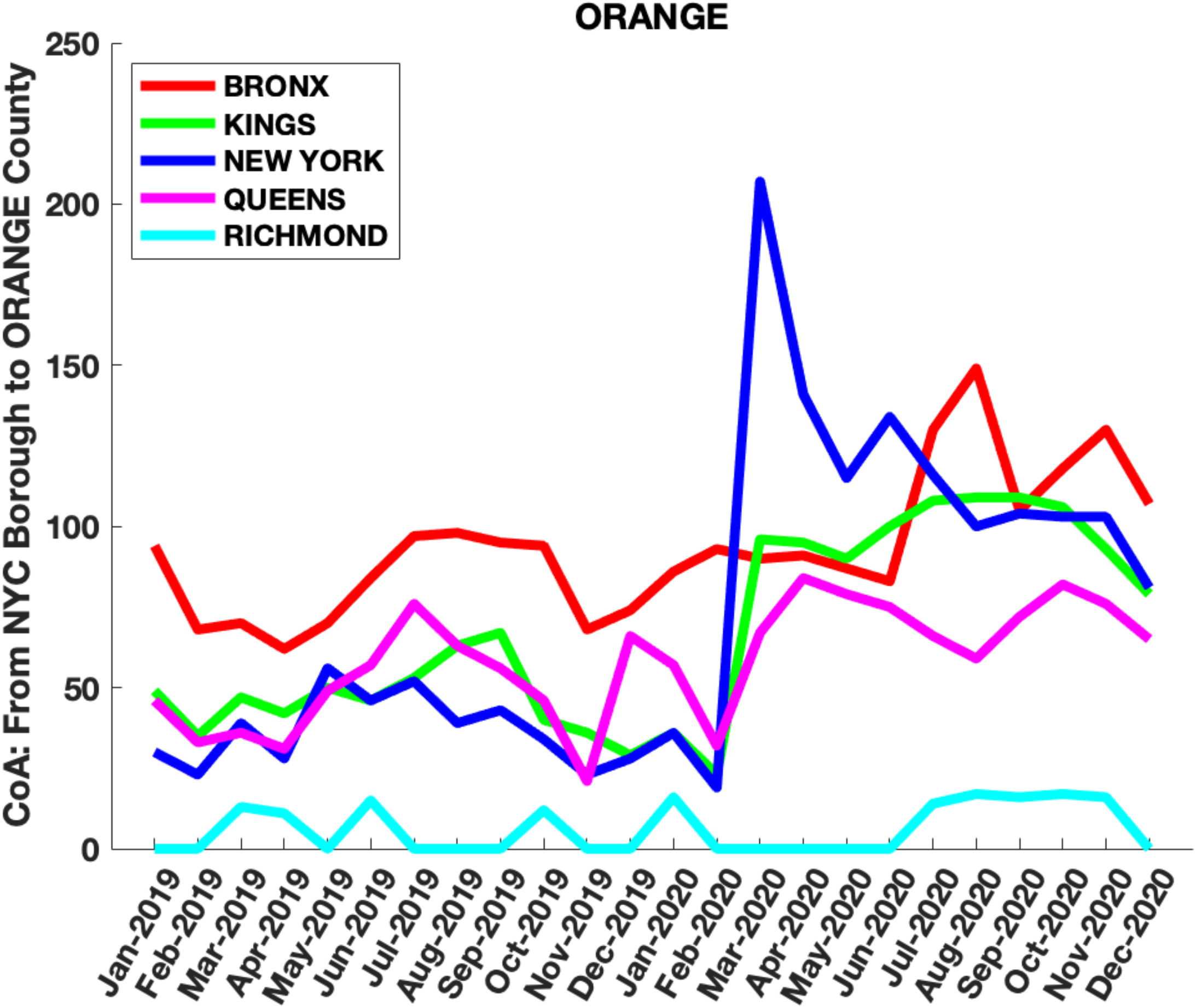
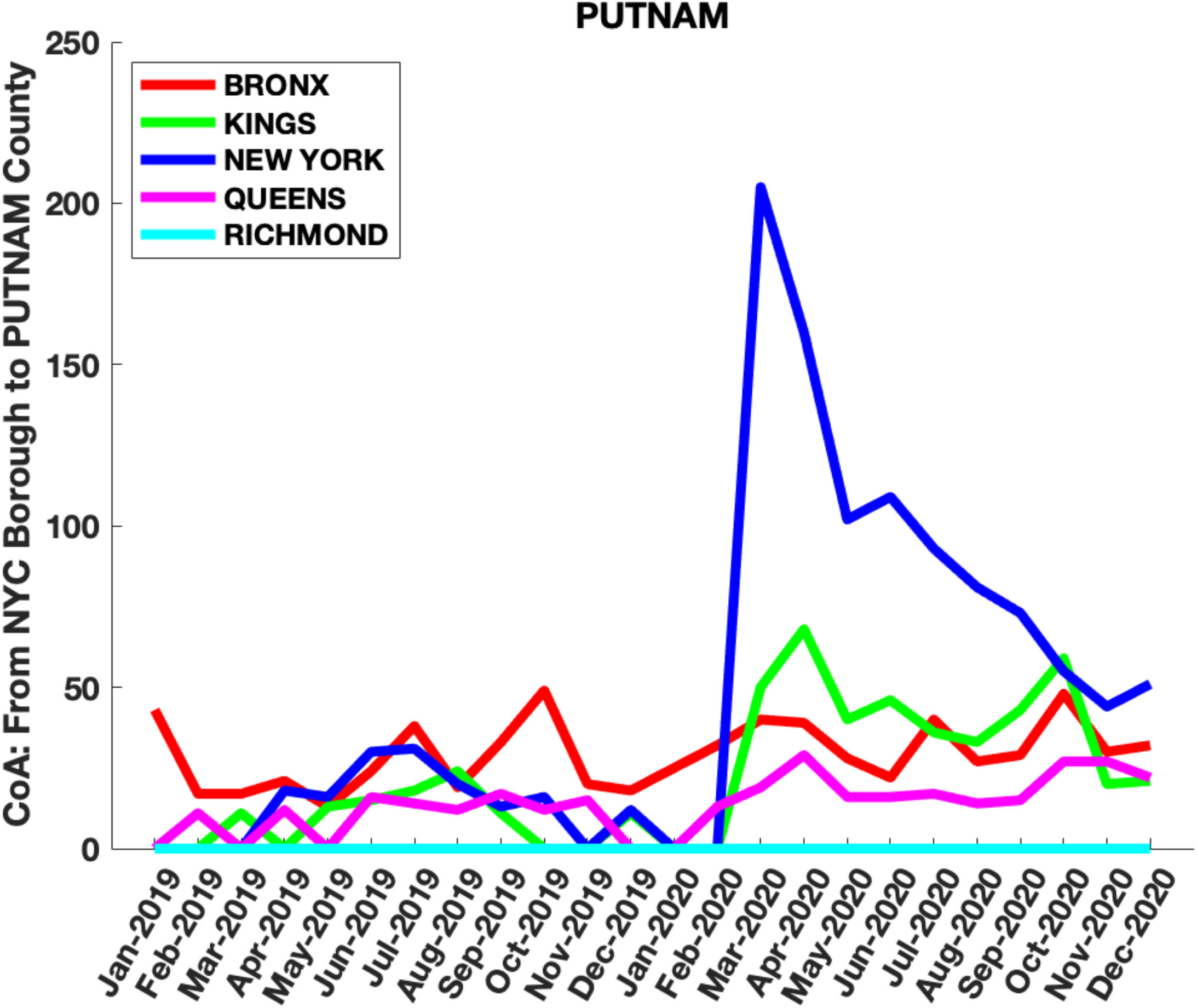
Monthly CoA counts from each NYC borough into the most densely populated counties in NJ, CT and NY States: **Figures 3a-f:** Hudson, Essex, Bergen, Union, Passaic and Middlesex counties in NJ, **Figures 3g-i** Fairfield, New Haven and Hartford counties in CT and **Figures 3j-o** Nassau, Westchester, Rockland, Suffolk, Orange and Putnam counties in NY State which are closest to NYC. Note the sharp rise in household moves from NYC boroughs, especially New York Borough (Manhattan), into the surrounding counties from March-December 2020 compared to the same period in 2019.

### Analysis of Covid-19 Daily Cases in NY, NJ, and CT

Seven-day averaged plots of daily Covid-19 cases in NYC, NJ, CT and NY State as a function of time show an initial first peak in daily cases around April 1, 2020, superimposed on secondary shoulders or peaks from mid-April to June (Figure 4 and Supplementary Figures 6,8,10). The first peak reflects the time dynamics of an initial exponential rise in infections in mid-March 2020 followed by a reduction in cases from quarantine and lockdown measures. However, the cause of the second peak has not yet been adequately explained. We propose that these second peaks were, in part, caused by the movement of households out of NYC to these counties, as identified by the CoA data described above.

**Figure 4:**
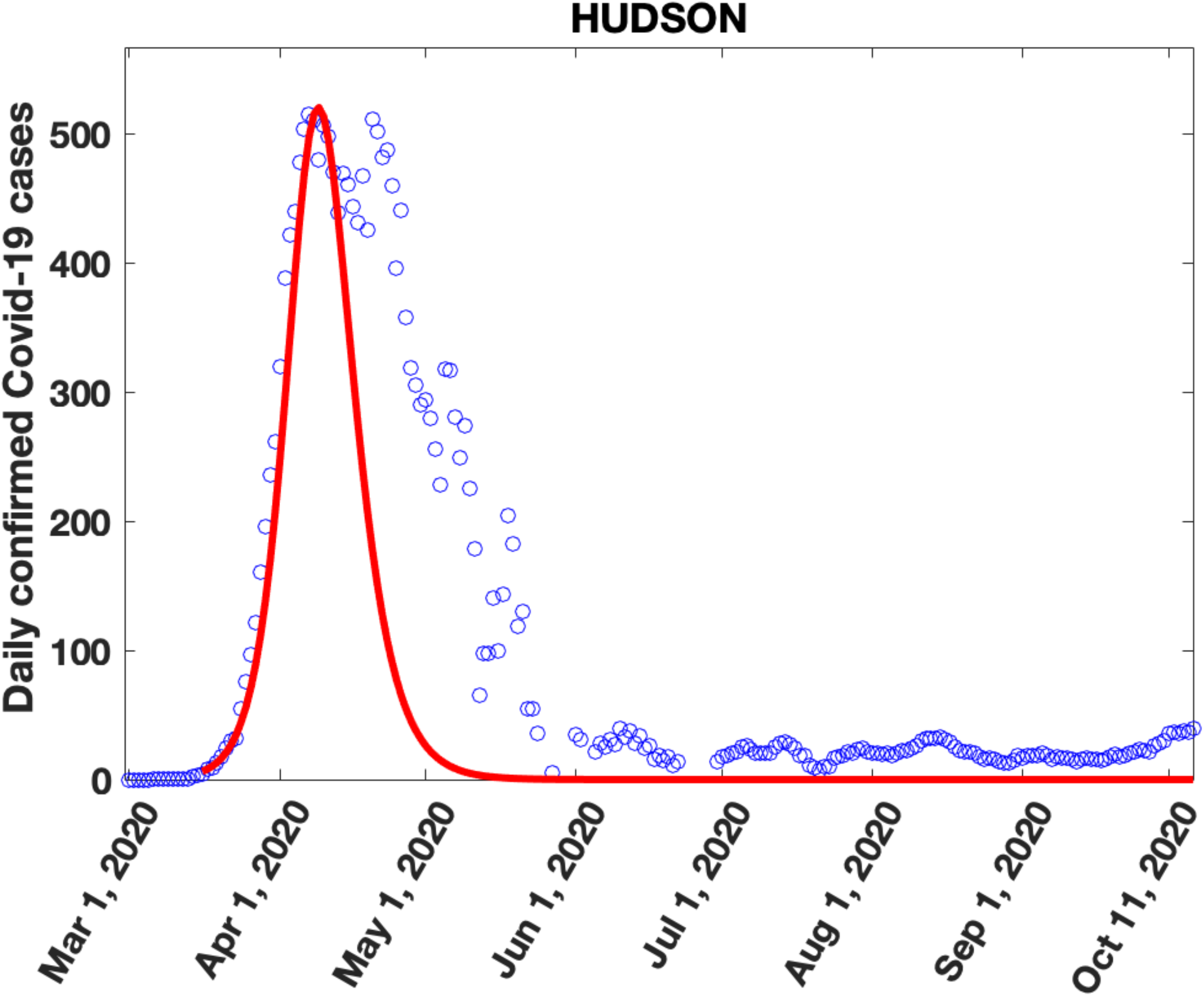
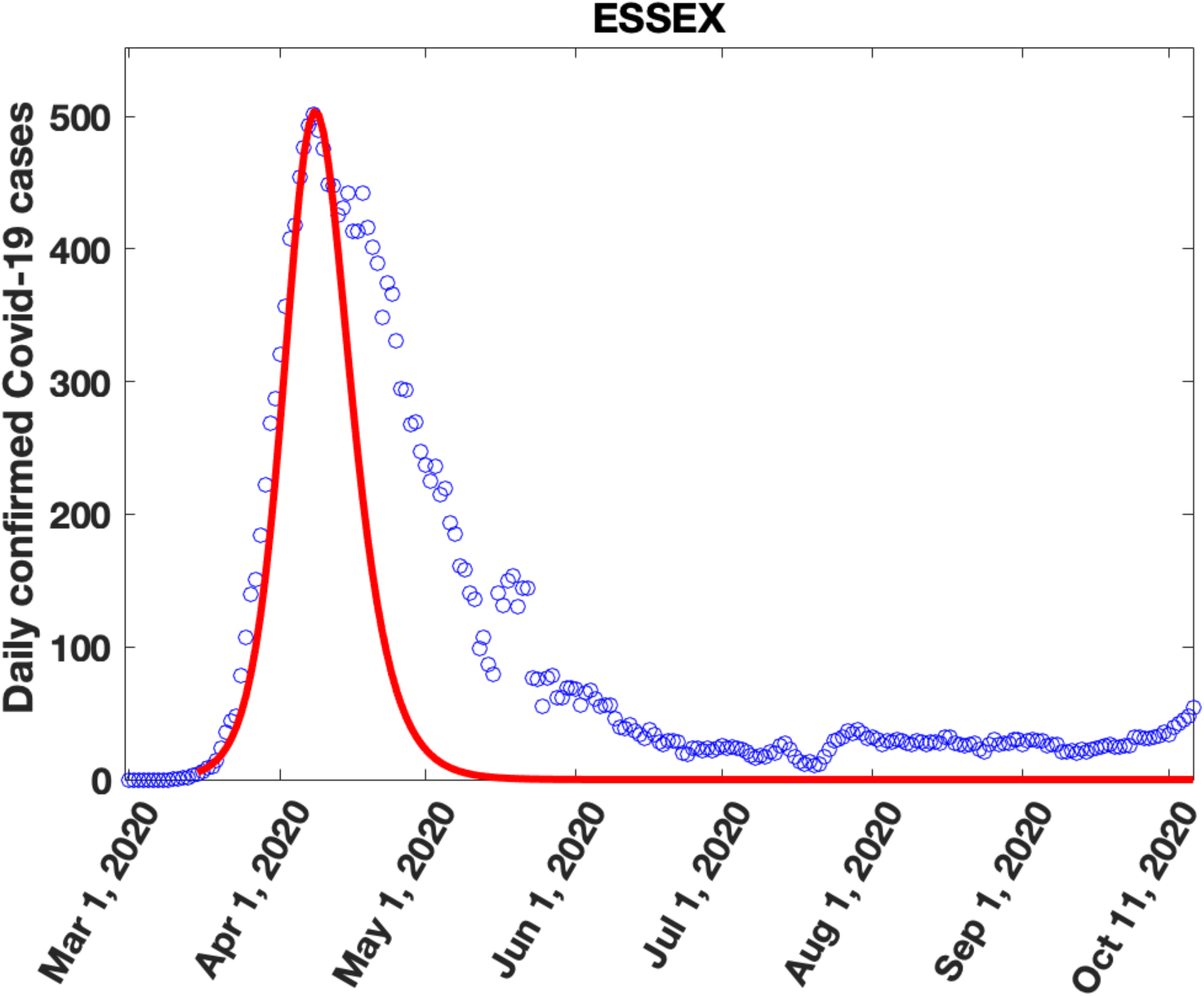
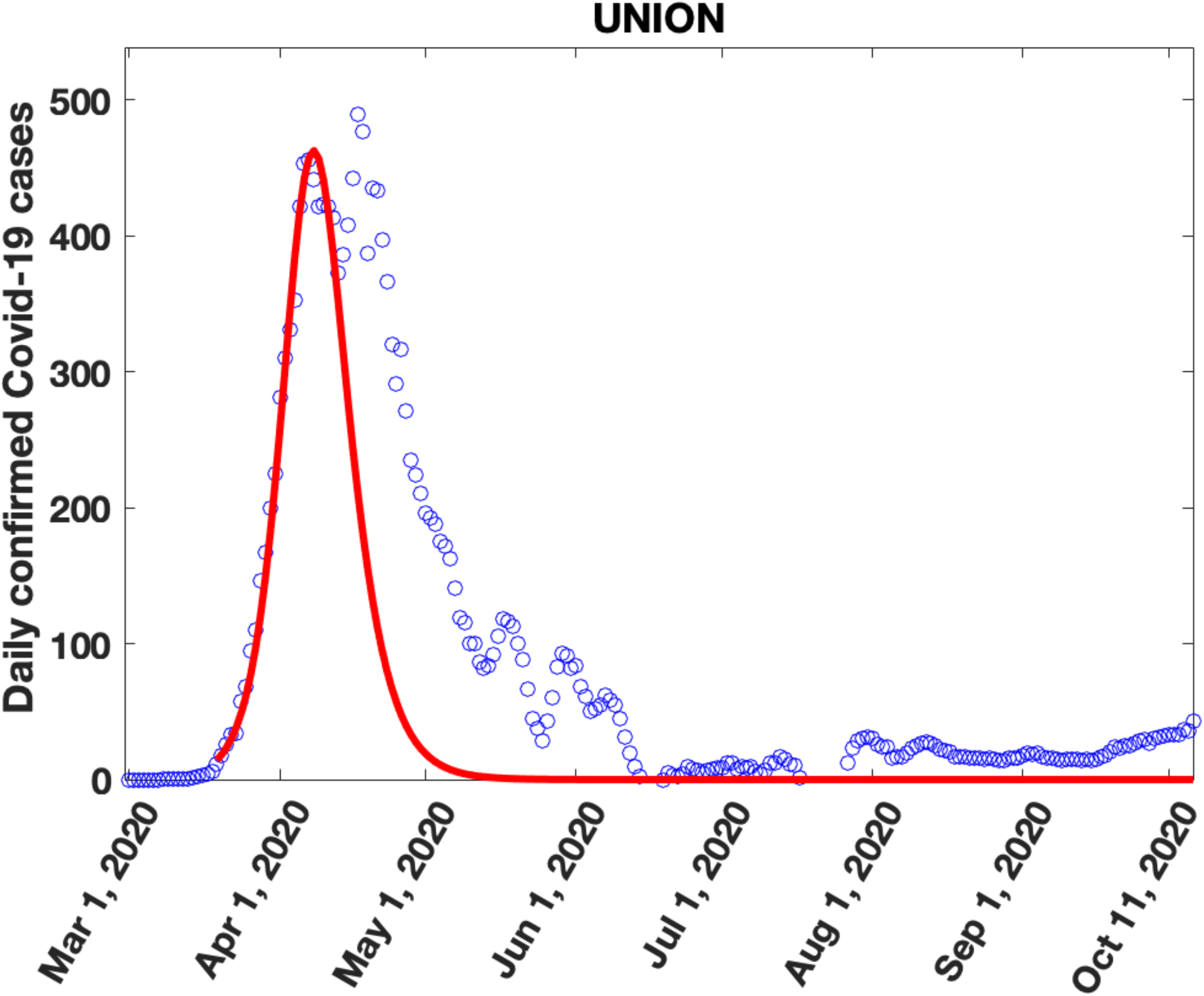
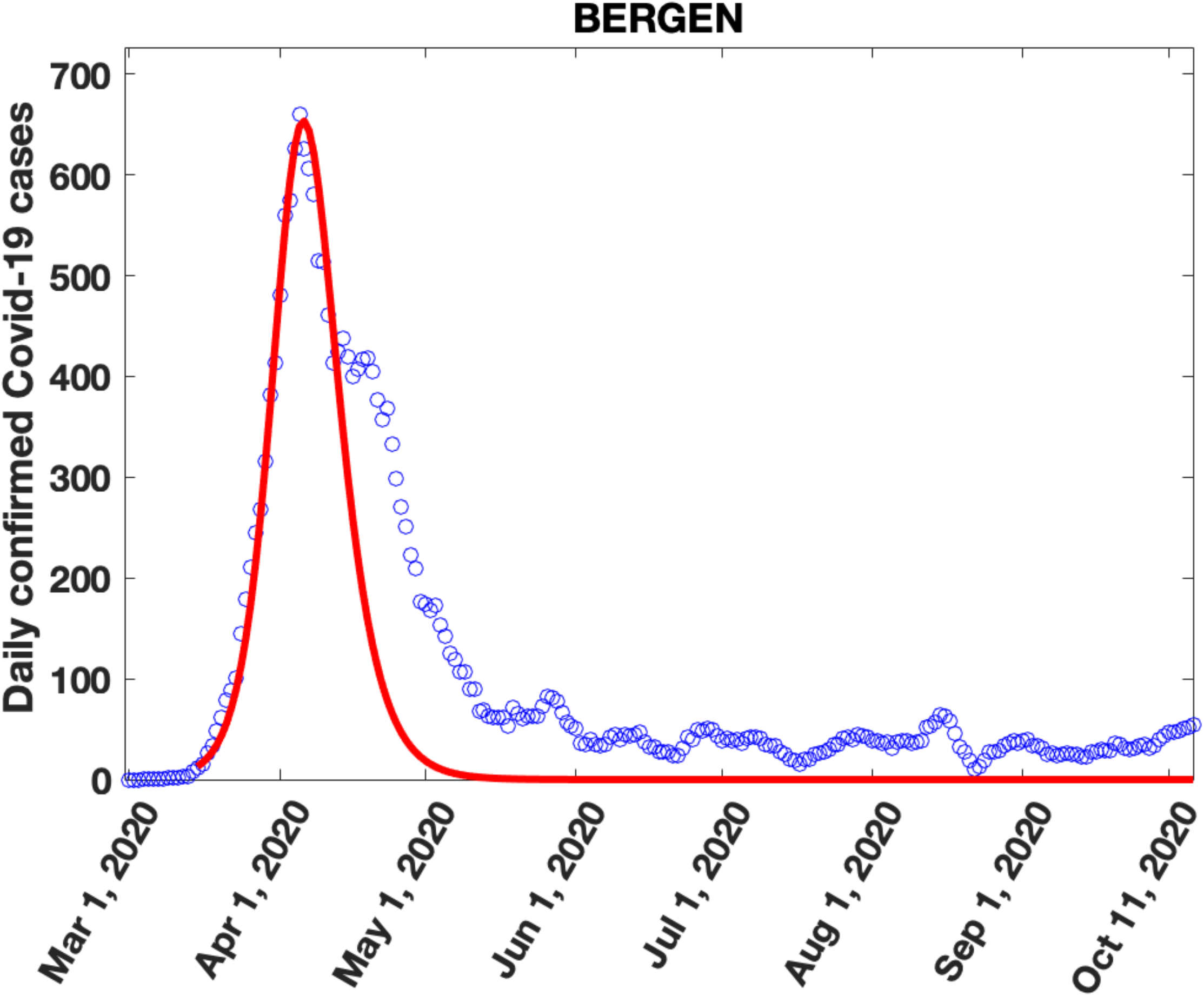
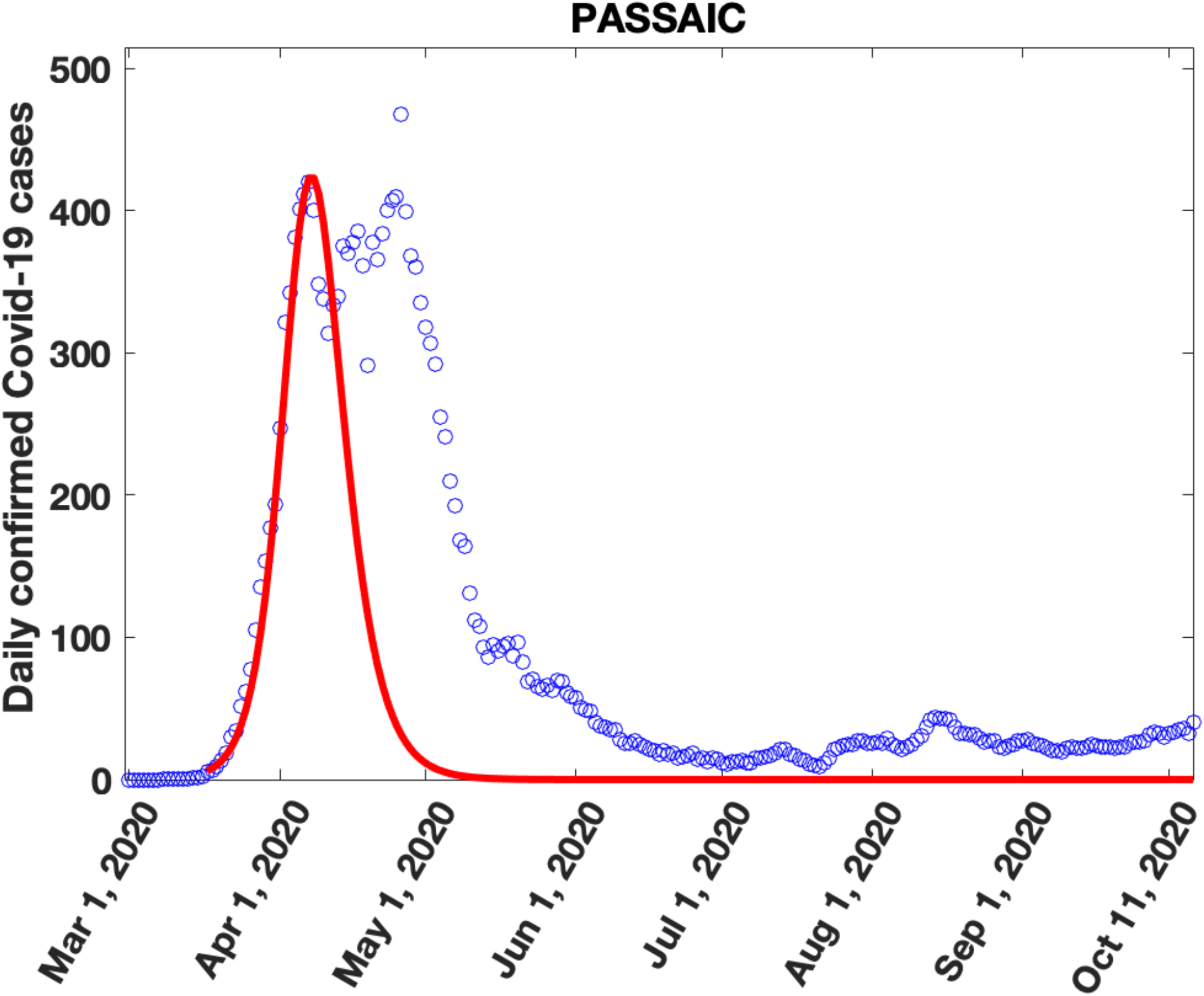
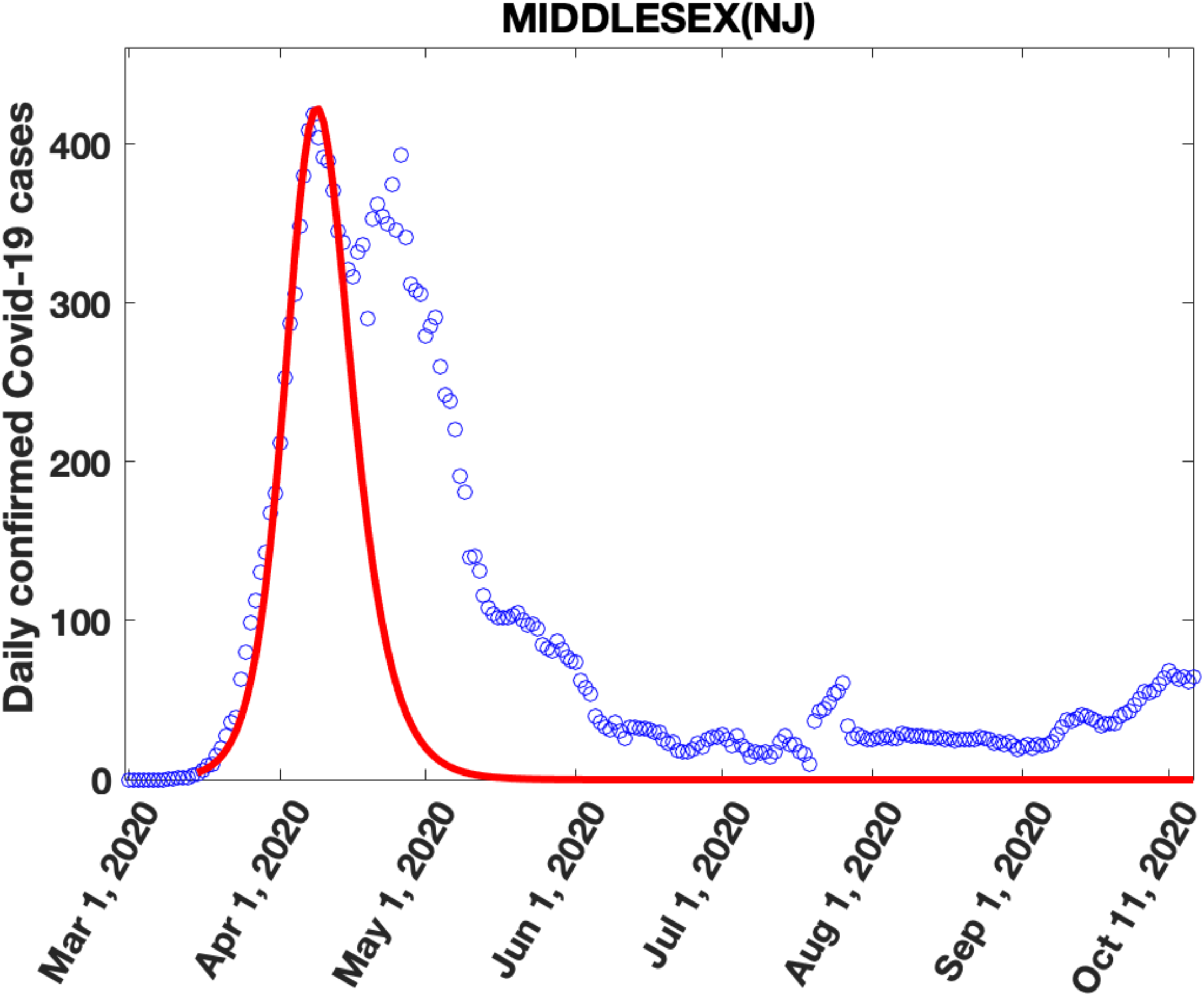
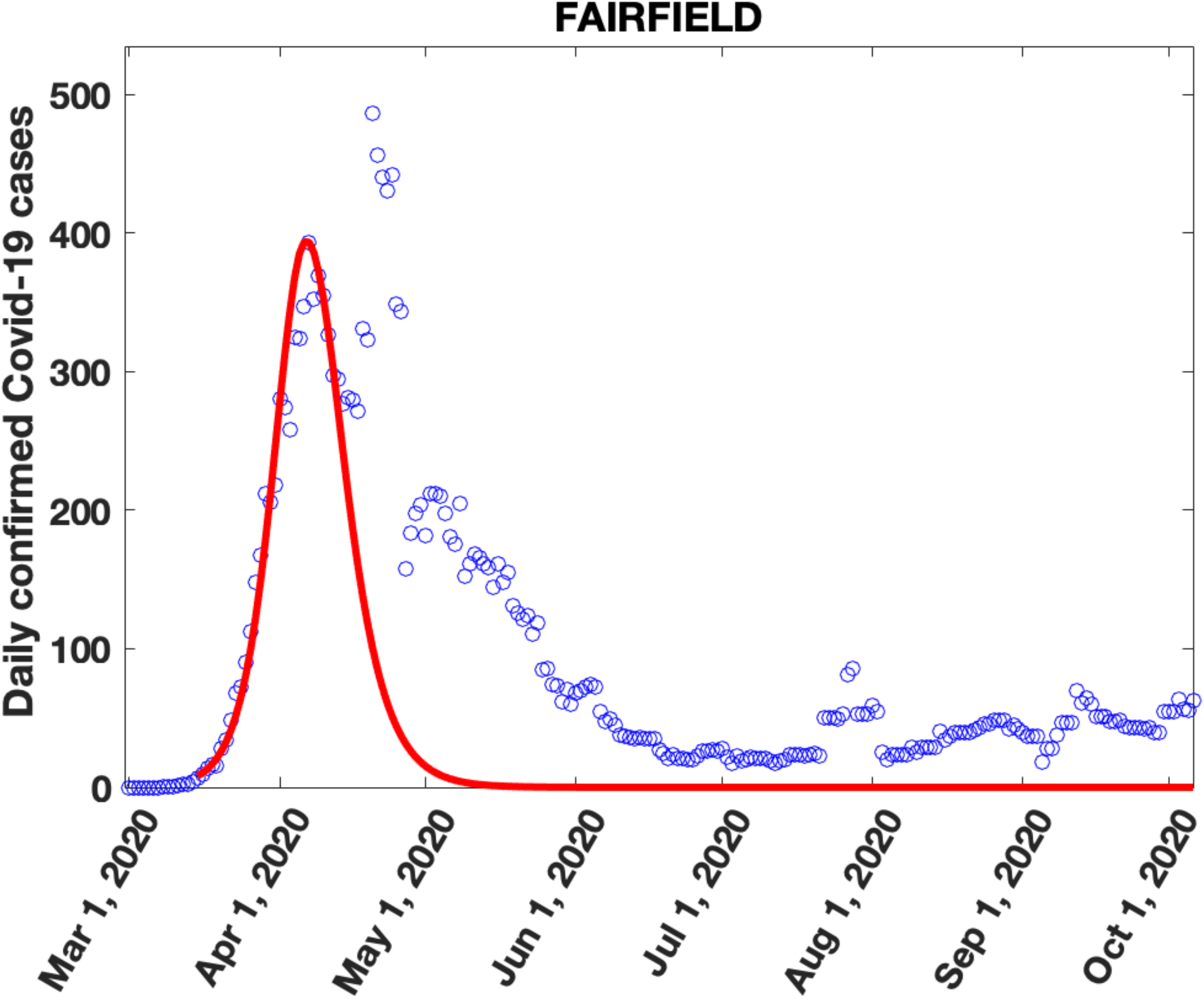
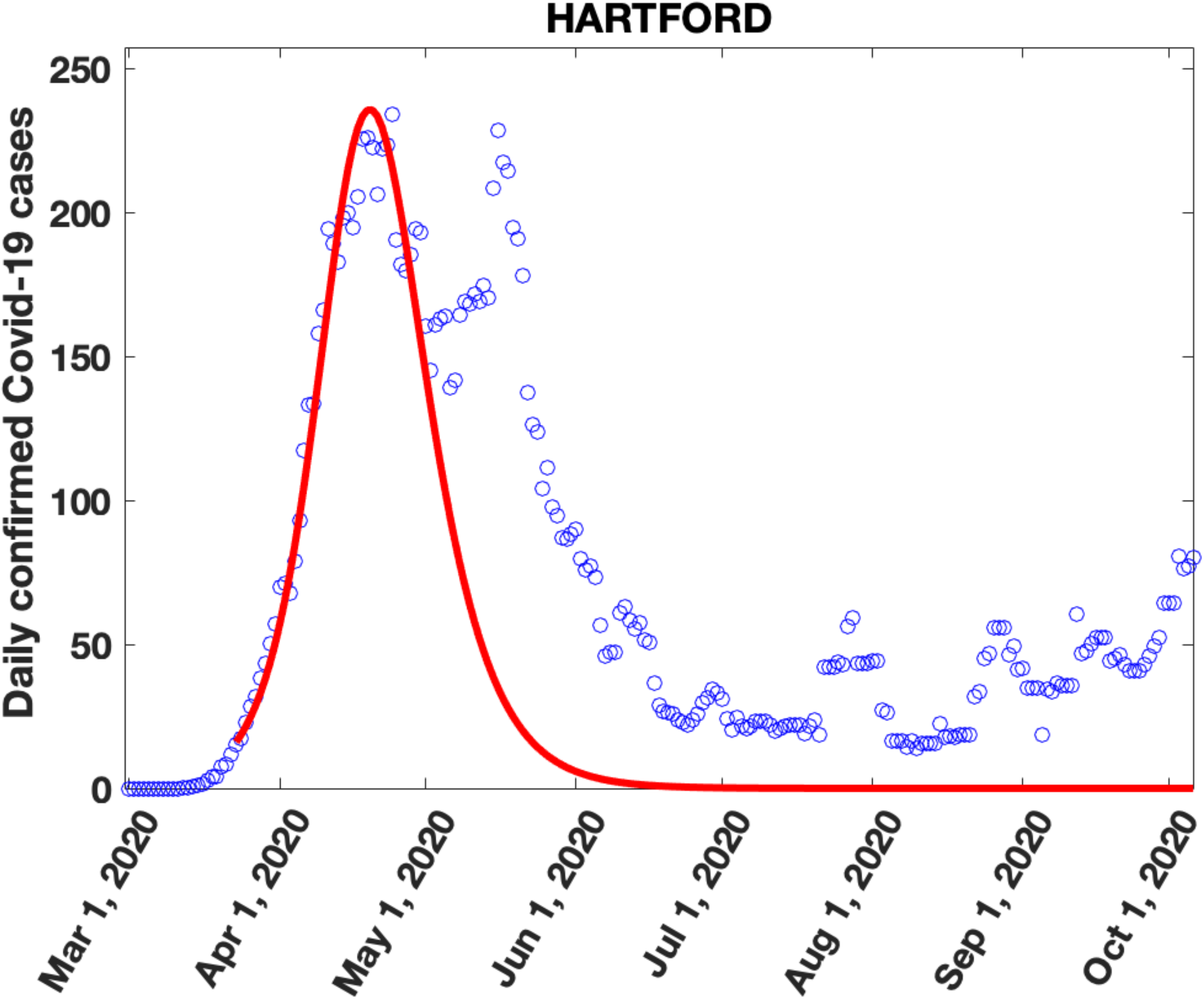
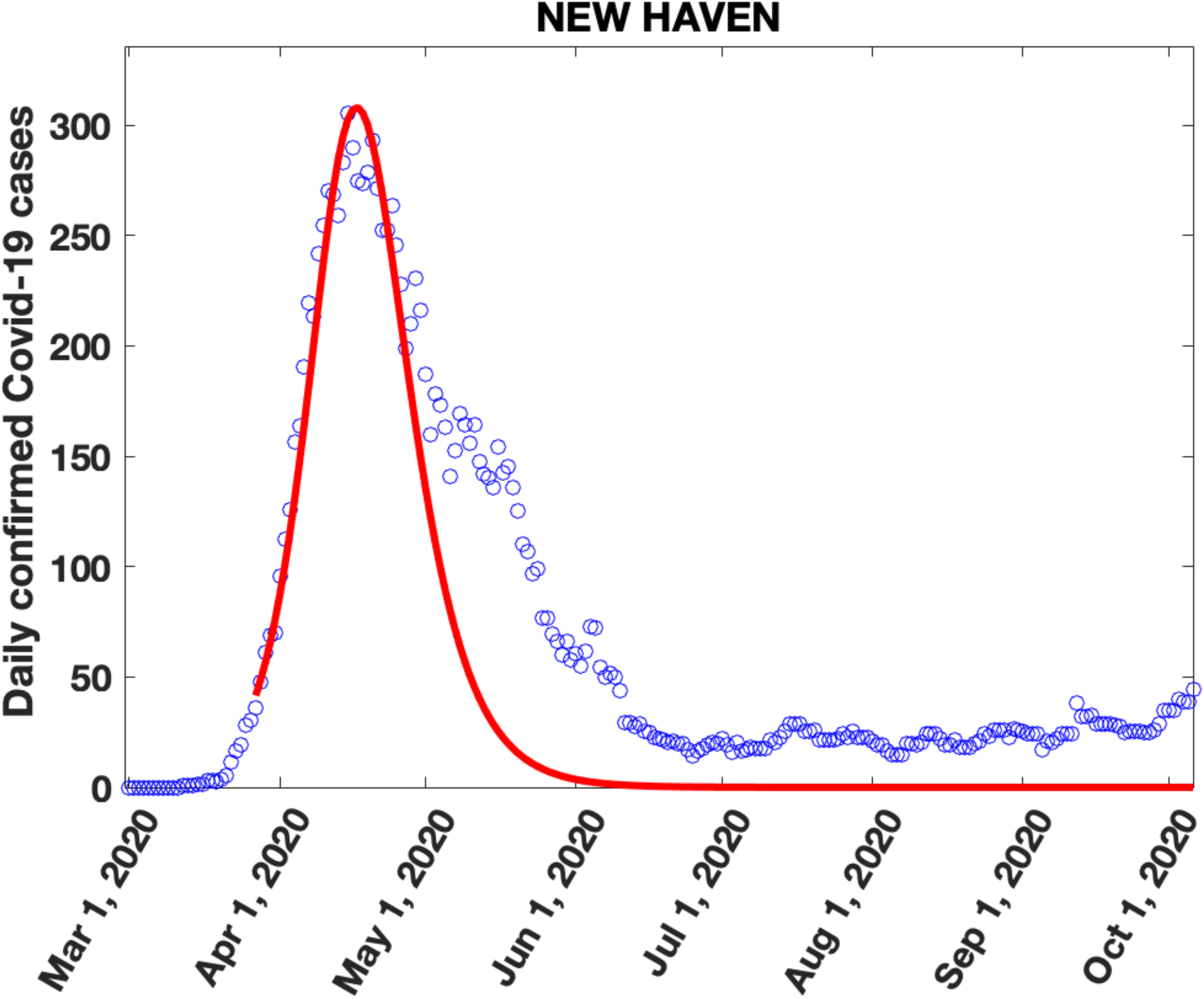
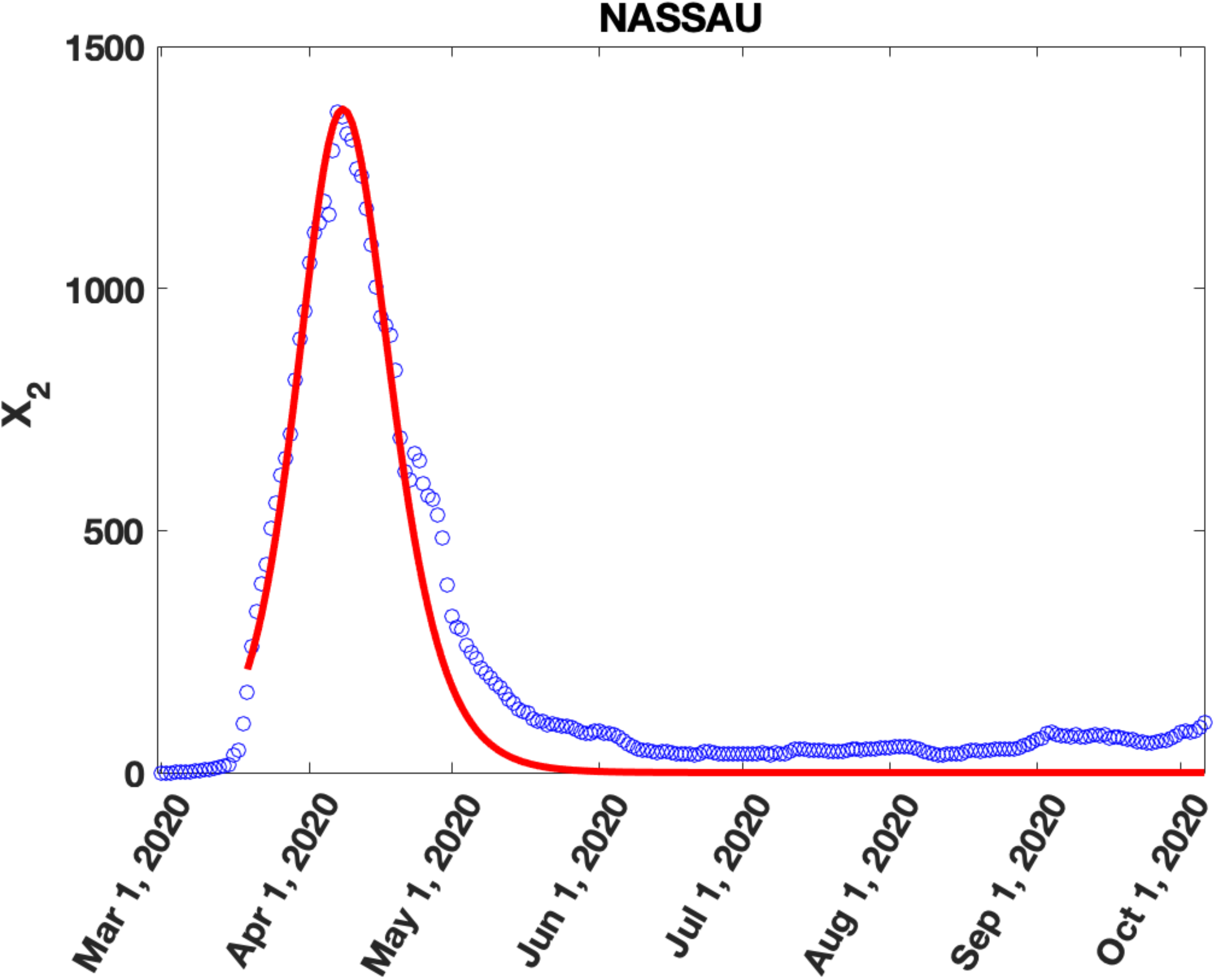
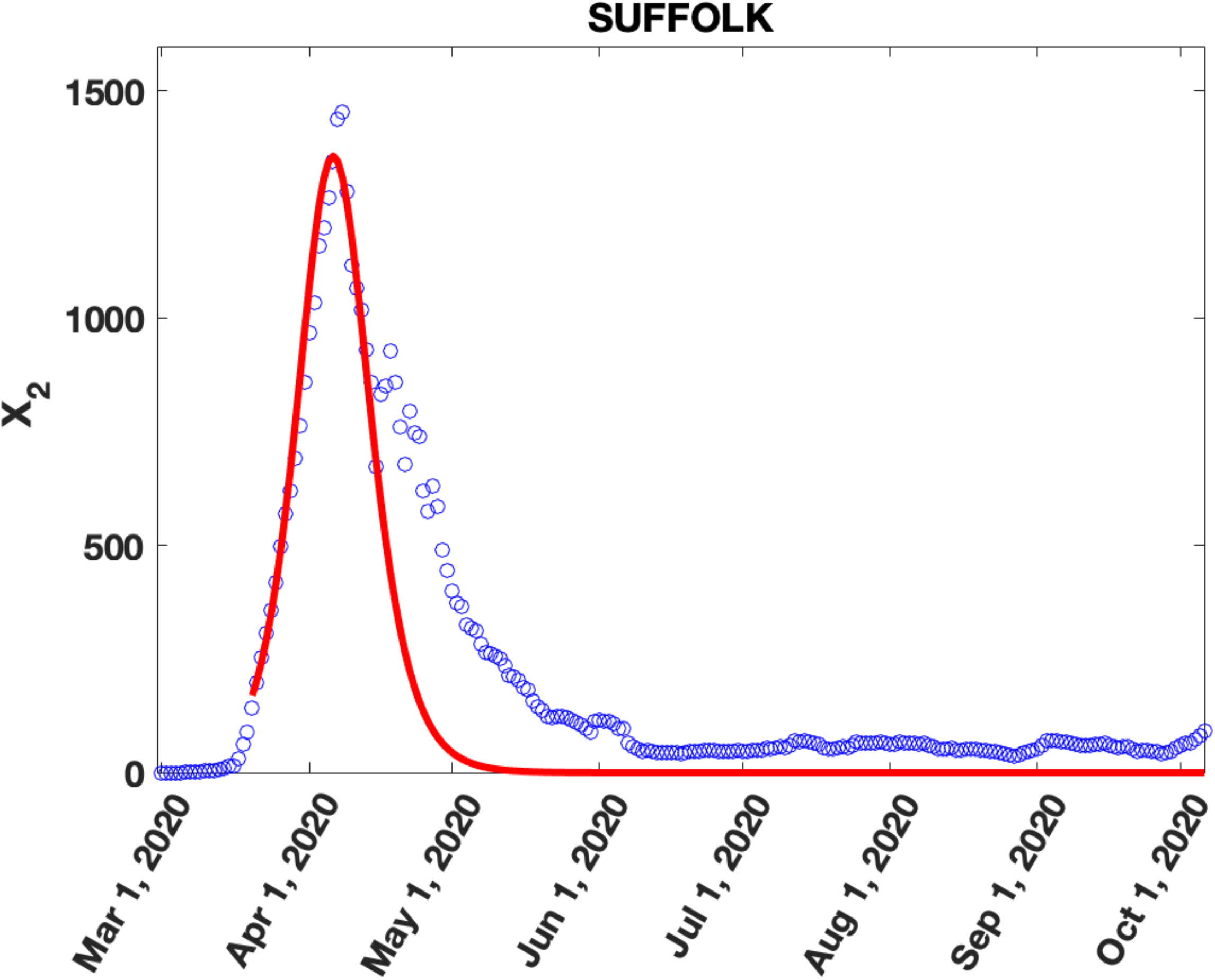
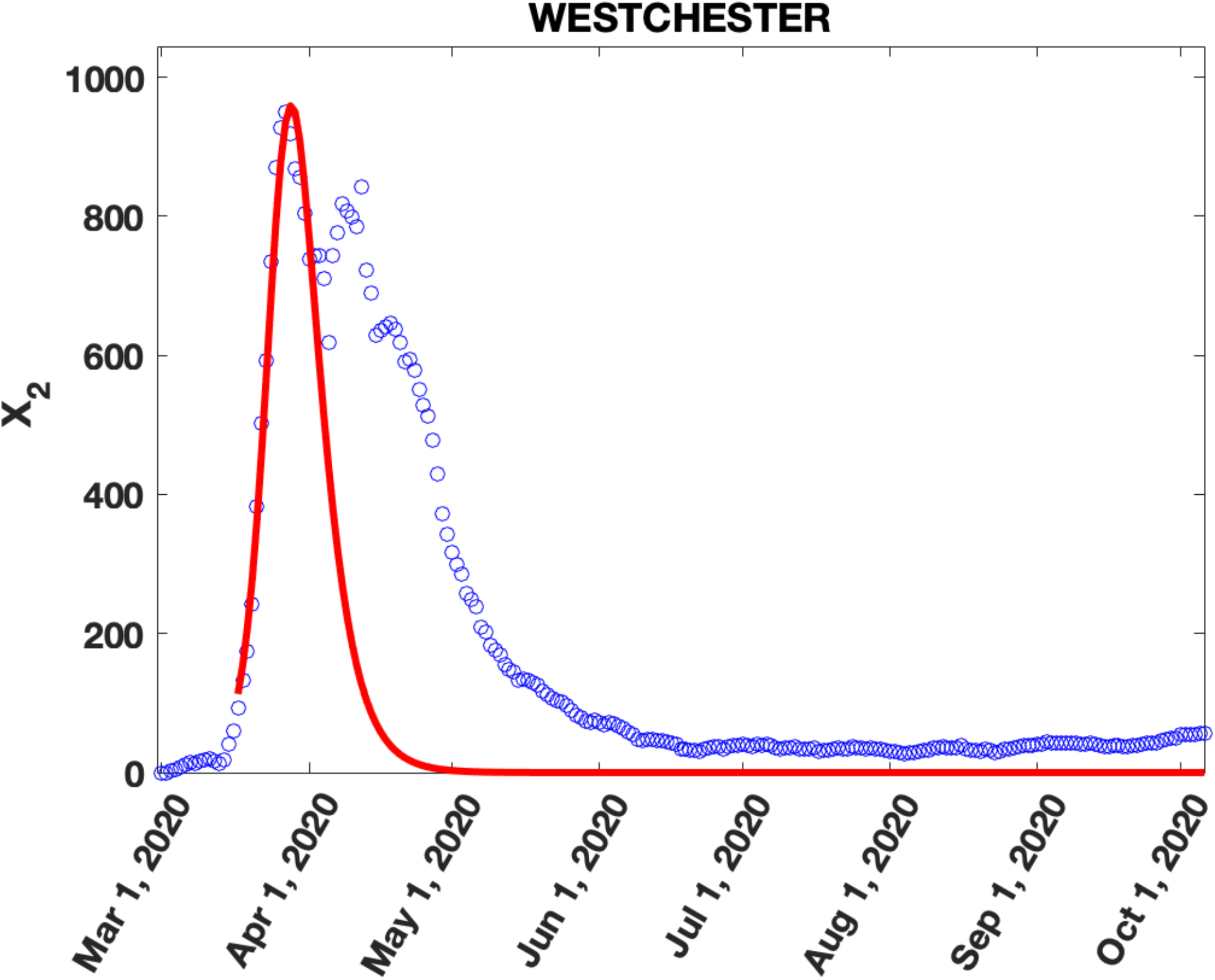
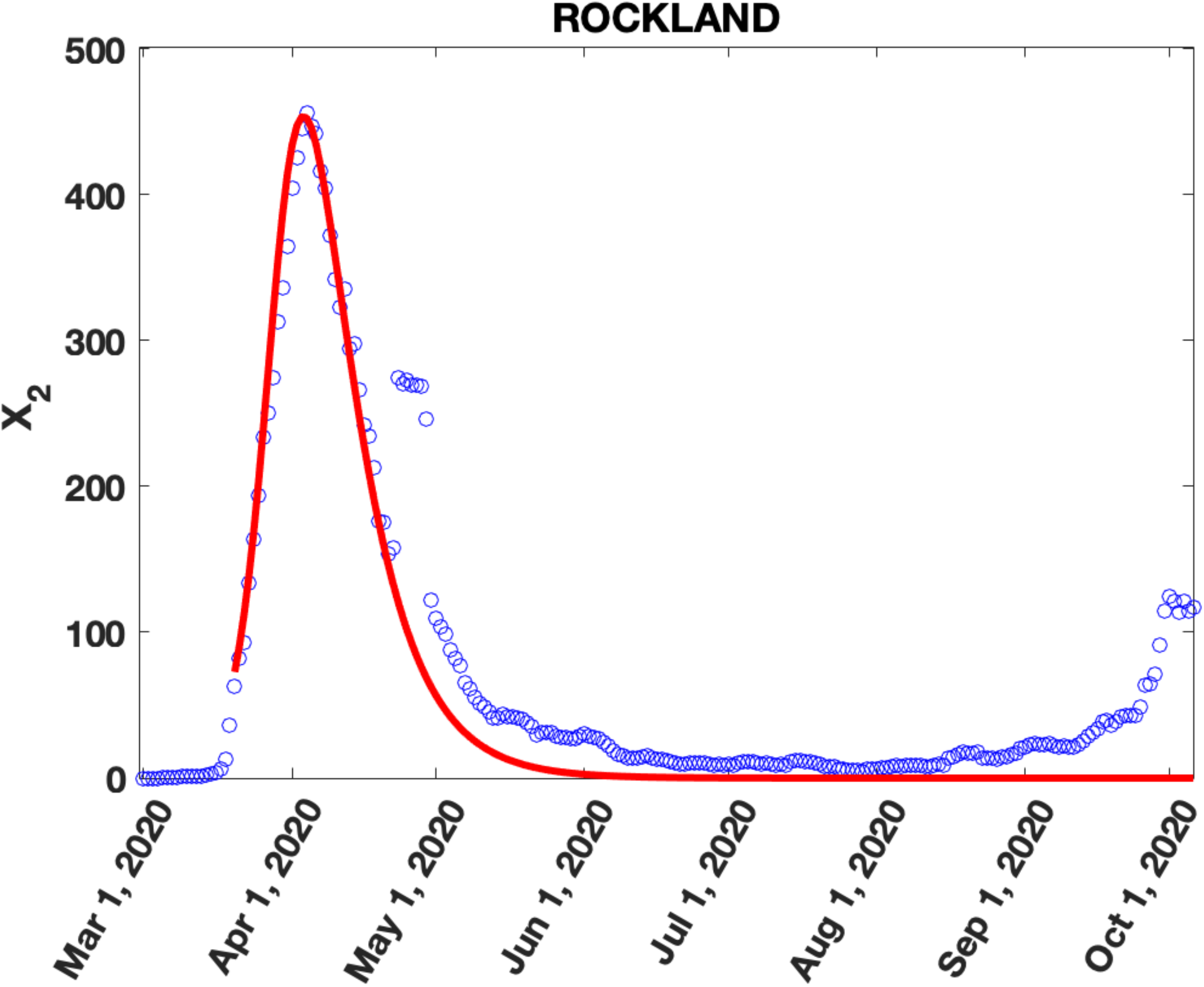
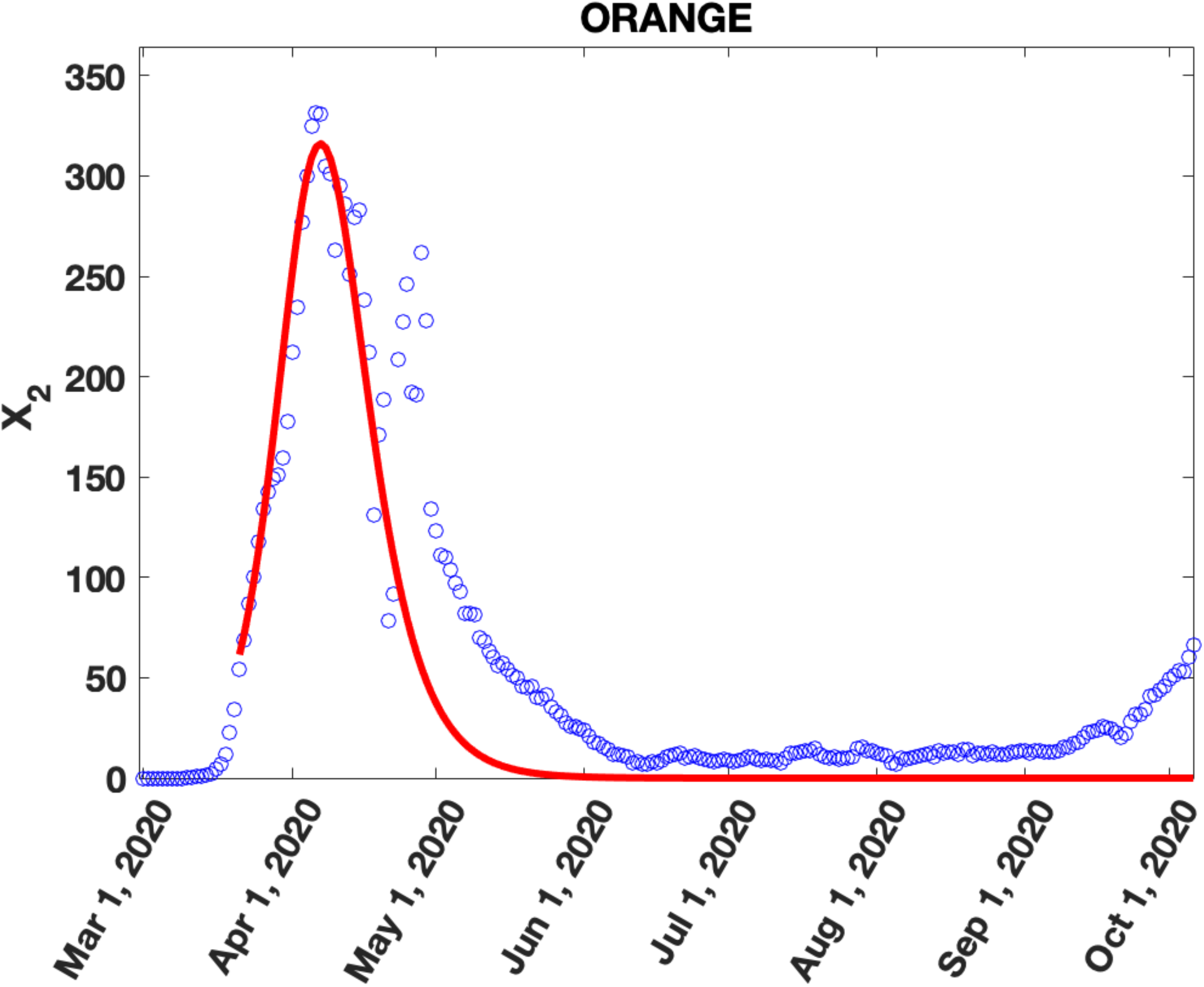
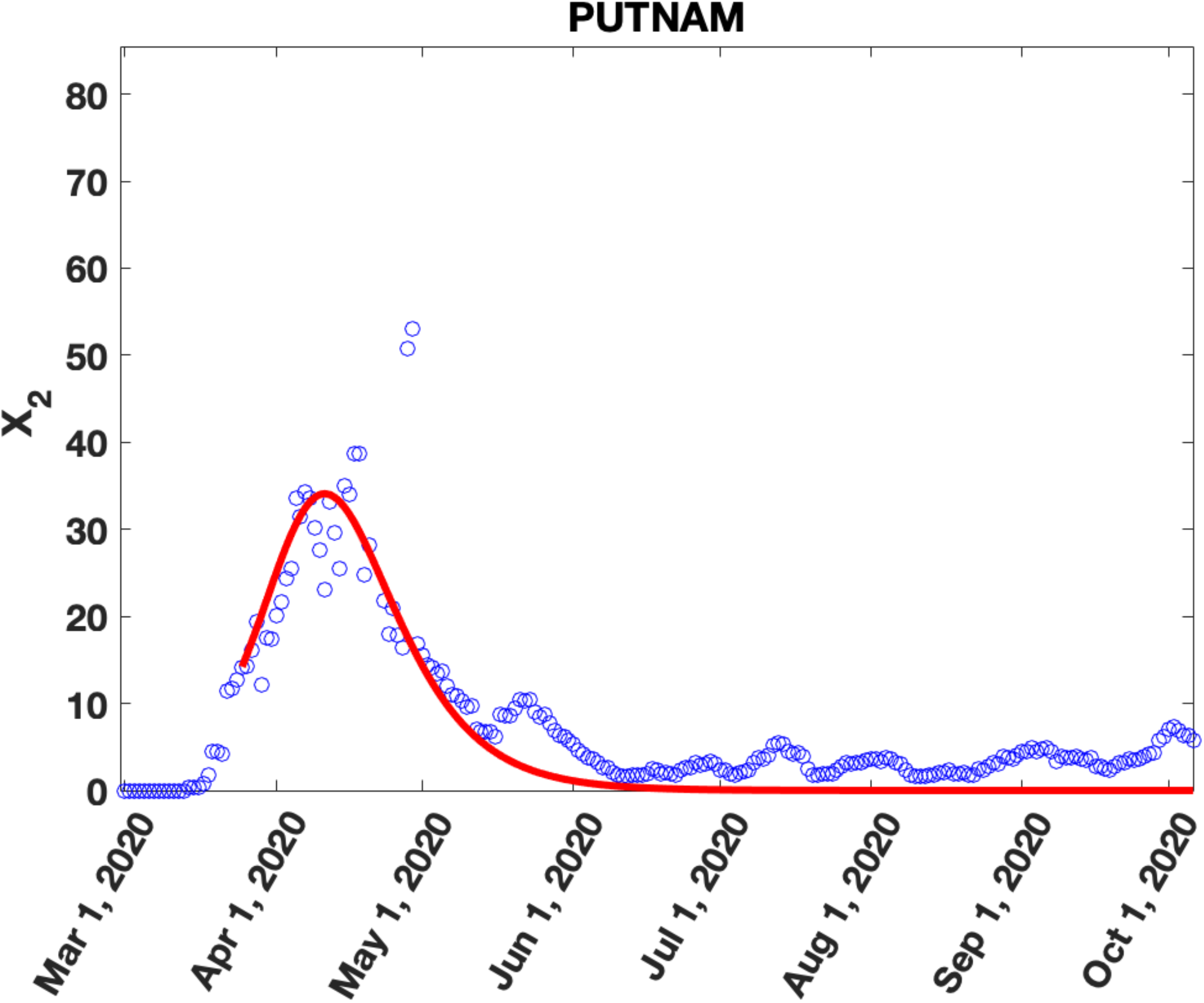
Data and Fits to Covid-19 daily cases: Seven day averaged daily Covid-19 cases (blue circles) and fits (red line) to the first peak using the SIR model in Appendix A: **Figures 4a-f, Figures 4g-i** and **Figures 4j-o** show results for the six most densely populated counties in NJ, three most densely populated counties in CT and six most densely populated counties in NY respectively. In all the counties, the first peak in cases appears in late March to early April 2020 followed by an increase in daily cases over the model predictions from late April-early June 2020 which shows up either as a shoulder or as a distinct peak.

To support our hypothesis, we will show that the number of cases in the second peak, N_CS_, is related to (a) the county population density, P, and (b) the excess CoA, E_CoA_, from NYC into these NJ/CT/NY counties by the relationship 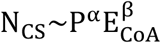. To demonstrate this relationship, we must identify the cases in the second peak in each county, which requires subtracting the cases in the first peak. To do this, we use a previously proposed SIR model (see Appendix A) which was successfully applied to analyze the lockdown/quarantine phase of the Covid-19 daily case load and mortality in eight European countries and the United Kingdom [36]. The model parameters are obtained by fitting the model to the data for the ascending limb (small time exponential increase) and the first peak in daily cases. Using these two as input, the pandemic parameter, R, is estimated by fitting the model to the entire first peak (see Appendix A for details of this model).

The model described in Appendix A was applied to confirmed, seven-day averaged daily Covid-19 case data for each NYC borough and each NJ, CT and NY county (Figure 4, Supplementary Figures 6, 8, 10). Details of the fits for NJ, CT and NYC/NY Counties are given in Supplementary Tables III, IV and V respectively. The solid red curve in Figure 4a-m shows the results of the fits for six counties in NJ, three counties in CT and four counties in NY with the highest population densities. Figure 5 shows the residual daily cases (the second peak) for these counties after subtracting the fits for the first peak (complete results for second peaks are in Supplementary Figures 7, 9, 11) The number of cases in the second peaks for each county was estimated by summing the second peak data (Figure 5, Supplementary Figures 7, 9, 11) between the time points marked with black dots. These time points were determined by starting at the time point with a daily case load near zero in the first peak and using the infection point at the end of the second peak (when the case load tapered off to a constant value) as the location of the second time point.

**Figure 5:**
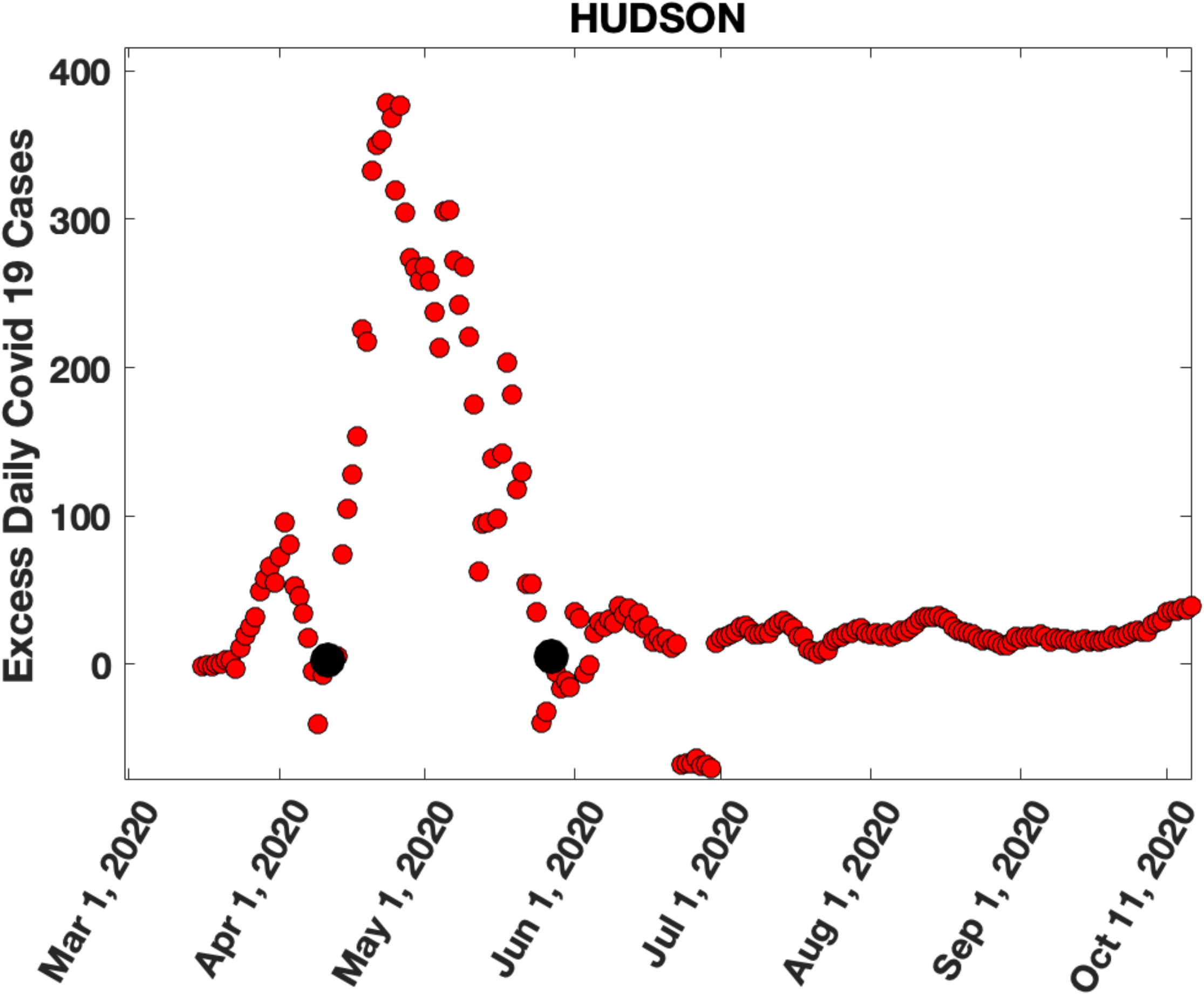
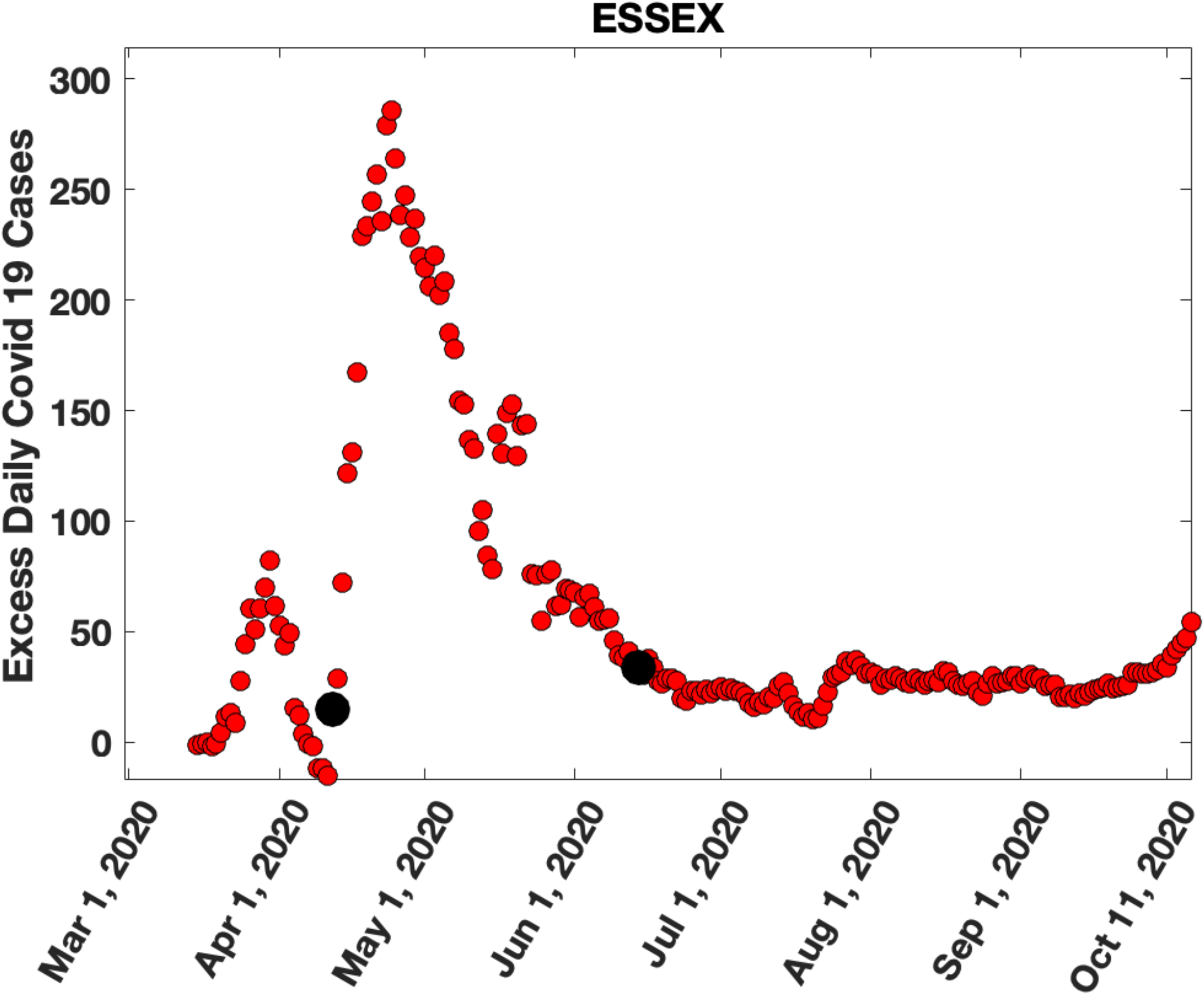
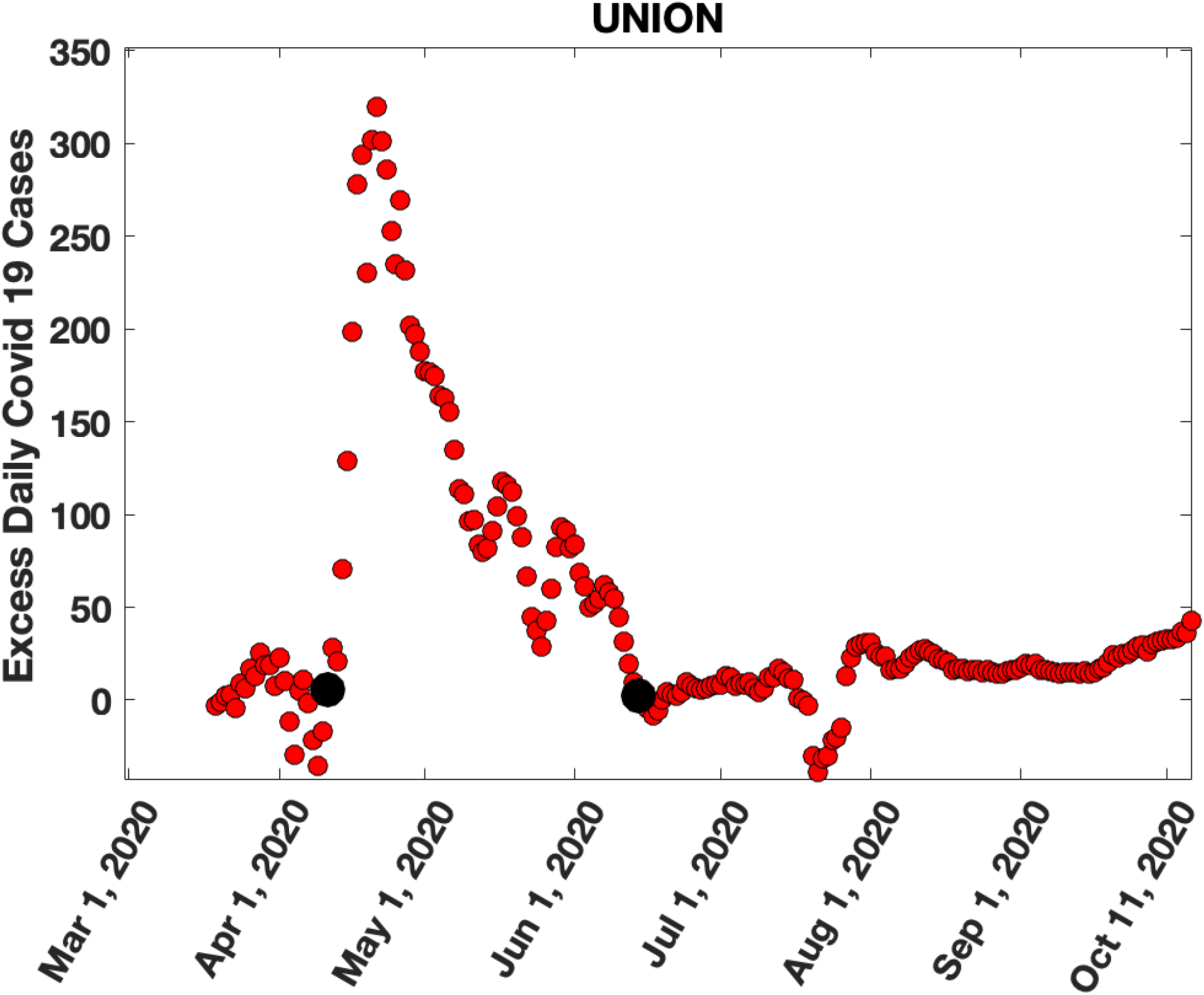
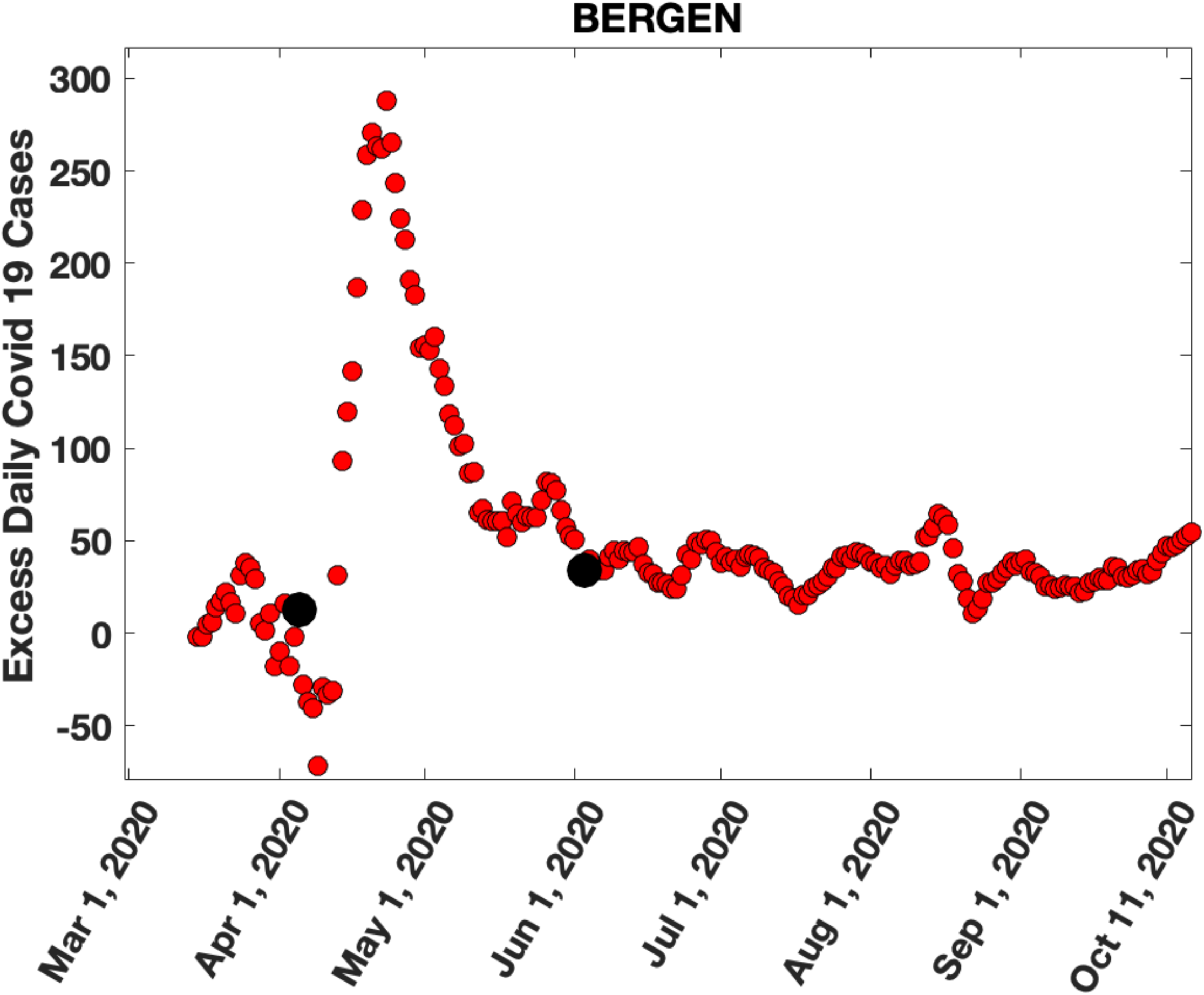
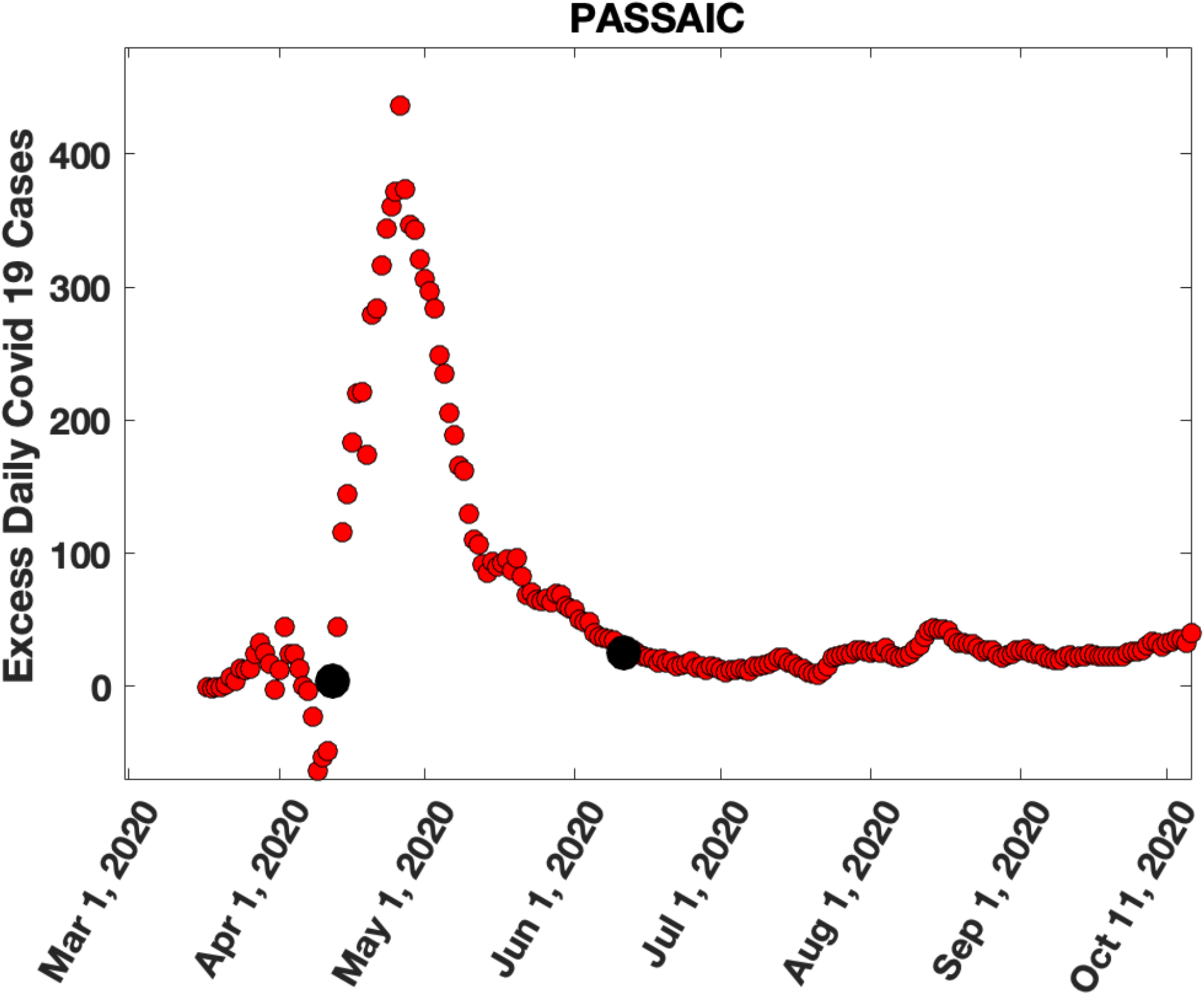
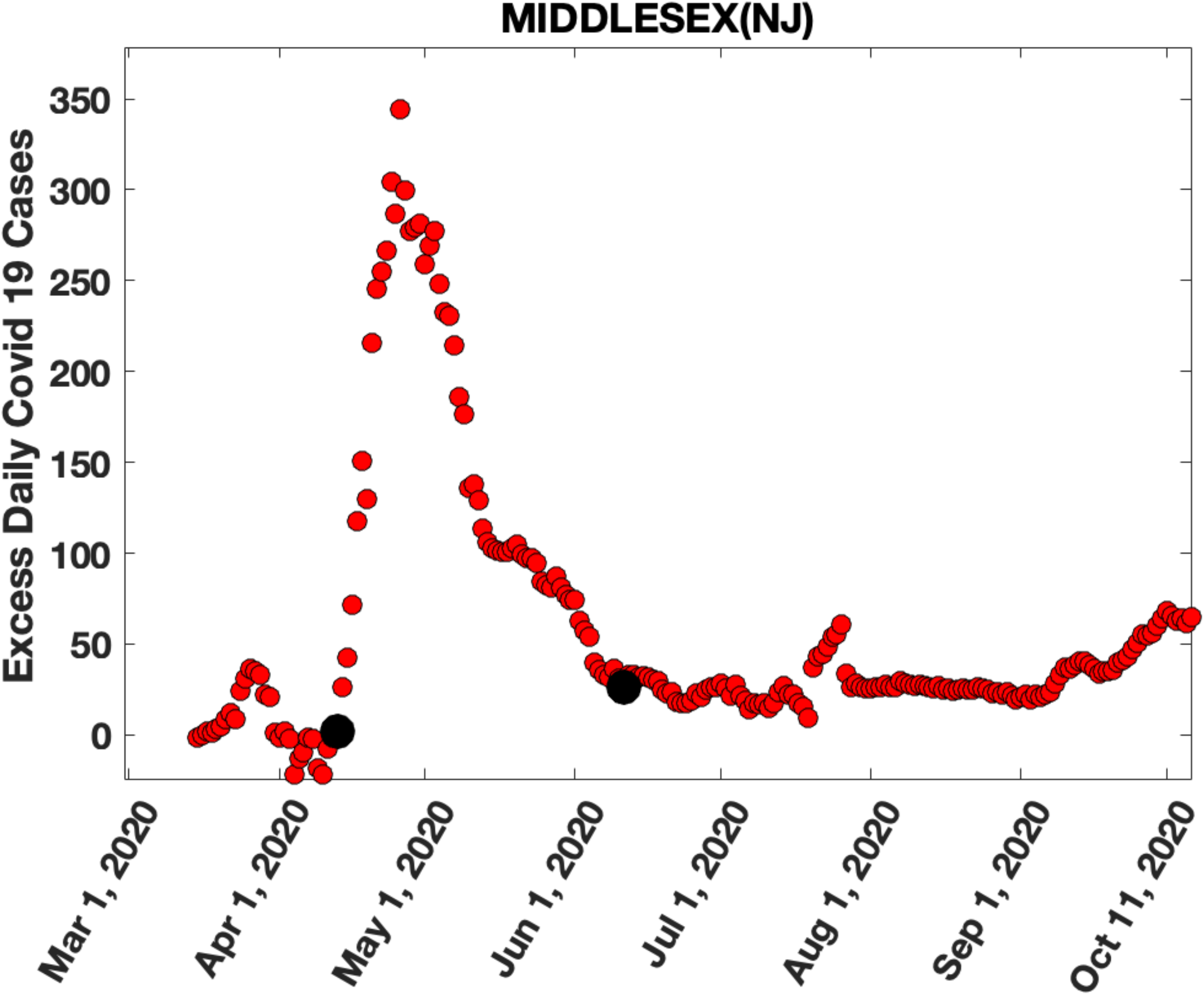
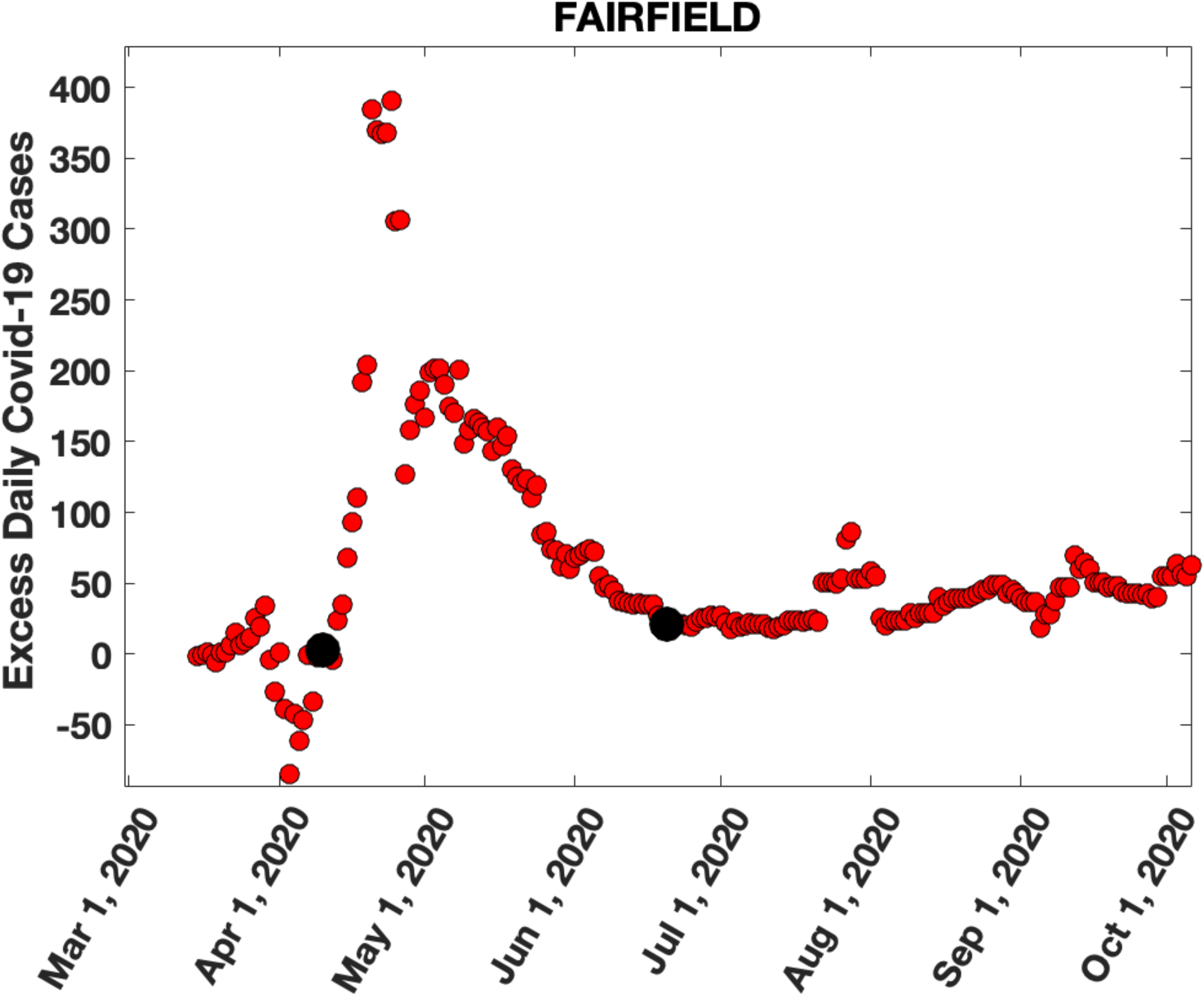
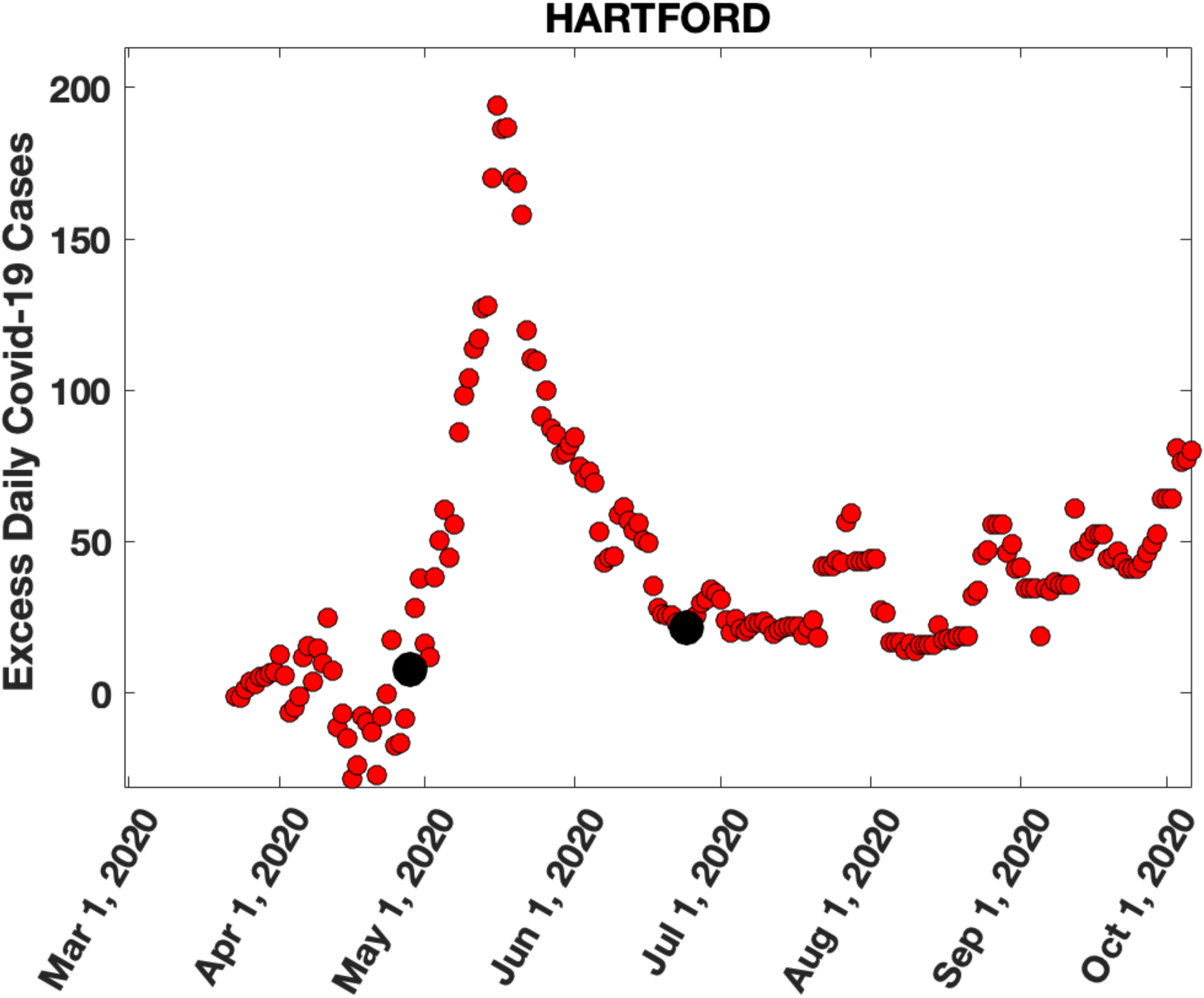
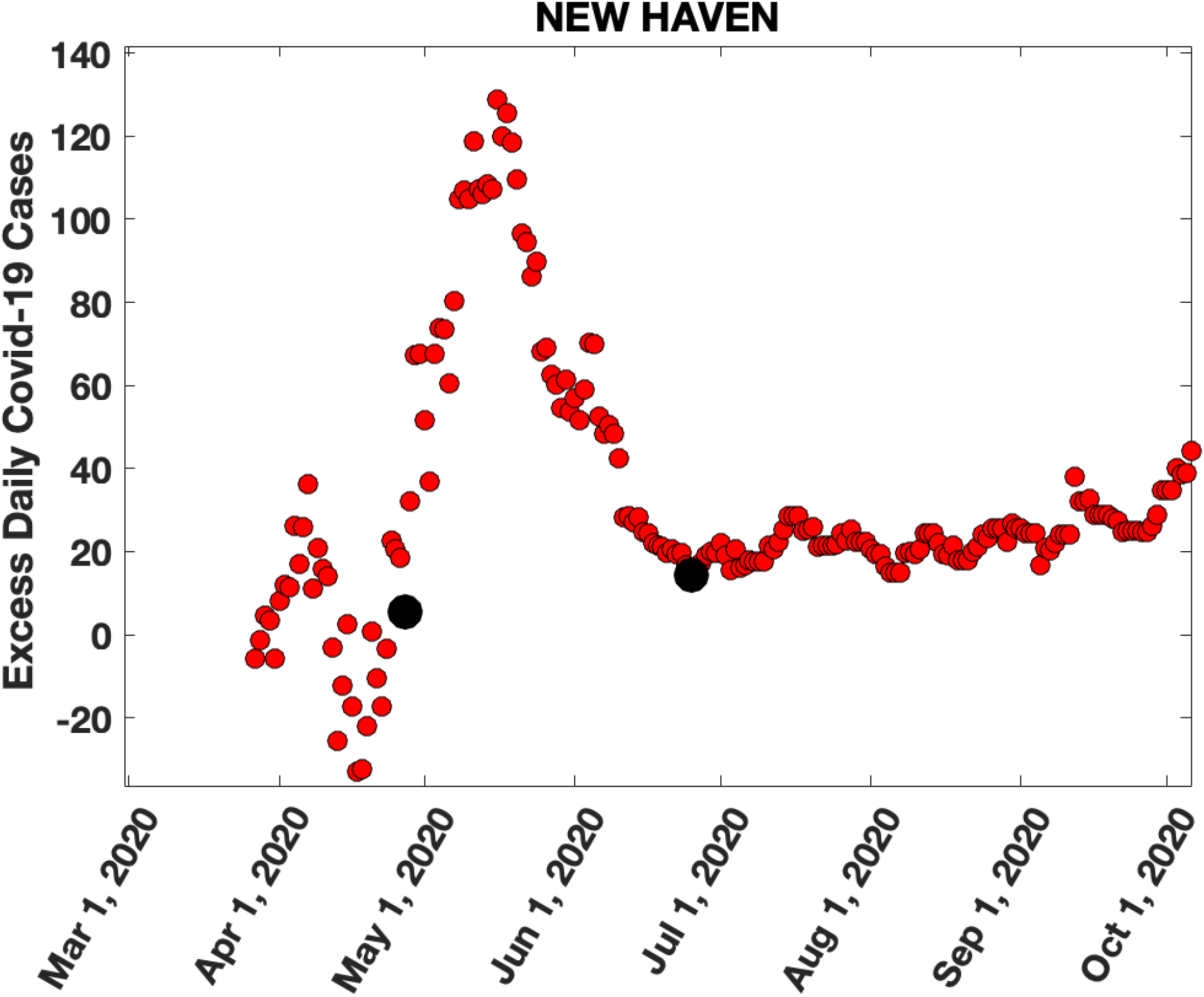
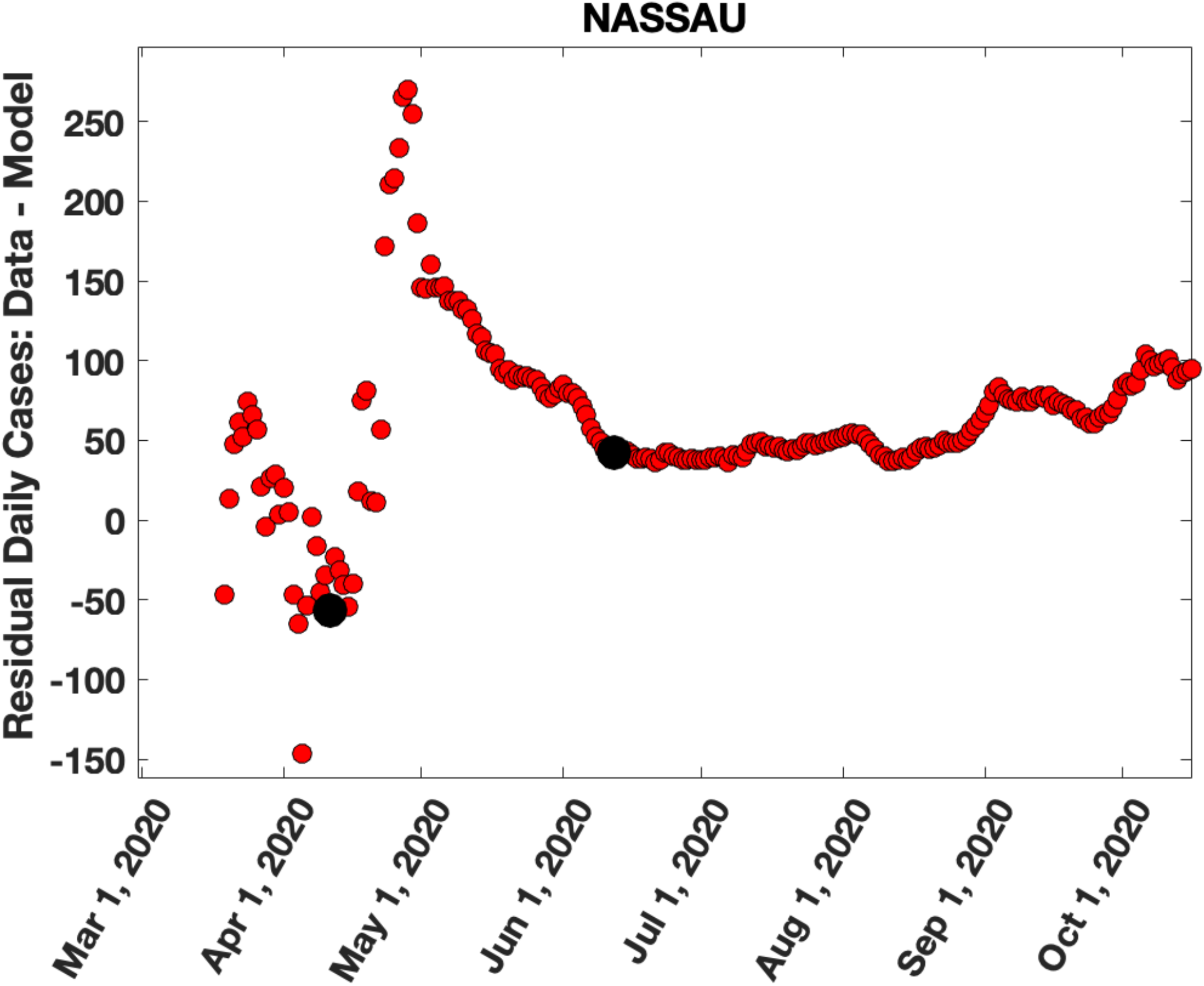
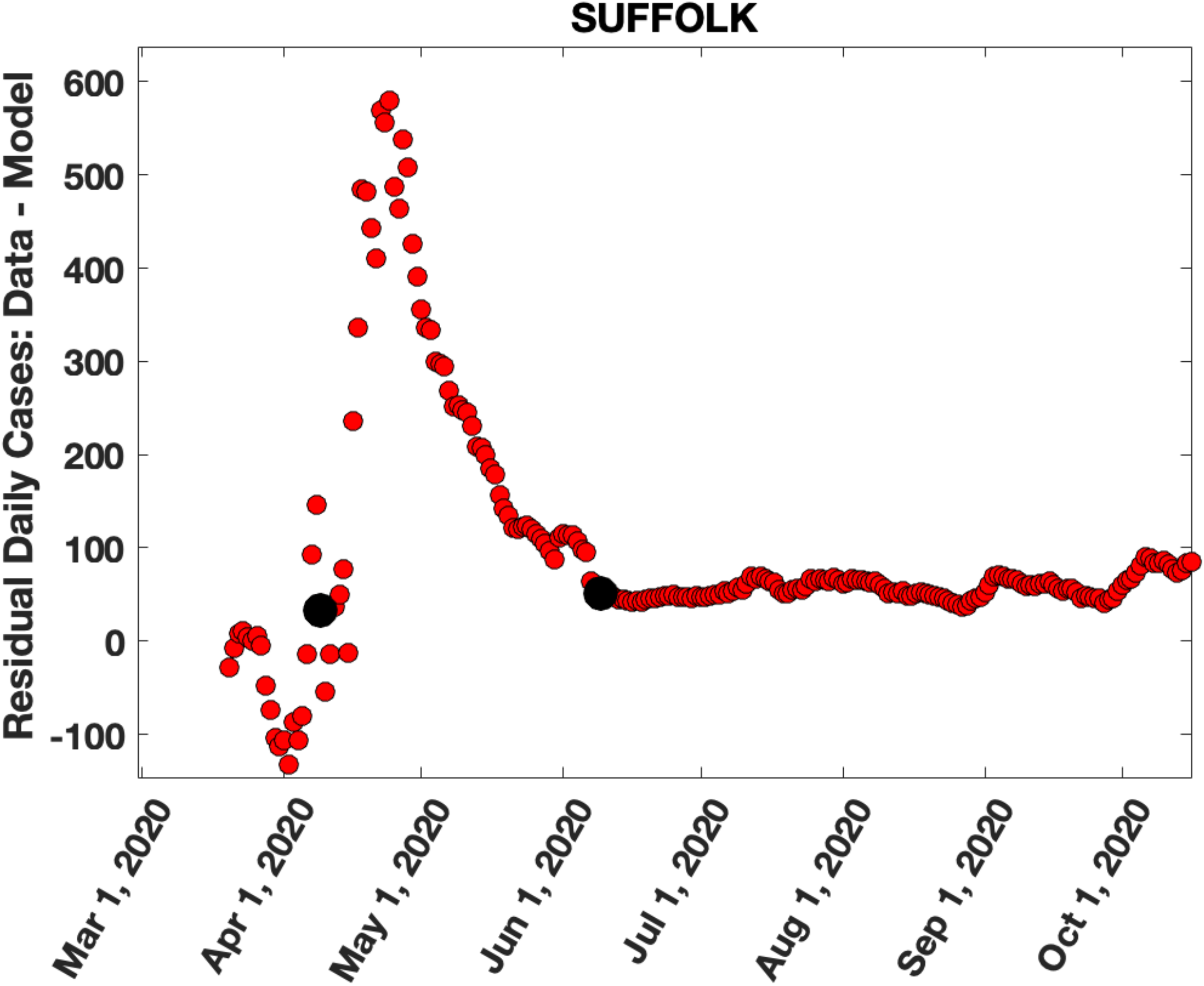
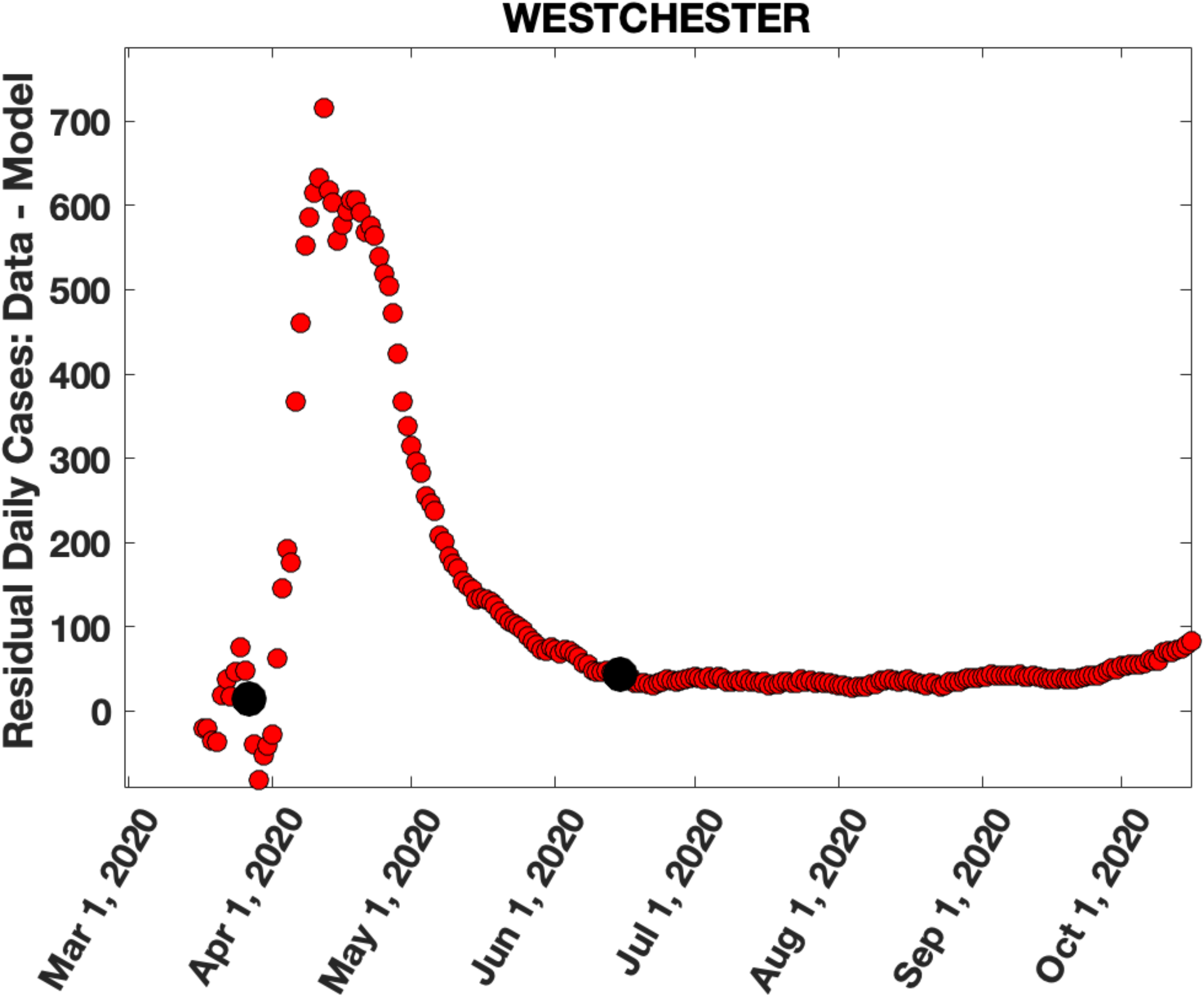
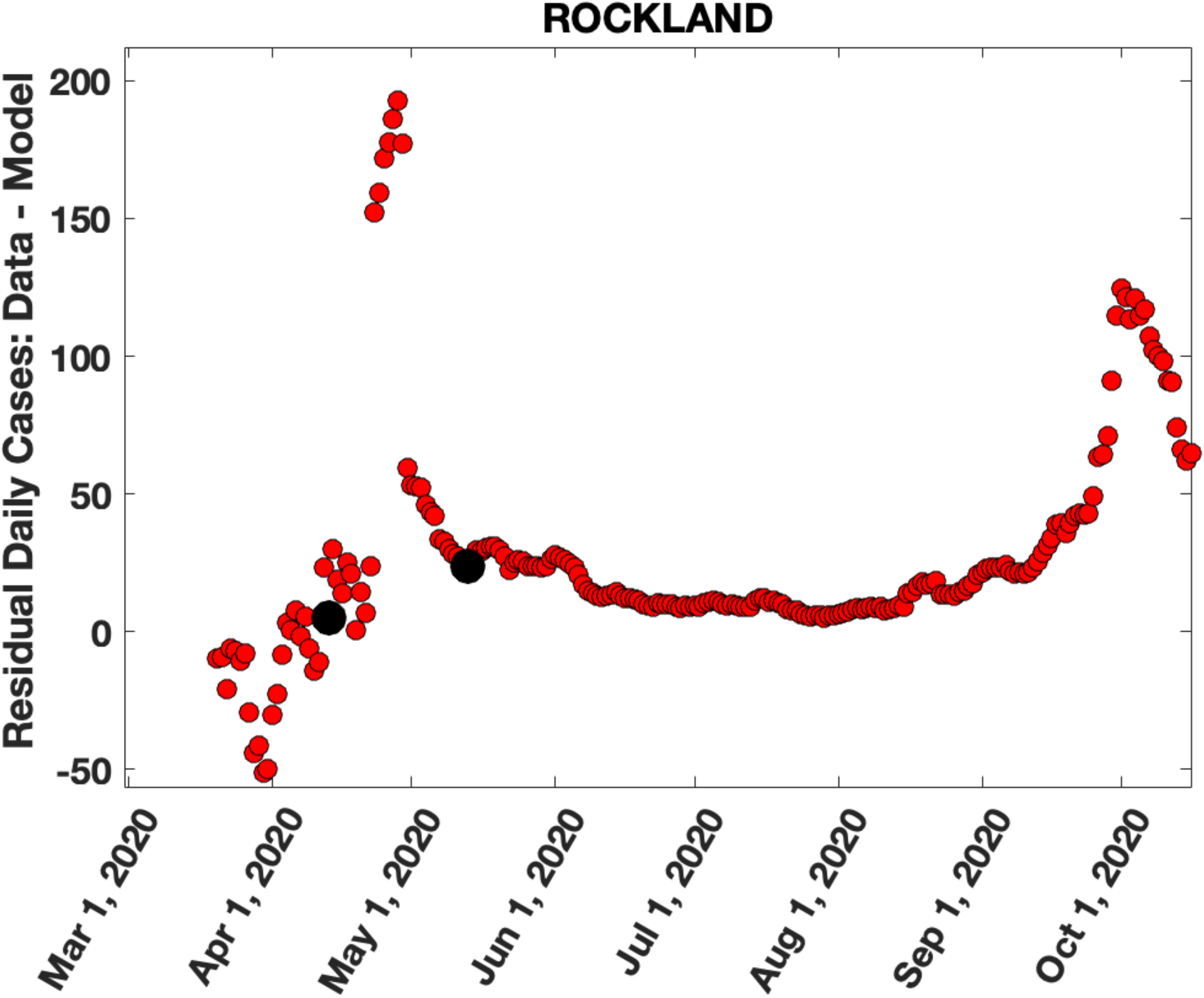
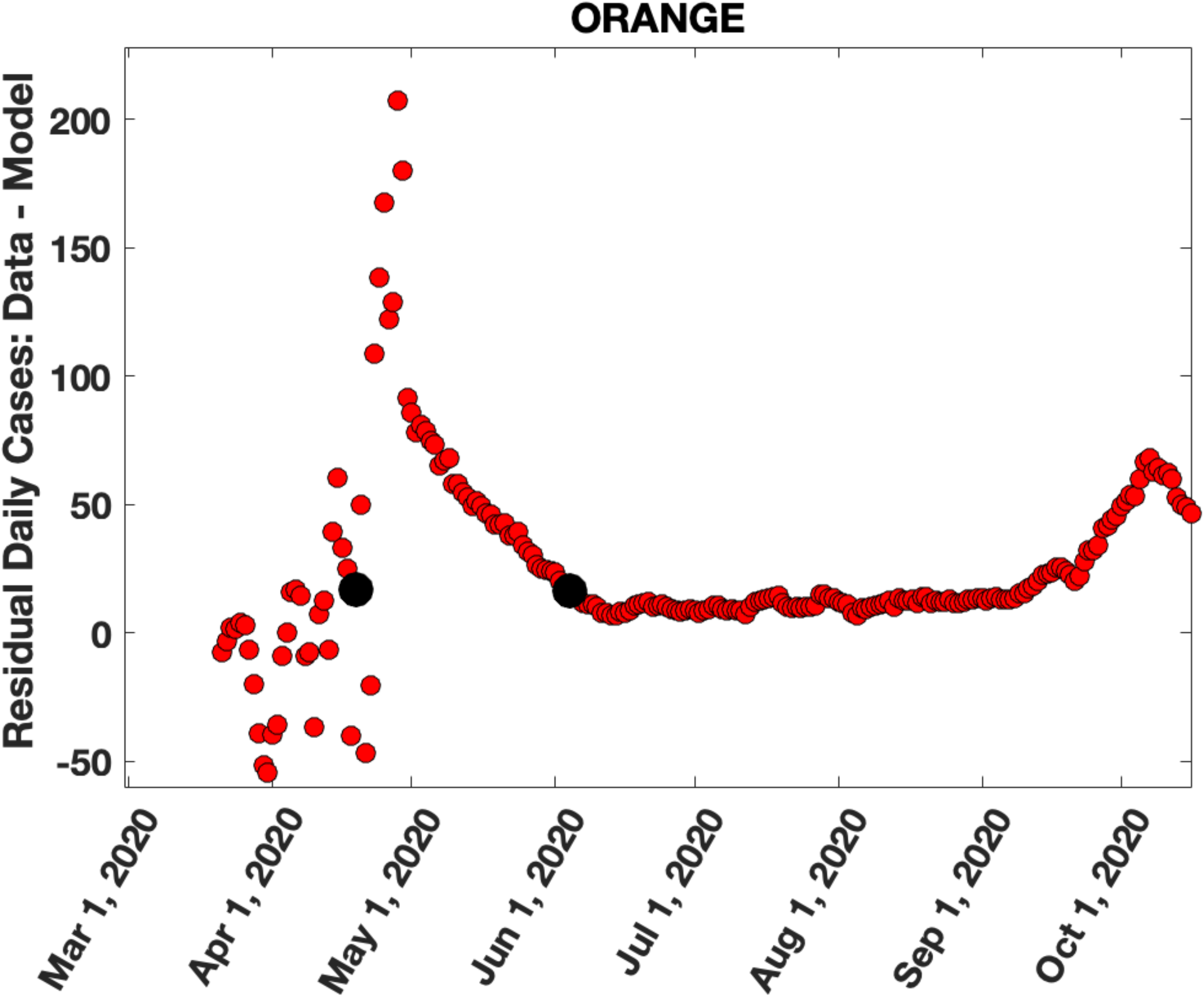
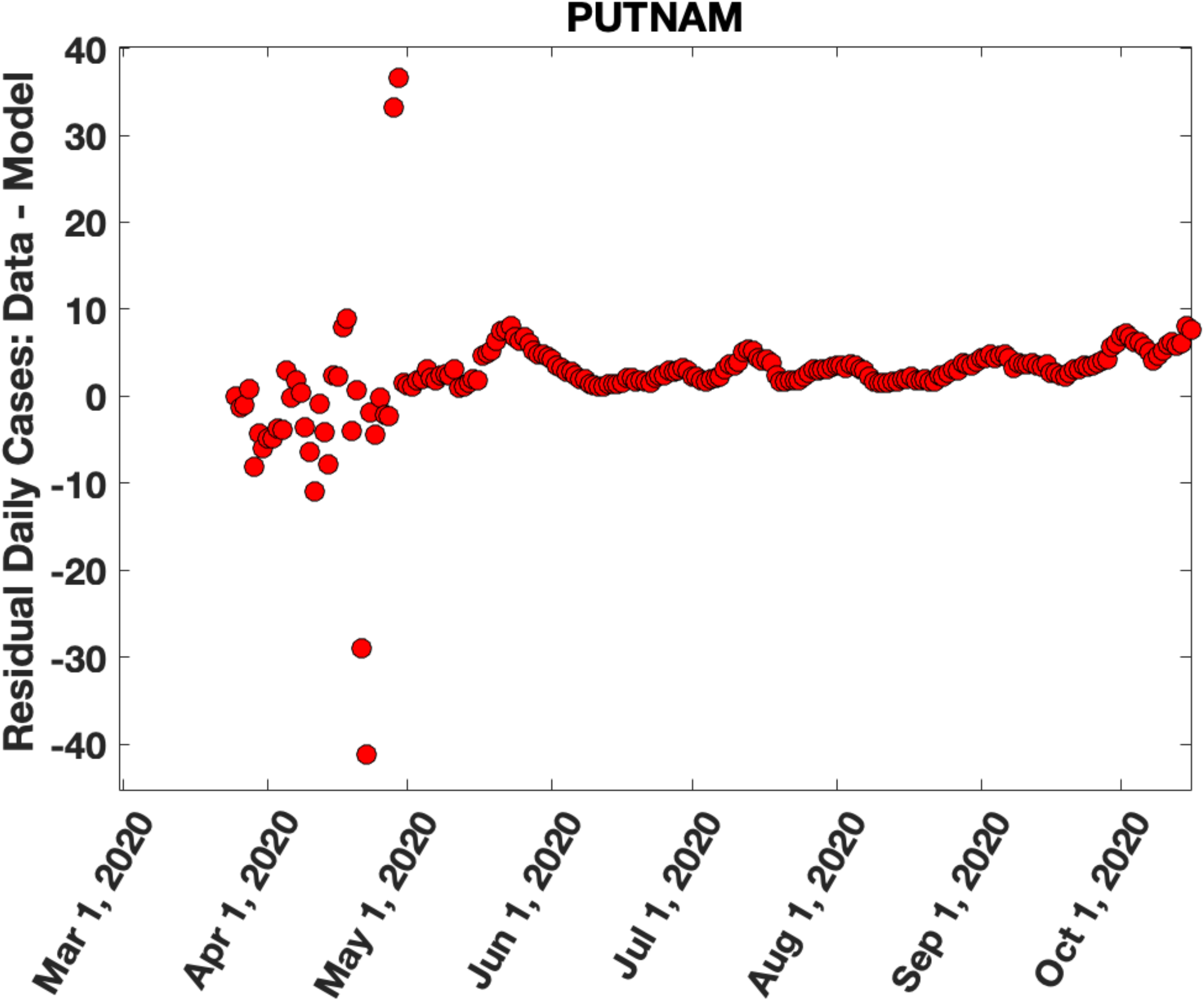
Excess in daily cases over model predictions. These figures show the difference in daily cases between the actual data (blue circles) and the SIR model predictions (red lines) in Figure 4. **Figure 5a-f, Figures 5g-i** and **Figures 5j-o** show the excess in daily cases for the six most densely populated counties in NJ, three most densely populated counties in CT and six most densely populated counties in NY respectively. The black dots show the time points between which we sum the counts to estimate the number of cases in the second peak.

### Relating the number of cases, N_CS_, in the second peak to the county population density, P, and excess CoA from NYC into the county, E_CoA_

The results for the location in time, amplitude and number of cases in the second peak, along with the CoA excess for each NJ/CT/NY county are given in Table I. If migration of households was responsible for the second peak, the cases in the second peak would be due to an interaction between infective people that moved to the county from NYC and uninfected residents in the county, with higher peaks expected in more densely populated counties. We propose the simplest model for this relationship: 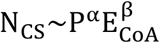 where N_CS_ is the number of cases in the second peak in a given county, P is the county population density and 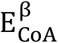 is the excess CoA into the county from NYC (Columns L, F and O in Table I). Taking the log on both sides allows a bivariate linear regression analysis to fit the model to the data. We find an excellent fit of the data to this model (Figure 6a) with α = 0.68 and β = 0.31, R^2^ = 0.74, F = 38.4, p-value = 1.3e-8, Spearman Rank Correlation = 0.88, p-value = 3.9e-7. Figure 6b and 6c show the projection of the fitted data points (crosses) onto the P and E_CoA_ coordinates respectively.

**Table 1:**
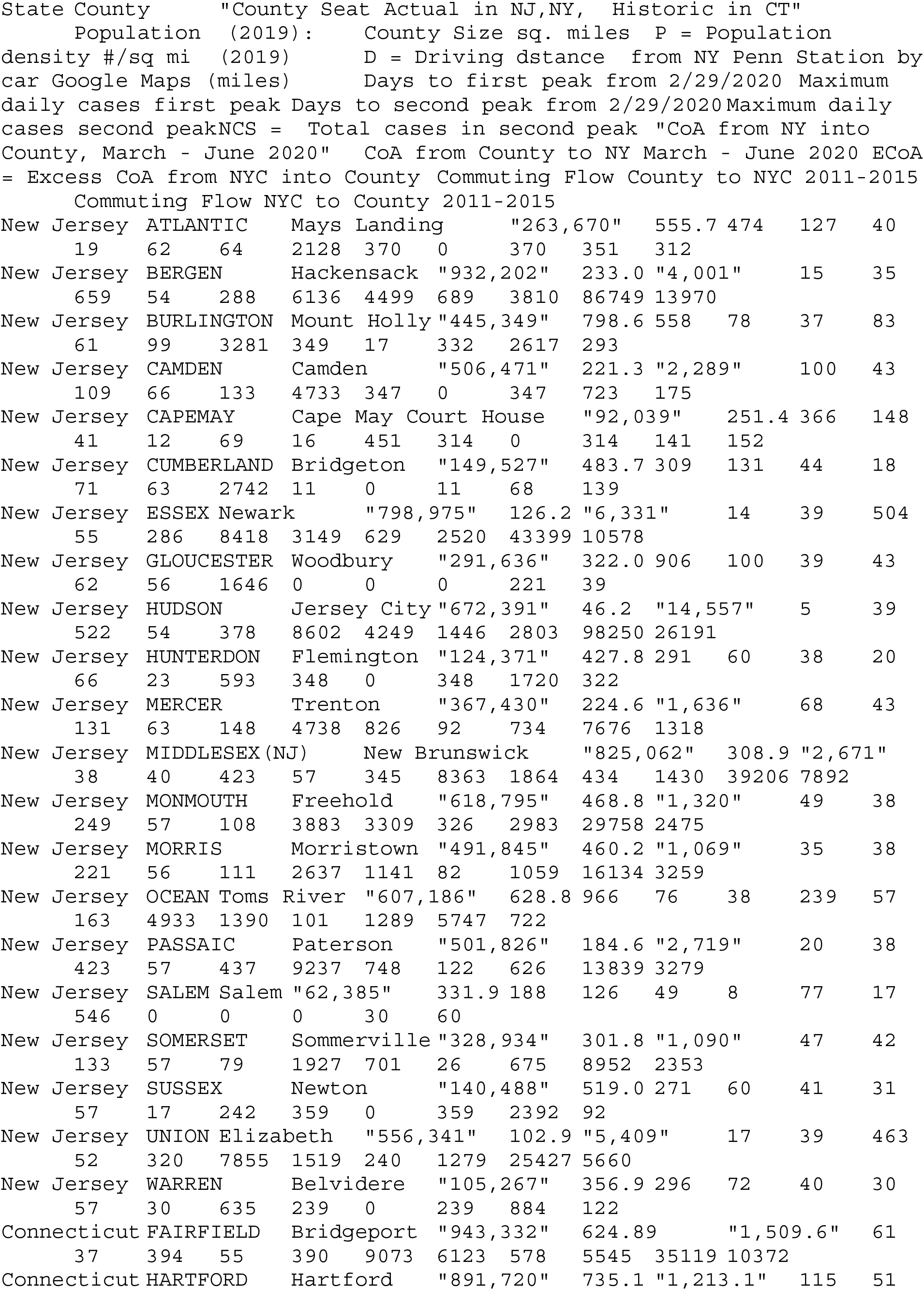
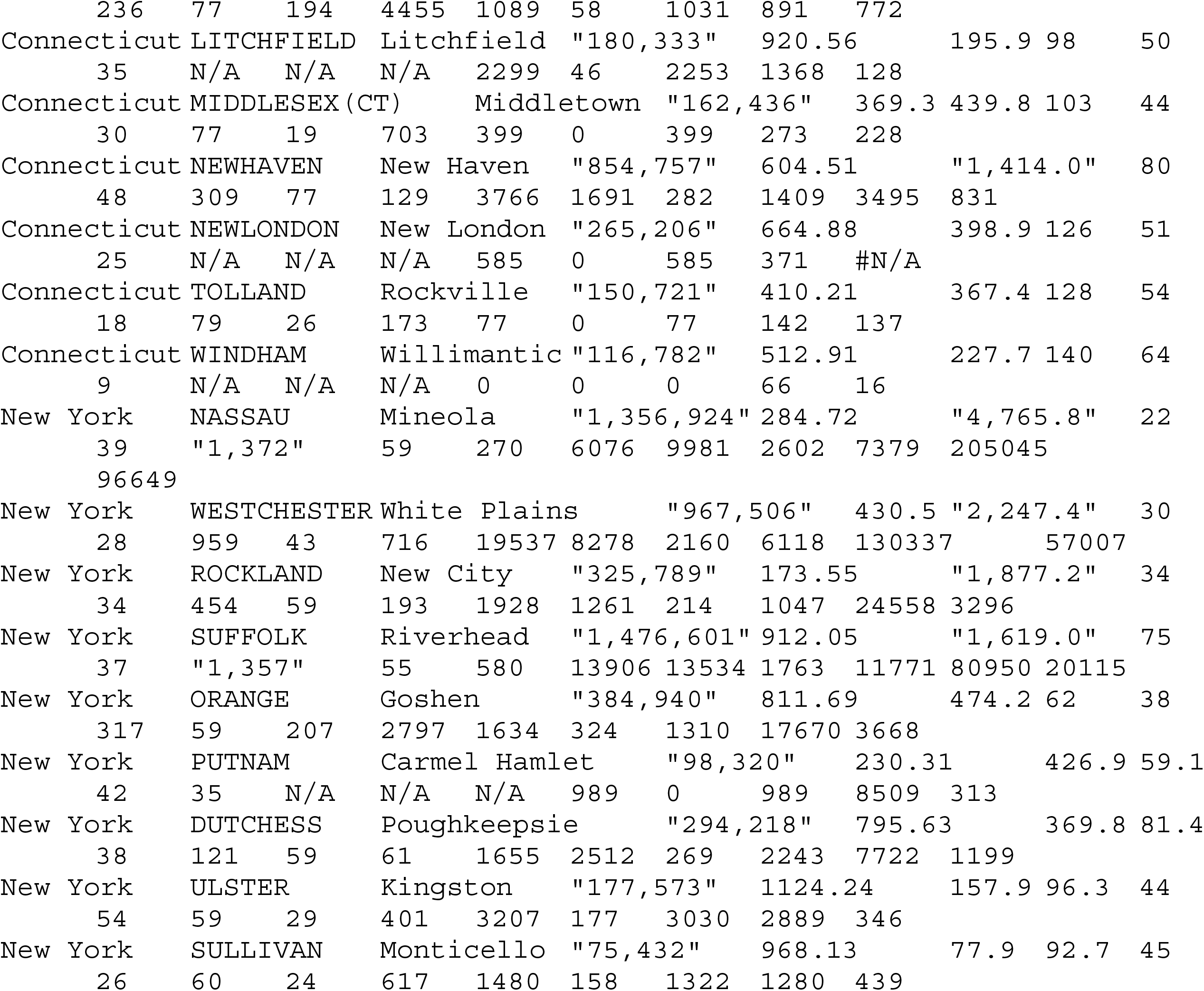
Demographics, statistics about caseloads, predictions from the SIR model, change of address information and commuter flow data for each county in NJ, CT, and the surrounding NYS.

**Figure 6a-c:**
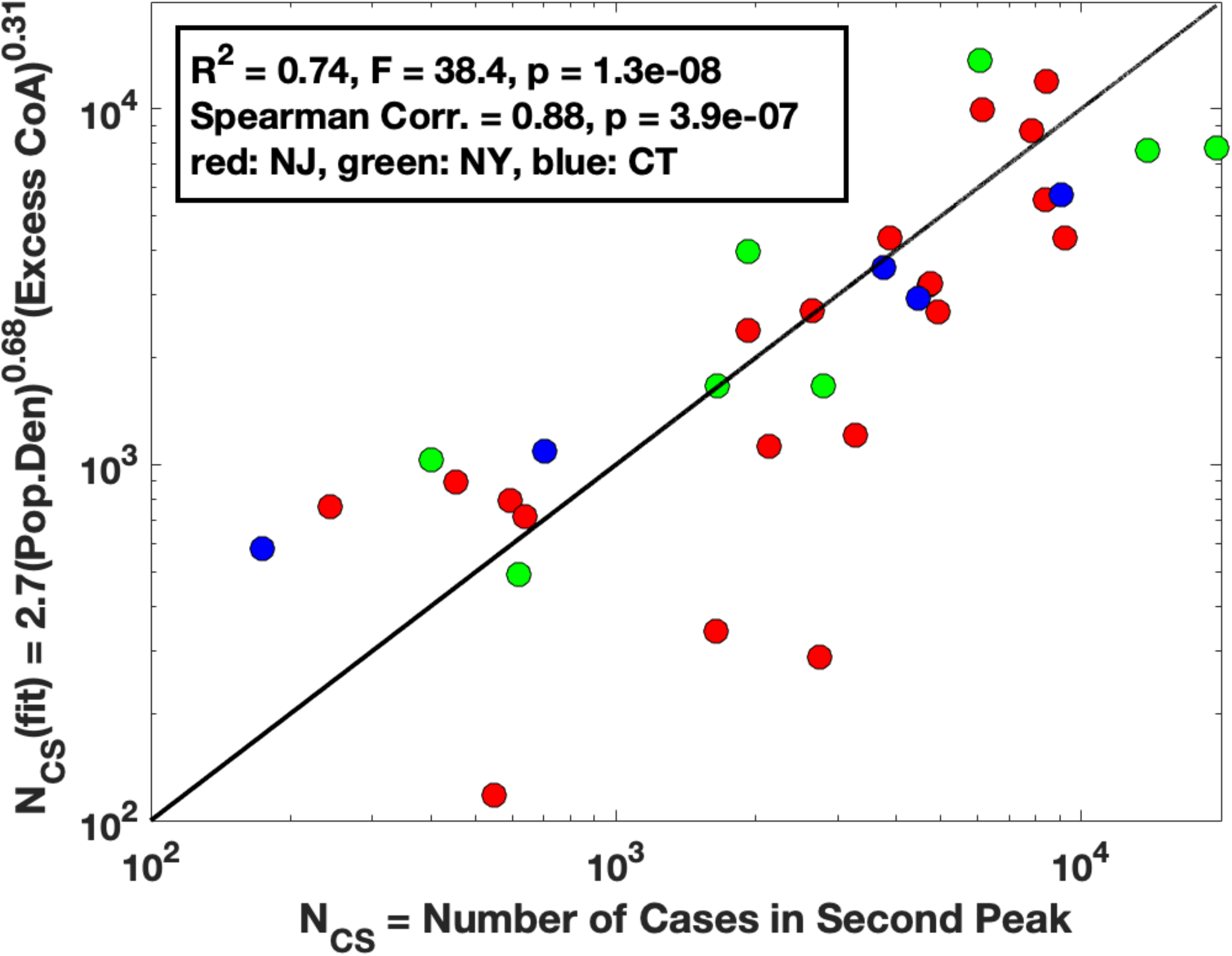
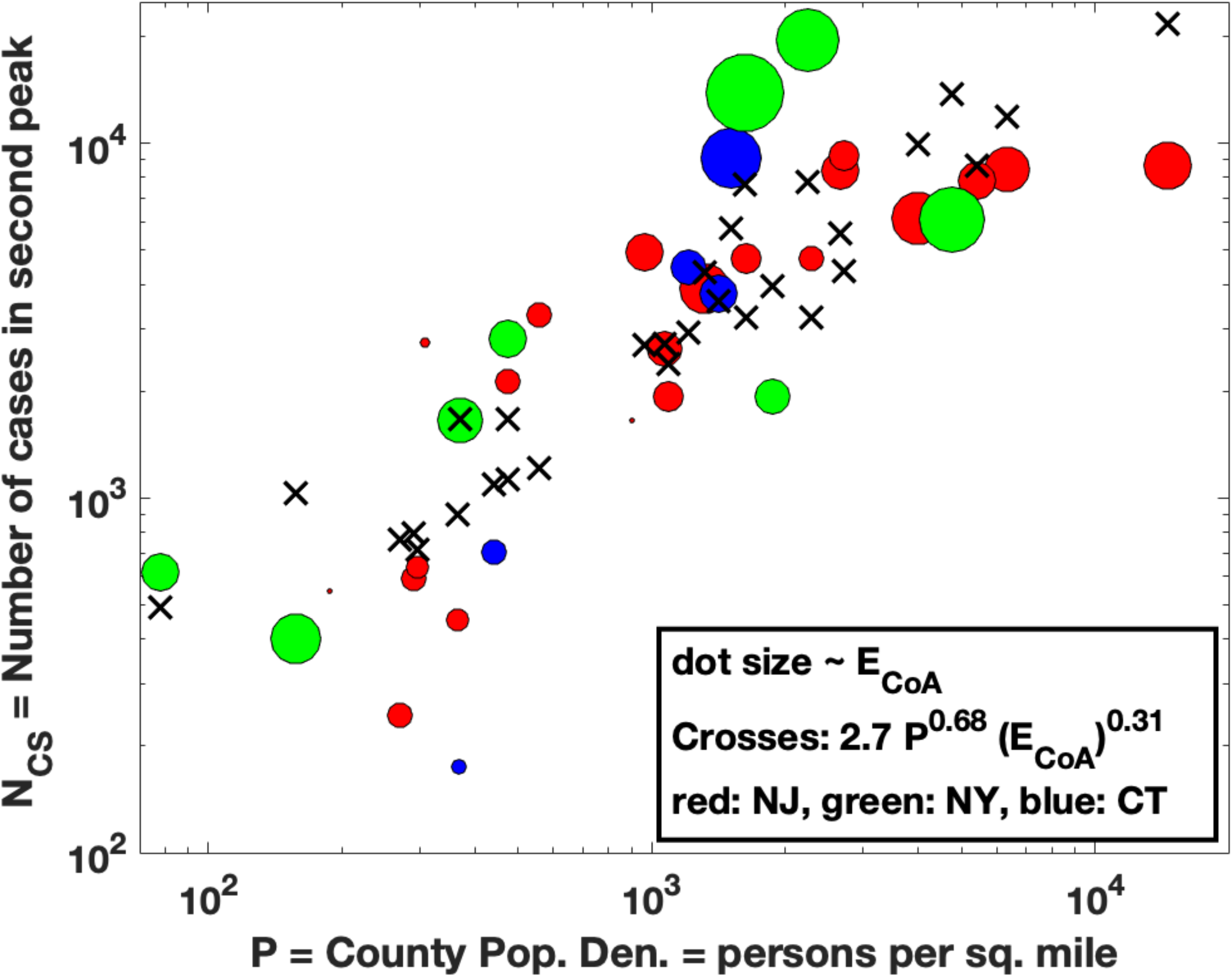
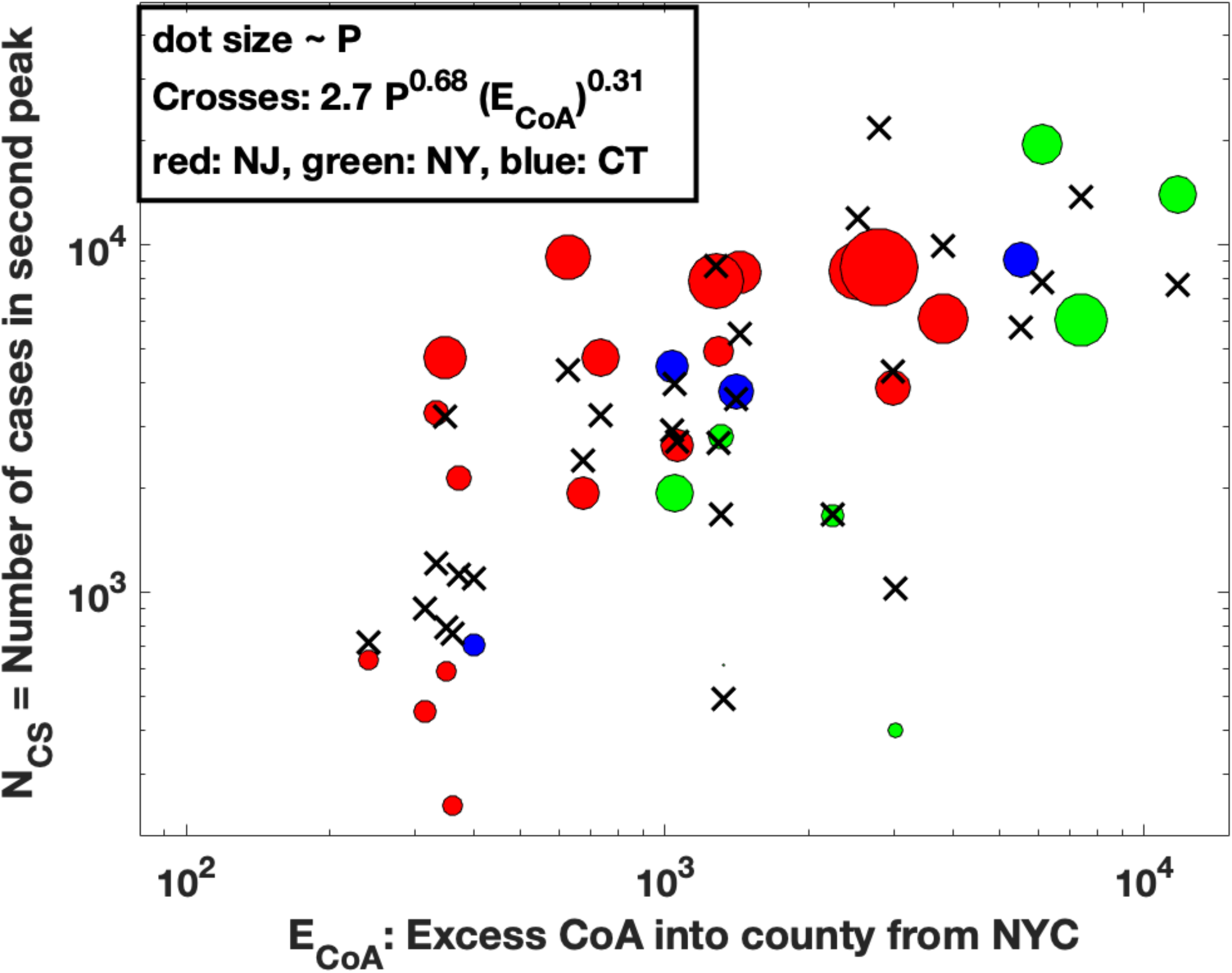
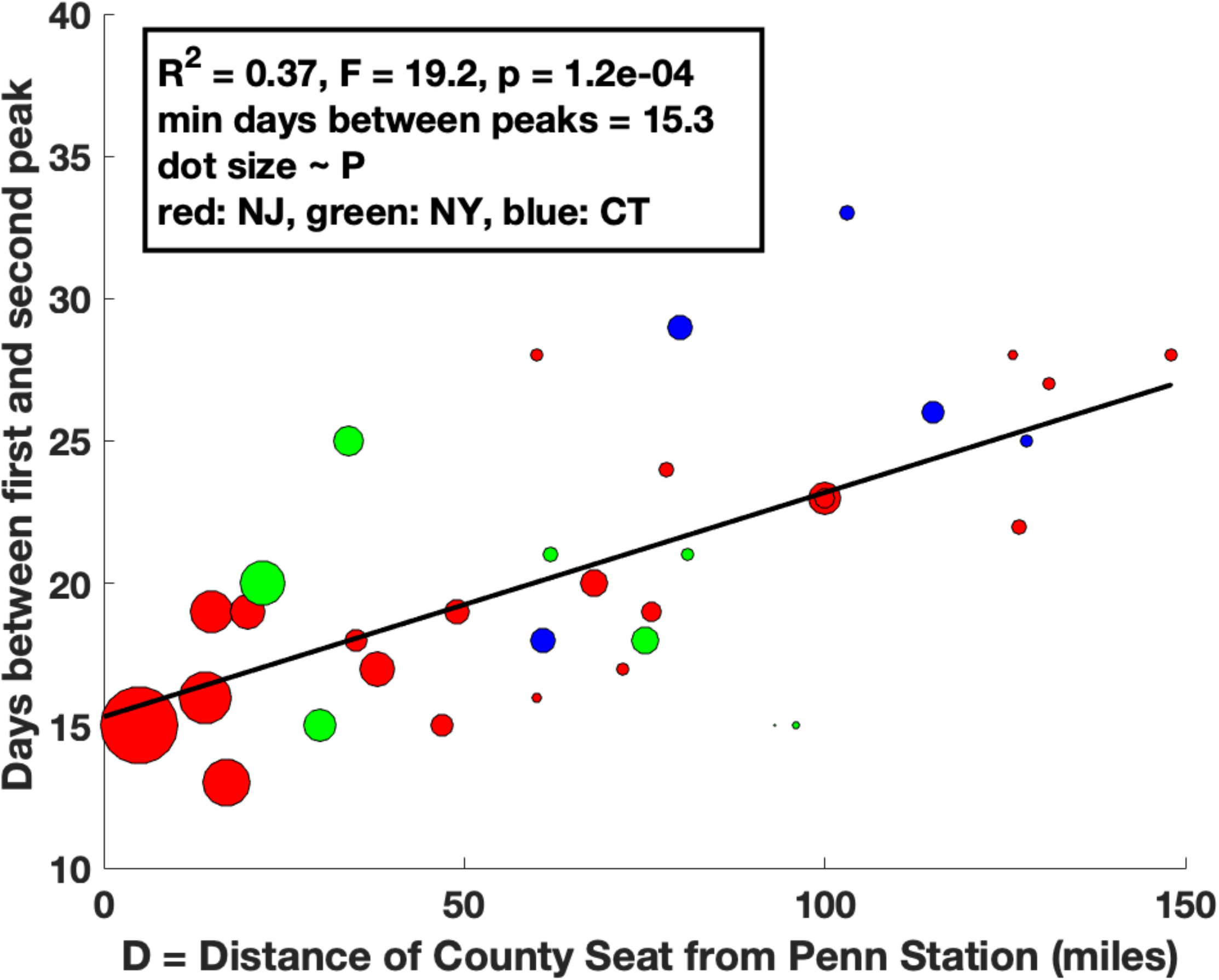
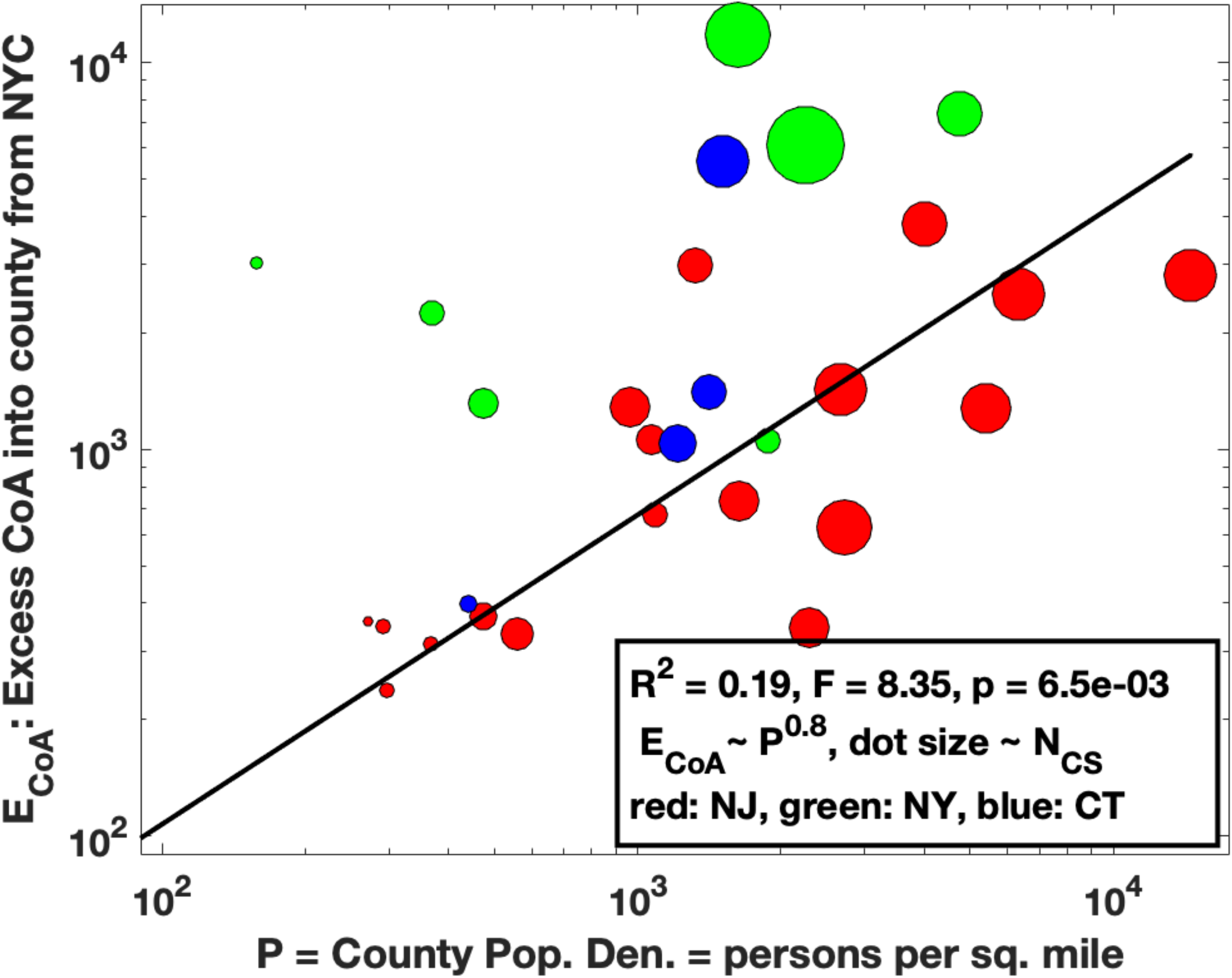
Bivariate linear regression fit to the model: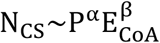 where N_CS_ is the number of cases in the second peak in a given county, P is the population density in the county and E_CoA_ is the excess households that moved into the county from NYC boroughs from March-June 2020. The red, green, blue colors represent NJ, CT and NY counties respectively. **Figure 6a:** The solid line shows the fit to the data (shown as dots) with α = 0.68 and *β* = 0.31, R^2^ = 0.74, F = 38.4, p-value = 1.3e-8, Spearman Rank Correlation = 0.88, p-value = 3.9e-7. **Figure 6b:** Projection of the fit of Figure 6a onto the population density (P) axis. The fitted results are shown as crosses and the actual data as circles, with the dot size proportional to E_CoA_. **Figure 6c:** Projection of the fit of Figure 6a onto the E_CoA_ axis, with the dot size now proportional to P, the county population density. **Figure 6d**: There is a linear relationship between the number of days between the first and second peak in cases and the driving distance from county seat to NY Penn Station. The red, green, blue colors represent NJ, CT and NY counties respectively and dot size is proportional to the population density in the county. The solid line represents the linear least square fit. The minimum time between the peaks (intercept on y-axis) is probably related to the quarantine time of 14 days which was required for move out of NYC during lockdown from March-April 2020. **Figure 6e**: There is a weak relationship between the excess CoA (E_CoA_) and the county population density (P) : E_CoA_~ P^0.8^, R^2^ = 0.19, so that these two variables are not completely independent. The solid line shows the fit to the data. The dot size is proportional to N_CS_, the number of cases in the second peak.

Finally, we find that the number of days from the first peak to the second peak is proportional to the distance of the respective county seat from NY Penn Station by car (Figure 6d, R^2^ = 0.37, Pearson Correlation = 0.61, p-value = 1.2e-04). Since the relationship is approximately linear, this suggests a directed movement and not a diffusion process. This observation lends credence to our hypothesis that the CoA excess likely caused the second peak. Although it is somewhat of a speculation, the intercept of ~15 days on the y-axis in Figure 6d may be because of the 14-day quarantine imposed on people moving to new locations in the early days of the pandemic.

Finally, as one might expect, the excess CoA is roughly proportional to the county population density (Figure 6e), so that these two variables are not independent. This makes sense because if the county population density P is low, there are very few cases in the second peak (Figure 6b). And likewise, for sufficiently low excess CoA, there are again only a few cases in the second peak (Figure 6c).

## DISCUSSION

Using household “Change of Address” data (CoA) from the United States Postal Service we have shown that there was a significant exodus of people from NYC boroughs to the surrounding counties in NJ, CT and NY, starting from the beginning of the coronavirus pandemic in March 2020 and extending through the lifting of regulations in May-June 2020 and beyond (Figures 1–3, Supplementary Figures 2-5). The majority of household moves to these counties were from New York City (Manhattan) (Supplementary Figures 2,3,4).

Our hypothesis is that this exodus of households into the counties surrounding NYC was partly responsible for second peaks in daily Covid-19 cases in these counties in May 2020. To test this hypothesis, we needed to identify the second peak in daily cases, both in time and amplitude. This is confounded by the presence of the first peak in daily cases during the initial phase of the pandemic in March-April 2020. Although the second peak in cases in each county is clearly visible in most counties (Figure 4, Supplementary Figures 6,8,10), we must use a model to subtract the cases in the first peak to estimate the number of cases in the second peak. To do this, we used a previously proposed SIR model (Appendix A), which accurately captured the initial pandemic dynamic (solid red lines in Figure 4, Supplementary Figures 6, 8, 10), from the early exponential rise in daily cases in March 2020 to the peak in early/mid-April 2020 and subsequent decrease in late April 2020, when the effects of quarantine and lockdown materialized. Although we could have fit the daily case data to some standard functional forms (Gaussian, Poisson, Negative Binomial, Beta function etc.), there is little justification for any of them, whereas our model not only captures the expected dynamics of the first peak but also fits the data very well. Subtracting the model fit to the first peak in daily cases clearly identified the daily cases in the second peak in each county (Figure 5, Supplementary Figures 7, 9, 11).

Table I summarizes our findings. We analyzed the data in Table I under the expectation that the number of cases, N_CS_, in the second Covid-19 peak should be related to the county population density P and the Excess CoA, E_CoA_, into the county from NYC. A bi-variate regression analysis showed that these were related by: 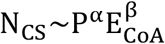 with α = 0.68 and *β* = 0.31, R^2^ = 0.74, F = 38.4, p-value = 1.3e-8, Spearman Rank Correlation = 0.88, p-value = 3.9e-7 (Fig6a, 6b, 6c). A possible interpretation of the exponents in the fit would be that from March-June 2020, the proportion of individuals in each county was ~ P^0.68^ and the number of infected/infective individuals that moved into each county was ~ (E_CoA_)^0.31^. We have also tried other types of fits to the data, which turn out not to be as good as the one we report. For example, both the number of cases in the second peak and the CoA could be normalized by either the population or the population density in the county and then one could try a fit of the form: 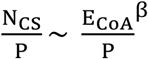 which becomes a linear relationship on taking logs. However, this fit has an R^2^~ 0.3. Curiously though, the value of the exponent *β* is still 0.31.

We also found that the number of days from the first to the second peak is linearly related to the distance by road of the county seat from Penn Station (Figure 6d). This again suggests that there was a non-random (directed) migration of households out of NYC that may have spread Covid-19 to surrounding counties. Although we have no proof, we conjecture that the intercept of 15 days on the y axis in Figure 6d may be due to the required quarantine period imposed after moving between locations during the initial phase of the pandemic.

We now list some caveats and limitations of our study, some of which can be mitigated by additional data and future research.

First, the USPS CoA data we analyzed did not include data for months when the moves out of or into the county totaled less than 10 per month. We do not expect this to affect our results by much, as the Excess CoA ranges over three orders of magnitude (Table I and Figure 6c) and the average excess CoA from the 15 counties with the highest population density was 298 changes per month from March–June 2020. A more serious limitation is the fact that the data only reports movements of “households” and does not specify the number of people that moved per household, and the latter will vary from county to county. It may be possible to correct this using data on average household sizes in each county and cell phone use data, especially with voice tagging.

A large number of people commute into NYC each day from counties close by [37]. In particular, there is a lot of commuter traffic into NYC from Westchester, Nassau, Suffolk Counties in NY, Bergen, Essex, Middlesex and Hudson Counties in NJ and Fairfield County in CT (Columns P and Q in Table I). It is possible that when households relocated out of NYC into these counties, some individuals in these households continued to commute into the city daily possibly because of their status as essential workers. Such individuals would increase the risk of Covid-19 infections into the counties. It should be possible to include this correction into our model by studying commuter traffic data between NYC and the surrounding counties in the period March-June 2020. Unfortunately, this data is not yet available [37].

An important limitation of our study is that we do not have data on whether an individual who moved was infected or healthy. To our knowledge, accurate data detailing movement of infected individuals does not exist, although confirmed hospital admissions of Covid-19 cases and tracking of their prior movements may provide a method to generate such data. We note however, that the virus spread more pervasively in NYC residents compared to those in surrounding counties, because there was a higher number of daily cases during the first peak in NYC boroughs compared the surrounding counties (Figures 4a-m compared to Supplementary Figures 10a-e). This means that households moving out of NYC were more likely to contain infected individuals than households moving in the other direction. An interesting way to extend our study to simulate these effects would be to use the CoA data to seed infected individuals moving into various counties and use fits to the actual location and height of the second peaks to find the fractions of these infected individuals, using the number of cases in the first peak to estimate the initial values for these fractions.

We determined the starting point of the second peak as the time when the daily cases in the second peak was near zero and the end point as the time when the case load tapered off to a constant value. Although this is somewhat arbitrary, we have checked that changing these start and end points by a few days is a 5-10% effect and does not change our conclusions.

## Data Availability

The authors confirm that the data supporting the findings of this study are available within the article and its supplementary materials.

## Author Contributions

The model, analysis method and ideas were developed jointly by both authors. AS was responsible for obtaining the data and GB did the data analysis. Both authors jointly wrote the paper and approve the final manuscript.

## Acknowledgements

GB thanks Professors Pablo Tamayo and Jill Mesirov for their hospitality at UC San Diego during his sabbatical in 2019-2020 when this work was begun. The model described in Appendix A is from a joint paper of GB with Professor Charles DeLisi from Boston University. We thank Dr. Meru Bhanot, Economist at Amazon and Professor Sahil Raina from the University of Alberta for several helpful discussions and useful pointers to data sources.

## SUPPLEMENTARY FIGURE LEGENDS

**Supplementary Figure 1:** Map of the counties in the New York metropolitan area, NJ and CT.

**Supplementary Figure 2a-t:** Monthly household moves (CoA counts) from New York City (NYC) boroughs into counties in New Jersey. Note the sharp increase in CoA from March-December 2020 compared to the same time period in 2019 from most NYC boroughs into NJ counties, especially from New York borough (Manhattan). Salem County is not shown because it had no CoA > 10, which was the threshold used by USPS in reporting this data.

**Supplementary Figure 3a-g:** Monthly household moves (CoA counts) from NYC boroughs into counties in Connecticut (CT). Note the sharp increase in CoA from March-December 2020 compared to the same time period in 2019 from most NYC boroughs into CT counties, especially from New York Borough (Manhattan). Windham County is not shown because it had no CoA > 10.

**Supplementary Figure 4a-i:** Monthly household moves (CoA counts) from NYC boroughs into nine counties in New York (NY) State which are closest to NYC. Note the sharp increase in CoA from March-December 2020 compared to the same time period in 2019 from most NYC boroughs into surrounding counties in NY State, especially from New York borough (Manhattan).

**Supplementary Figure 5a-e:** Monthly household moves (CoA counts) from each NJ/CT/NY county into each of the NYC boroughs: Bronx (a), Kings (b), New York (c), Queens (d) and Richmond (e). Note the lack of excess household movements in 2020 compared to 2019.

**Supplementary Figure 6a-u:** Seven-day averaged plots of daily Covid-19 cases for each NJ county and the fits to the SIR model for the first peak (Appendix A). The data is shown as blue dots and the model fits as solid red lines.

**Supplementary Figure 7a-u:** The excess daily cases in each county in NJ. The excess cases are estimated as the difference between the seven-day averaged data for daily Covid-19 cases (blue dots) and the fits to the SIR model (red lines) from Supplementary Figure 6. The solid black dots show the time points between which we sum the excess cases to estimate the number of cases in the second peak (Table I).

**Supplementary Figure 8a-h:** Seven-day averaged plots of daily Covid-19 cases for each CT county and the fits to the SIR model for the first peak (Appendix A). The data is shown as blue dots and the model fits as solid red lines.

**Supplementary Figure 9a-h:** The excess daily cases in each county in CT. The excess cases are estimated as the difference between the seven-day averaged data for daily Covid-19 cases (blue dots) and the fits to the SIR model (red lines) from Supplementary Figure 6. The solid black dots show the time points between which we sum the excess cases to estimate the number of cases in the second peak (Table I).

**Supplementary Figure 10a-e:** Seven-day averaged plots of daily Covid-19 cases for each NYC borough and the fits to the SIR model for the first peak (Appendix A). The data is shown as blue dots and the model fits as solid red lines.

**Supplementary Figure 10f-n:** Seven-day averaged plots of daily Covid-19 cases for nine NY counties adjoining NYC and the fits to the SIR model for the first peak (Appendix A). The data is shown as blue dots and the model fits as solid red lines.

**Supplementary Figure 11a-e:** The excess daily cases in each NYC Borough. The excess cases are estimated as the difference between the seven-day averaged data for daily Covid-19 cases (blue dots) and the fits to the SIR model (red lines) from Supplementary Figure 6. The solid black dots show the time points between which we sum the excess cases to estimate the number of cases in the second peak (Table I).

**Supplementary Figure 11f-n:** The excess daily cases in nine NY counties adjoining NYC. The excess cases are estimated as the difference between the seven-day averaged data for daily Covid-19 cases (blue dots) and the fits to the SIR model (red lines) from Supplementary Figure 6. The solid black dots show the time points between which we sum the excess cases to estimate the number of cases in the second peak (Table I).

## SUPPLEMENTARY TABLE LEGENDS

**Supplementary Table 1a-b:** Monthly change of address (CoA) counts for household moves between NYC boroughs and counties in NJ/CT/NYS for each month in 2019 and 2020. This data was obtained from the United States Postal Service.

**Supplementary Table 2a-c:** Cumulative daily reported Covid-19 case counts for NJ, CT and NY counties and NYC Boroughs that was used in the analysis presented in this paper. The data was obtained from https://github.com/CSSEGISandData/COVID-19/tree/master/csse_covid_19_data/csse_covid_19_time_series

**Supplementary Table 3a-c:** Demographics, fitted SIR model parameters and predictions from the fits for NJ, CT, NY counties and NYC boroughs.

