## Appendix for "Migration of households from New York City and the Second Peak in Covid-19 cases in New Jersey, Connecticut and New York Counties"

### Appendix A:

#### A simplified SIR model for the first peak in daily cases

We model the Covid-19 pandemic using a simplified version of the SIR model [A1], which partitions the population into three compartments, Susceptibles (S), Infectious (I) and Removed R: Recovered or Dead after being infected. Such models have been used to study the global spread of diseases in a variety of contexts (For some recent reviews, see [A2-A4]).

So far, the Covid-19 pandemic, in places such as NYC Boroughs and surrounding counties, which implemented measures such as limiting movement of people via partial or total lockdowns for some time, had the following dynamics, at least in the early stages of the pandemic: After being infected, an individual remains able to infect others for an average of $T_L$ days. After a time $T_L$, the infected individual becomes sick, gets tested, is identified as infected and is removed from the pool by quarantine or hospitalization.

Assuming this dynamics, our simplified model is defined as follows: At t=0, there is a pool of N interacting individuals, almost all in the S compartment, except for the few infected cases in the I compartment. The R compartment is empty at t=0. Over time, individuals move from S to I and from I to R. In R, they either recover or die. Since the Recovered pool is populated only from the Infected pool, on average, the number removed each day must equal the number infected sometime in the past; i.e. the two are related by a fixed time displacement and a "mortality probability" factor. We assume that the number of deaths and the number of individuals recovered each day are proportional to the number Removed each day by fixed probabilities, and that these remain invariant over the time of the lockdown/quarantine. In other words, the number of dead or recovered each day are proportional to the number infected on some previous day, with different time delays and probabilities.

Define:

$X_1(t)$ = number of Susceptible individuals at time t, (A1)
$X_2(t)$ = number of Infected individuals at time t, (A2)
$X_3(t)$ = number of individuals Removed at time t (A3)
$X_1(t) + X_2(t) + X_3(t) = N$ = constant (A4)

A fraction $\delta$ of the infected individuals will die after being identified as infected. On average, there will be a time delay $T_D$ between when a person is identified to be infected (tests positive) and when he/she dies of the disease. $T_D$ will depend on a variety of factors, such as quality of care, age, severity of disease, co-morbidities, immune status, whether vaccinated or not etc.

Under these assumptions, the number of deaths $X_4(t)$ at time $t$ is related to the number of Infected cases $X_2$ by:

$X_4(t) = \delta X_2(t - T_D)$ = number of individuals that died on day t (A5a)

Similarly, the number of Recovered at time $t$ will be:

$X_5(t) = \beta X_2(t - T_{Re})$ = number of individuals that recovered on day t (A5b)

Further define:

- $\alpha$: the transmission rate, the probability of infection per day per contact between an infected person and a susceptible person. (A6a)
- $\gamma$: the rate at which individuals leave the infected population (A6b)

Note that $\frac{1}{\gamma} = T_L$: is the time period when an infected individual is able to infect a susceptible individual (A6c)

With these assumptions, the basic equations of our model are:

$$\frac{dX_1(t)}{dt} = -\alpha X_1(t) X_2(t) \qquad (A7a)$$

$$\frac{dX_2(t)}{dt} = \alpha X_1(t) X_2(t) - \gamma X_2(t) \qquad (A7b)$$

and the boundary conditions at $t = 0$ are:

$$X_1(t = 0) = (N - a) \text{ and } X_2(t = 0) = a \qquad (A7c)$$

Eliminating t from (A7a) and (A7b) by taking their ratio, it is easy to show that
i$X_1$and $X_2$ are related by :

$$X_2(t) = N - a + \frac{N}{R} \log\left(\frac{X_1(t)}{N-a}\right) - X_1(t) \qquad (A8)$$

From (A7b) we find that the maximum in $X_2$ is at $X_1 = \frac{\gamma}{\alpha} = \frac{N}{R}$ (A9)

where $R \equiv \frac{\alpha N}{\gamma}$ (A10)

R is the so called “Pandemic Parameter”. In our simple model, R is a constant.

Substituting (A9) into (A8) gives:

Maximum value of $X_2 \equiv P = N - \frac{N}{R}[1 + \log(R)]$ (A11)

For small t, we can expand the log in (A8) as follows:

$$\log\left(\frac{X_1(t)}{N-a}\right) = \log\left[1 - \left(1 - \left(\frac{X_1(t)}{N-a}\right)\right)\right] \sim - \left(1 - \left(\frac{X_1(t)}{N-a}\right)\right) \qquad (A12)$$

Substituting (A12) into (A8) gives, after some simple algebra,

$$X_2(t) = \frac{R-1}{R}\left[N - X_1(t)\left(1 - \frac{a}{(R-1)N}\right)\right] \sim \frac{R-1}{R}\,[N - X_1(t)] \qquad \text{(A13)}$$

Finally, substituting (A13) into (A7a) gives a Logistic Equation for $X_1$, which is correct to O(1/N):

$$\frac{dX_1}{dt} = -\gamma(R-1)X_1(t)[1 - \frac{X_1(t)}{N}] \qquad \text{(A14)}$$

whose solution, with the boundary condition (A7c) is:

$$X_1(t) = \frac{N}{\left[1+\frac{a}{N-a}e^{\gamma(R-1)t}\right]} \qquad \text{(A15)}$$

For small t, we can expand the denominator to get:

$$X_1(t) \sim N\,[\,1 - \frac{a}{N-a}e^{\gamma(R-1)t}],\ t\ \text{small} \qquad \text{(A16)}$$

Substituting into (A13) gives,

$$X_2(t) \sim \frac{(R-1)aN}{R(N-a)}e^{\gamma(R-1)t} \qquad \text{(A17)}$$

Thus, for small t, $X_2(t)$ increases exponentially, fed by a movement of individuals from the S compartment to the I compartment.

The coefficient of t in the exponential rise of $X_2(t)$ for small t is $\gamma(R-1)$. (A18)

**Fitting the Model to Data**

The first confirmed cases in NY were registered on March 1, in NJ on March 4, and in CT on March 8. Each of the tri-states responded by declaring a state of emergency (NY and NJ on March 7, and CT on March 10) and issuing a joint mandate that banned crowds of over 50 people, closed casinos, movie theatres, and gyms, limited access to bars and restaurants to takeout service, and set a night curfew. This was followed by each state ending in-person schooling, closing all non-essential businesses, and issuing a "stay-at-home" order in an attempt at full-scale quarantine. All three states stayed in these lockdown conditions until a phased reopening began with Phase 1 (NY on June 8, NJ on June 10, CT on May 20). This reopening extended throughout June and July.

Data on Covid-19 cases and deaths for NYC Boroughs and the surrounding counties in NJ, CT and NY was downloaded from https://github.com/CSSEGISandData/COVID-19/tree/master/csse_covid_19_data/csse_covid_19_time_series

(a) The cumulative number $I(t)$ and daily number $X_2(t)$ of confirmed cases (A19)

and
(b) The cumulative number $\mathrm{D(t)}$ and daily number $\mathrm{X_4(t)}$ of deaths (A20)

These are related by:
$$\mathrm{X_2(t) = I(t) - I(t-1)} \tag{A21}$$

and,

$$\mathrm{X_4(t) = D(t) - D(t-1)}\ . \tag{A22}$$

We note that the data identifies *Confirmed* cases, whereas the model described above requires the number of *Infected* cases. These are in general quite different, since many infected cases are likely not included in the confirmed cases. This raises the following two complications which we cannot model for lack of data:

- An infected, unconfirmed individual infects a susceptible individual who is then counted among the confirmed cases: $\mathrm{I_{unconfirmed}} \rightarrow \mathrm{infects\ Susceptible} \rightarrow \mathrm{who\ is\ confirmed}$
- An infected and confirmed individual infects a susceptible individual who is not counted among the confirmed infected. $\mathrm{I_{confirmed}} \rightarrow \mathrm{infects\ Susceptible} \rightarrow \mathrm{who\ is\ not\ confirmed}$

To deal with these complications, for which data is not available, we have to make the following simplifying assumptions:

- At any given time, the number of confirmed cases is proportional to the number of infected cases.
- The fraction of individuals that move in the two directions described above is fixed.

With these assumptions, we can use the data of confirmed cases at a given time as an approximation for the number of infected cases.

Although we do not analyze the data for the deaths in this paper, it is possible to extend the model to find the parameters $\delta, \beta, T_D, T_{Re}$. It is worth noting that the dynamics of the number removed each day and the total number of removed do not enter our analysis explicitly. Some connection to the full SIR model [A1] can be made by noting that the total number of removed individuals increases asymptotically at large times to $\mathrm{N(1 - S_1(\infty))}$, where,

$$\mathrm{S_1(\infty) = \frac{X_1(\infty)}{N}} \tag{A23a}$$

This quantity is also related to R by:

$$\mathrm{R} = -\ \frac{\mathrm{log(S_1(\infty))}}{\mathrm{[1-S_1(\infty)]}} \tag{A23b}$$

We determine the following parameters for NYC Boroughs and each county in NJ, CT and NY that we analyzed:

- $\mathbf{N}$, as defined in (A4)
- $\boldsymbol{\alpha}$, the transmission rate or the number of infections per day per contact (A6)
- $\boldsymbol{\gamma}$, the average rate at which individuals leave the infected pool (A7)
- $\mathbf{R}$ **,** the average number of transmissions per individual
- $\mathbf{T_L}$**,** the infective period = $1/\boldsymbol{\gamma}$.

The parameters $\mathrm{N}, \alpha, \gamma, \mathrm{R}$ were obtained as follows:

- Using (A10), we can define $\alpha$ in terms of $\mathrm{N}, \gamma, \mathrm{R}$. This eliminates $\alpha$.
- Choosing a value for R and estimating P from the data as the maximum in the daily cases determines N in terms of R using (A11). This eliminates N.
- $\gamma(\mathrm{R}-1)$ is determined as the coefficient of $\mathrm{t}$ in the exponential increase in $\mathrm{X}_2(\mathrm{t})$ for small t (A17-18). This eliminates $\gamma$.

Finally, the optimum value of R is obtained by iterating the sequence above, using a numerical solver to fit the observed data for $\mathrm{X}_2(\mathrm{t})$ to find the value of R that best fits the data. We emphasize that in determining these parameters, we are only using the ascending limb and the peak in the data for $\mathrm{X}_2(\mathrm{t})$ and using these to predict how $\mathrm{X}_2(\mathrm{t})$ will evolve in time for some period past the peak.

A1. Kermack W, McKendric A (1991), *Contributions to the mathematical theory of epidemics – I. Bulletin of Mathematical Biology*. 53 (1–2): 33–55; ibid. *Contributions to the mathematical theory of epidemics – II. The problem of endemicity*", Bulletin of Mathematical Biology. 53 (1–2): 57–87. doi:10.1007/BF02464424. PMID 2059742; ibid. *Contributions to the mathematical theory of epidemics – III. Further studies of the problem of endemicity*. Bulletin of Mathematical Biology. 53 (1–2): 89–118. doi:10.1007/BF02464425. PMID 2059743.

A2. Huppert A, Katriel G. *Mathematical modelling and prediction in infectious disease epidemiology*. Clin Microbiol Infect. 2013;19(11):999-1005. doi:10.1111/1469-0691.12308

A3. Brauer F, *Mathematical epidemiology: Past, present, and future*. Infect Dis Model (2017) 2(2):113-127. Published 2017 Feb 4. doi:10.1016/j.idm.2017.02.001

A4. Colizza V, Barrat A, Barthelemy M, Valleron AJ, Vespignani A, *Modeling the worldwide spread of pandemic influenza: baseline case and containment interventions*. PLoS Med. 2007;4(1):e13. doi:10.1371/journal.pmed.0040013.
