## Supplementary figures and images for "Migration of households from New York City and the Second Peak in Covid-19 cases in New Jersey, Connecticut and New York Counties"

### Supplementary Figure 1

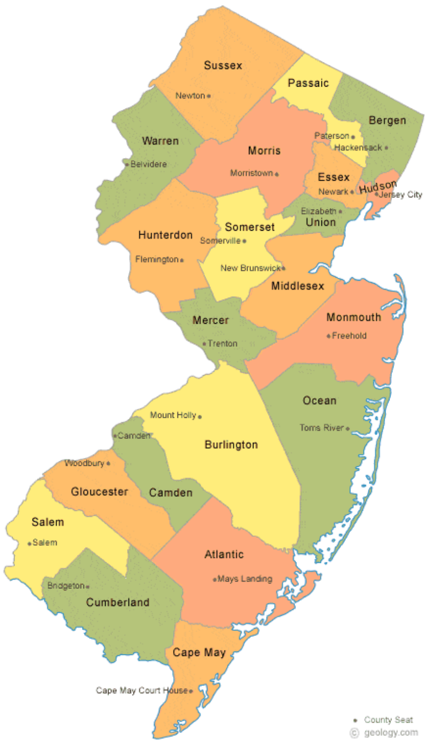

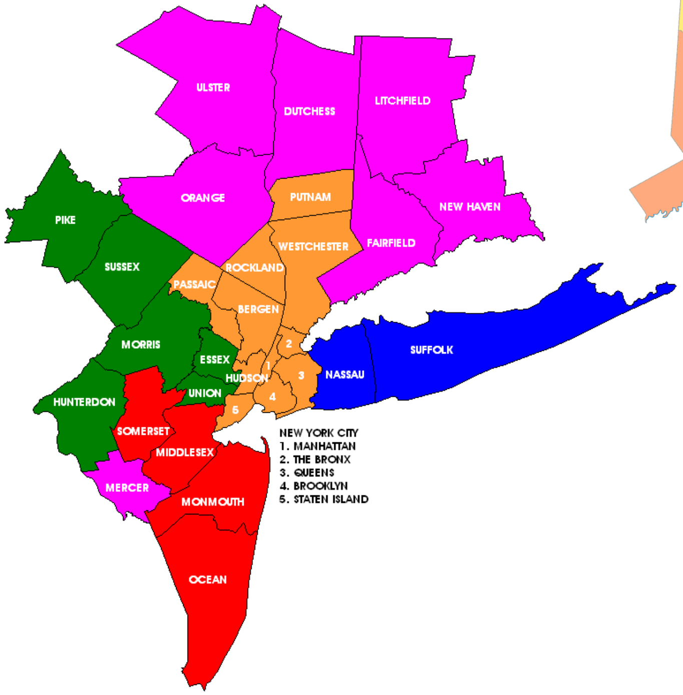

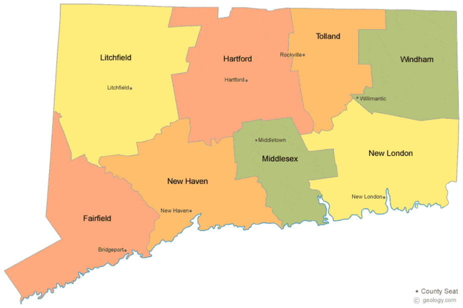

### Supplementary Figure 3

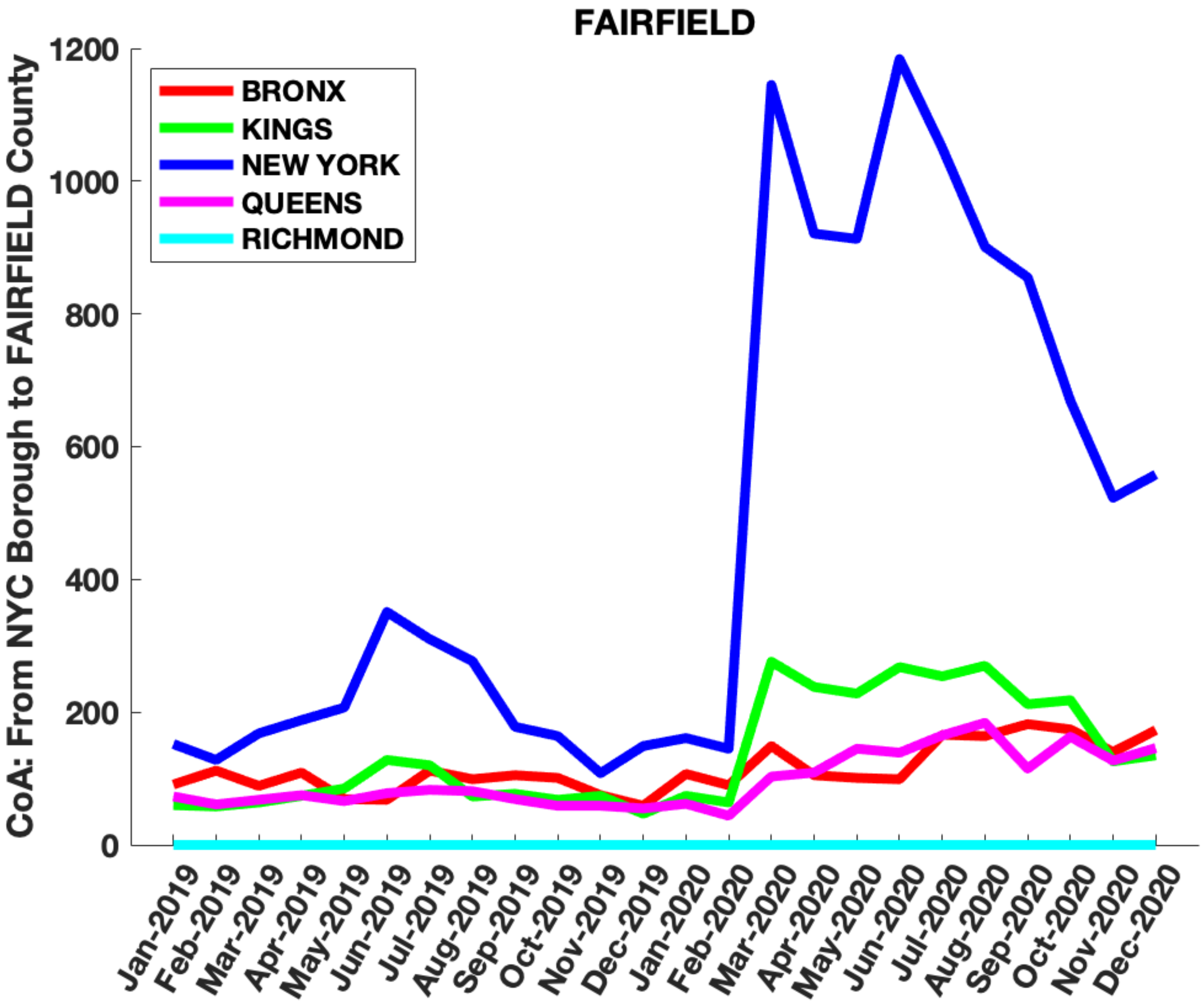

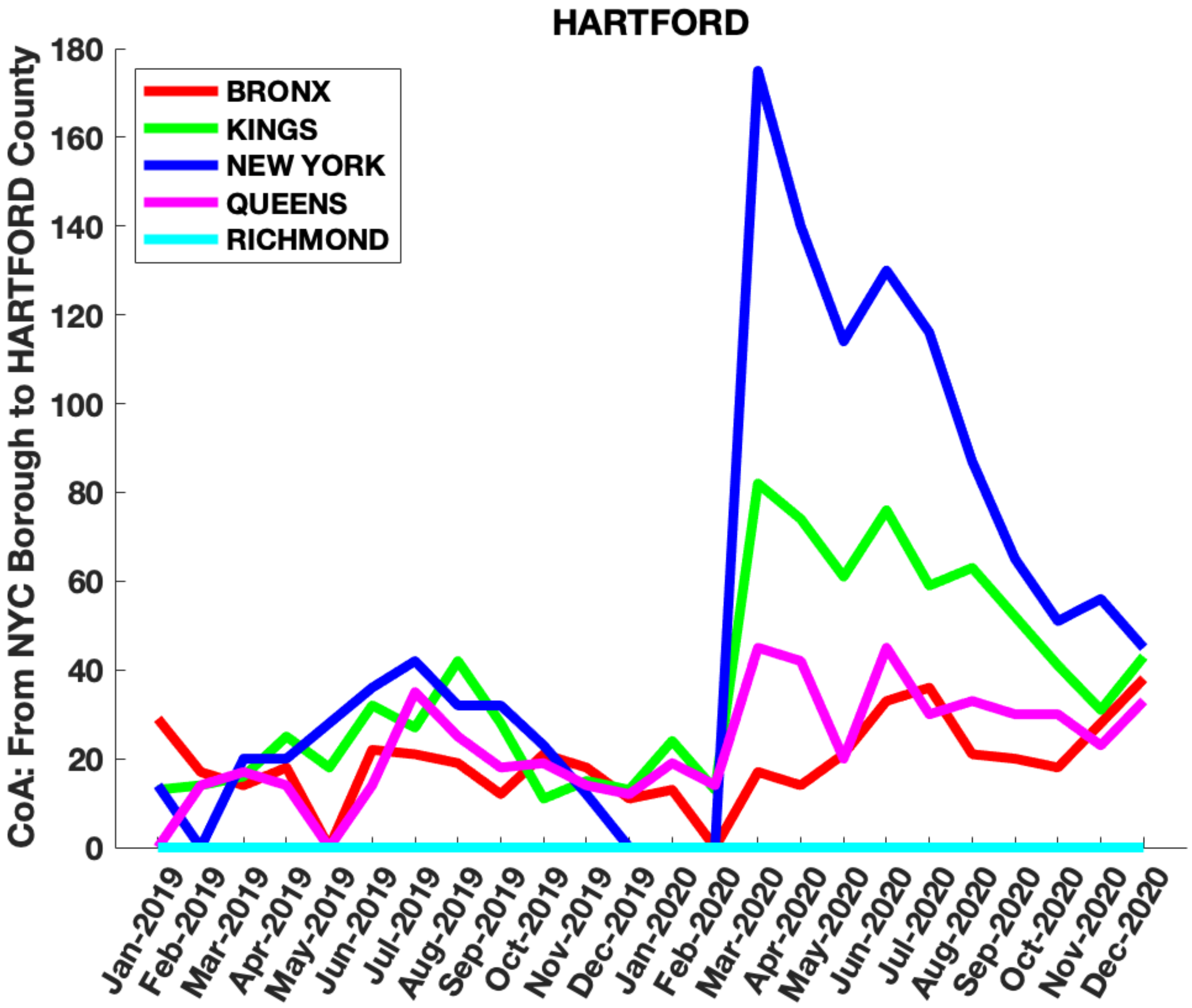

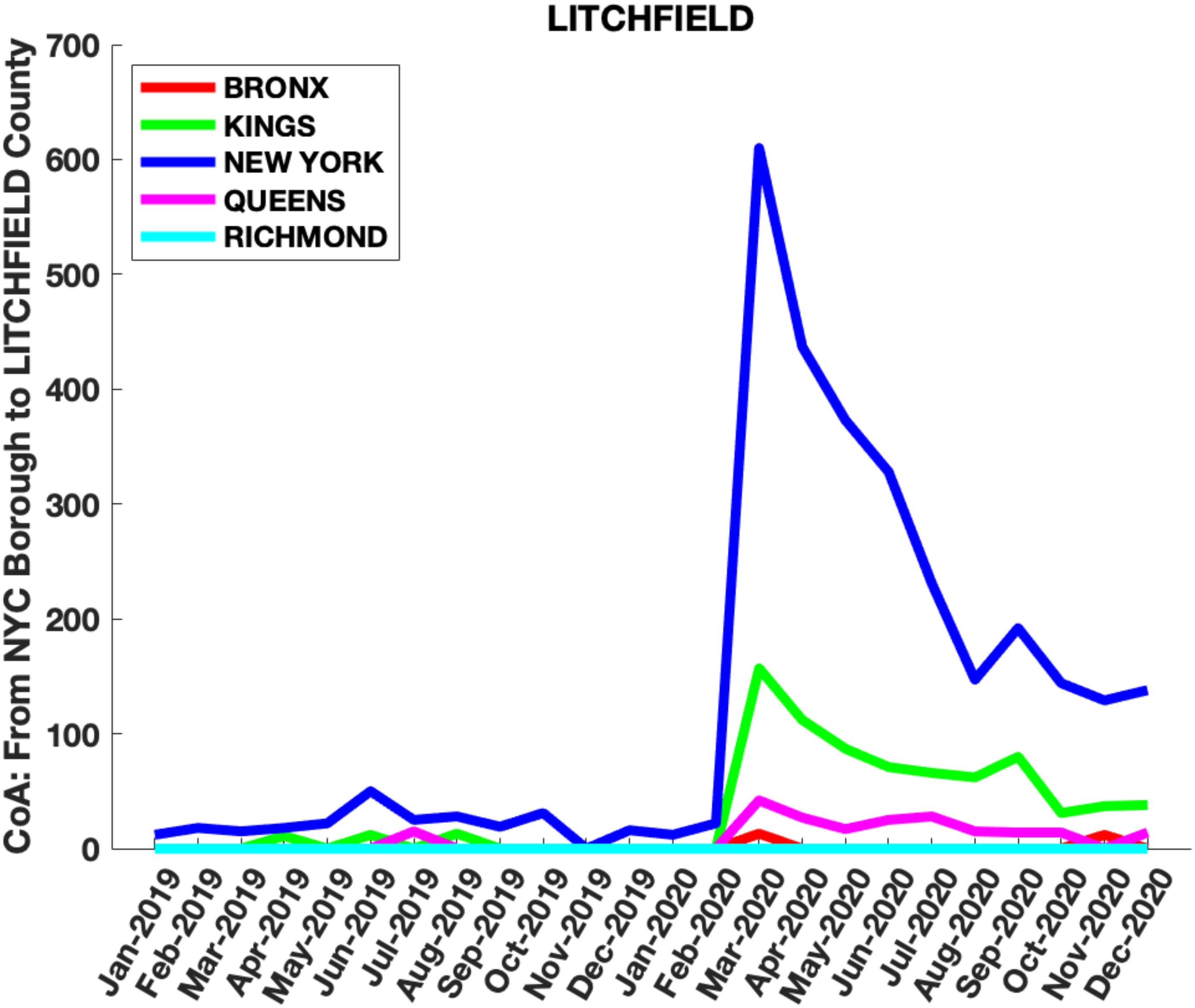

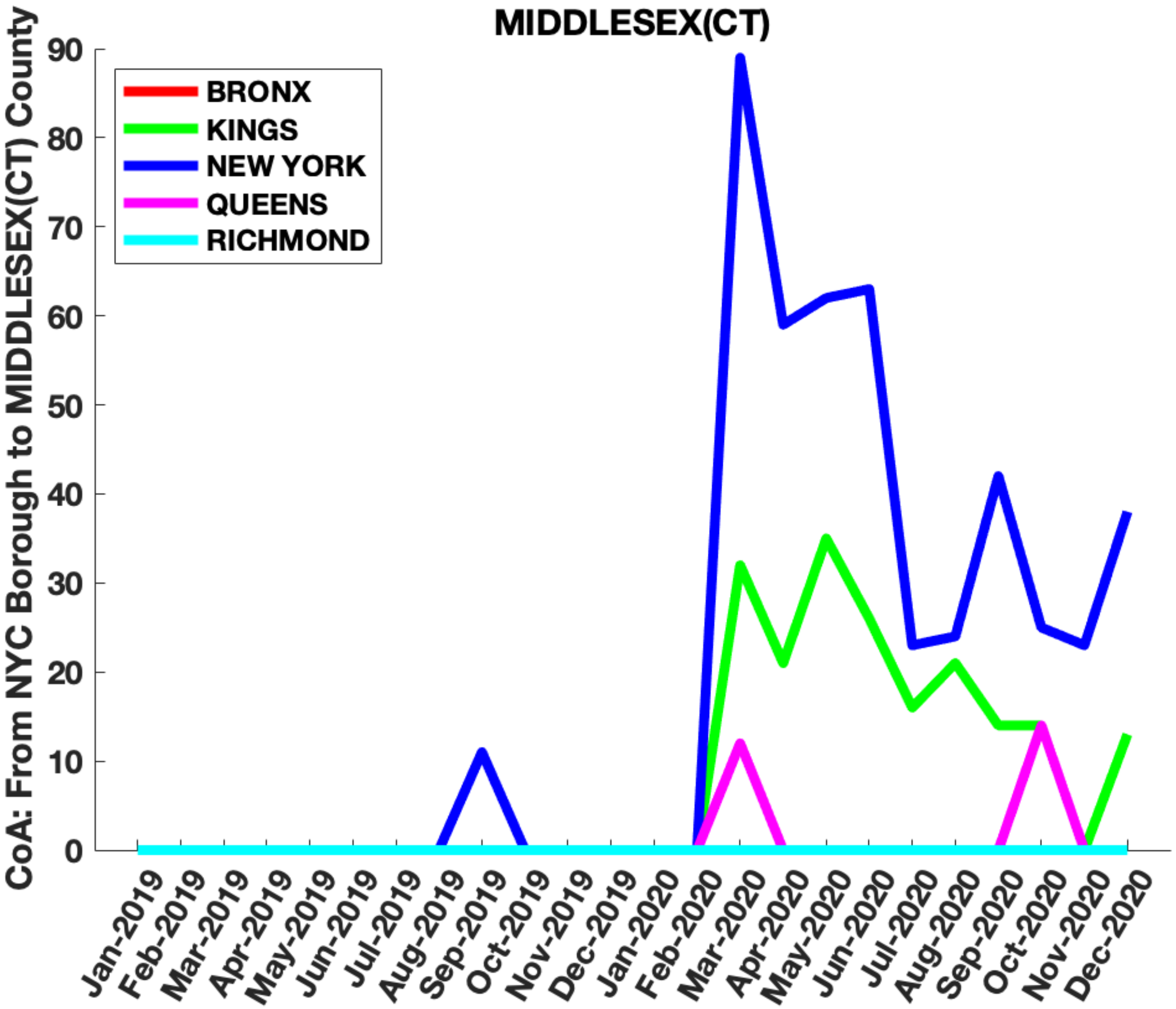

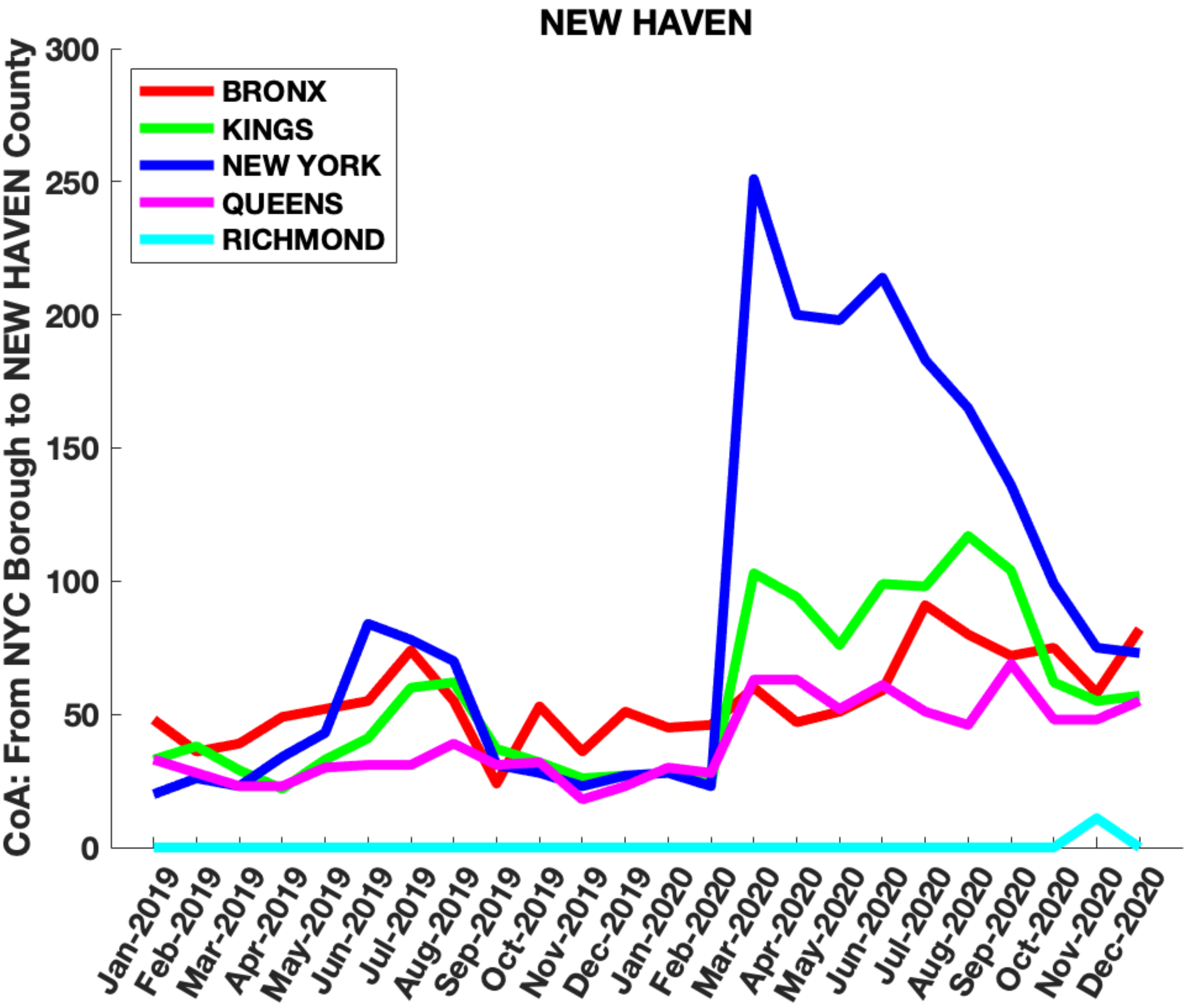

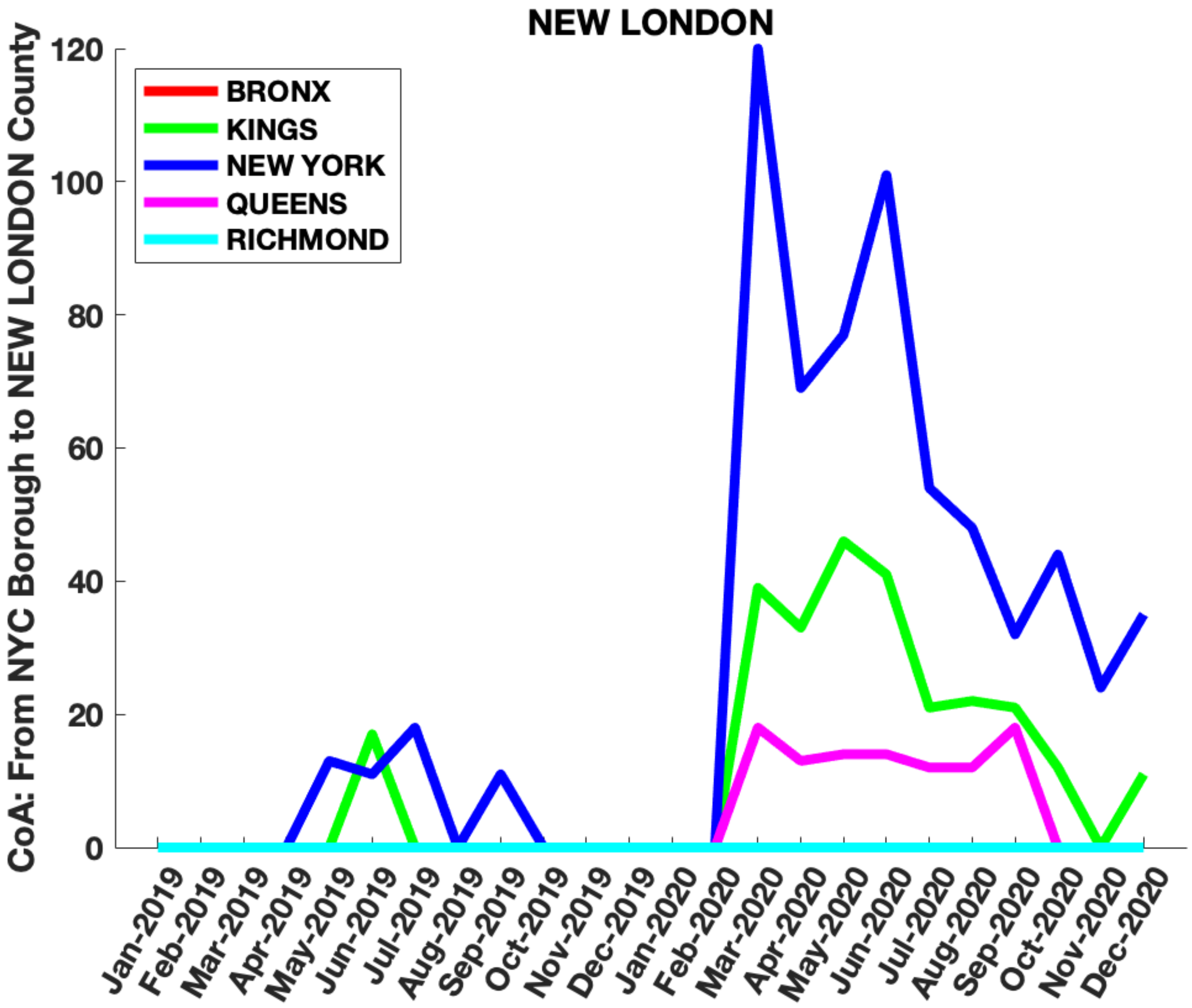

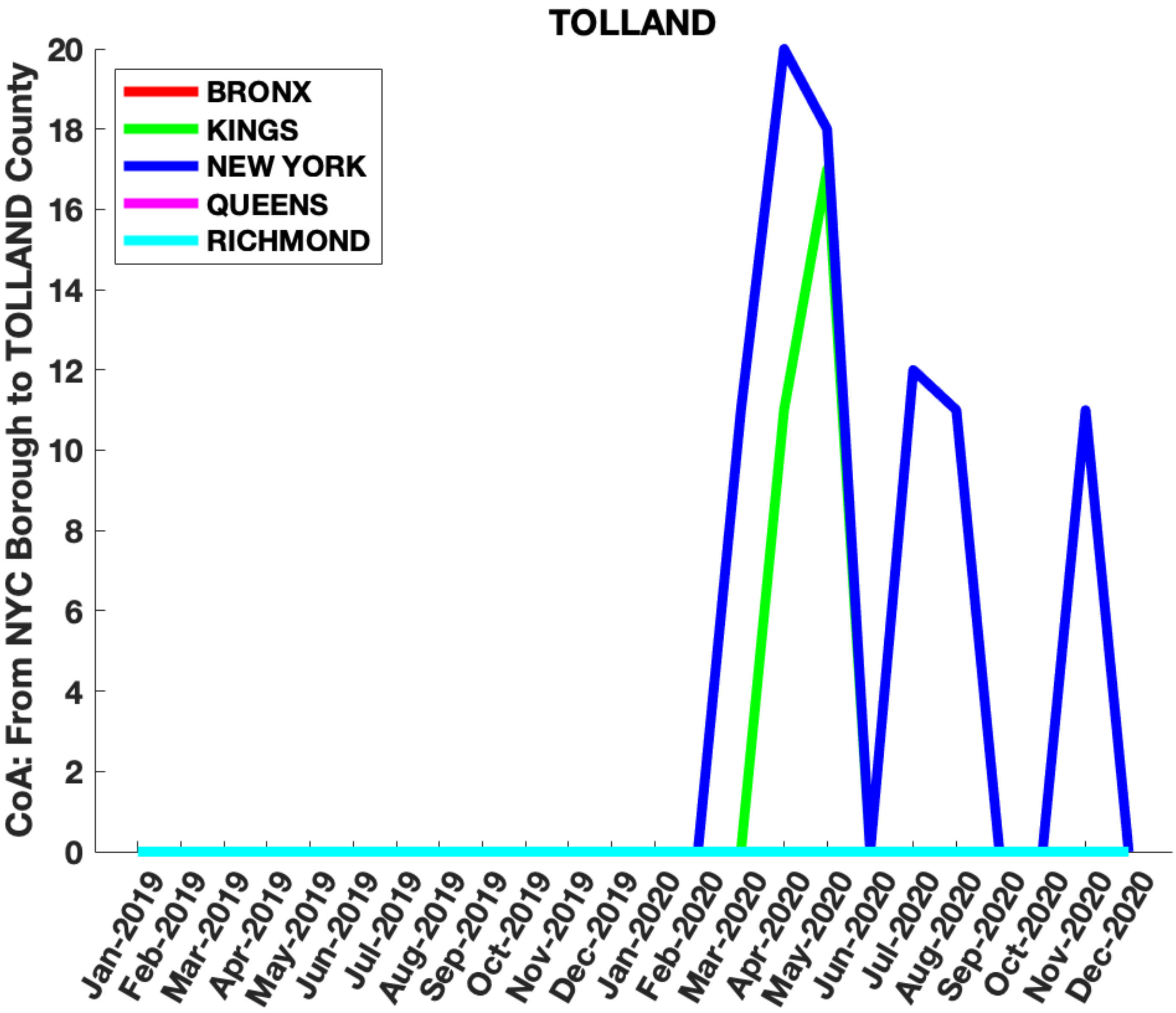

### Supplementary Figure 4

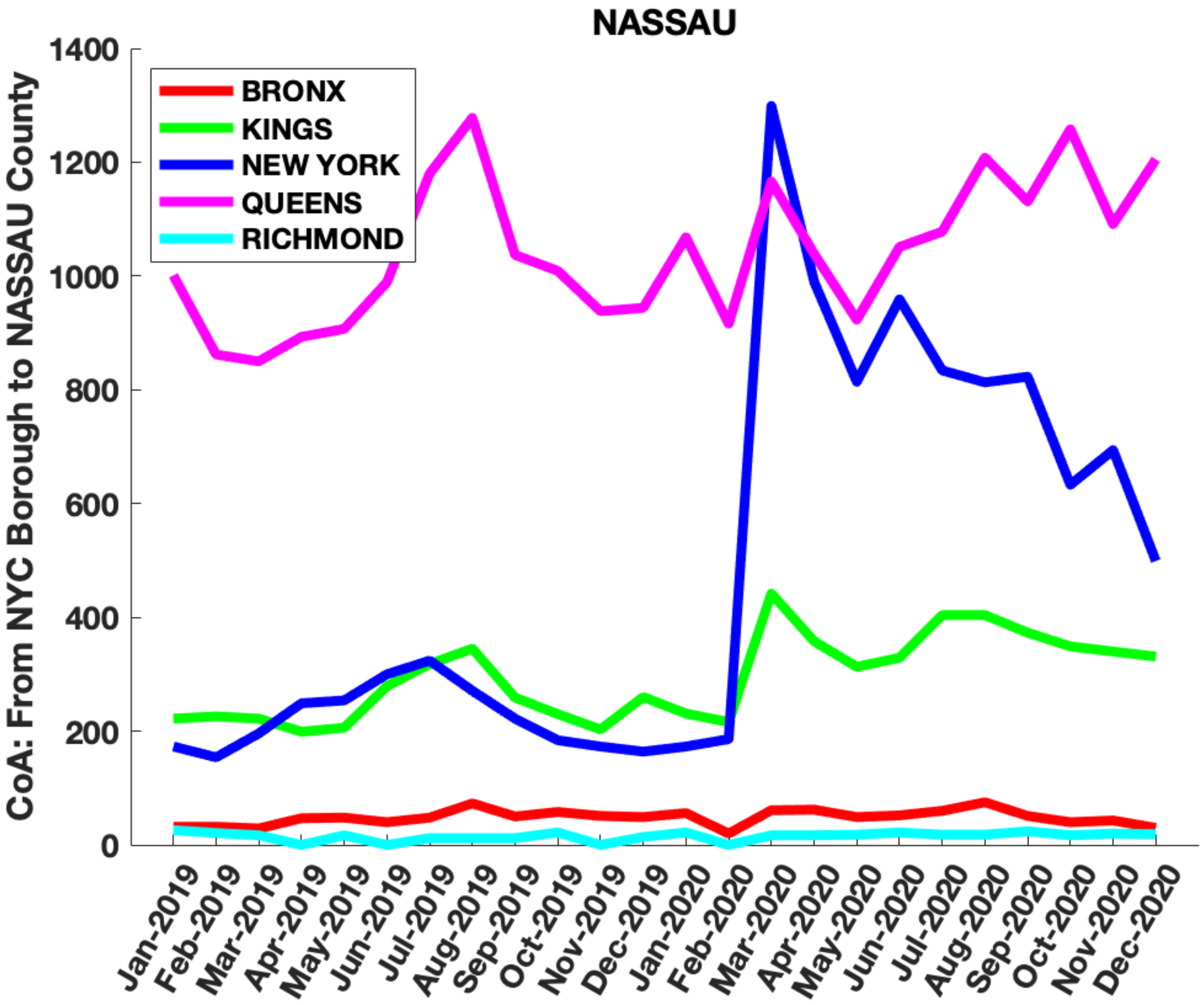

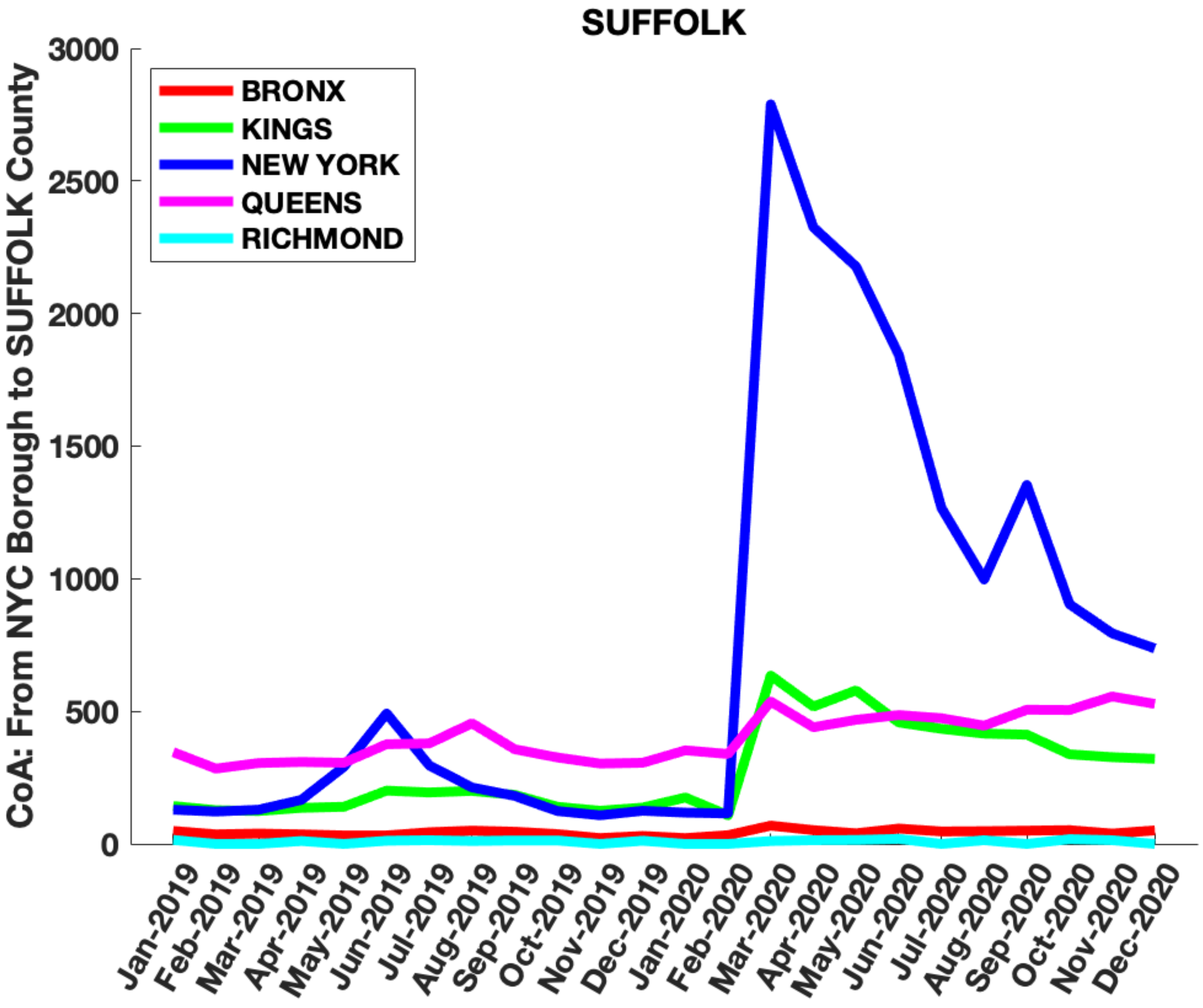

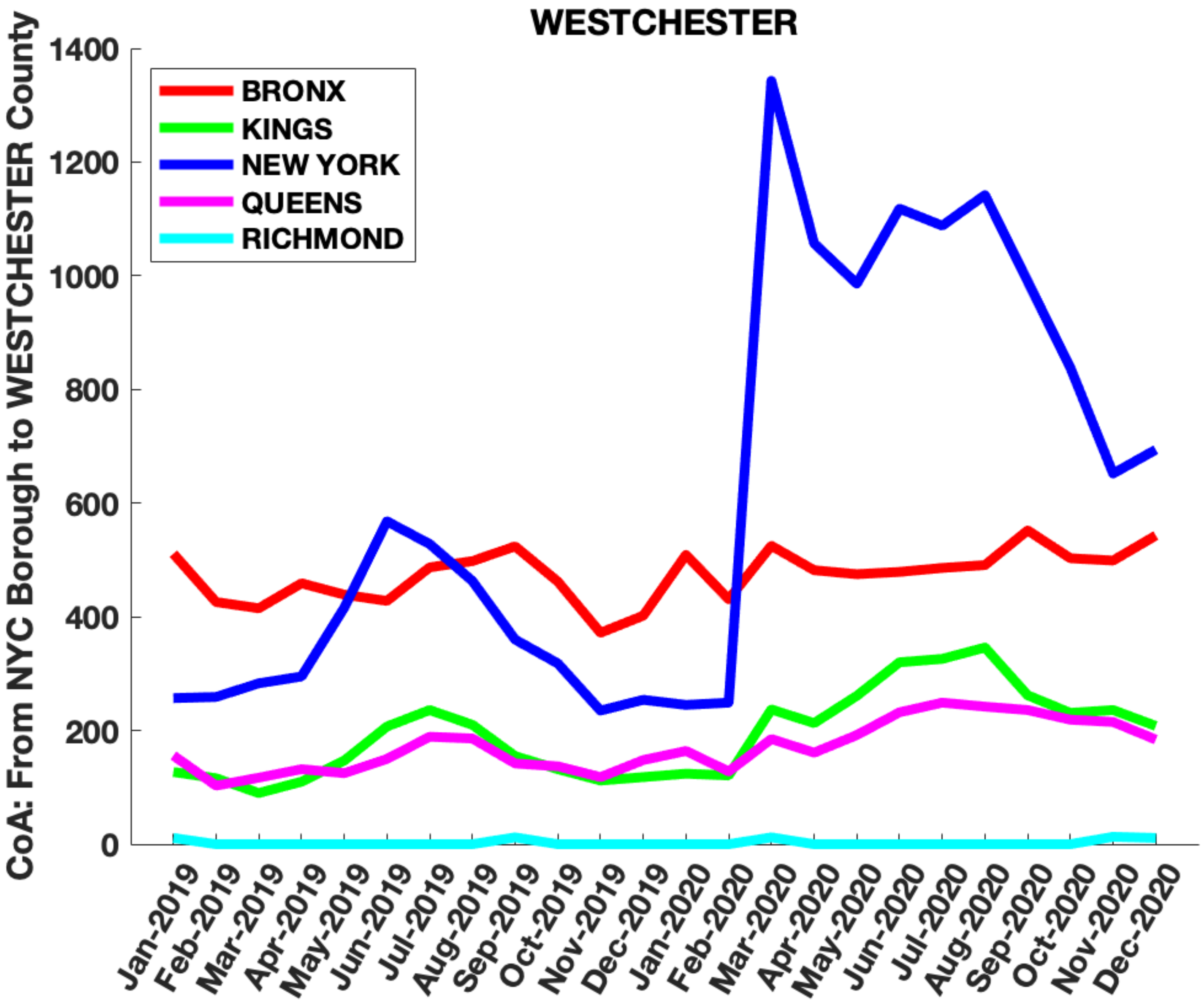

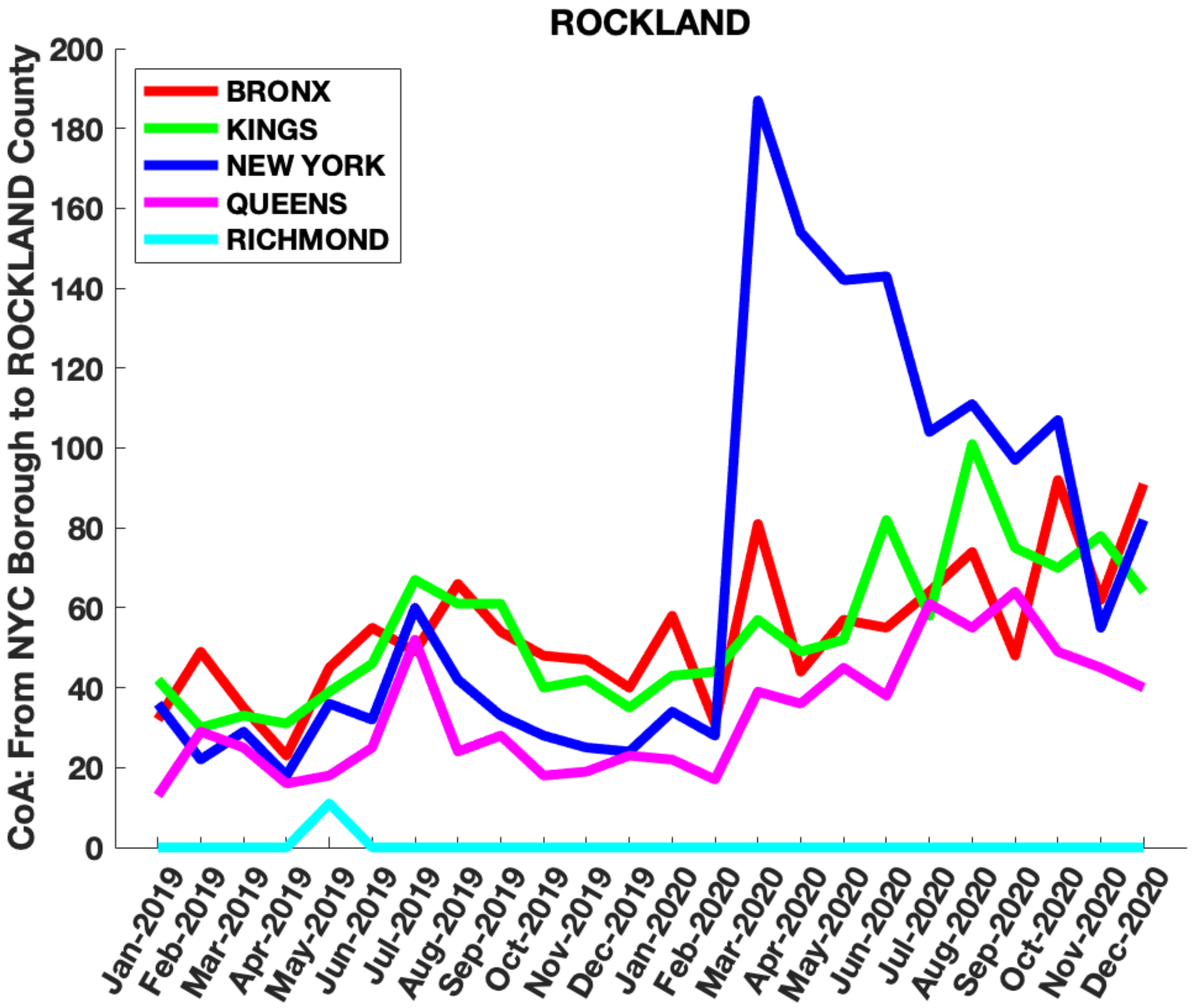

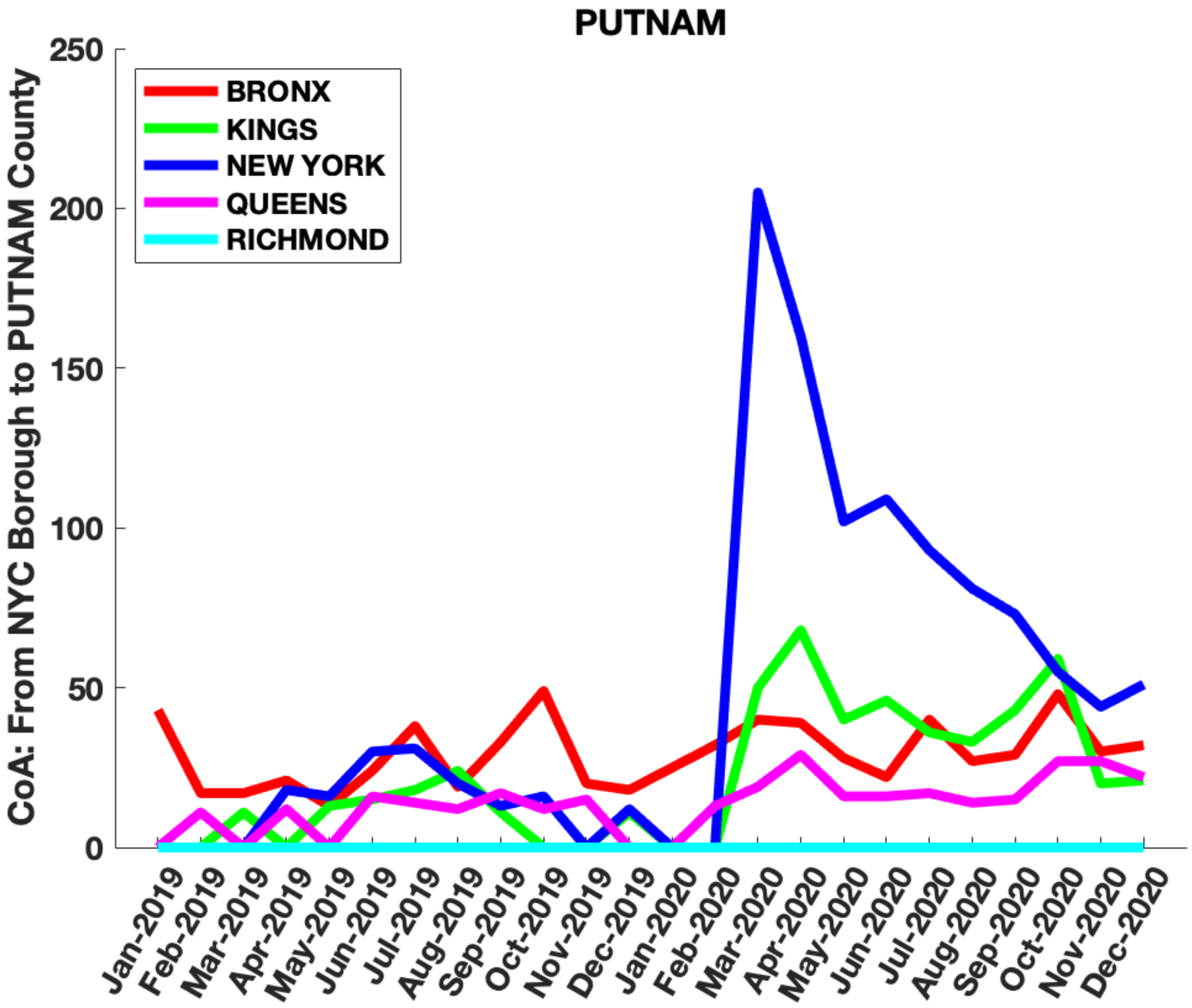

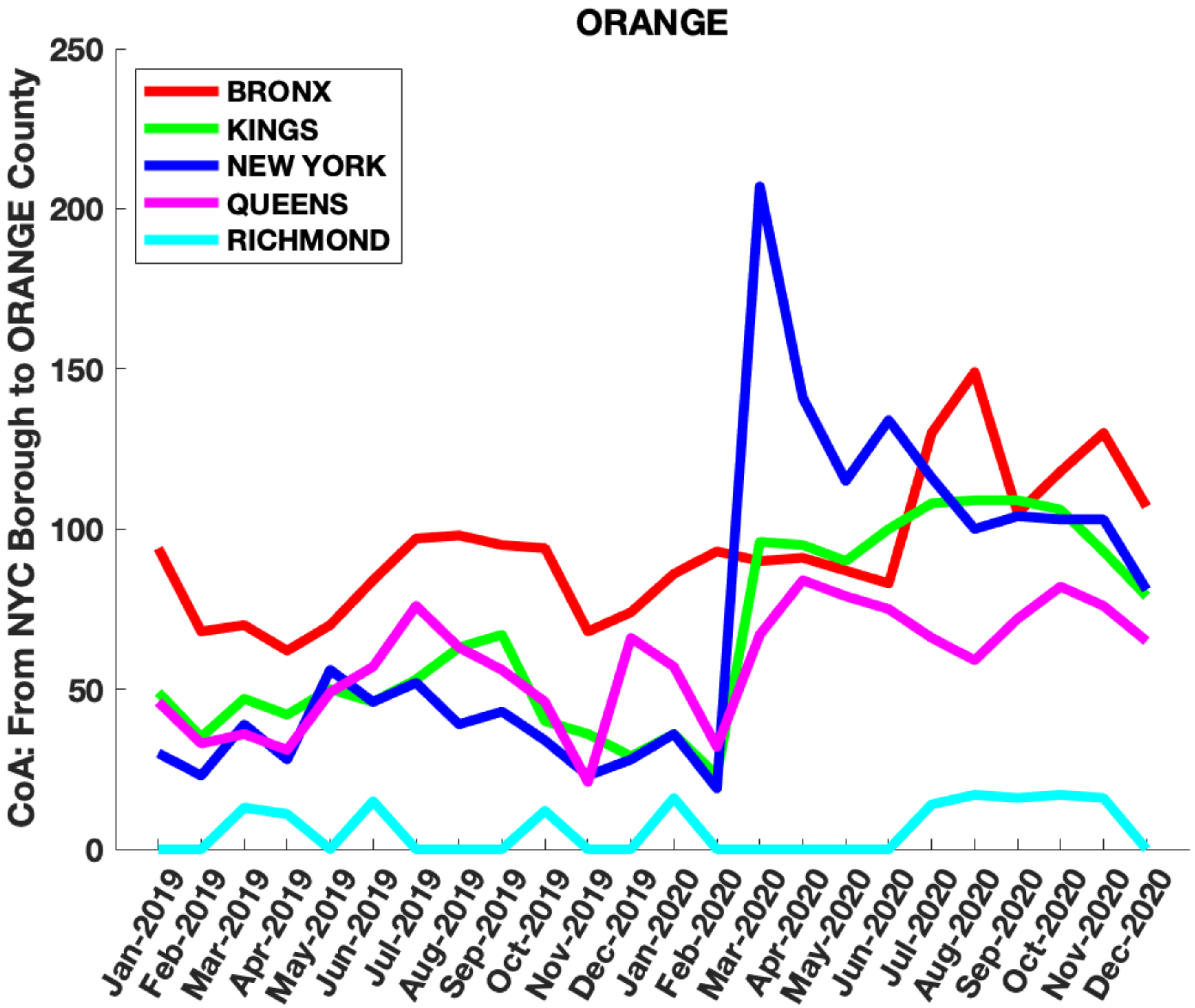

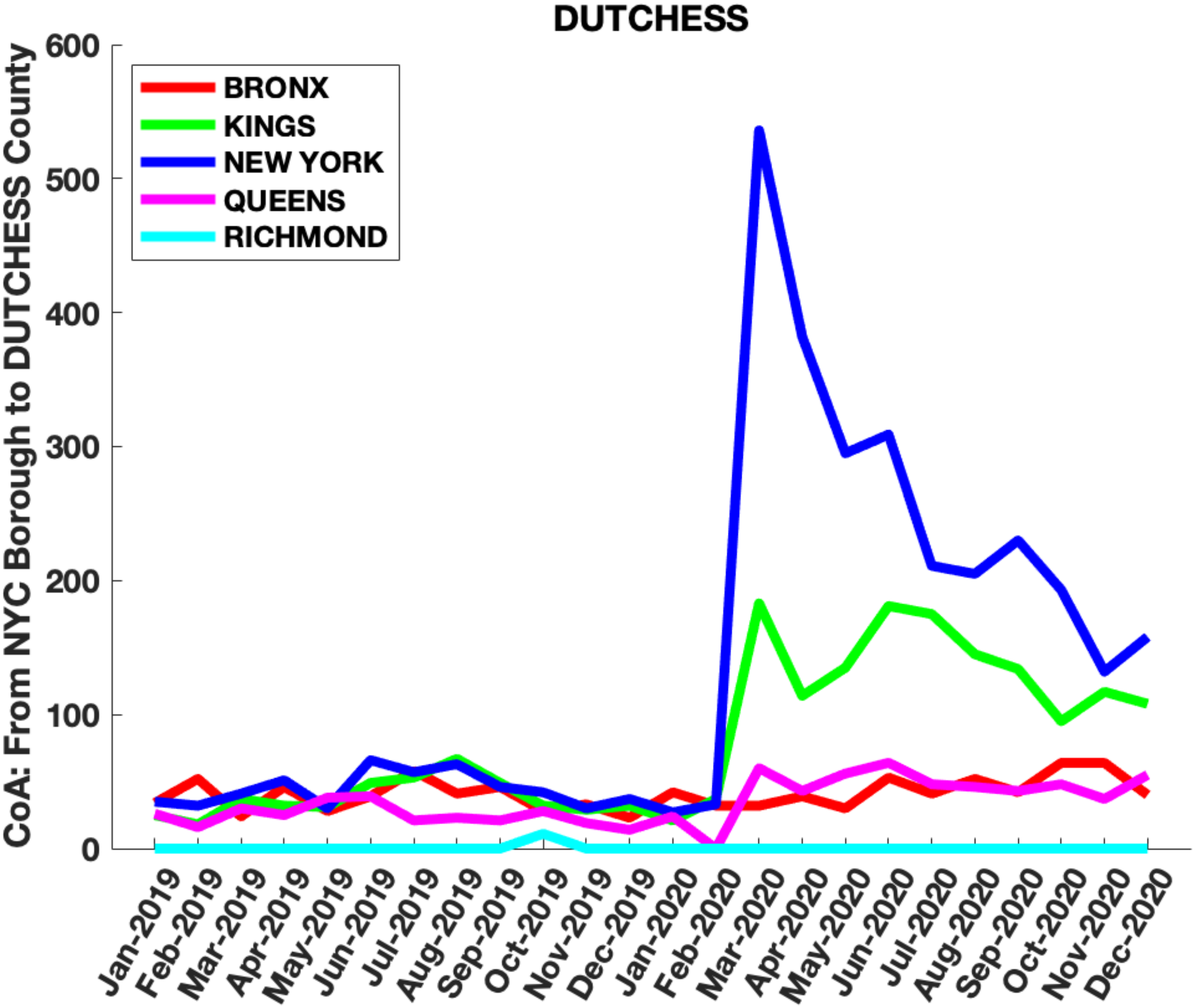

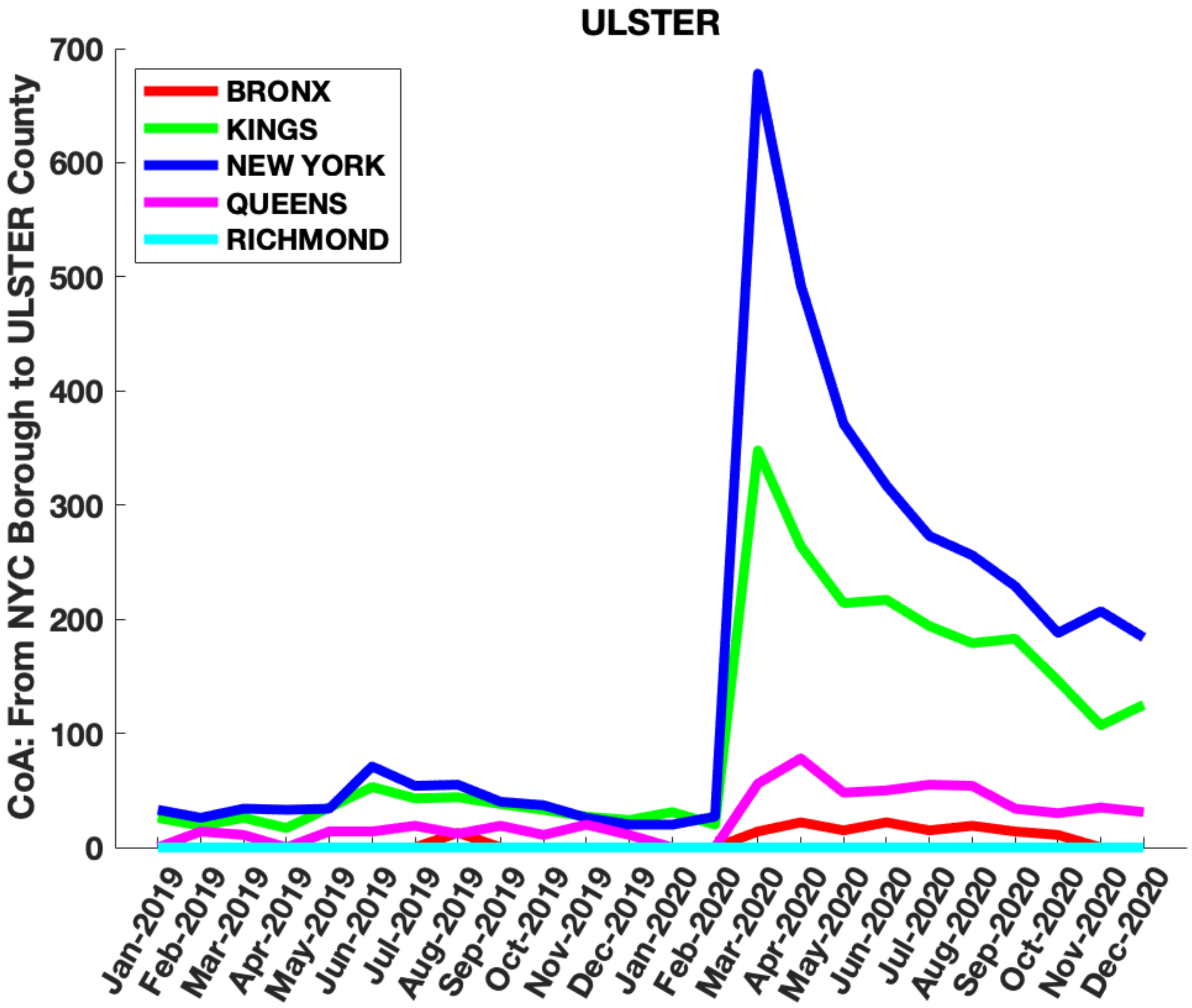

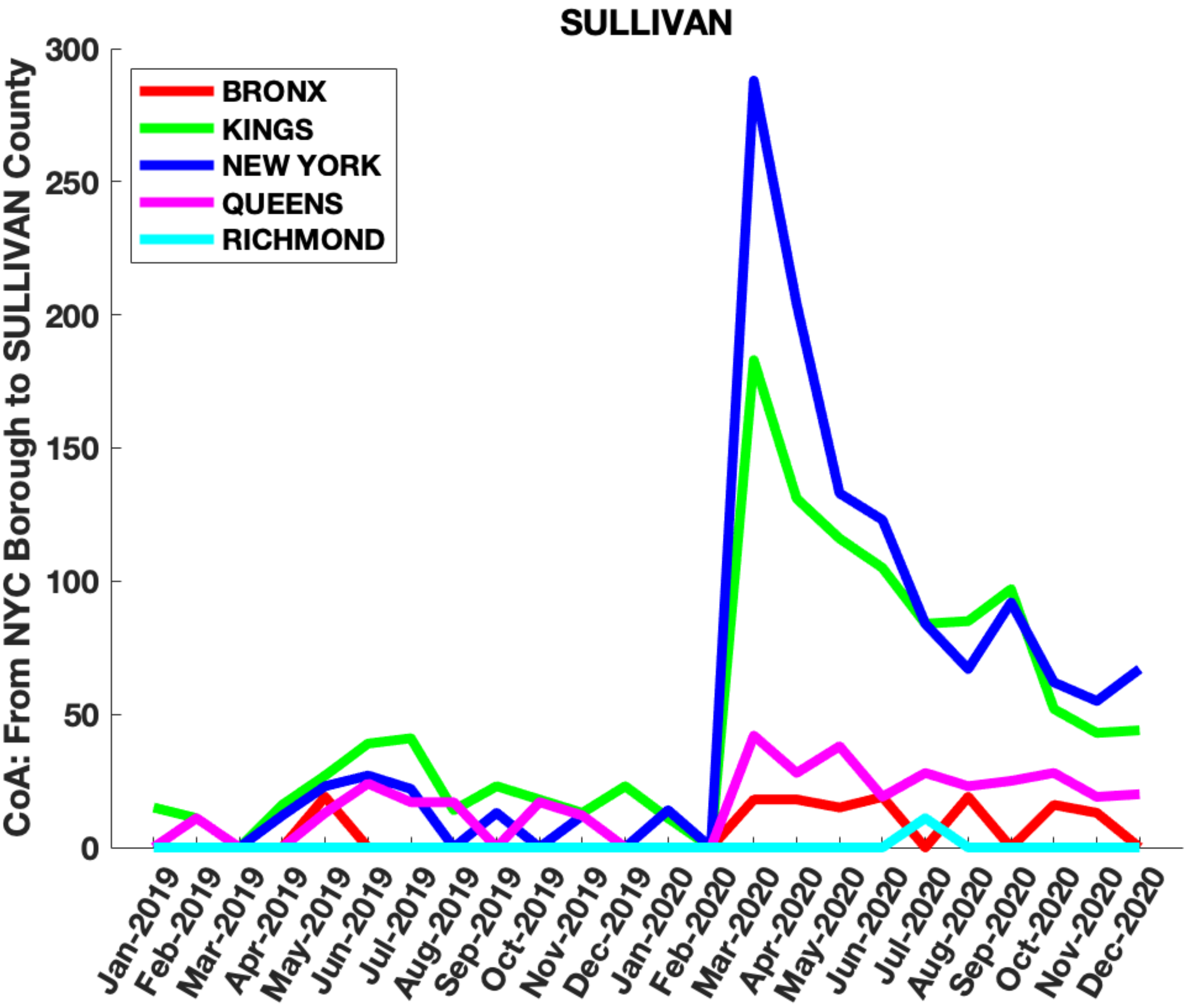

### Supplementary Figure 5

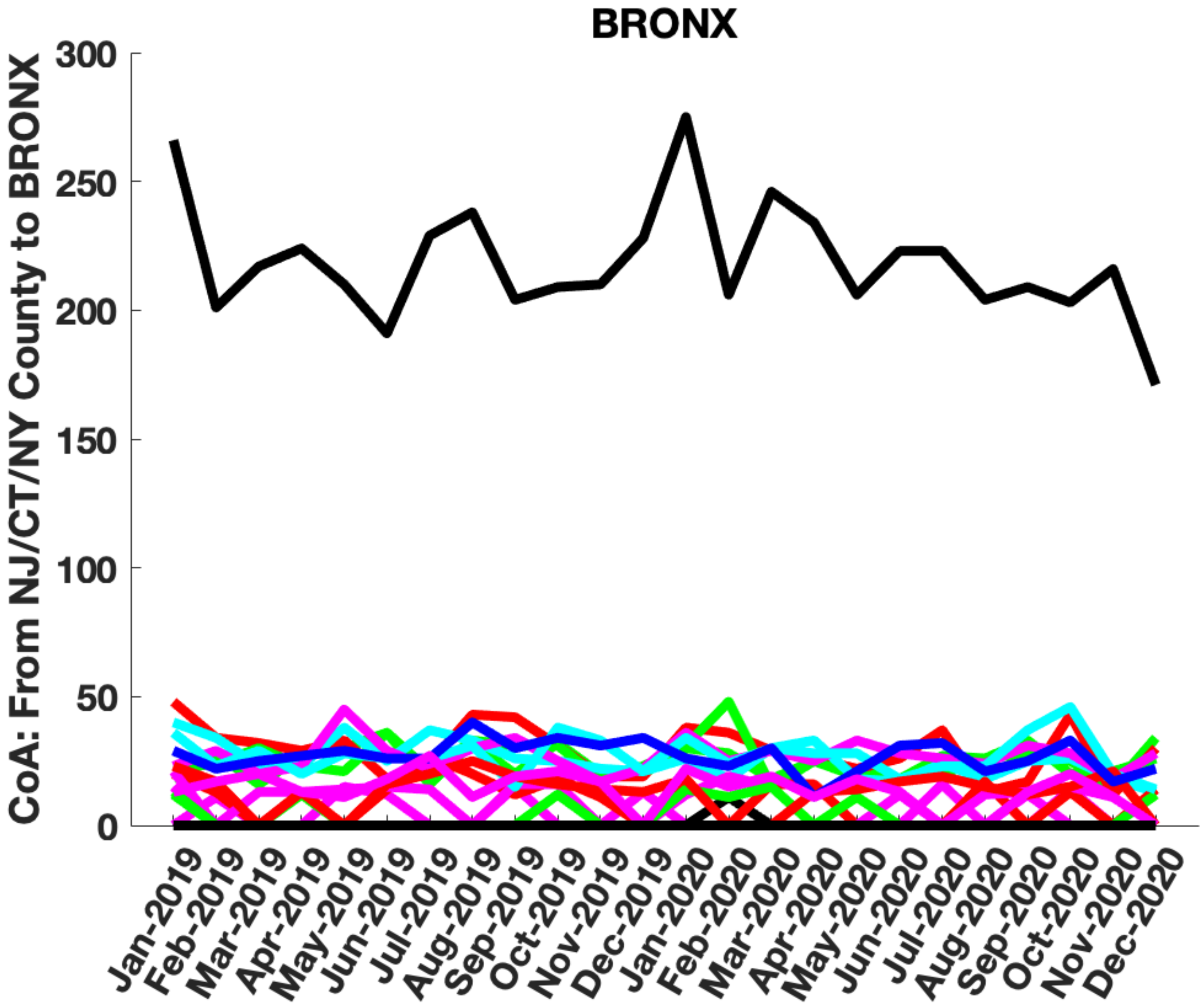

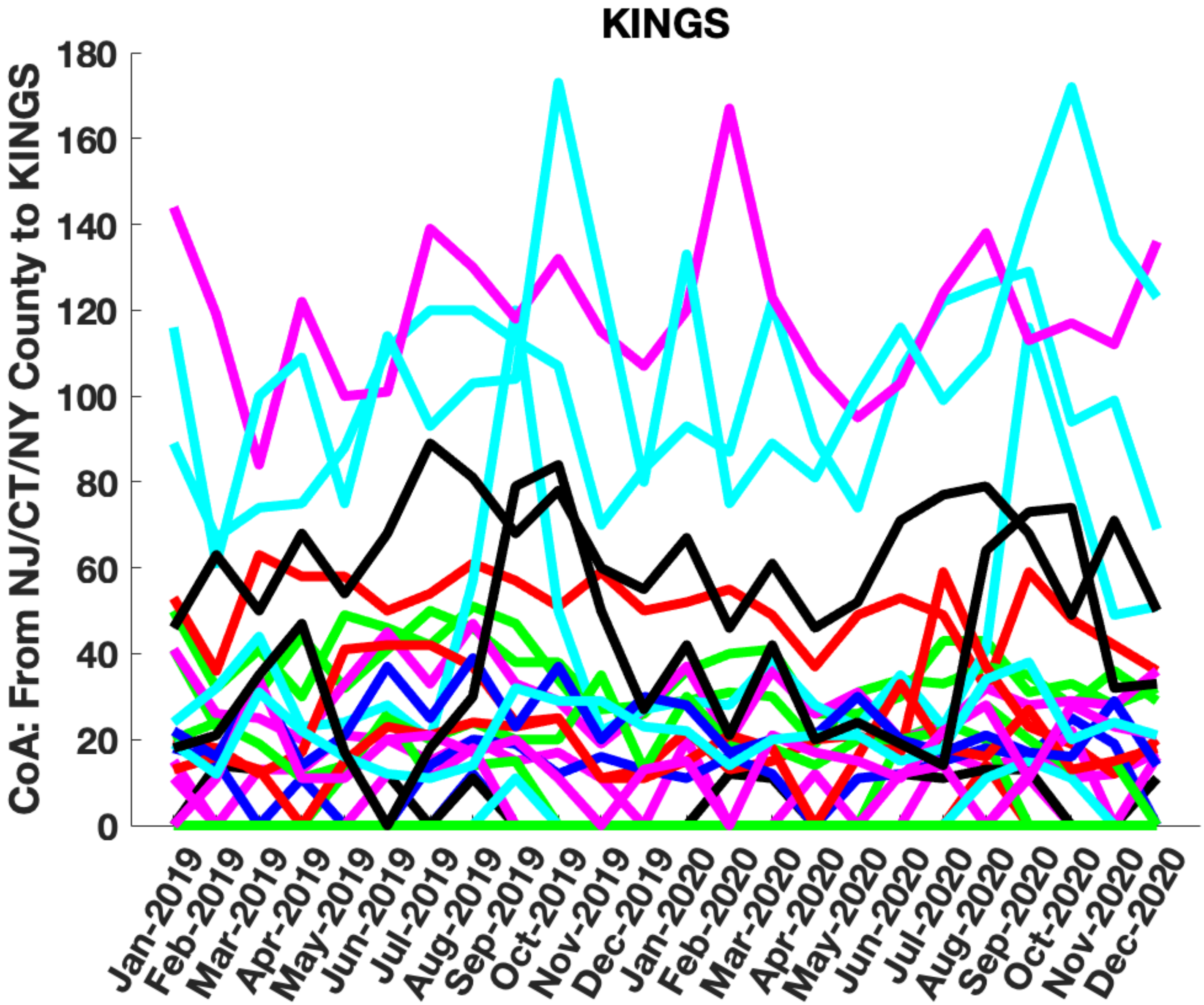

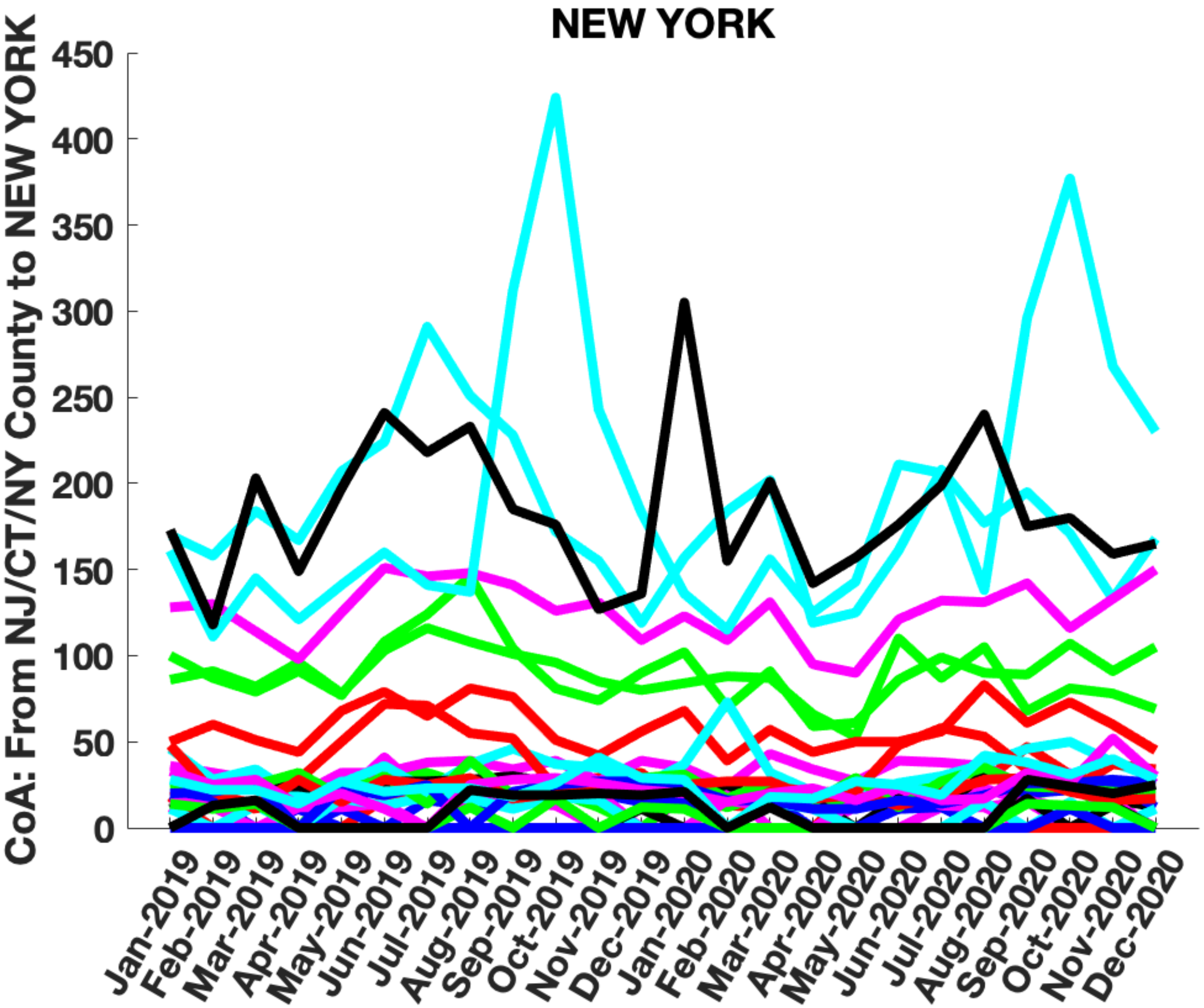

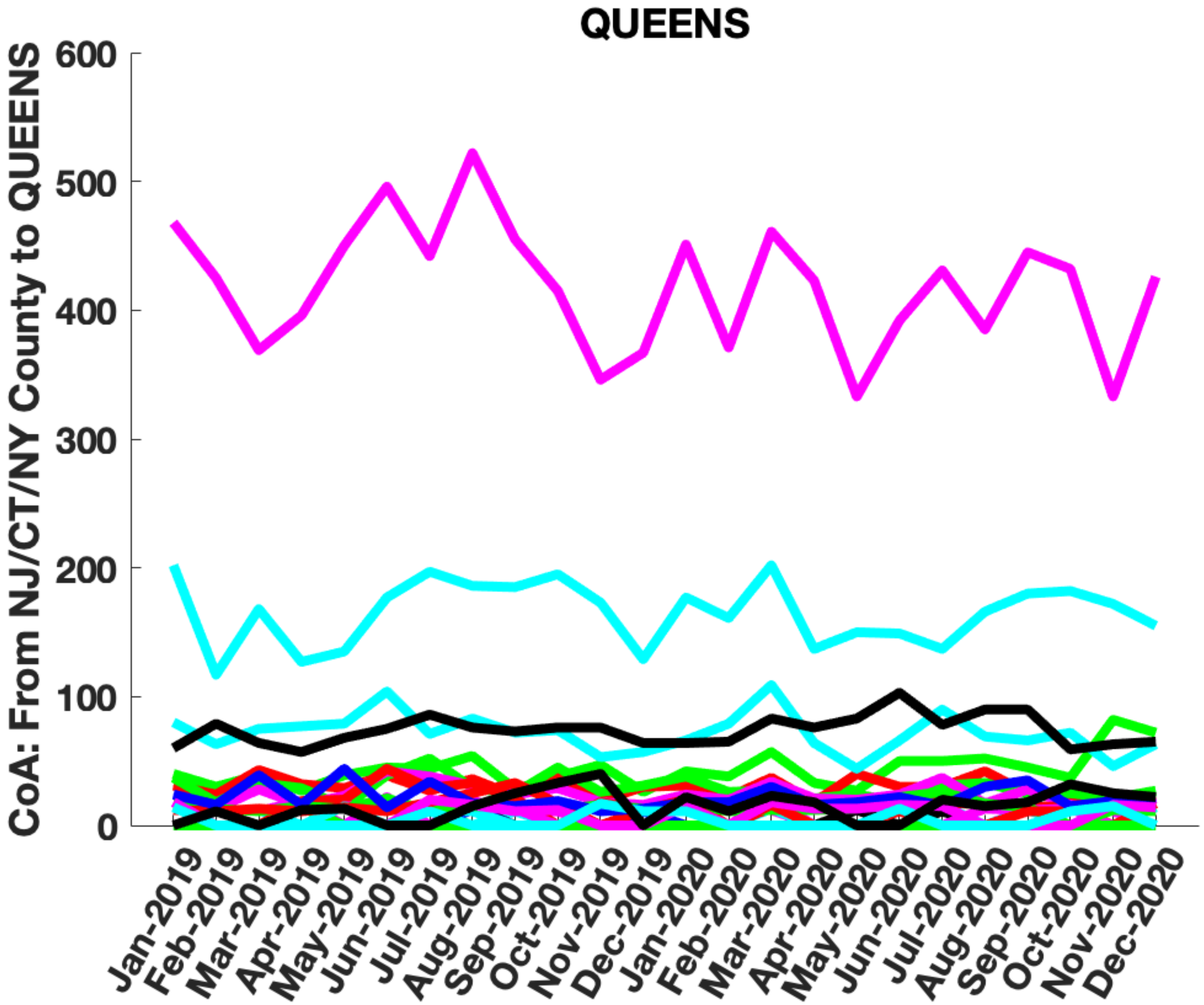

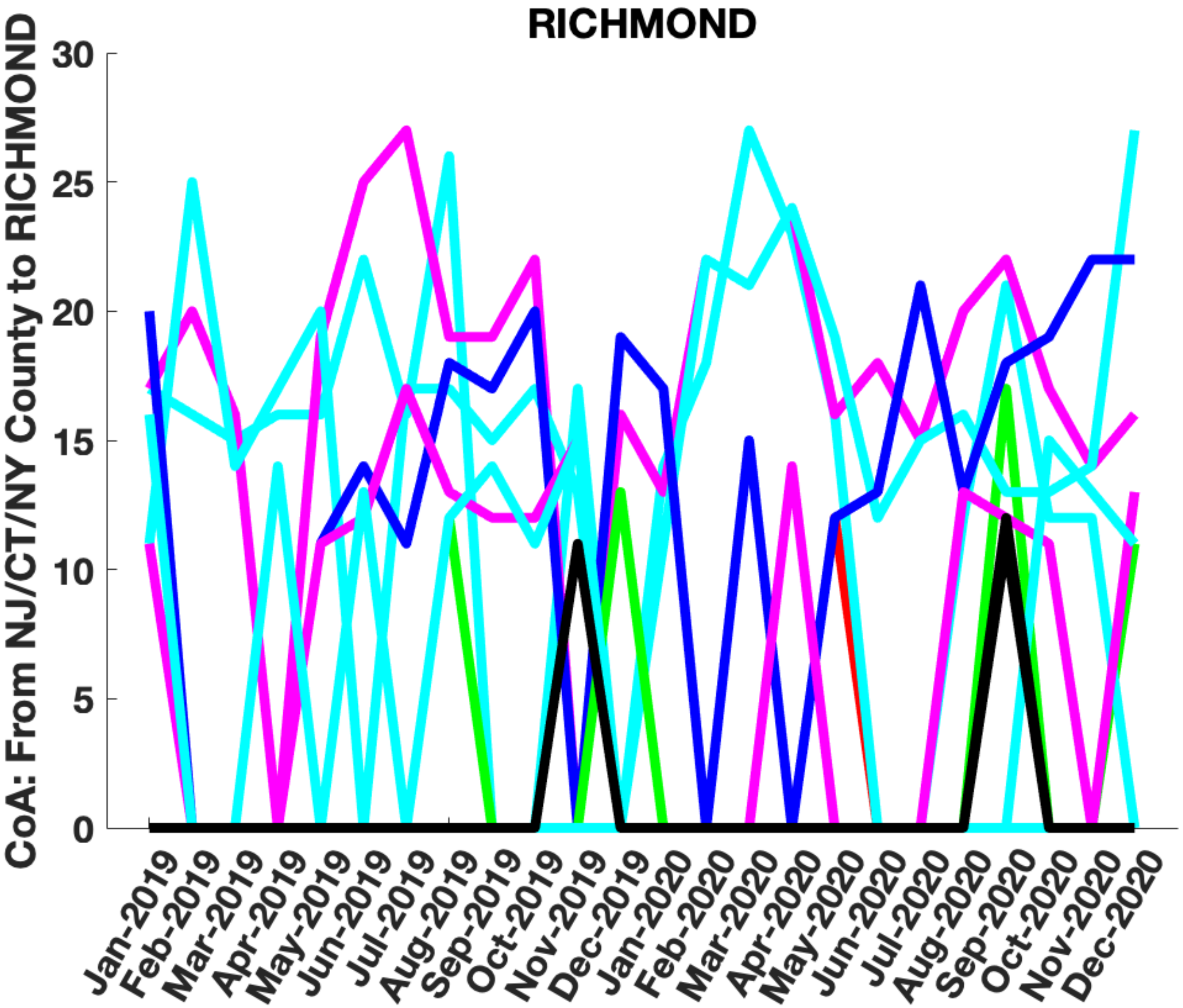

### Supplementary Figure 8

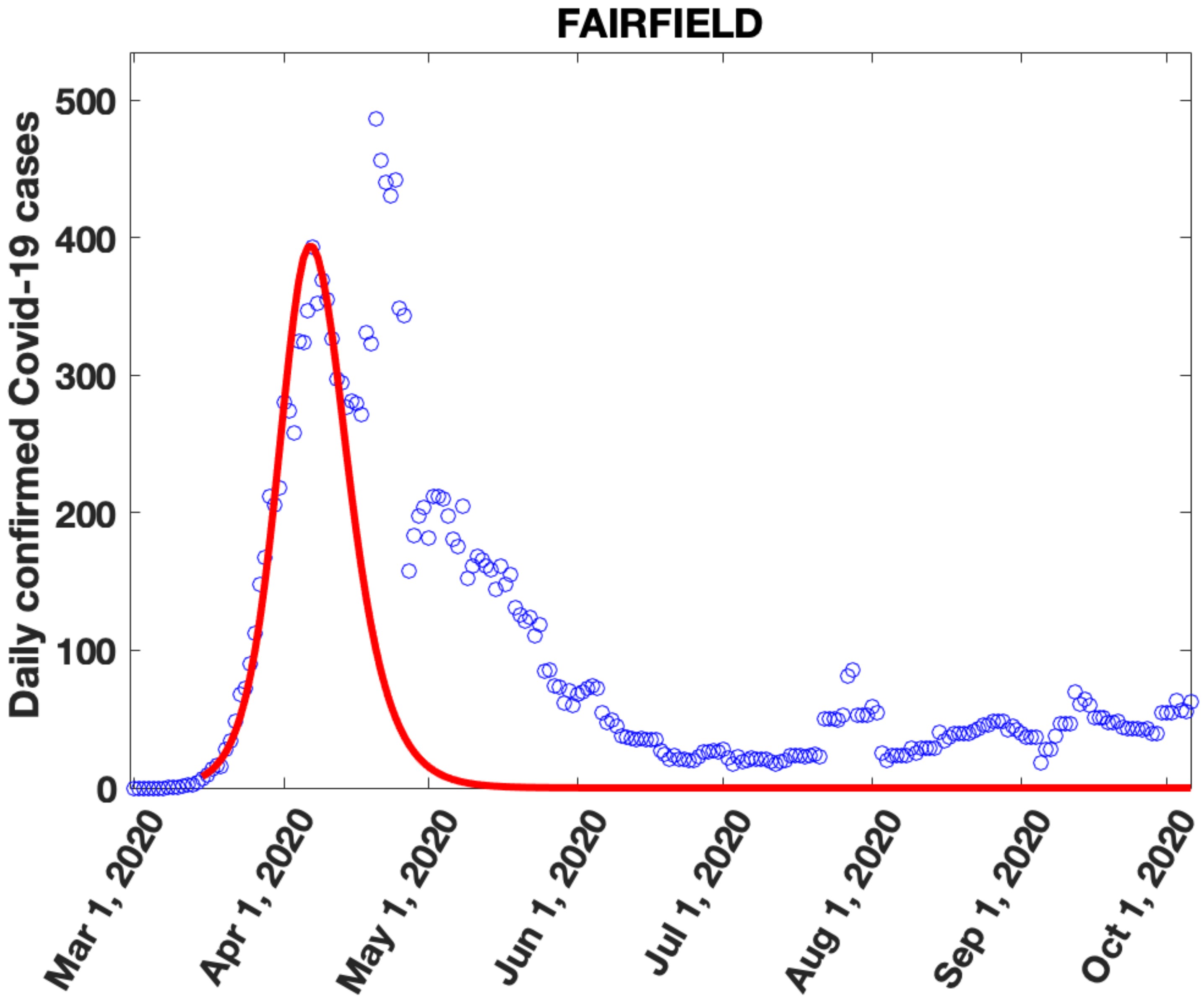

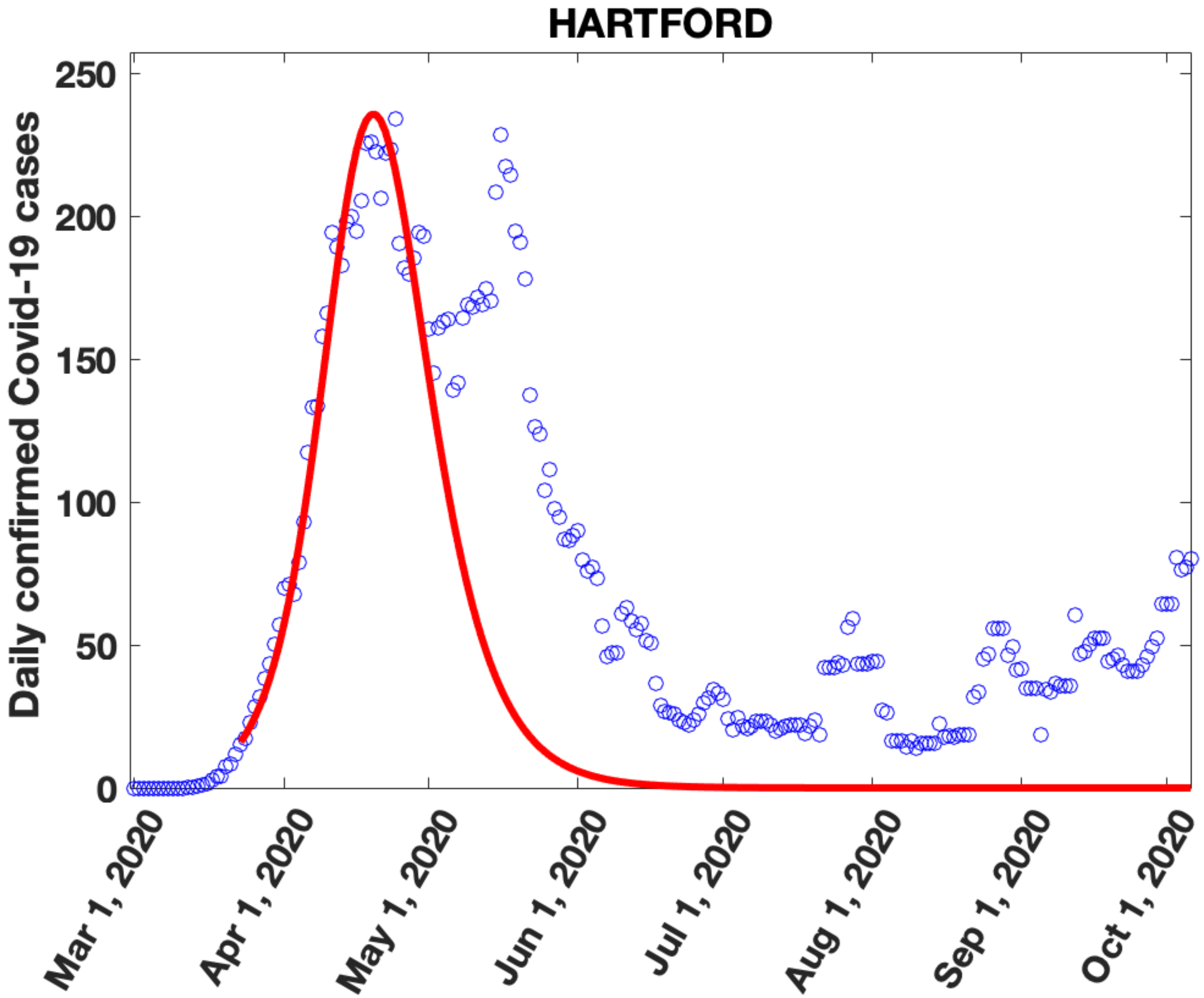

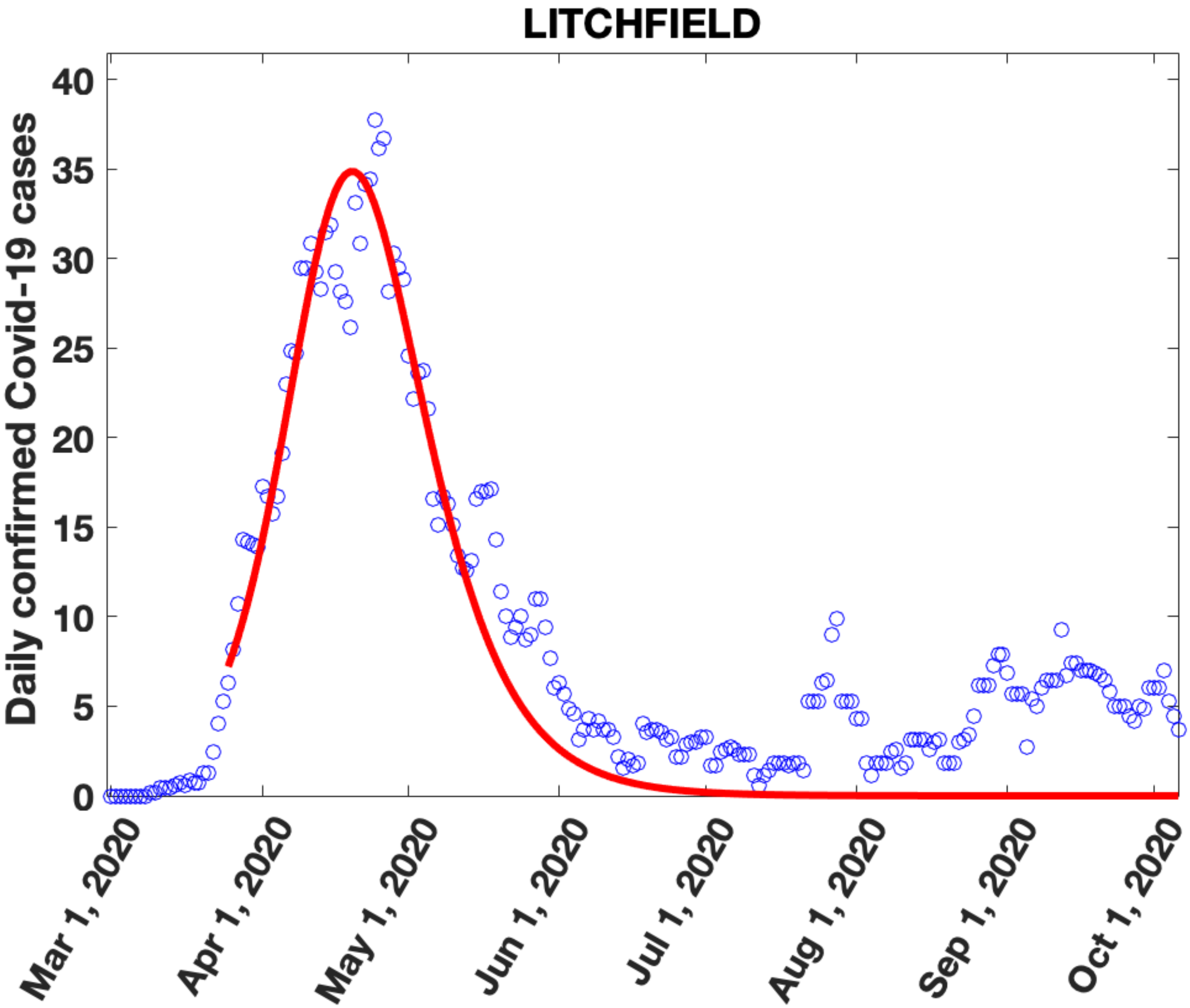

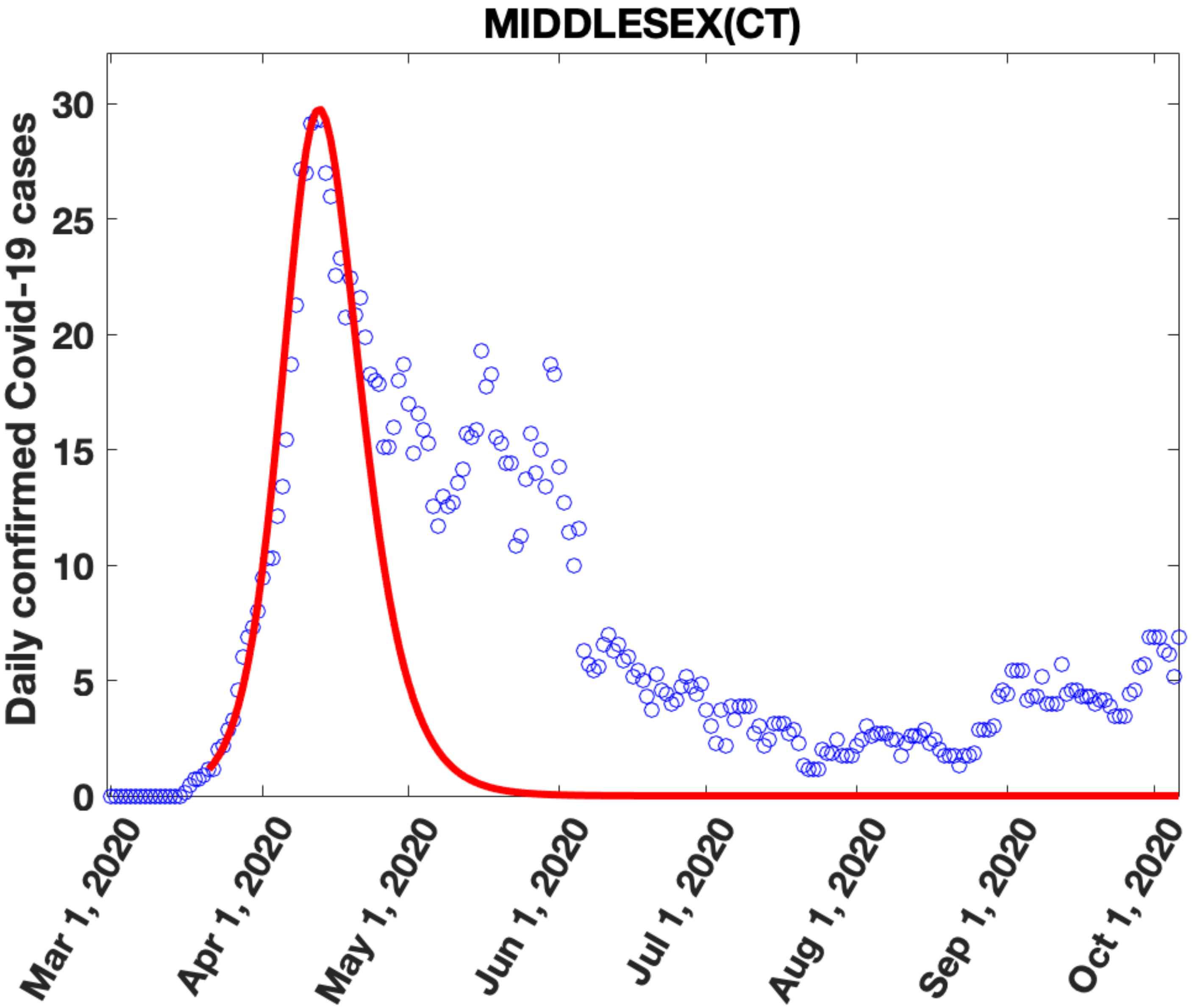

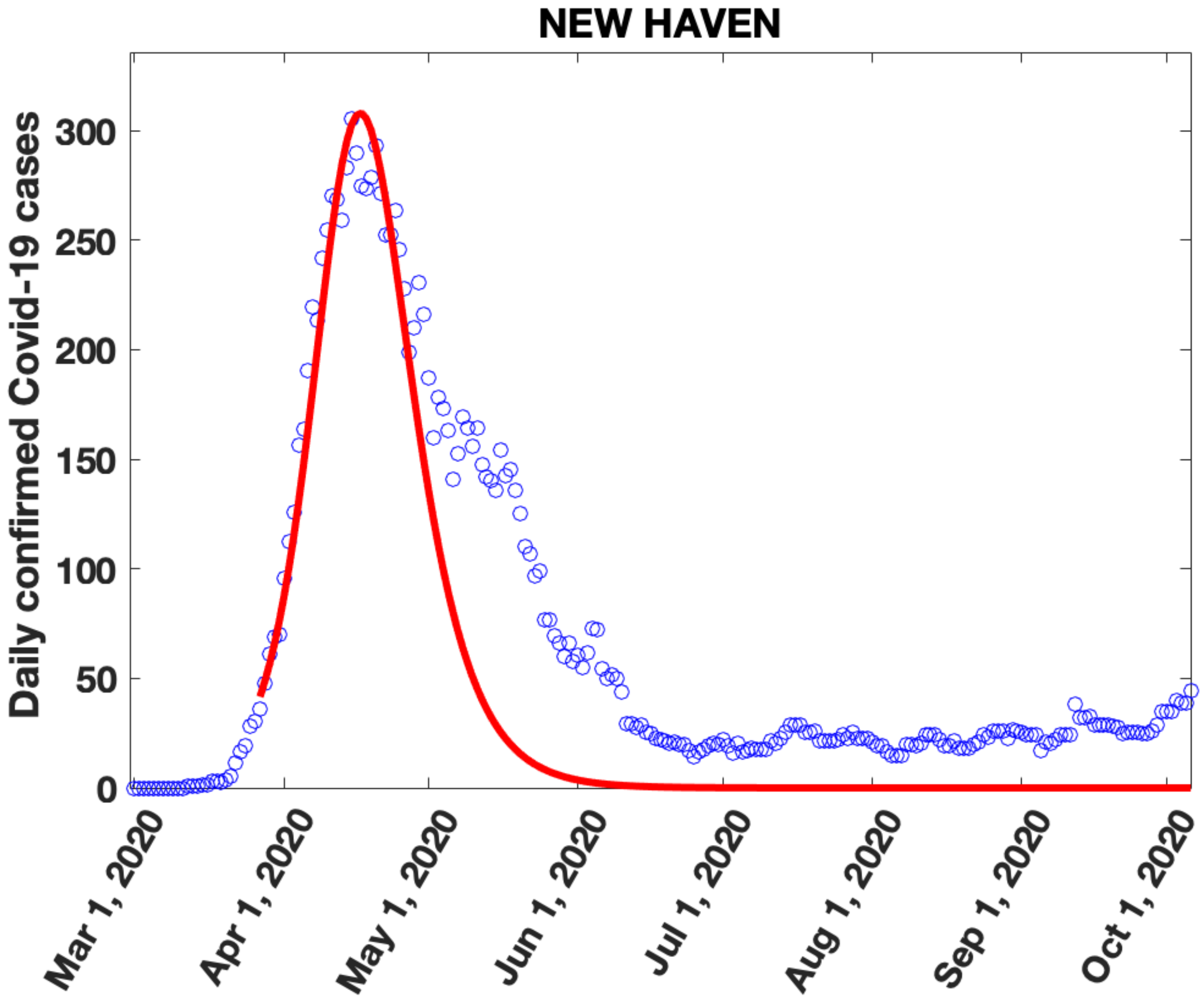

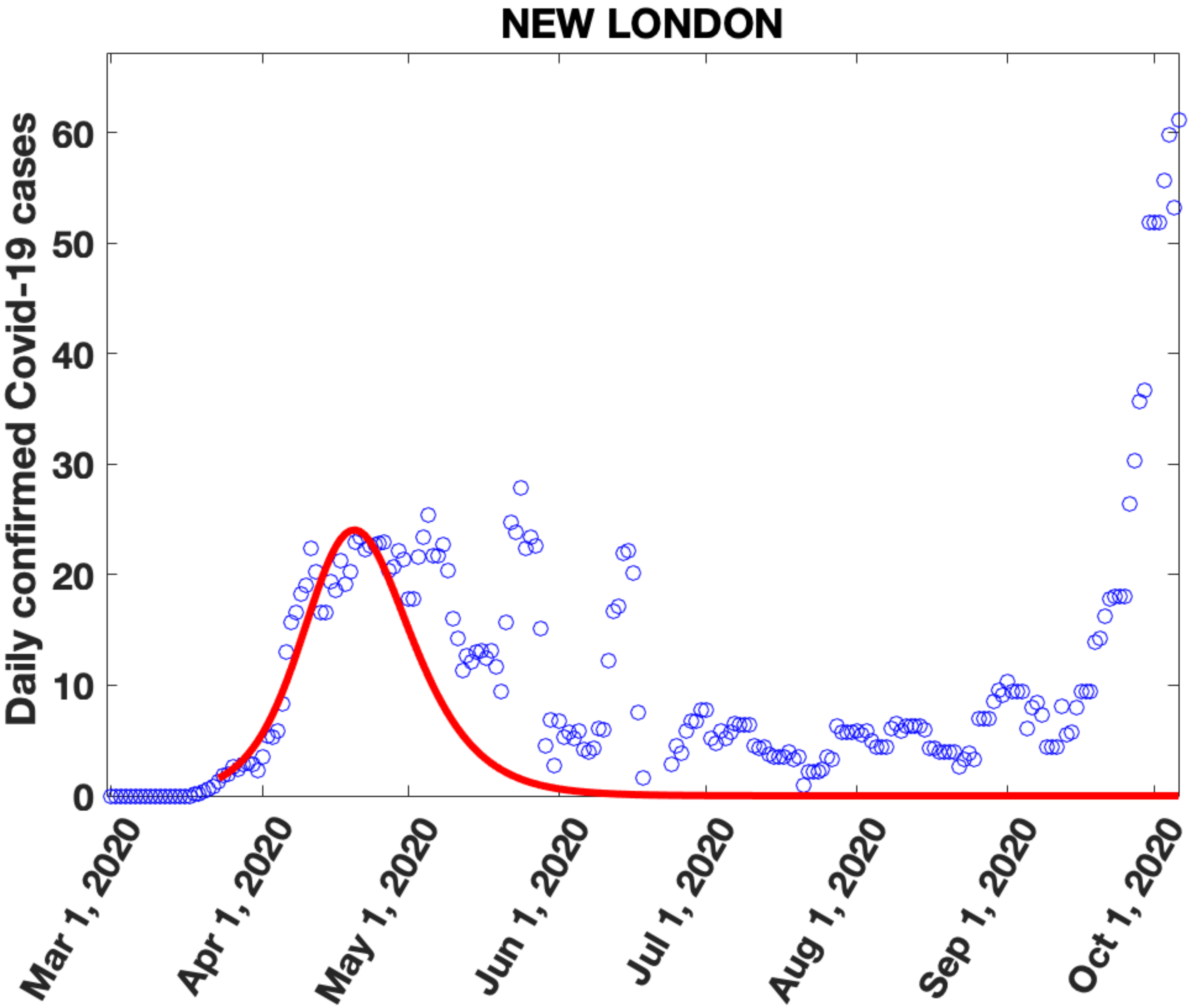

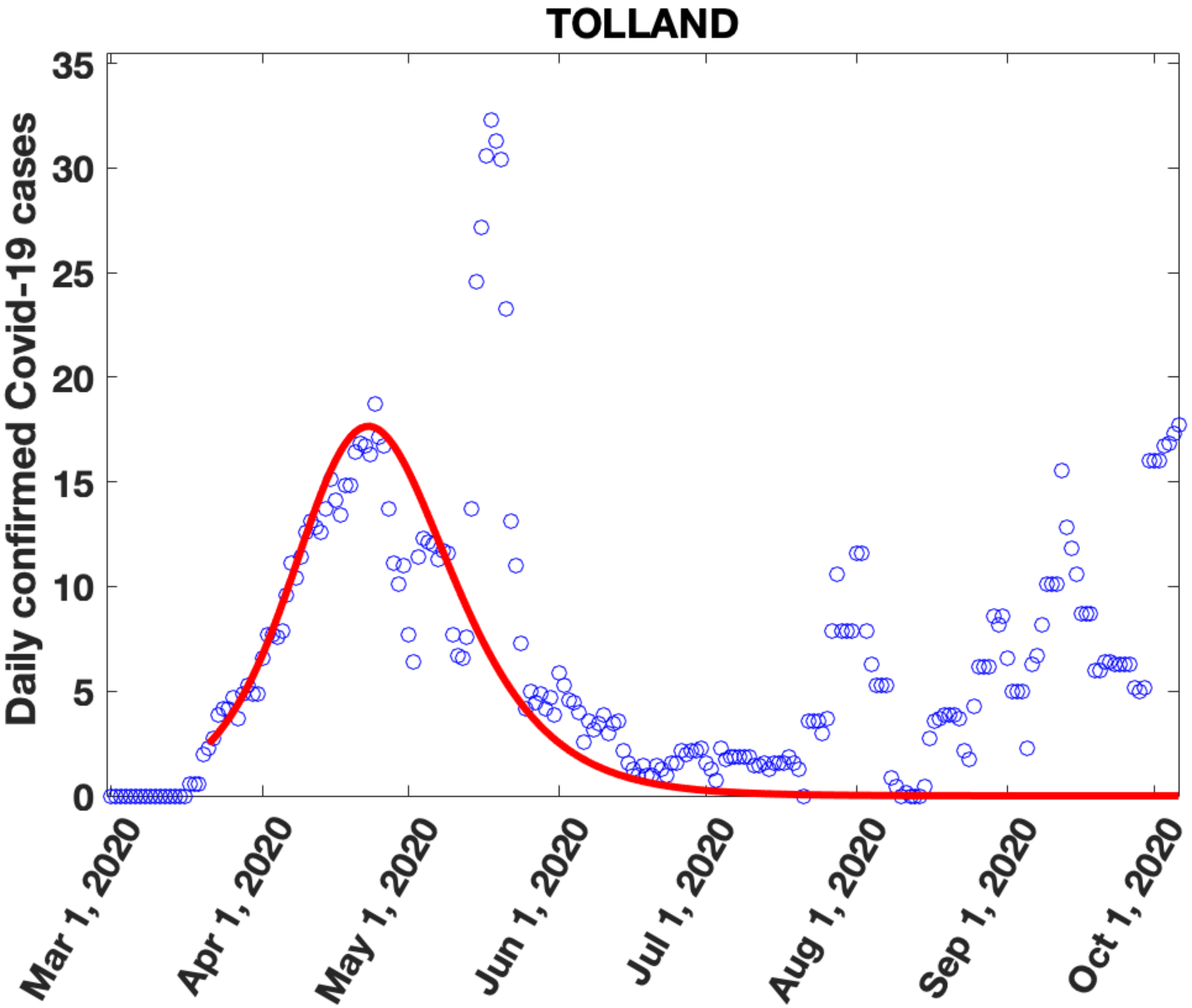

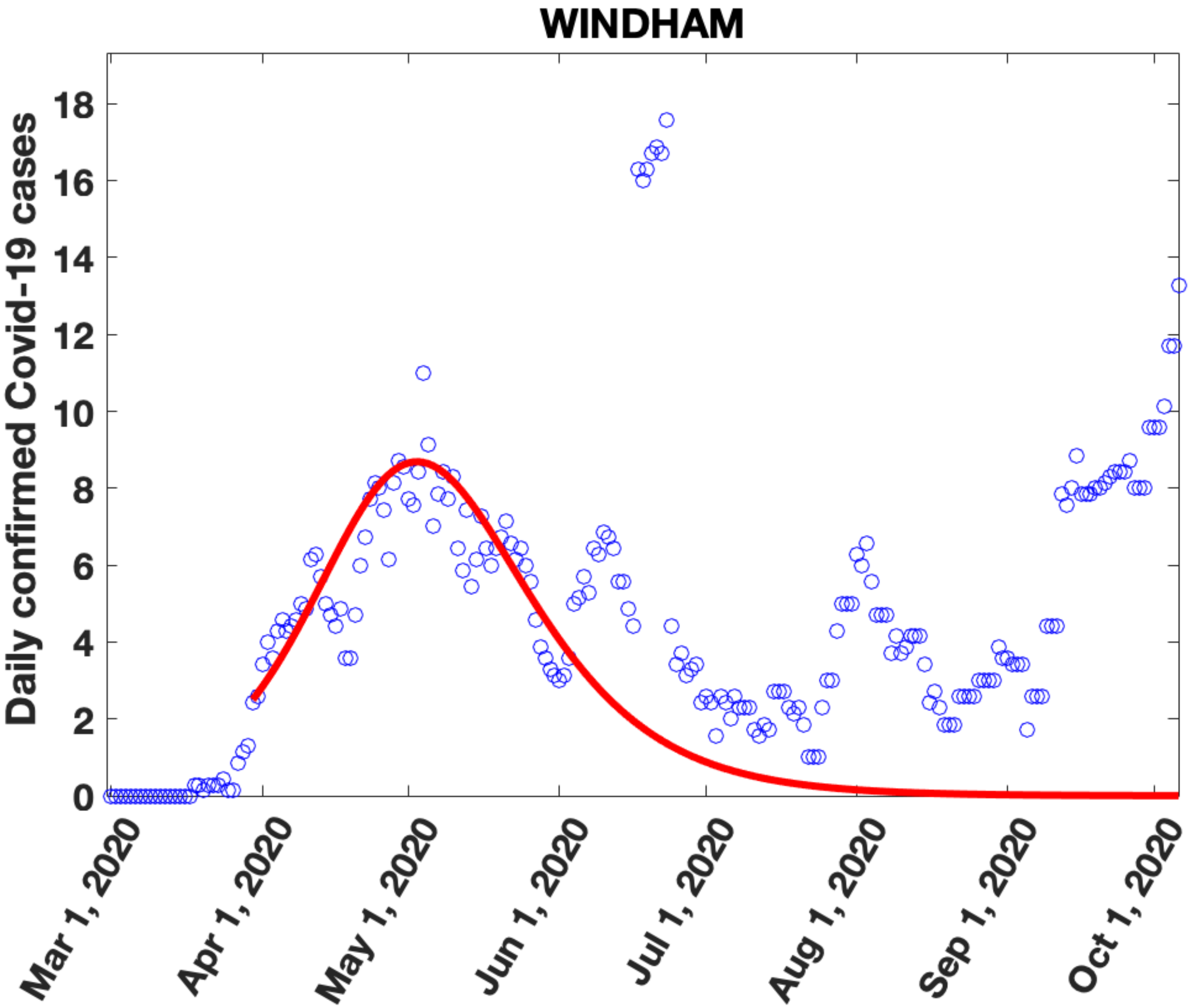

### Supplementary Figure 9

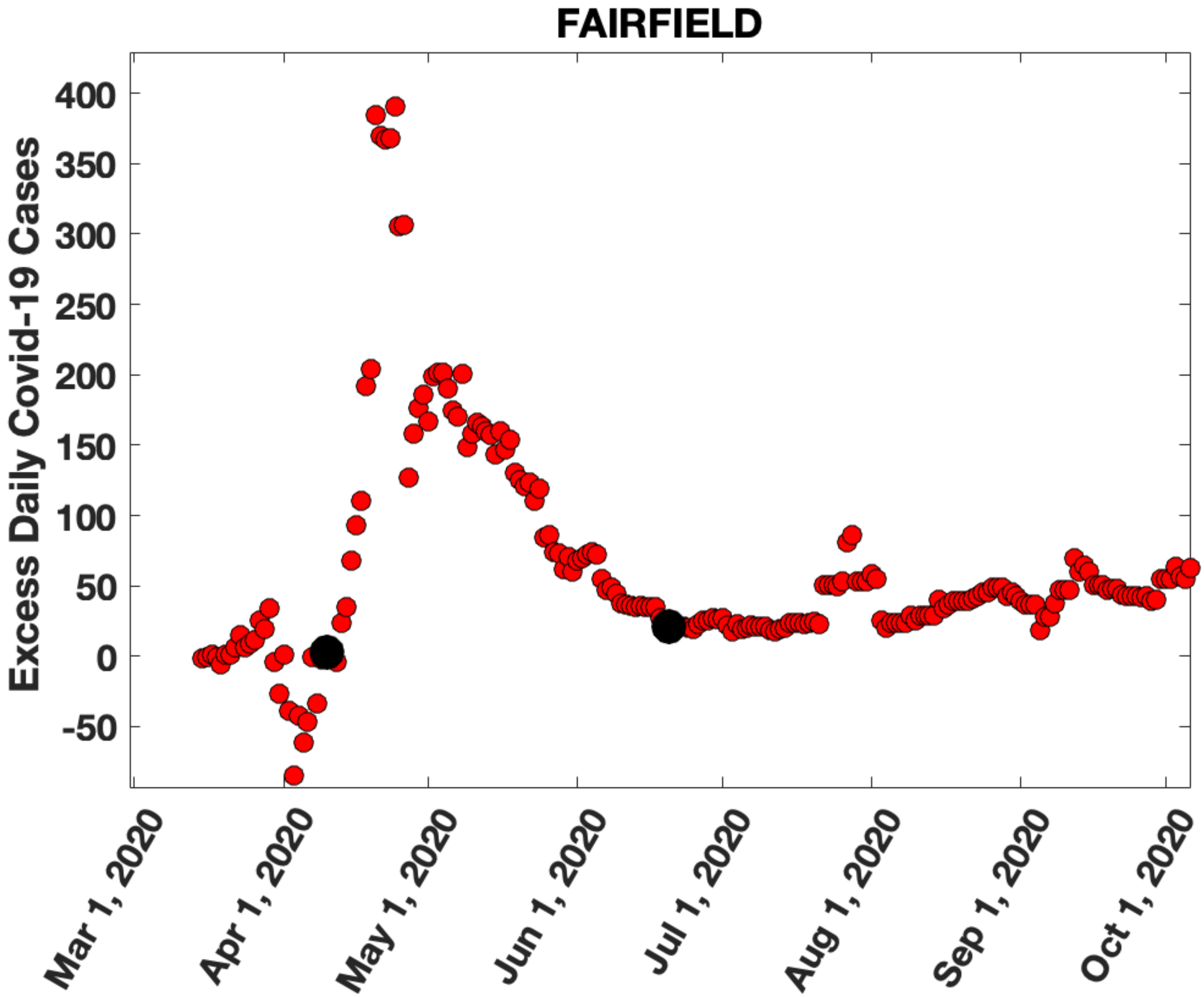

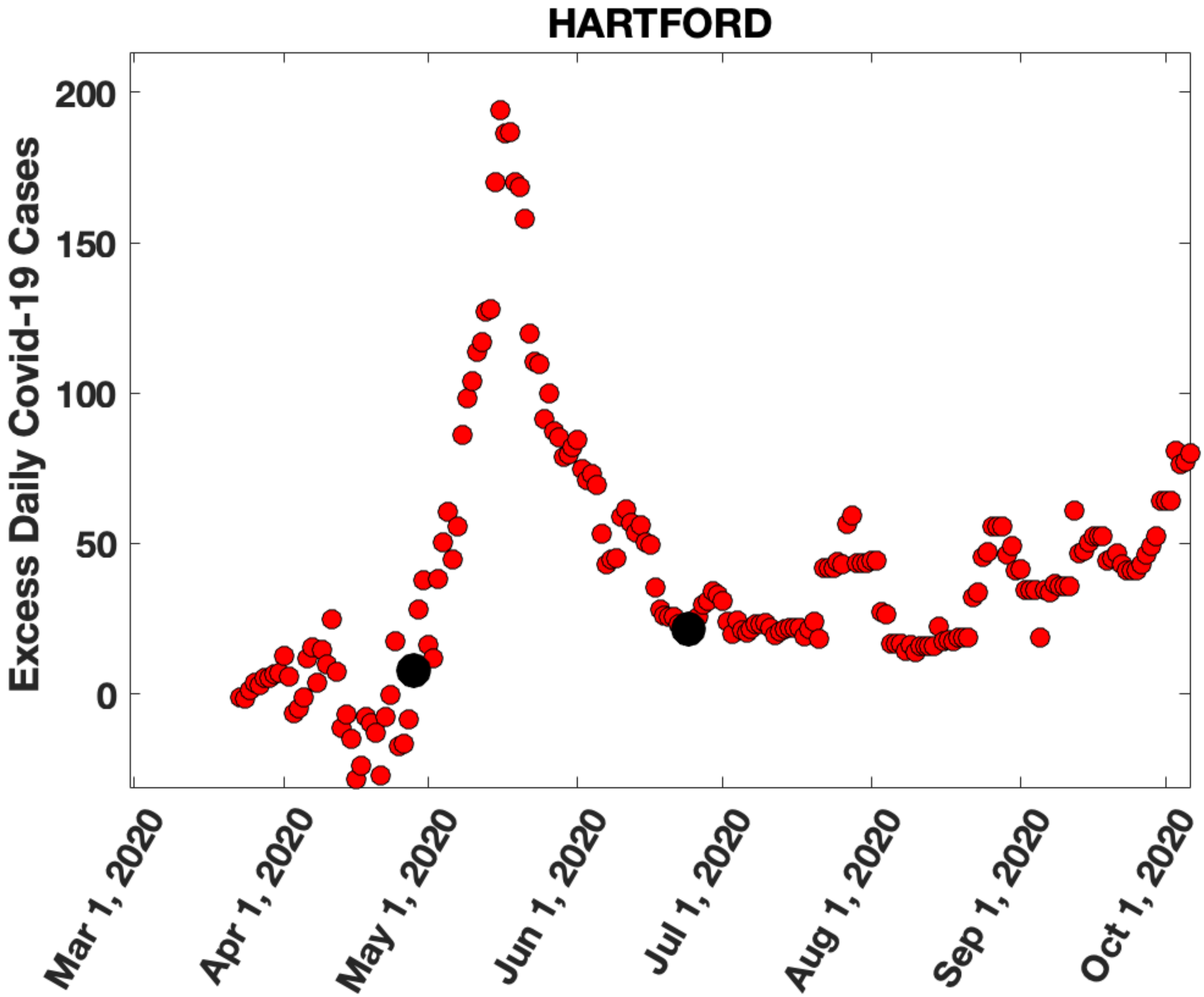

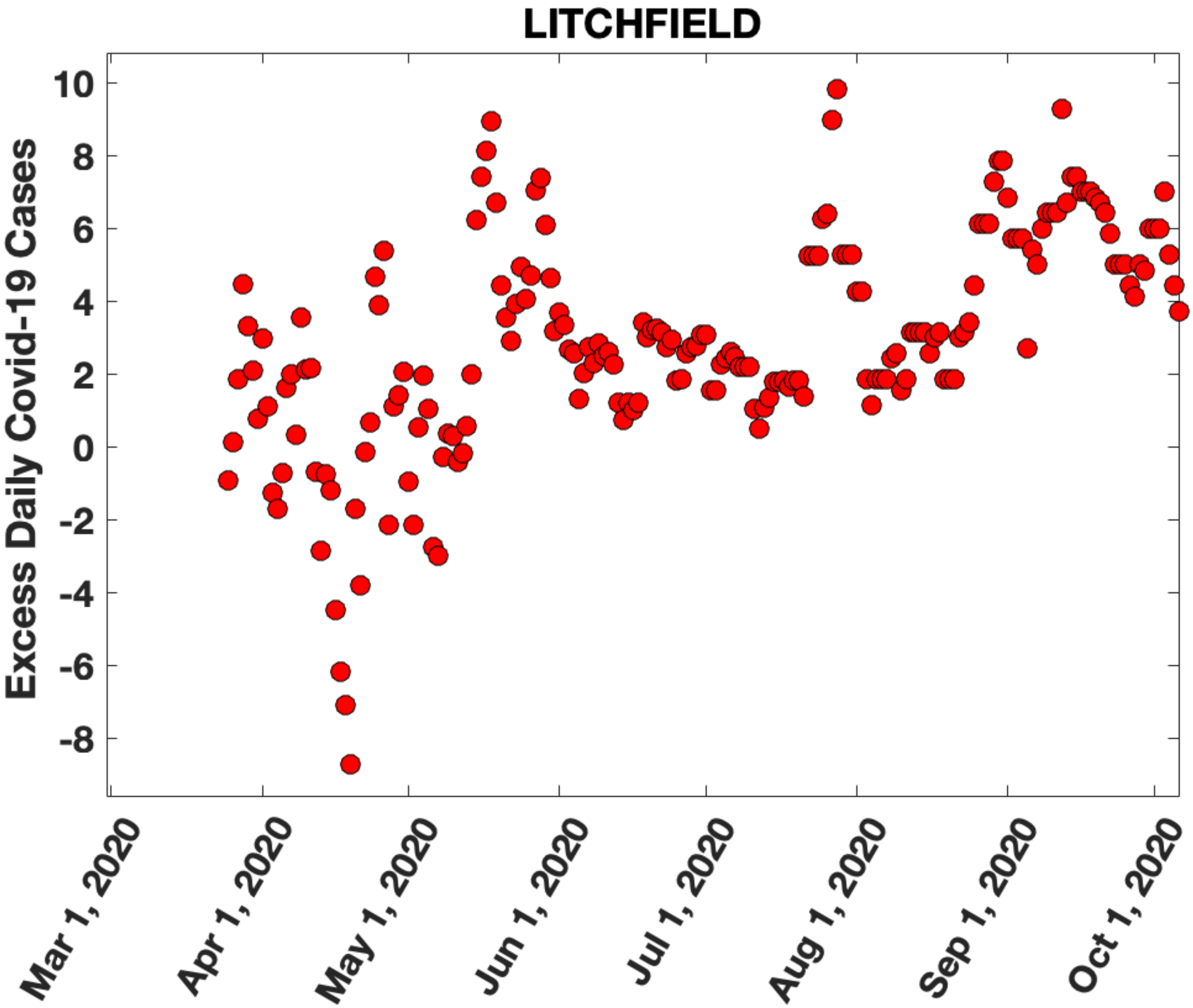

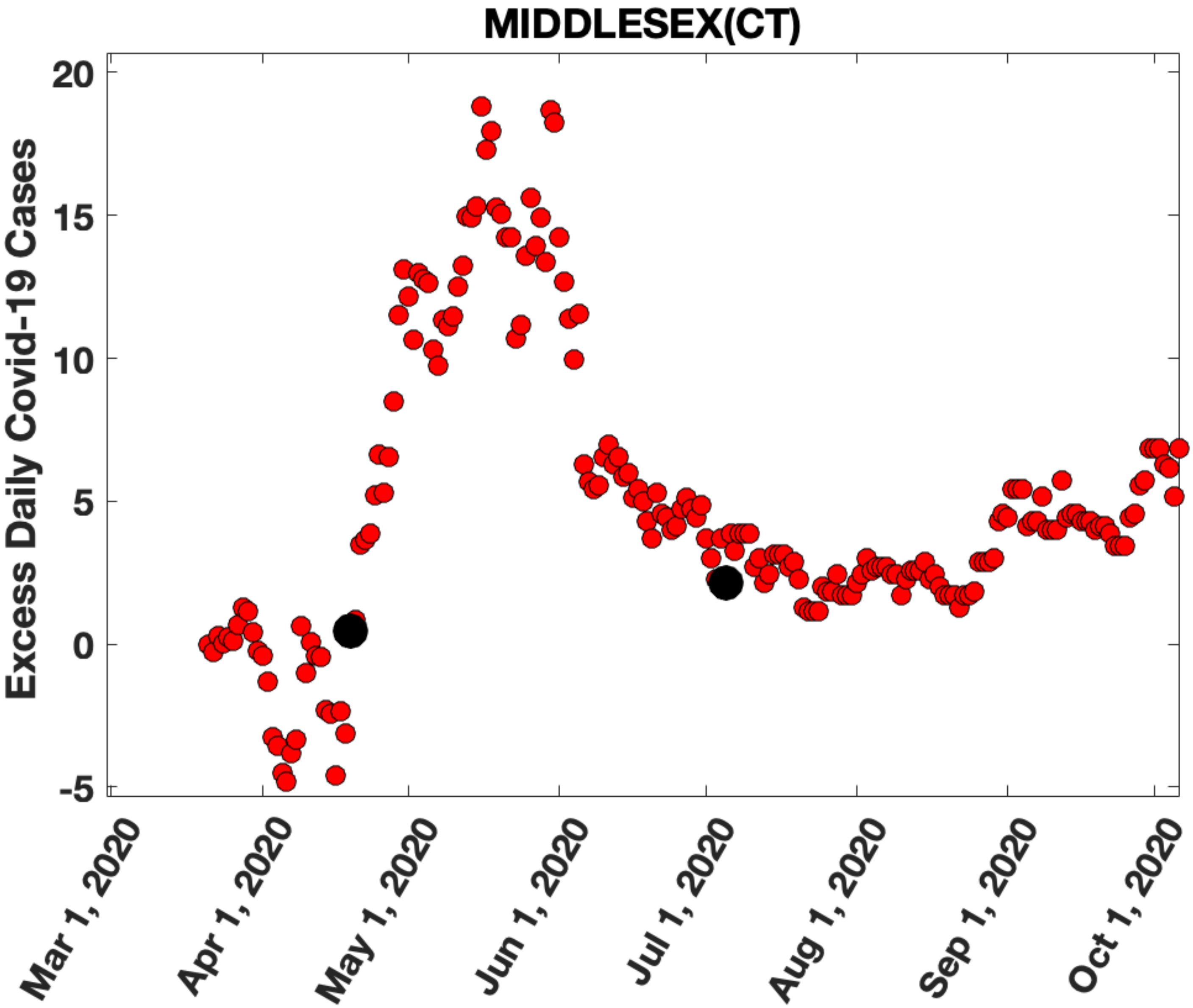

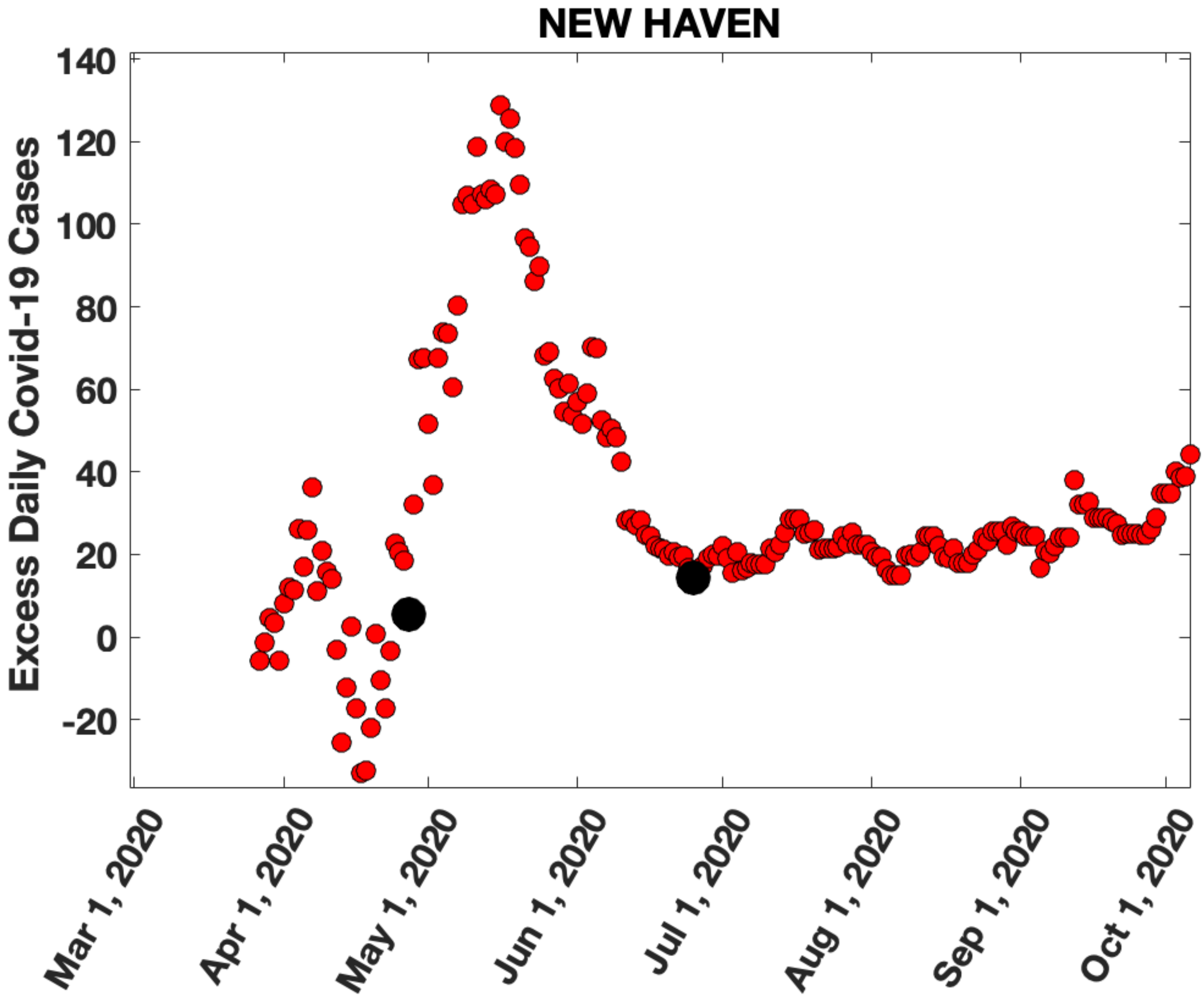

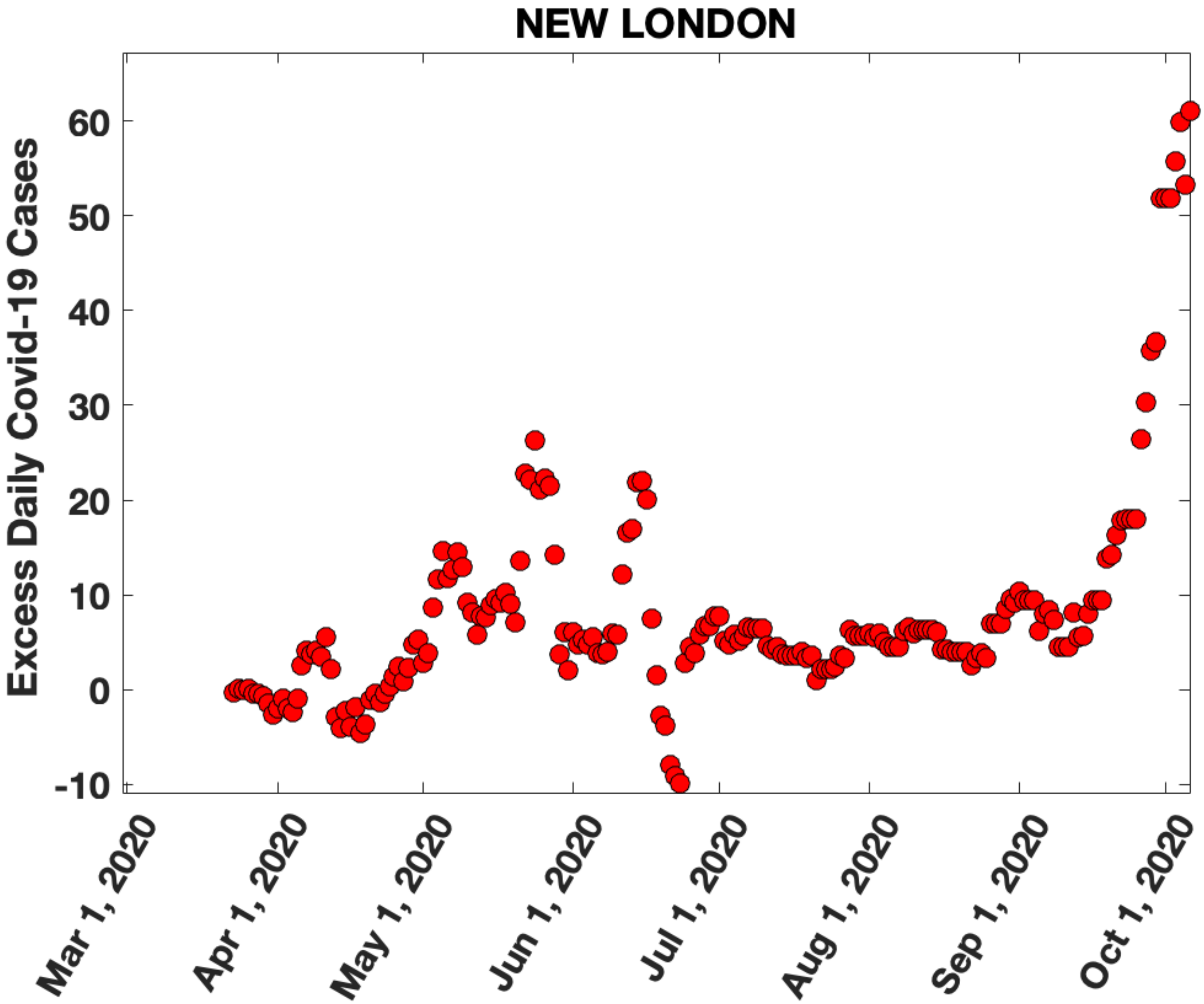

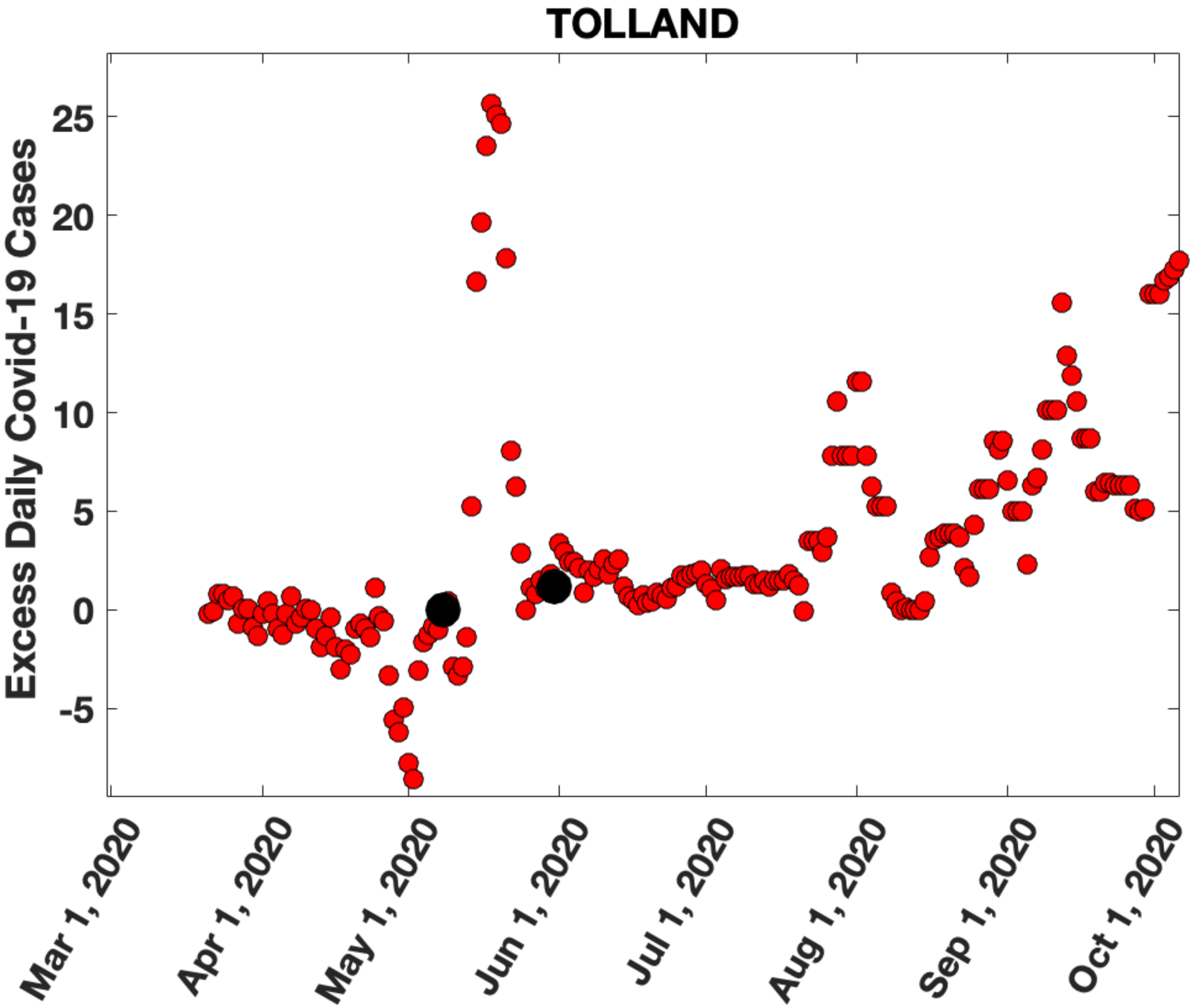

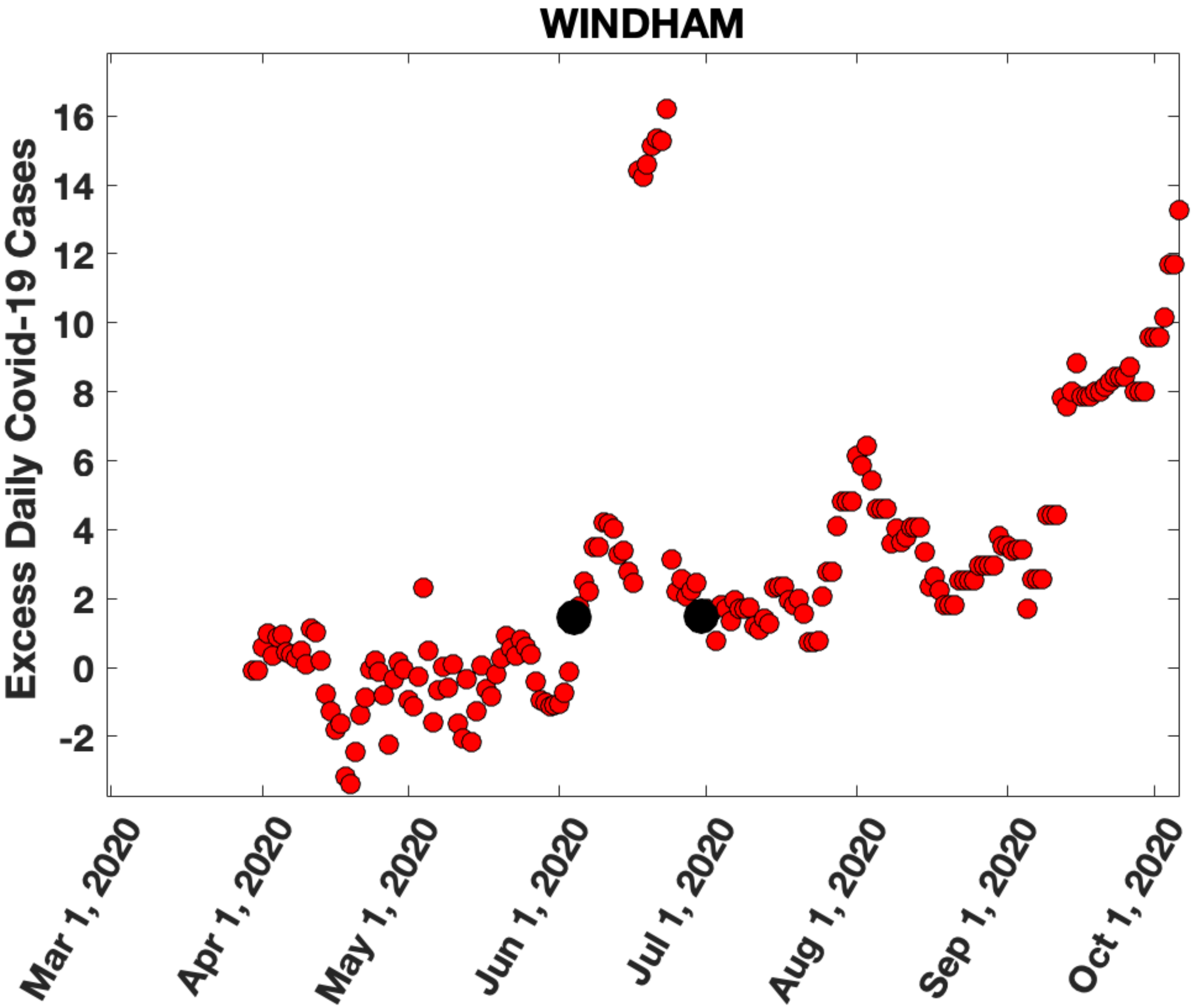
