## Supplementary Figure 2 for "Migration of households from New York City and the Second Peak in Covid-19 cases in New Jersey, Connecticut and New York Counties"

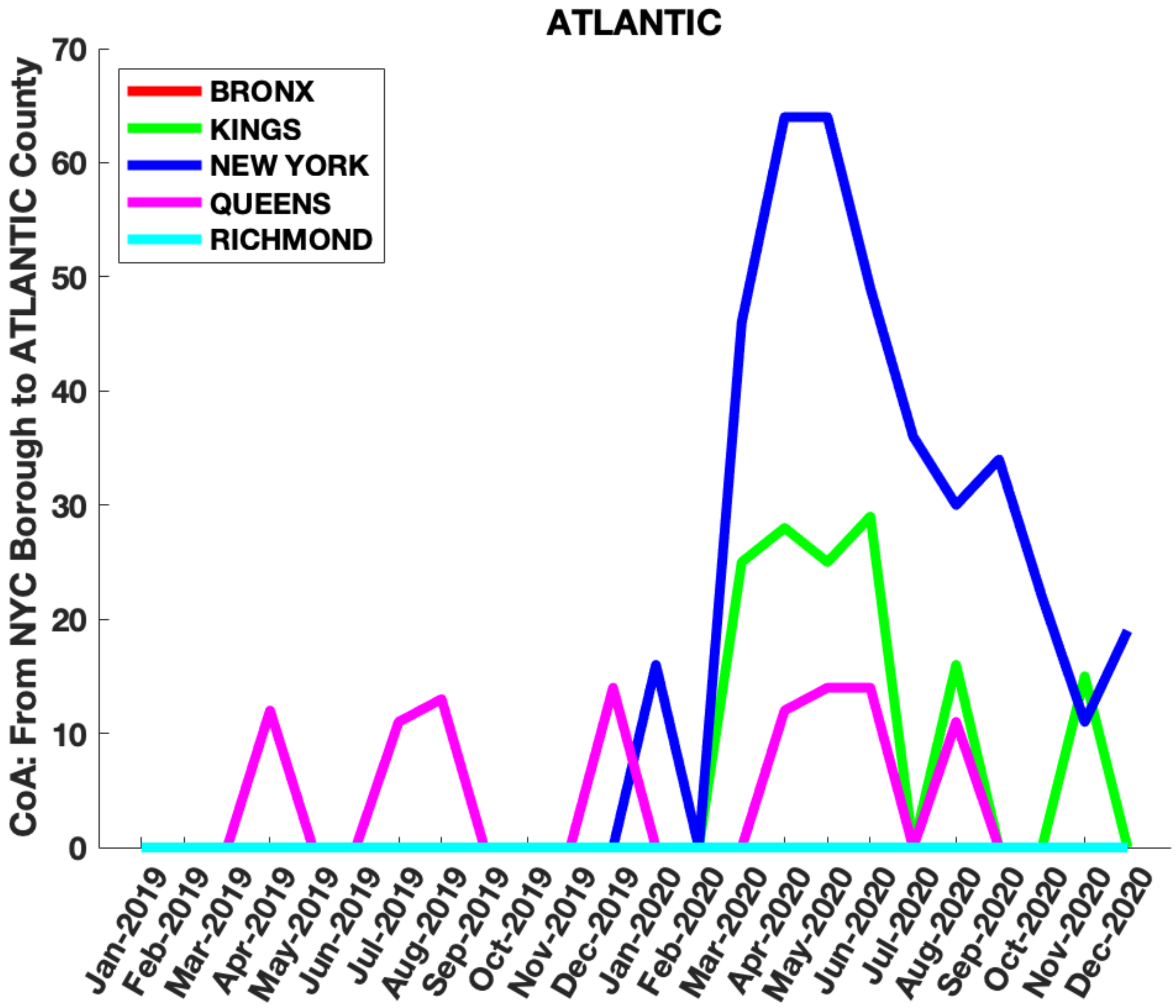
ATLANTIC
CoA: From NYC Borough to ATLANTIC County
BRONX
KINGS
NEW YORK
QUEENS
RICHMOND
0
10
20
30
40
50
60
70
Jan-2019
Feb-2019
Mar-2019
Apr-2019
May-2019
Jun-2019
Jul-2019
Aug-2019
Sep-2019
Oct-2019
Nov-2019
Dec-2019
Jan-2020
Feb-2020
Mar-2020
Apr-2020
May-2020
Jun-2020
Jul-2020
Aug-2020
Sep-2020
Oct-2020
Nov-2020
Dec-2020

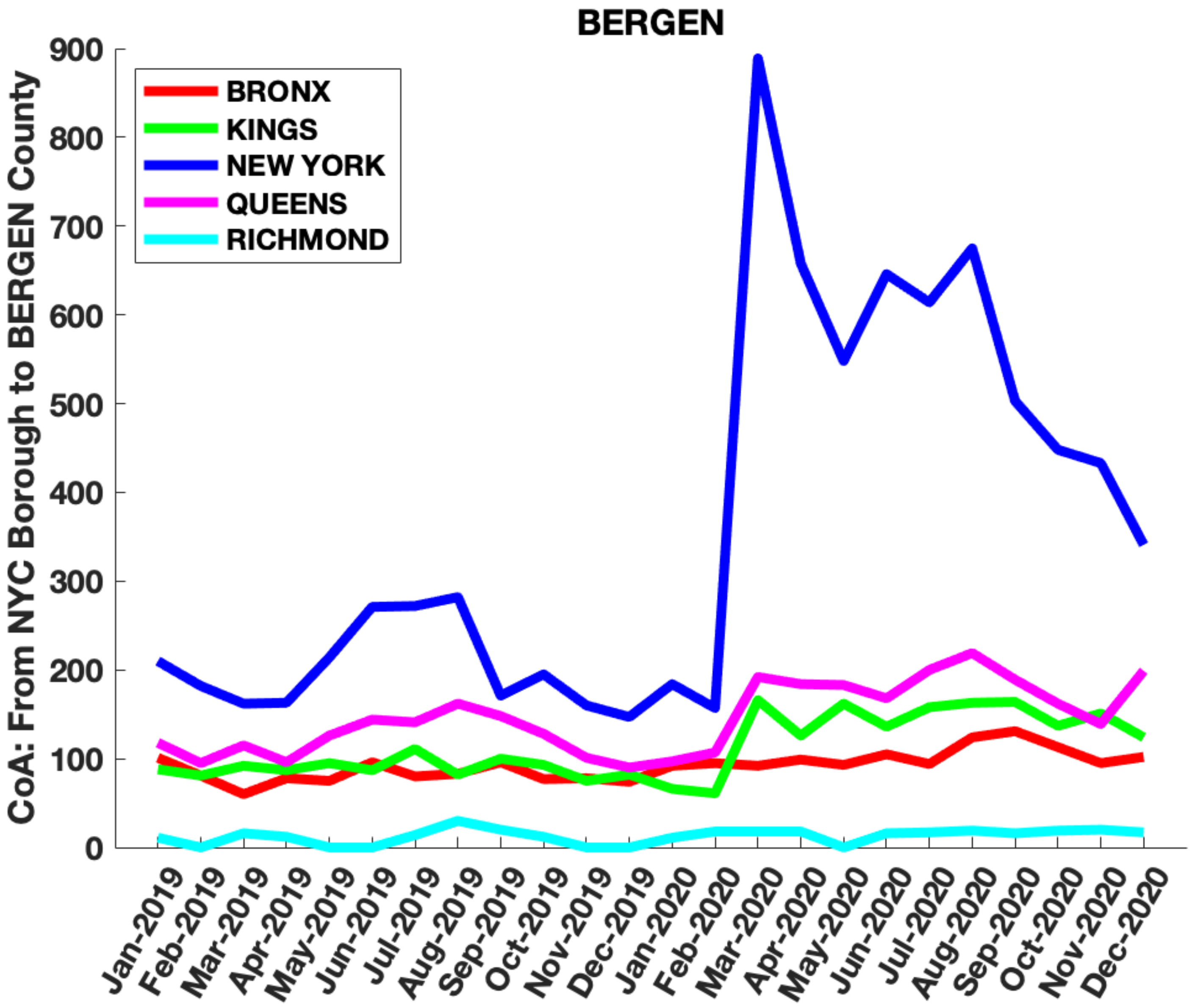

BERGEN
CoA: From NYC Borough to BERGEN County
BRONX
KINGS
NEW YORK
QUEENS
RICHMOND
0
100
200
300
400
500
600
700
800
900
Jan-2019
Feb-2019
Mar-2019
Apr-2019
May-2019
Jun-2019
Jul-2019
Aug-2019
Sep-2019
Oct-2019
Nov-2019
Dec-2019
Jan-2020
Feb-2020
Mar-2020
Apr-2020
May-2020
Jun-2020
Jul-2020
Aug-2020
Sep-2020
Oct-2020
Nov-2020
Dec-2020

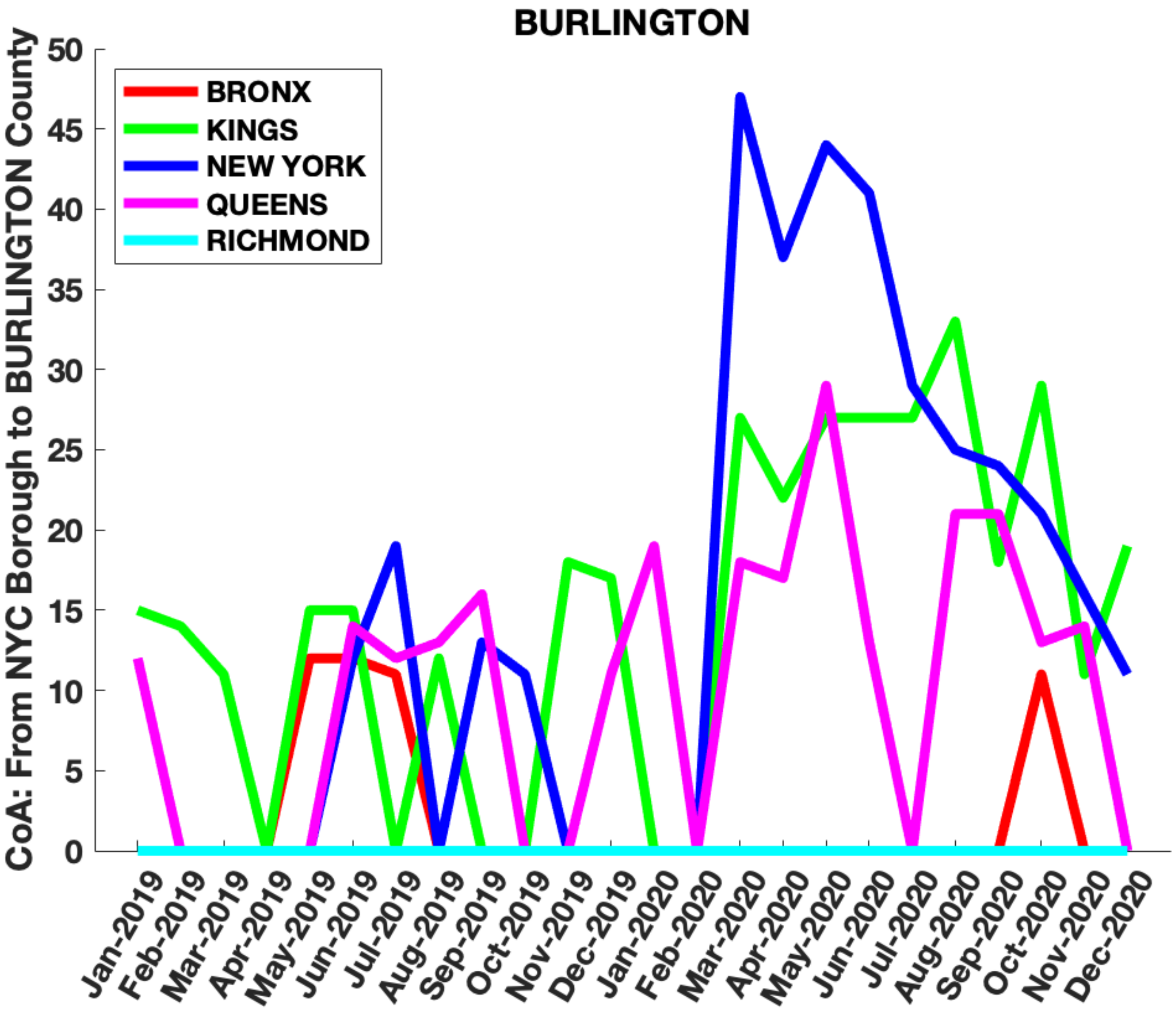

BURLINGTON
CoA: From NYC Borough to BURLINGTON County
BRONX
KINGS
NEW YORK
QUEENS
RICHMOND
0
5
10
15
20
25
30
35
40
45
50
Jan-2019
Feb-2019
Mar-2019
Apr-2019
May-2019
Jun-2019
Jul-2019
Aug-2019
Sep-2019
Oct-2019
Nov-2019
Dec-2019
Jan-2020
Feb-2020
Mar-2020
Apr-2020
May-2020
Jun-2020
Jul-2020
Aug-2020
Sep-2020
Oct-2020
Nov-2020
Dec-2020

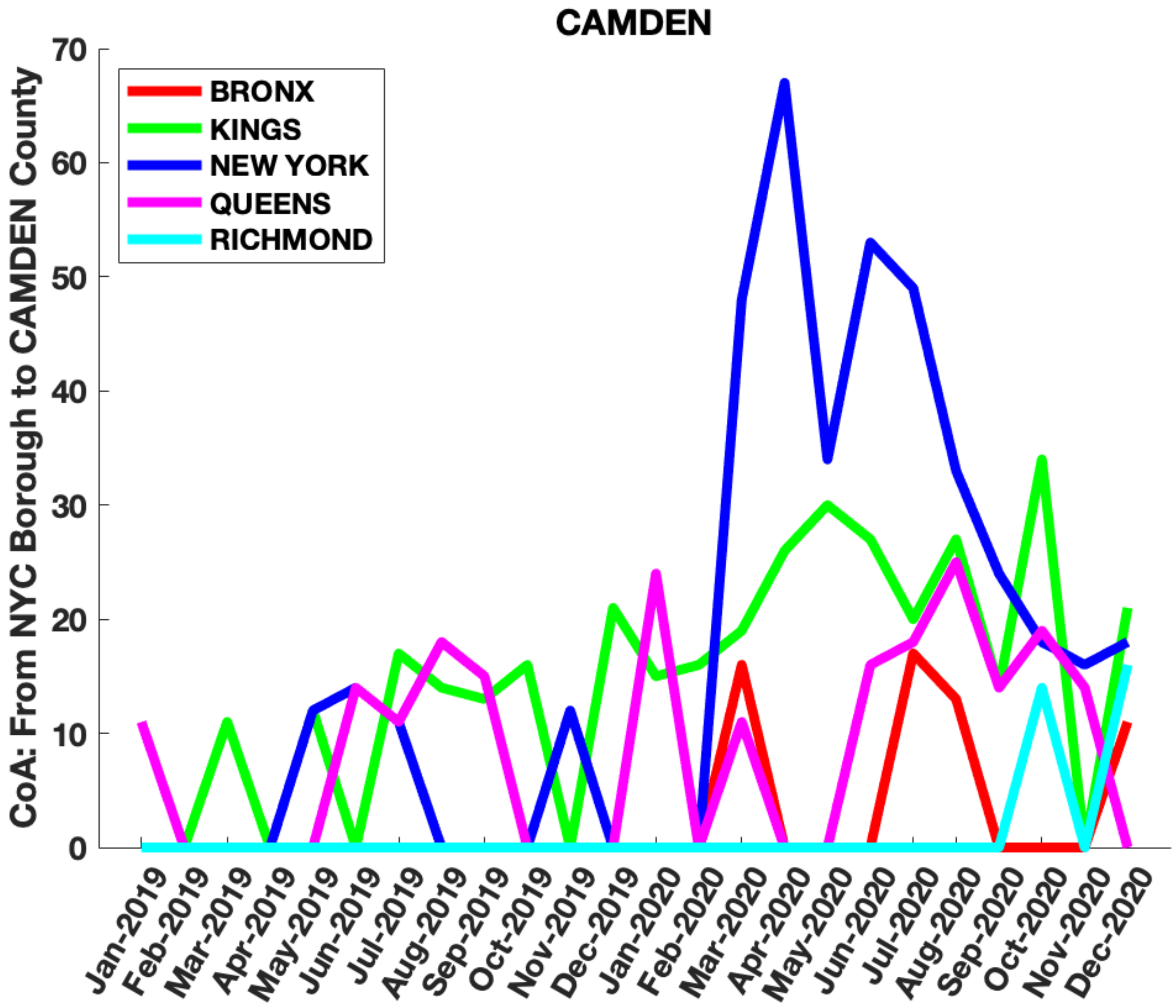

CAMDEN
CoA: From NYC Borough to CAMDEN County
BRONX
KINGS
NEW YORK
QUEENS
RICHMOND
0
10
20
30
40
50
60
70
Jan-2019
Feb-2019
Mar-2019
Apr-2019
May-2019
Jun-2019
Jul-2019
Aug-2019
Sep-2019
Oct-2019
Nov-2019
Dec-2019
Jan-2020
Feb-2020
Mar-2020
Apr-2020
May-2020
Jun-2020
Jul-2020
Aug-2020
Sep-2020
Oct-2020
Nov-2020
Dec-2020

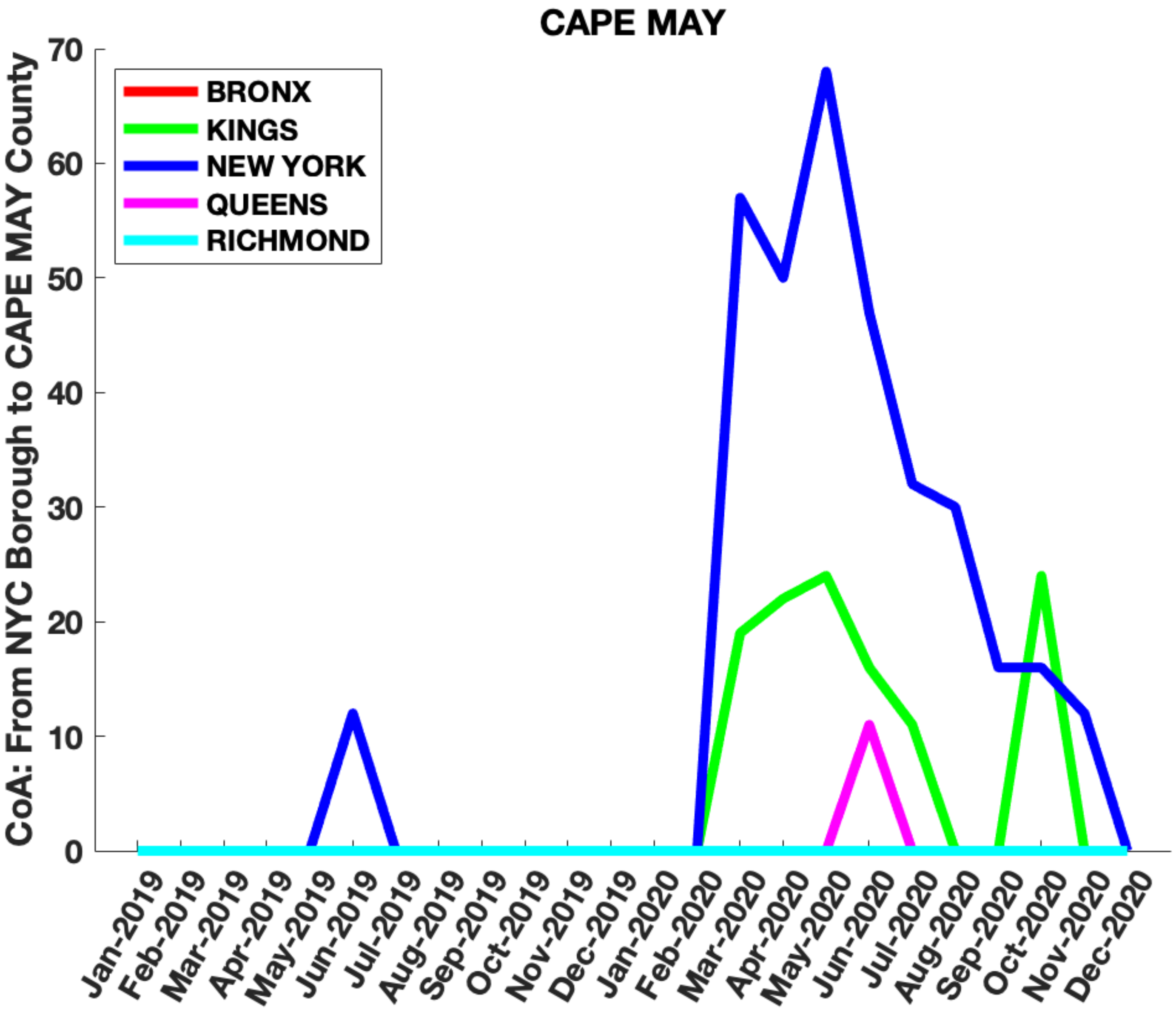
CAPE MAY
CoA: From NYC Borough to CAPE MAY County
BRONX
KINGS
NEW YORK
QUEENS
RICHMOND
0
10
20
30
40
50
60
70
Jan-2019
Feb-2019
Mar-2019
Apr-2019
May-2019
Jun-2019
Jul-2019
Aug-2019
Sep-2019
Oct-2019
Nov-2019
Dec-2019
Jan-2020
Feb-2020
Mar-2020
Apr-2020
May-2020
Jun-2020
Jul-2020
Aug-2020
Sep-2020
Oct-2020
Nov-2020
Dec-2020

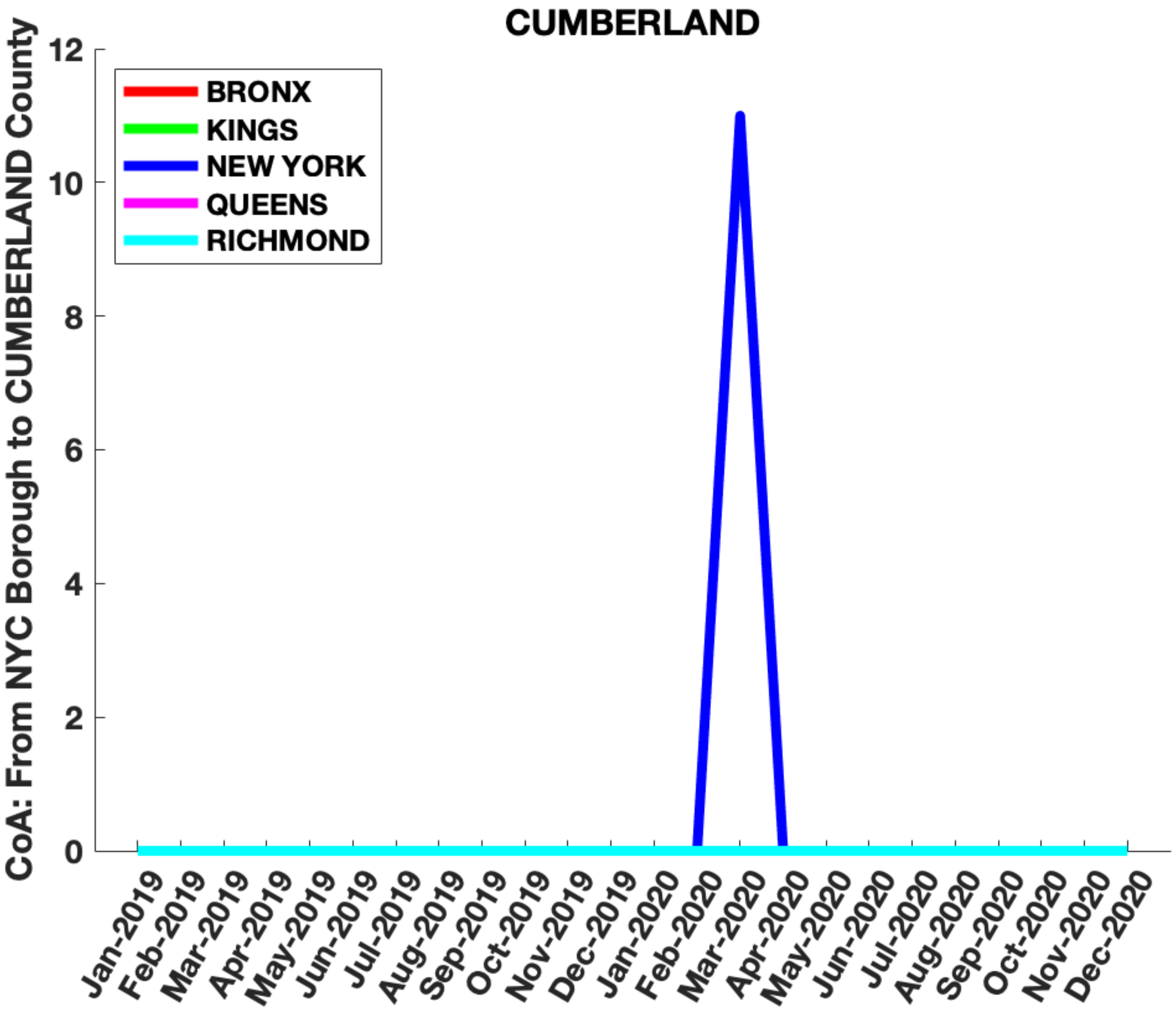

CUMBERLAND
CoA: From NYC Borough to CUMBERLAND County
BRONX
KINGS
NEW YORK
QUEENS
RICHMOND
0
2
4
6
8
10
12
Jan-2019
Feb-2019
Mar-2019
Apr-2019
May-2019
Jun-2019
Jul-2019
Aug-2019
Sep-2019
Oct-2019
Nov-2019
Dec-2019
Jan-2020
Feb-2020
Mar-2020
Apr-2020
May-2020
Jun-2020
Jul-2020
Aug-2020
Sep-2020
Oct-2020
Nov-2020
Dec-2020

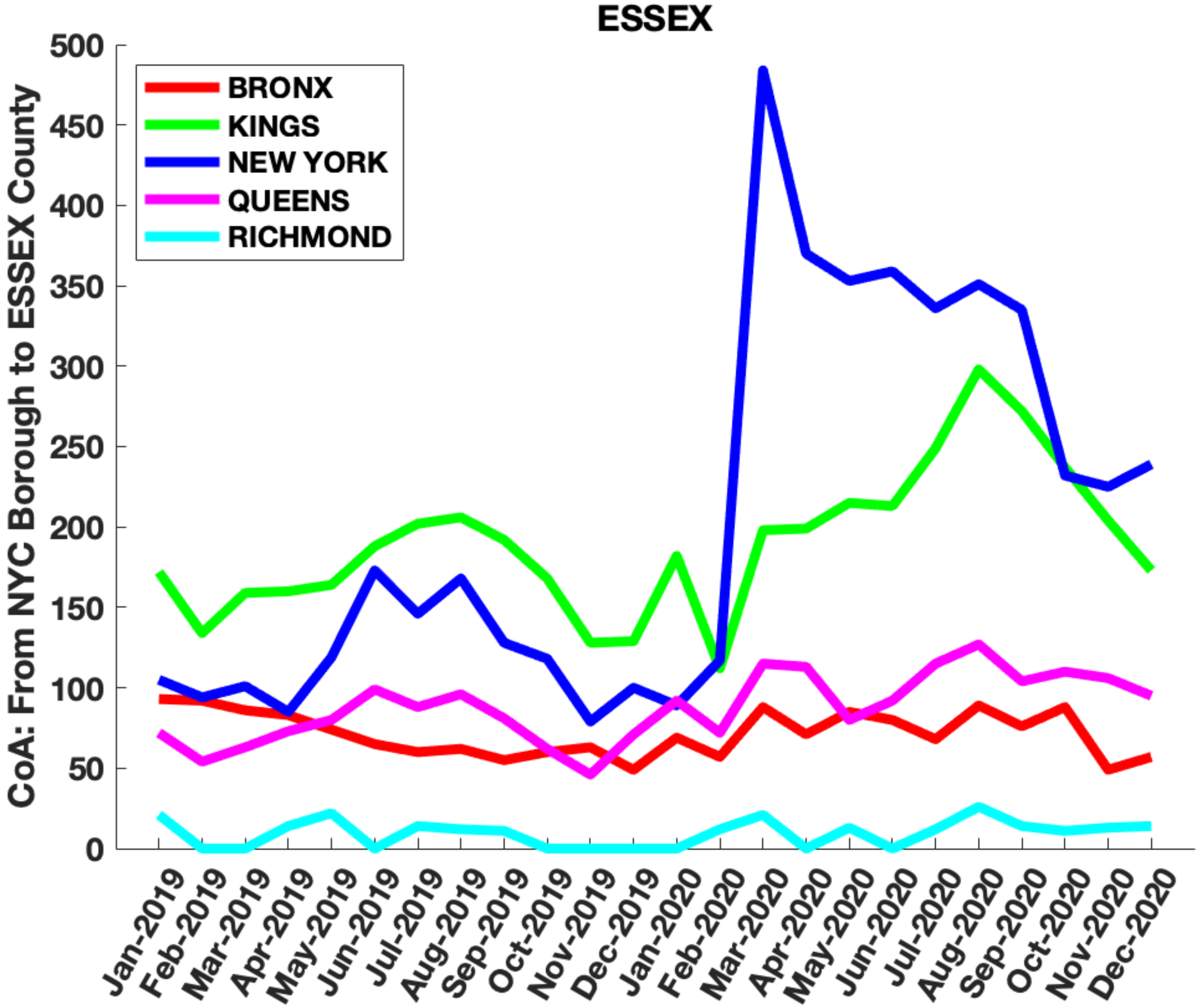

ESSEX
CoA: From NYC Borough to ESSEX County
BRONX
KINGS
NEW YORK
QUEENS
RICHMOND
0
50
100
150
200
250
300
350
400
450
500
Jan-2019
Feb-2019
Mar-2019
Apr-2019
May-2019
Jun-2019
Jul-2019
Aug-2019
Sep-2019
Oct-2019
Nov-2019
Dec-2019
Jan-2020
Feb-2020
Mar-2020
Apr-2020
May-2020
Jun-2020
Jul-2020
Aug-2020
Sep-2020
Oct-2020
Nov-2020
Dec-2020

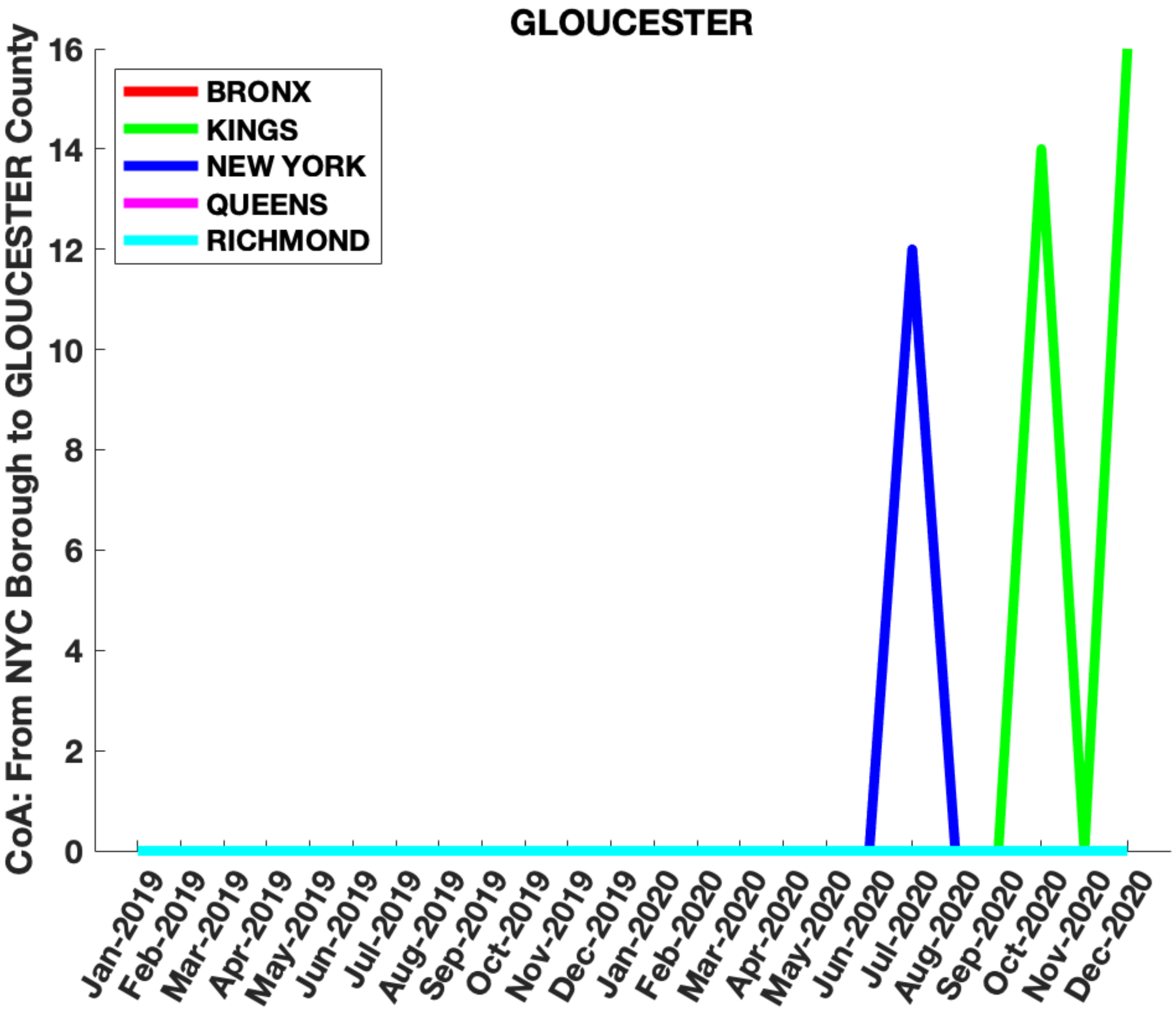
GLOUCESTER
CoA: From NYC Borough to GLOUCESTER County
BRONX
KINGS
NEW YORK
QUEENS
RICHMOND
0
2
4
6
8
10
12
14
16
Jan-2019
Feb-2019
Mar-2019
Apr-2019
May-2019
Jun-2019
Jul-2019
Aug-2019
Sep-2019
Oct-2019
Nov-2019
Dec-2019
Jan-2020
Feb-2020
Mar-2020
Apr-2020
May-2020
Jun-2020
Jul-2020
Aug-2020
Sep-2020
Oct-2020
Nov-2020
Dec-2020

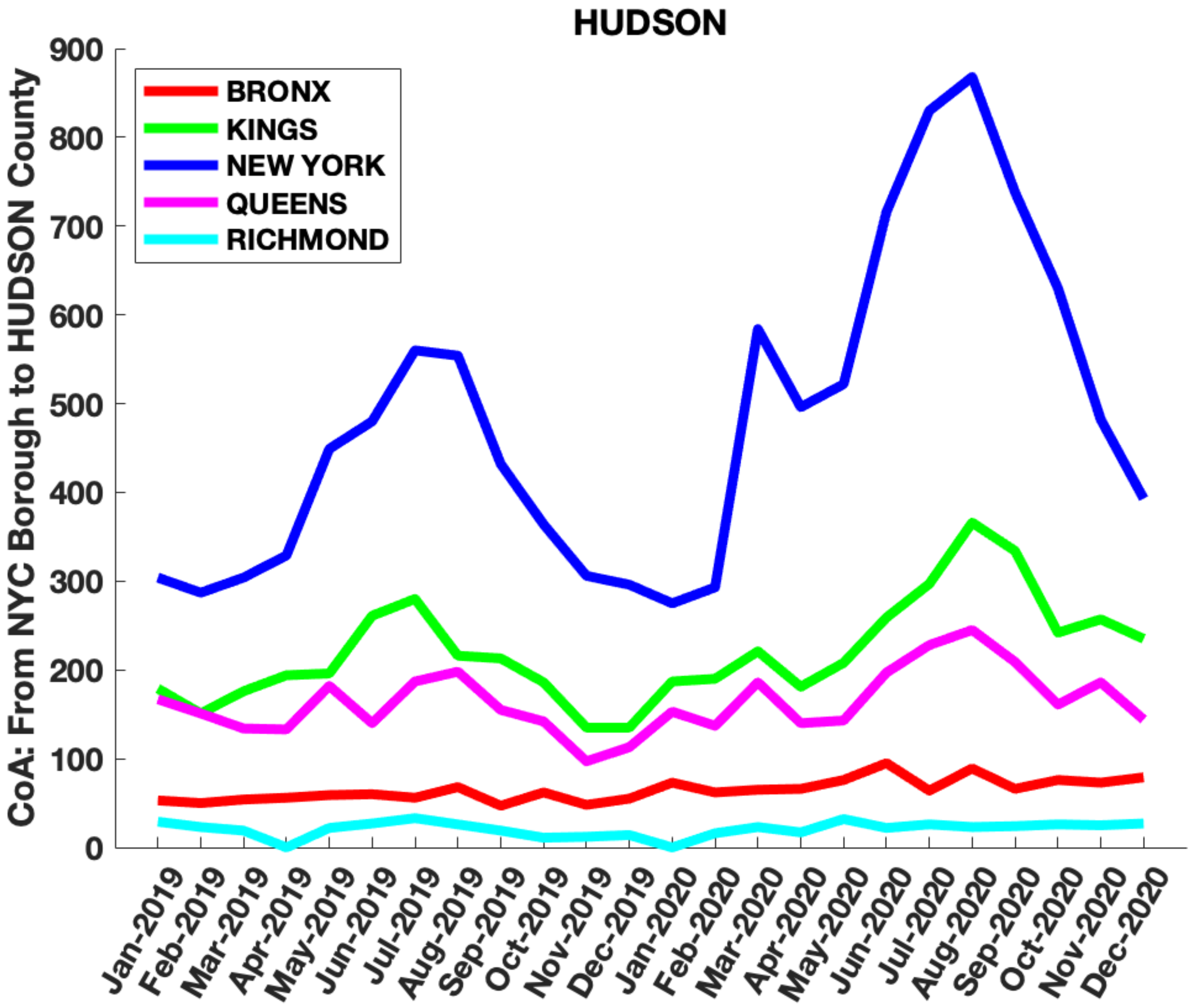
HUDSON
CoA: From NYC Borough to HUDSON County
BRONX
KINGS
NEW YORK
QUEENS
RICHMOND
0
100
200
300
400
500
600
700
800
900
Jan-2019
Feb-2019
Mar-2019
Apr-2019
May-2019
Jun-2019
Jul-2019
Aug-2019
Sep-2019
Oct-2019
Nov-2019
Dec-2019
Jan-2020
Feb-2020
Mar-2020
Apr-2020
May-2020
Jun-2020
Jul-2020
Aug-2020
Sep-2020
Oct-2020
Nov-2020
Dec-2020

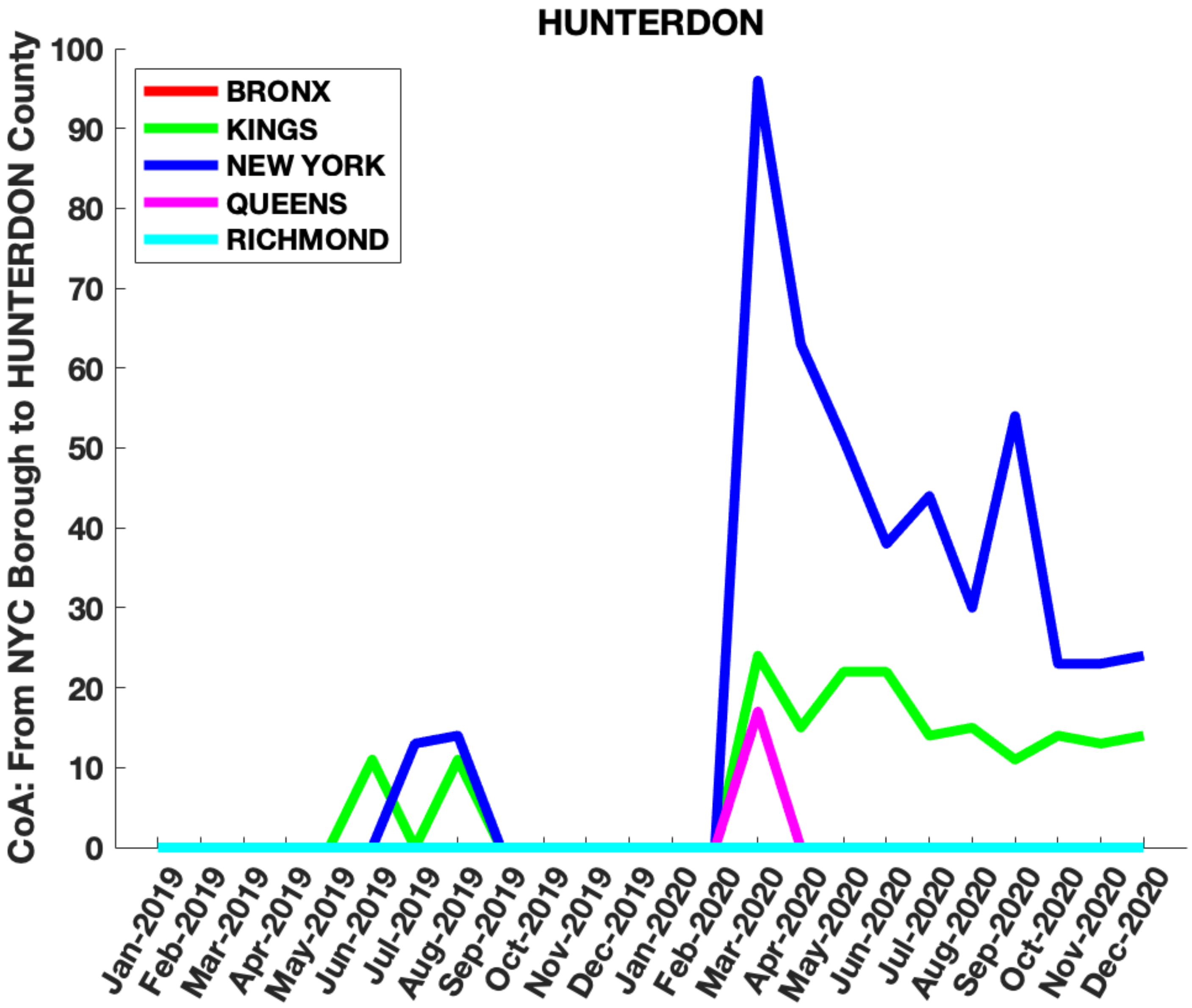
HUNTERDON
CoA: From NYC Borough to HUNTERDON County
BRONX
KINGS
NEW YORK
QUEENS
RICHMOND
0
10
20
30
40
50
60
70
80
90
100
Jan-2019
Feb-2019
Mar-2019
Apr-2019
May-2019
Jun-2019
Jul-2019
Aug-2019
Sep-2019
Oct-2019
Nov-2019
Dec-2019
Jan-2020
Feb-2020
Mar-2020
Apr-2020
May-2020
Jun-2020
Jul-2020
Aug-2020
Sep-2020
Oct-2020
Nov-2020
Dec-2020

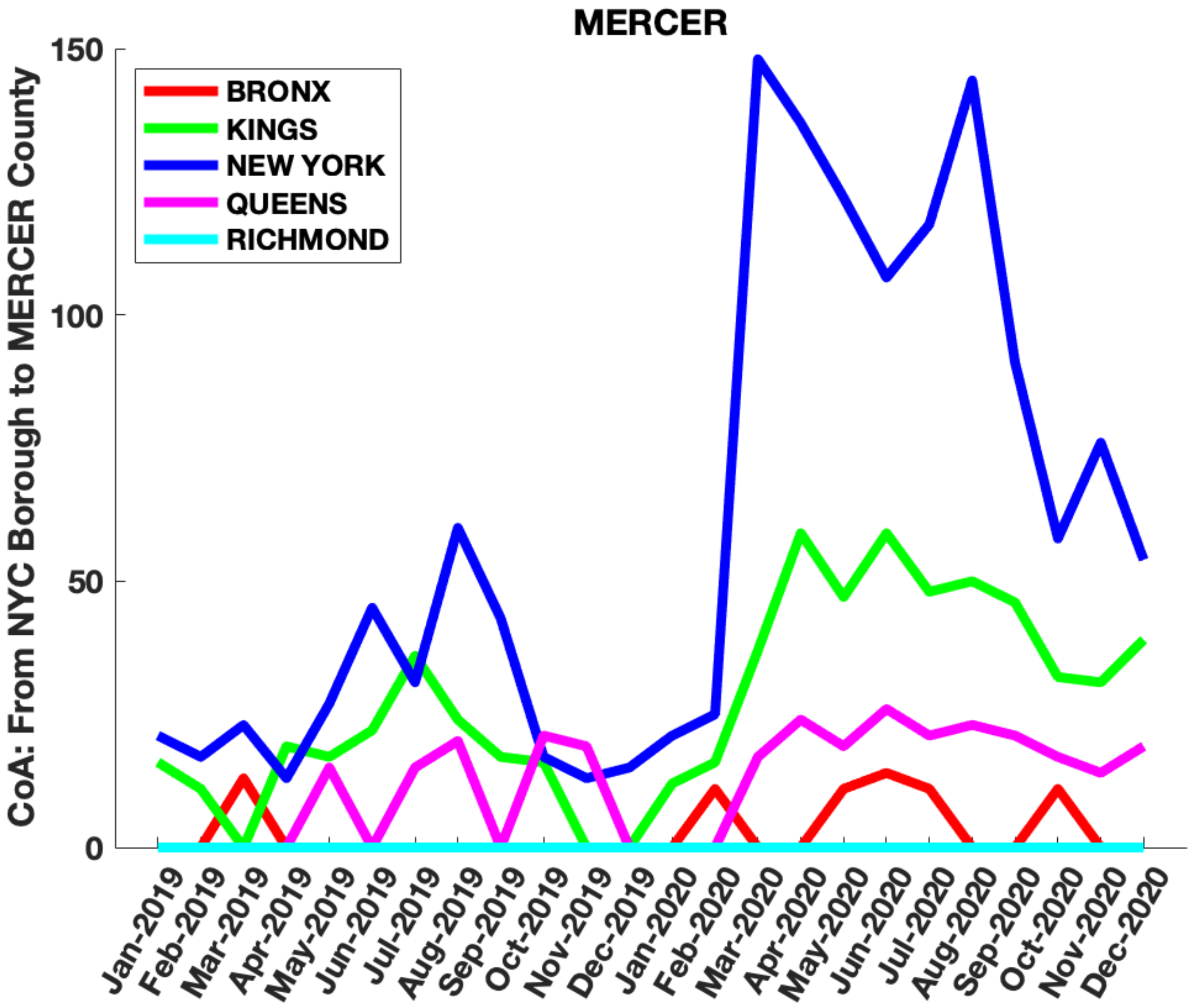

MERCER
CoA: From NYC Borough to MERCER County
BRONX
KINGS
NEW YORK
QUEENS
RICHMOND
150
100
50
0
Jan-2019
Feb-2019
Mar-2019
Apr-2019
May-2019
Jun-2019
Jul-2019
Aug-2019
Sep-2019
Oct-2019
Nov-2019
Dec-2019
Jan-2020
Feb-2020
Mar-2020
Apr-2020
May-2020
Jun-2020
Jul-2020
Aug-2020
Sep-2020
Oct-2020
Nov-2020
Dec-2020

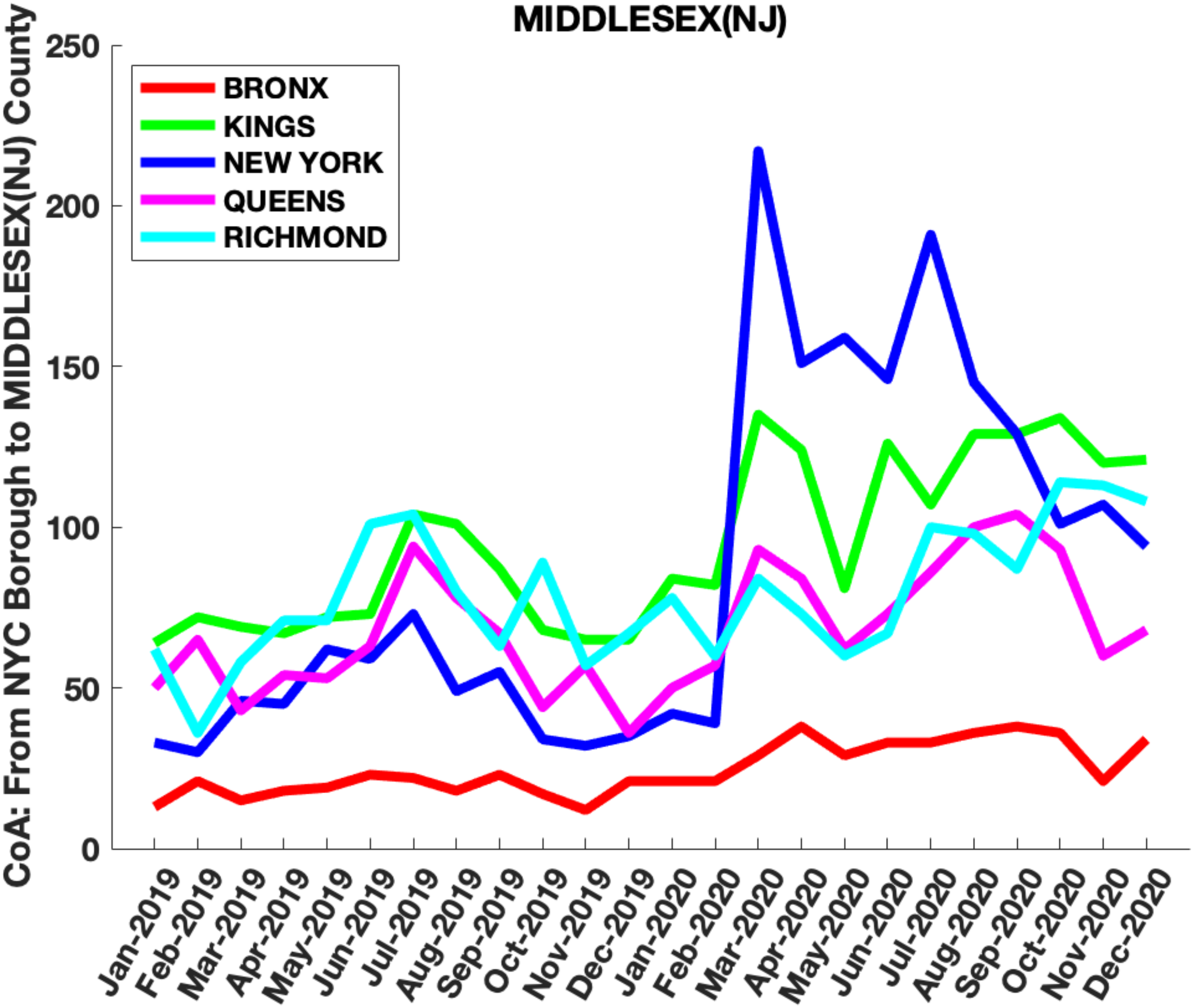

MIDDLESEX(NJ)
CoA: From NYC Borough to MIDDLESEX(NJ) County
BRONX
KINGS
NEW YORK
QUEENS
RICHMOND
0
50
100
150
200
250
Jan-2019
Feb-2019
Mar-2019
Apr-2019
May-2019
Jun-2019
Jul-2019
Aug-2019
Sep-2019
Oct-2019
Nov-2019
Dec-2019
Jan-2020
Feb-2020
Mar-2020
Apr-2020
May-2020
Jun-2020
Jul-2020
Aug-2020
Sep-2020
Oct-2020
Nov-2020
Dec-2020

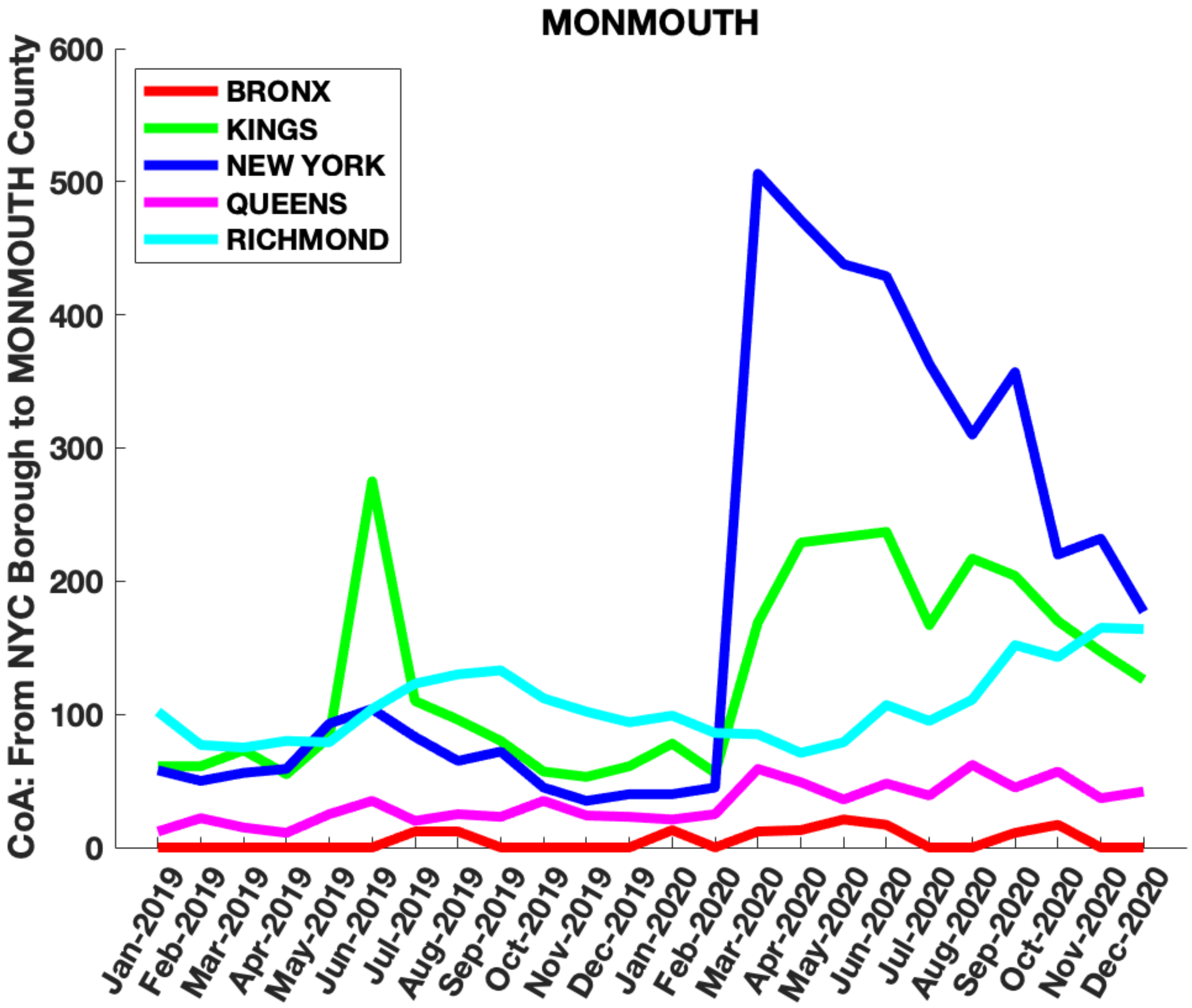

MONMOUTH
CoA: From NYC Borough to MONMOUTH County
BRONX
KINGS
NEW YORK
QUEENS
RICHMOND
0
100
200
300
400
500
600
Jan-2019
Feb-2019
Mar-2019
Apr-2019
May-2019
Jun-2019
Jul-2019
Aug-2019
Sep-2019
Oct-2019
Nov-2019
Dec-2019
Jan-2020
Feb-2020
Mar-2020
Apr-2020
May-2020
Jun-2020
Jul-2020
Aug-2020
Sep-2020
Oct-2020
Nov-2020
Dec-2020

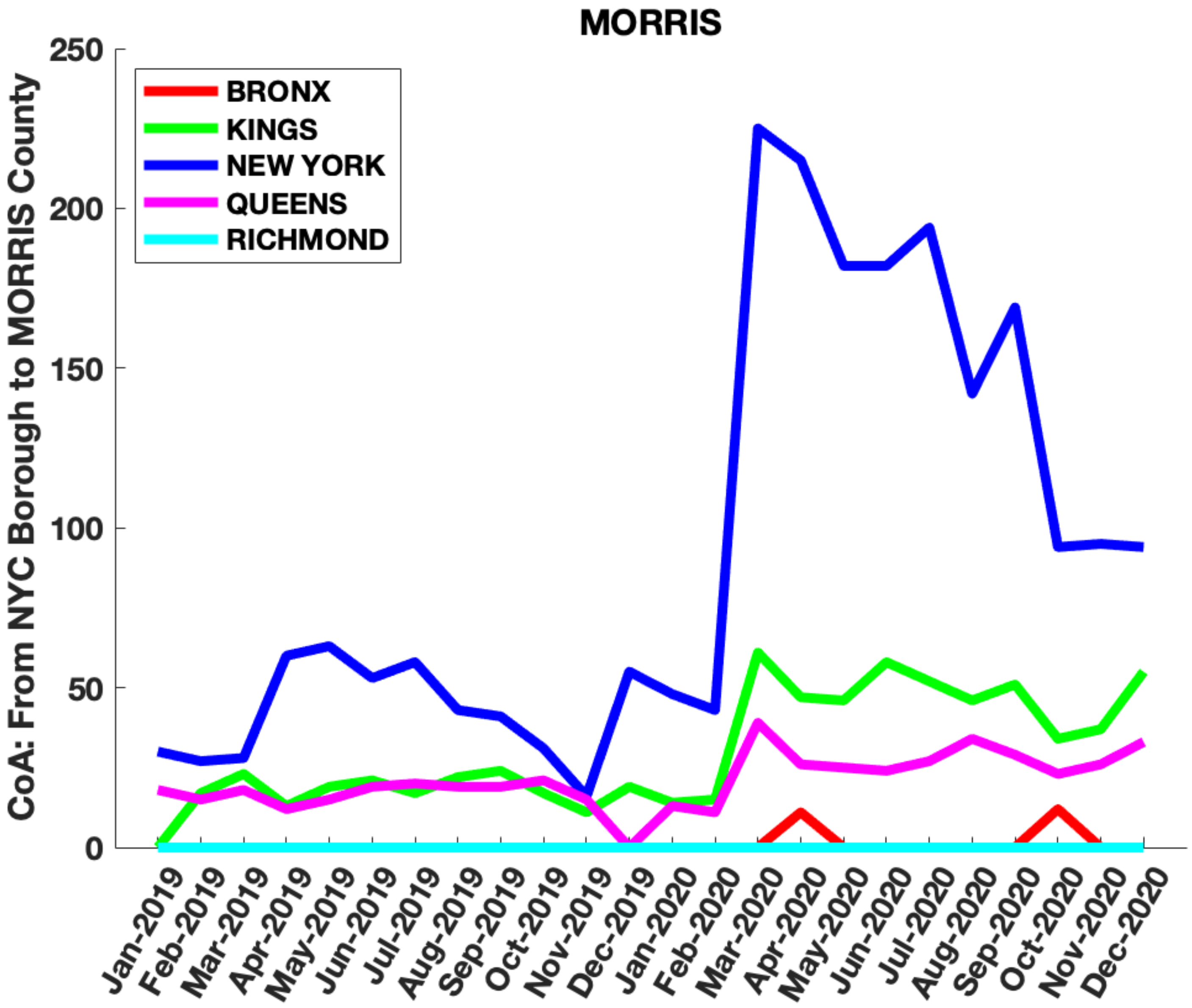
MORRIS
CoA: From NYC Borough to MORRIS County
BRONX
KINGS
NEW YORK
QUEENS
RICHMOND
250
200
150
100
50
0
Jan-2019
Feb-2019
Mar-2019
Apr-2019
May-2019
Jun-2019
Jul-2019
Aug-2019
Sep-2019
Oct-2019
Nov-2019
Dec-2019
Jan-2020
Feb-2020
Mar-2020
Apr-2020
May-2020
Jun-2020
Jul-2020
Aug-2020
Sep-2020
Oct-2020
Nov-2020
Dec-2020

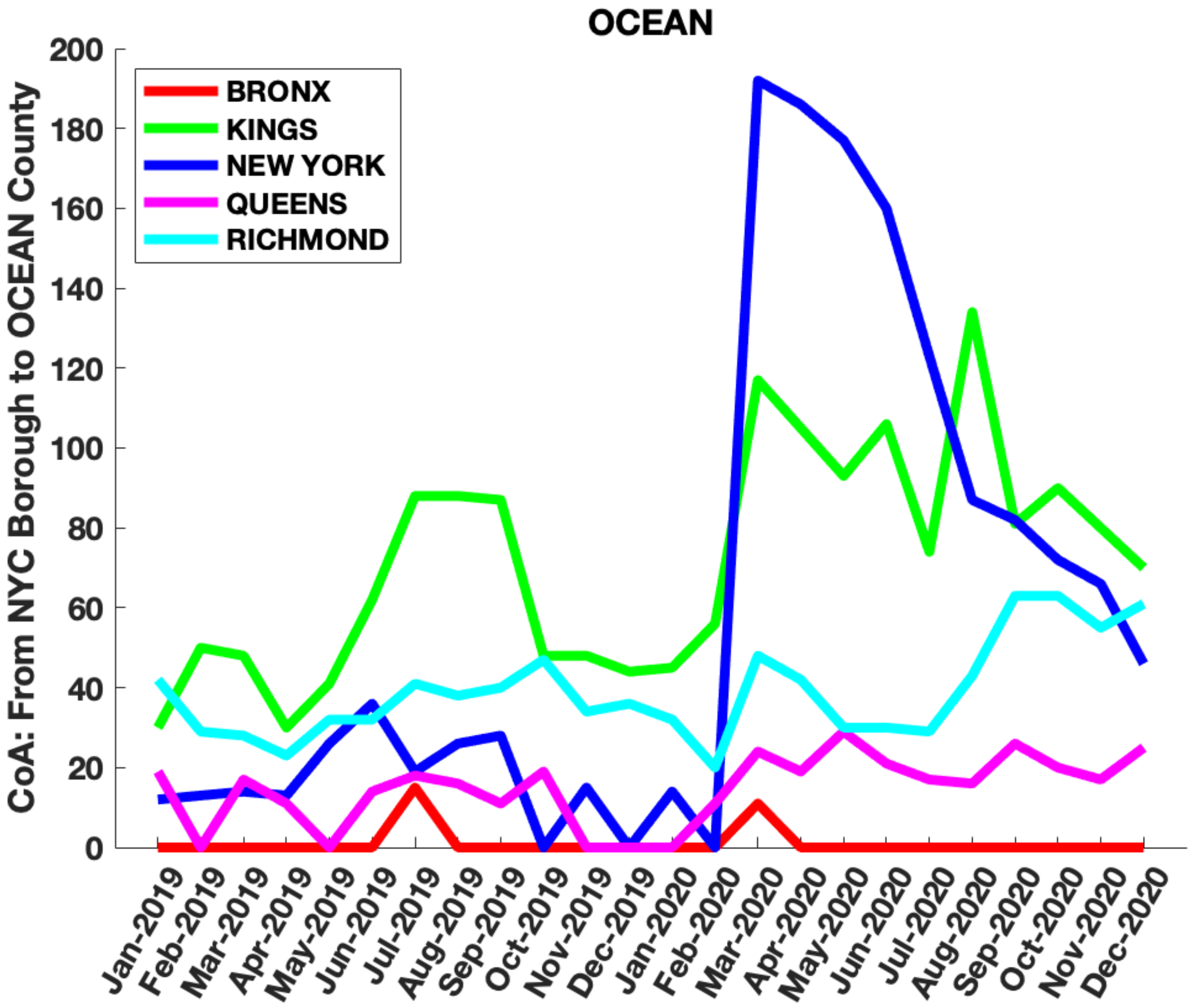
OCEAN
BRONX
KINGS
NEW YORK
QUEENS
RICHMOND
CoA: From NYC Borough to OCEAN County
0
20
40
60
80
100
120
140
160
180
200
Jan-2019
Feb-2019
Mar-2019
Apr-2019
May-2019
Jun-2019
Jul-2019
Aug-2019
Sep-2019
Oct-2019
Nov-2019
Dec-2019
Jan-2020
Feb-2020
Mar-2020
Apr-2020
May-2020
Jun-2020
Jul-2020
Aug-2020
Sep-2020
Oct-2020
Nov-2020
Dec-2020

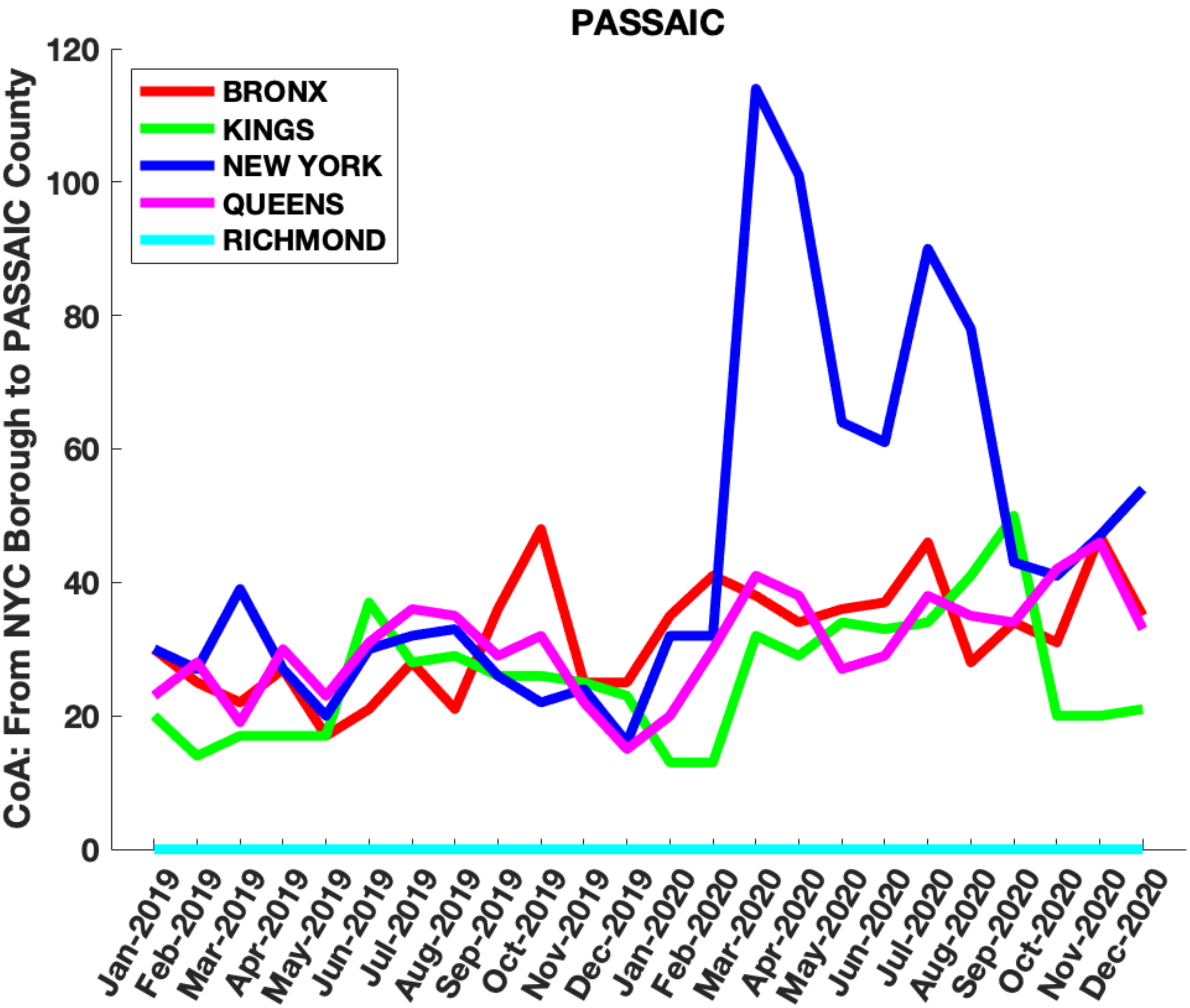

PASSAIC
CoA: From NYC Borough to PASSAIC County
BRONX
KINGS
NEW YORK
QUEENS
RICHMOND
0
20
40
60
80
100
120
Jan-2019
Feb-2019
Mar-2019
Apr-2019
May-2019
Jun-2019
Jul-2019
Aug-2019
Sep-2019
Oct-2019
Nov-2019
Dec-2019
Jan-2020
Feb-2020
Mar-2020
Apr-2020
May-2020
Jun-2020
Jul-2020
Aug-2020
Sep-2020
Oct-2020
Nov-2020
Dec-2020

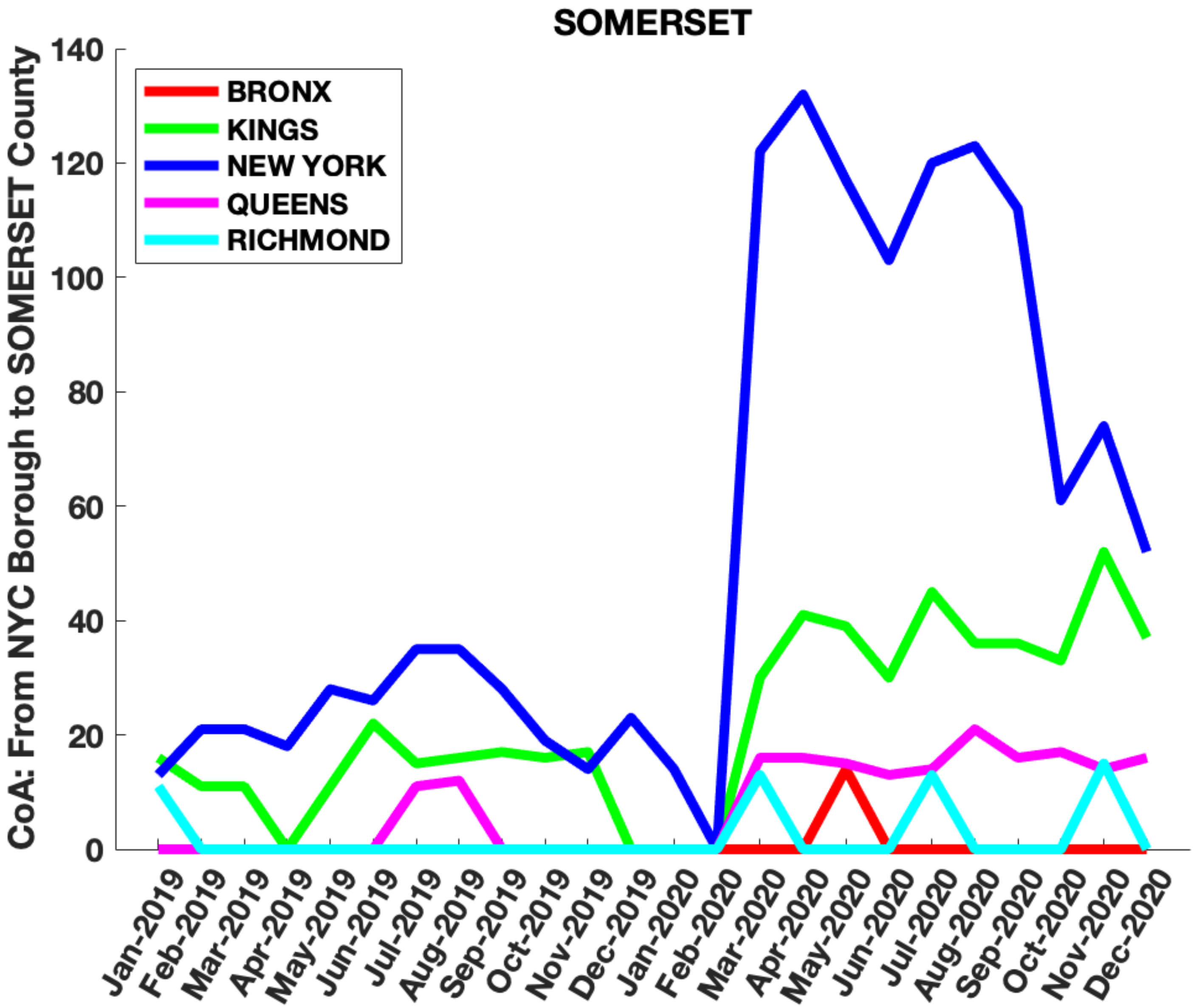

SOMERSET
BRONX
KINGS
NEW YORK
QUEENS
RICHMOND
CoA: From NYC Borough to SOMERSET County
0
20
40
60
80
100
120
140
Jan-2019
Feb-2019
Mar-2019
Apr-2019
May-2019
Jun-2019
Jul-2019
Aug-2019
Sep-2019
Oct-2019
Nov-2019
Dec-2019
Jan-2020
Feb-2020
Mar-2020
Apr-2020
May-2020
Jun-2020
Jul-2020
Aug-2020
Sep-2020
Oct-2020
Nov-2020
Dec-2020

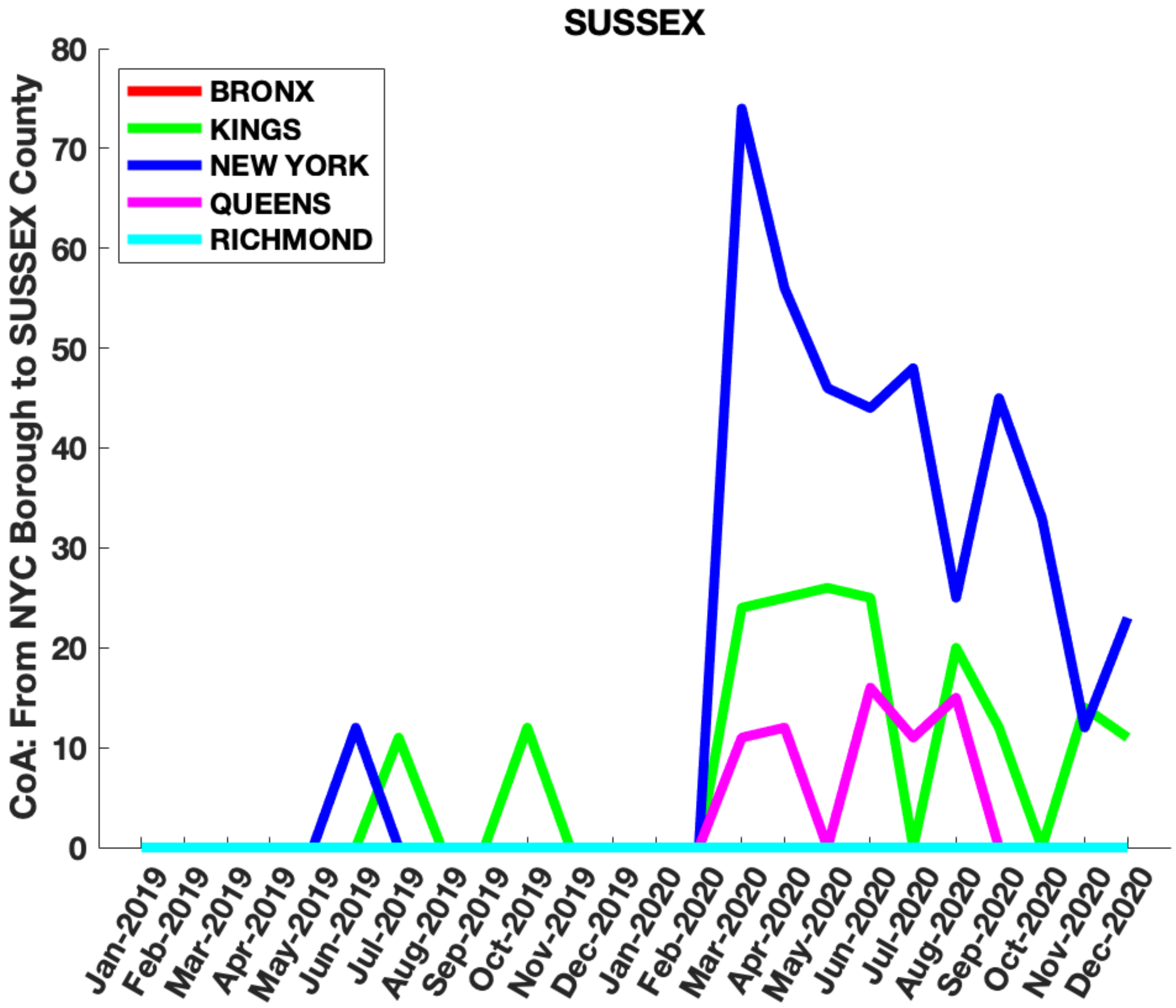

SUSSEX
CoA: From NYC Borough to SUSSEX County
BRONX
KINGS
NEW YORK
QUEENS
RICHMOND
0
10
20
30
40
50
60
70
80
Jan-2019
Feb-2019
Mar-2019
Apr-2019
May-2019
Jun-2019
Jul-2019
Aug-2019
Sep-2019
Oct-2019
Nov-2019
Dec-2019
Jan-2020
Feb-2020
Mar-2020
Apr-2020
May-2020
Jun-2020
Jul-2020
Aug-2020
Sep-2020
Oct-2020
Nov-2020
Dec-2020

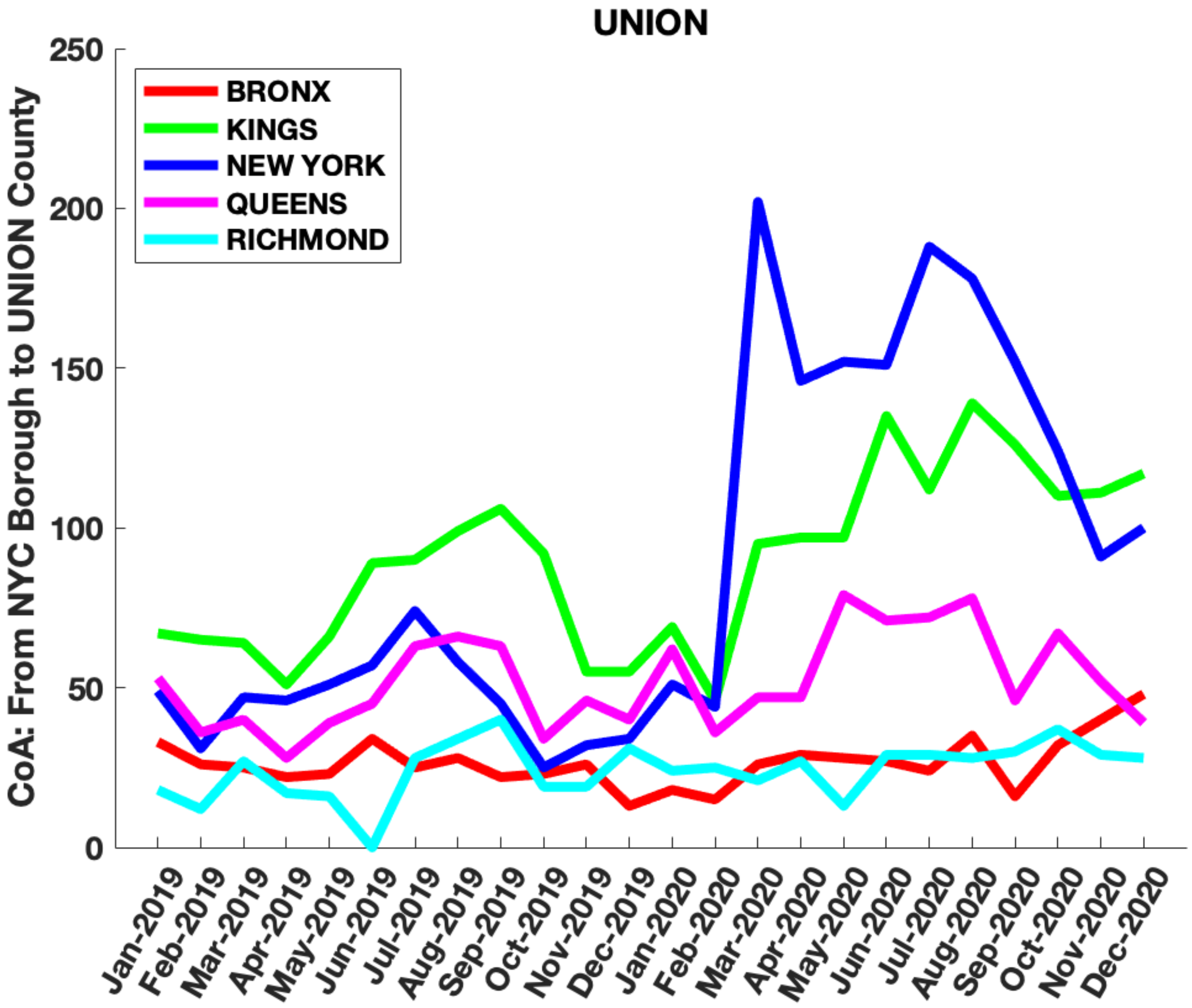

UNION
CoA: From NYC Borough to UNION County
BRONX
KINGS
NEW YORK
QUEENS
RICHMOND
0
50
100
150
200
250
Jan-2019
Feb-2019
Mar-2019
Apr-2019
May-2019
Jun-2019
Jul-2019
Aug-2019
Sep-2019
Oct-2019
Nov-2019
Dec-2019
Jan-2020
Feb-2020
Mar-2020
Apr-2020
May-2020
Jun-2020
Jul-2020
Aug-2020
Sep-2020
Oct-2020
Nov-2020
Dec-2020

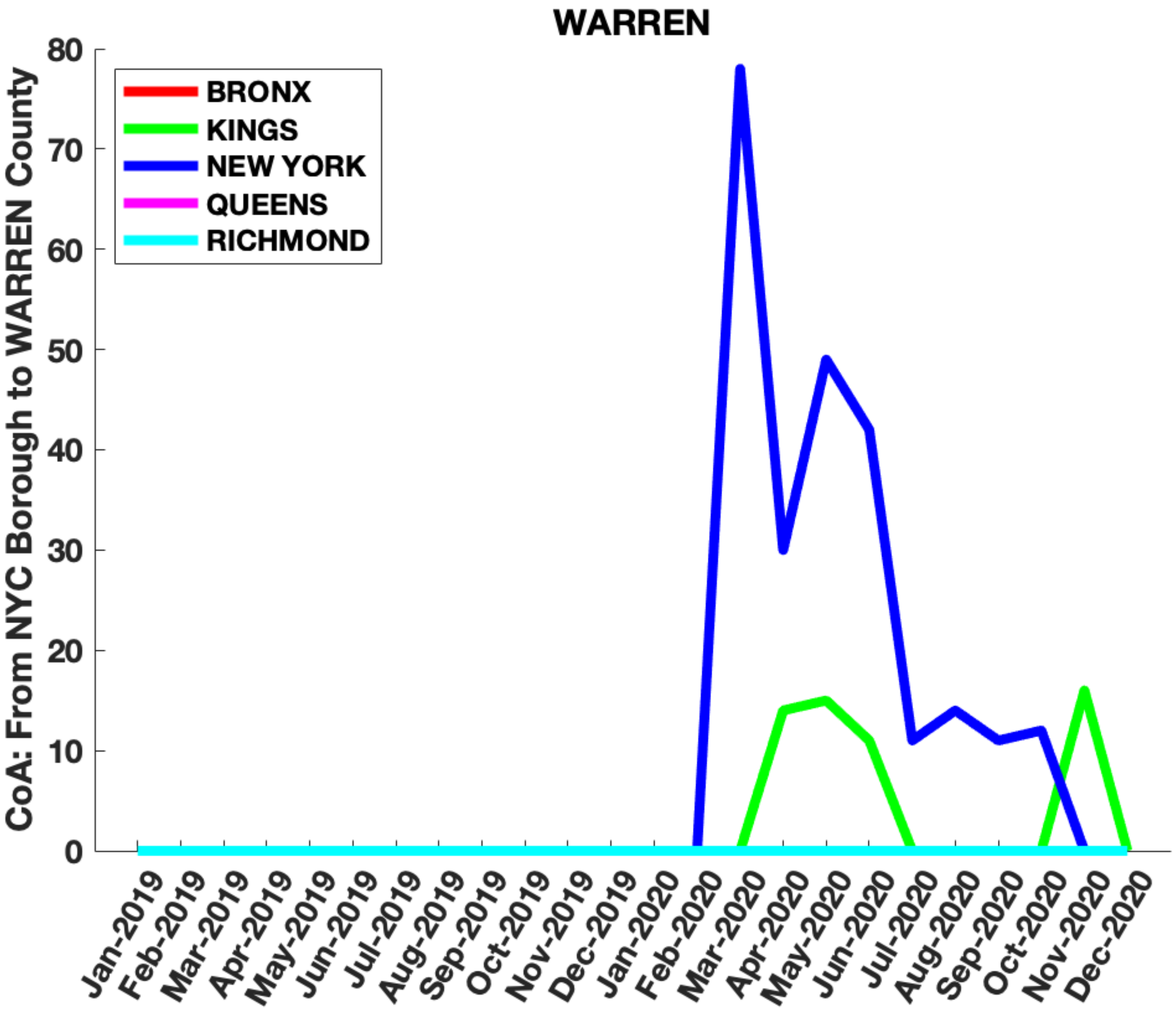

WARREN
CoA: From NYC Borough to WARREN County
BRONX
KINGS
NEW YORK
QUEENS
RICHMOND
0
10
20
30
40
50
60
70
80
Jan-2019
Feb-2019
Mar-2019
Apr-2019
May-2019
Jun-2019
Jul-2019
Aug-2019
Sep-2019
Oct-2019
Nov-2019
Dec-2019
Jan-2020
Feb-2020
Mar-2020
Apr-2020
May-2020
Jun-2020
Jul-2020
Aug-2020
Sep-2020
Oct-2020
Nov-2020
Dec-2020
