## Supplementary Figure 6 for "Migration of households from New York City and the Second Peak in Covid-19 cases in New Jersey, Connecticut and New York Counties"

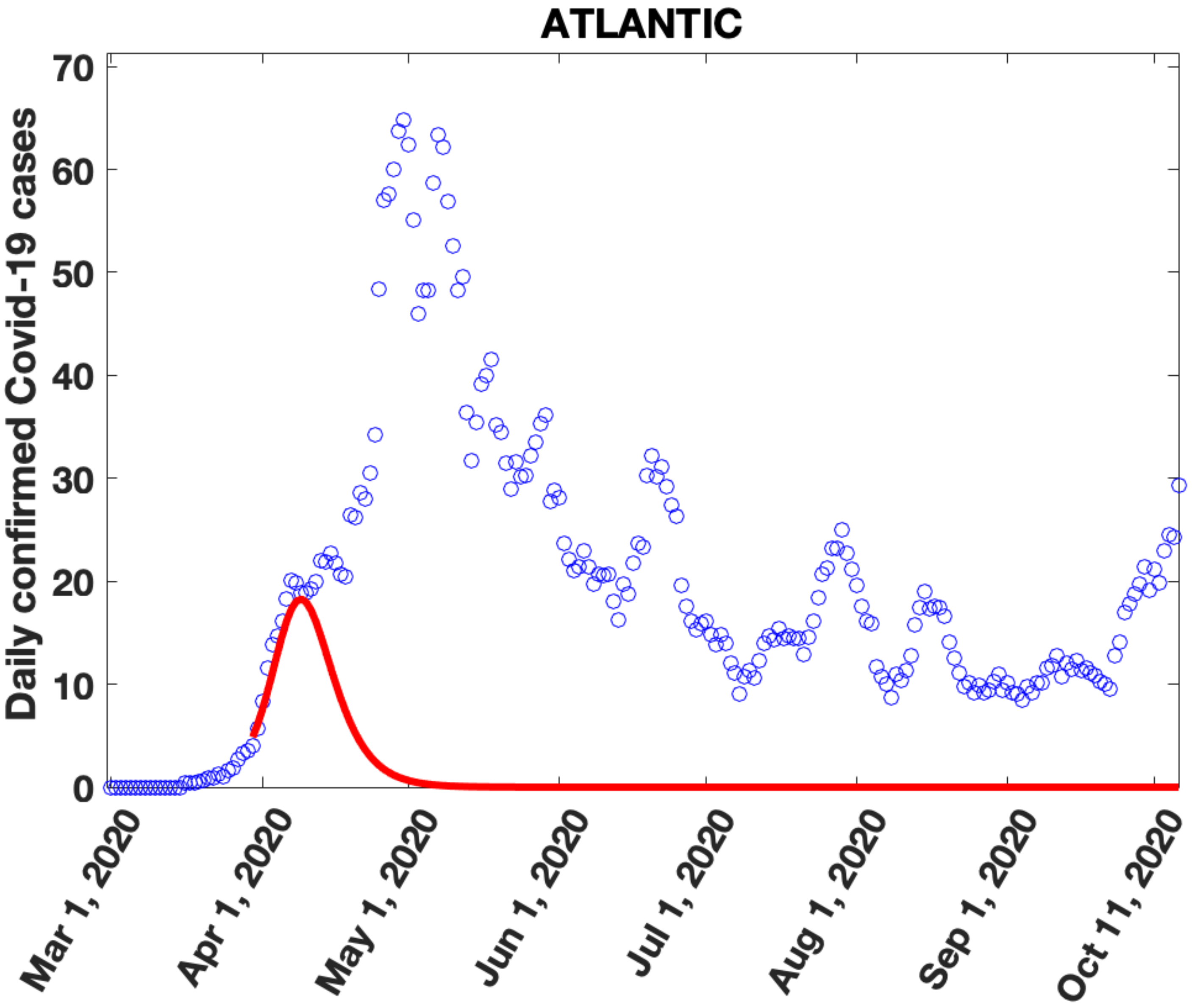
ATLANTIC
Daily confirmed Covid-19 cases
0
10
20
30
40
50
60
70
Mar 1, 2020
Apr 1, 2020
May 1, 2020
Jun 1, 2020
Jul 1, 2020
Aug 1, 2020
Sep 1, 2020
Oct 11, 2020

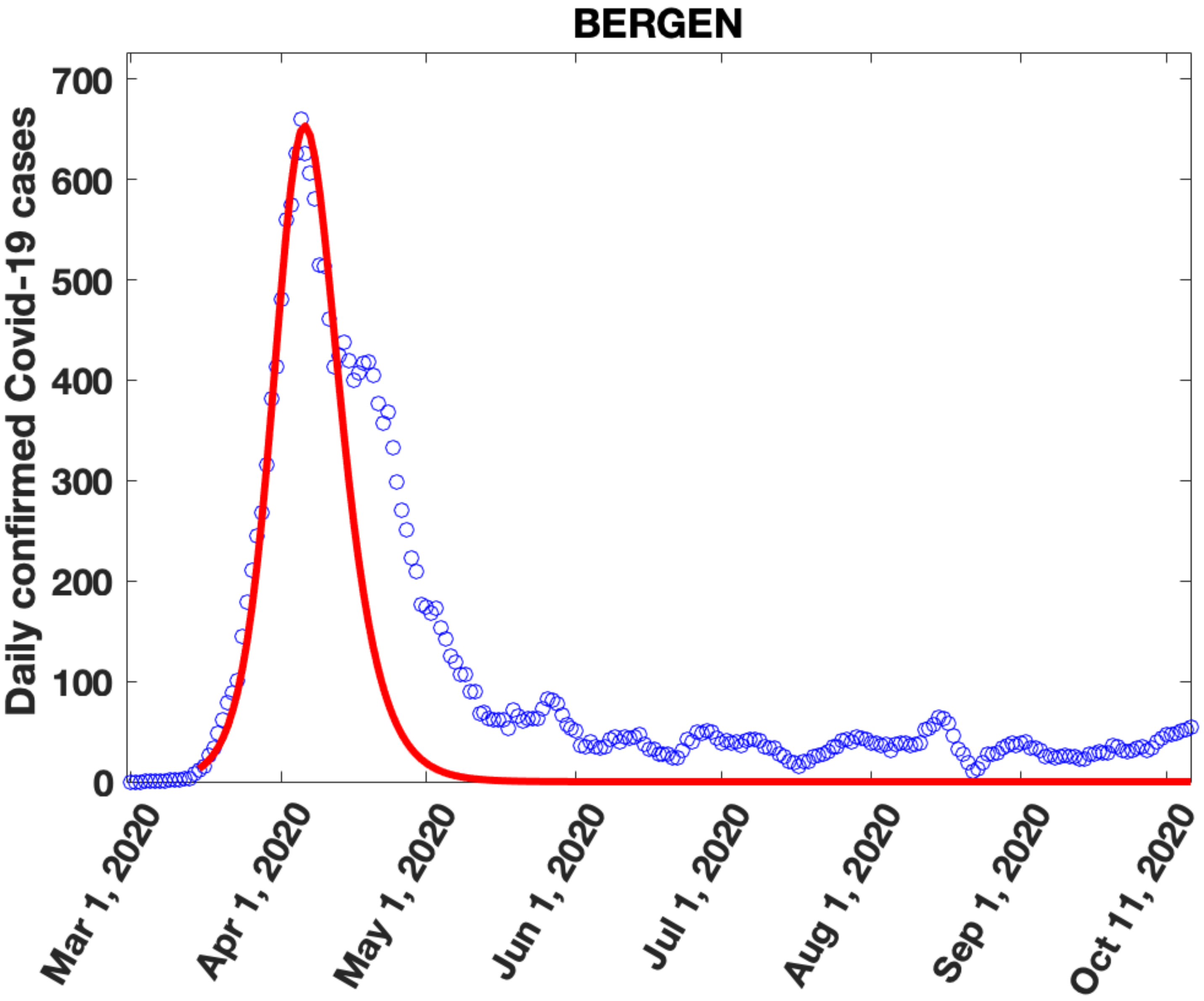
BERGEN
Daily confirmed Covid-19 cases
0
100
200
300
400
500
600
700
Mar 1, 2020
Apr 1, 2020
May 1, 2020
Jun 1, 2020
Jul 1, 2020
Aug 1, 2020
Sep 1, 2020
Oct 11, 2020

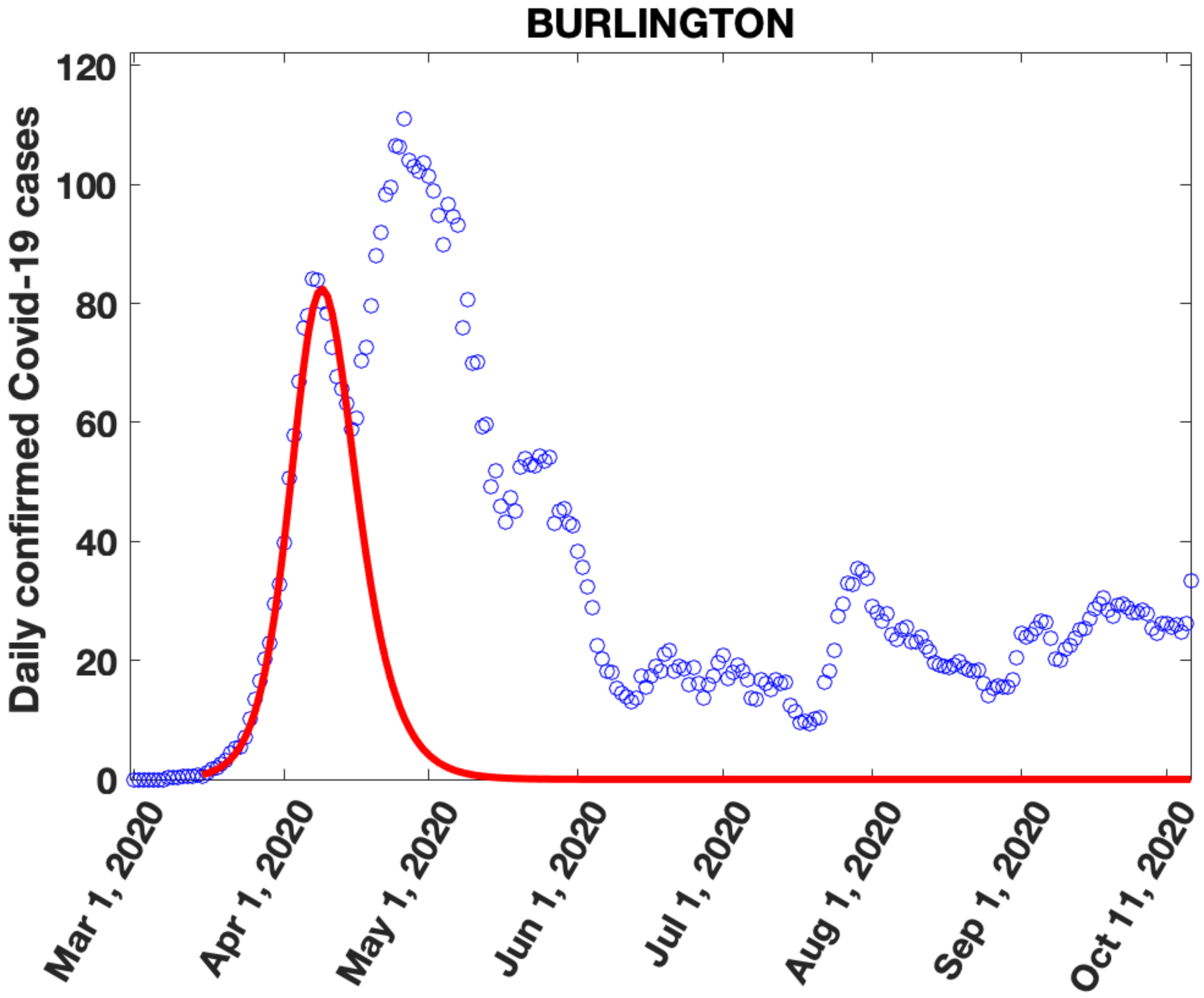

BURLINGTON
Daily confirmed Covid-19 cases
120
100
80
60
40
20
0
Mar 1, 2020
Apr 1, 2020
May 1, 2020
Jun 1, 2020
Jul 1, 2020
Aug 1, 2020
Sep 1, 2020
Oct 11, 2020

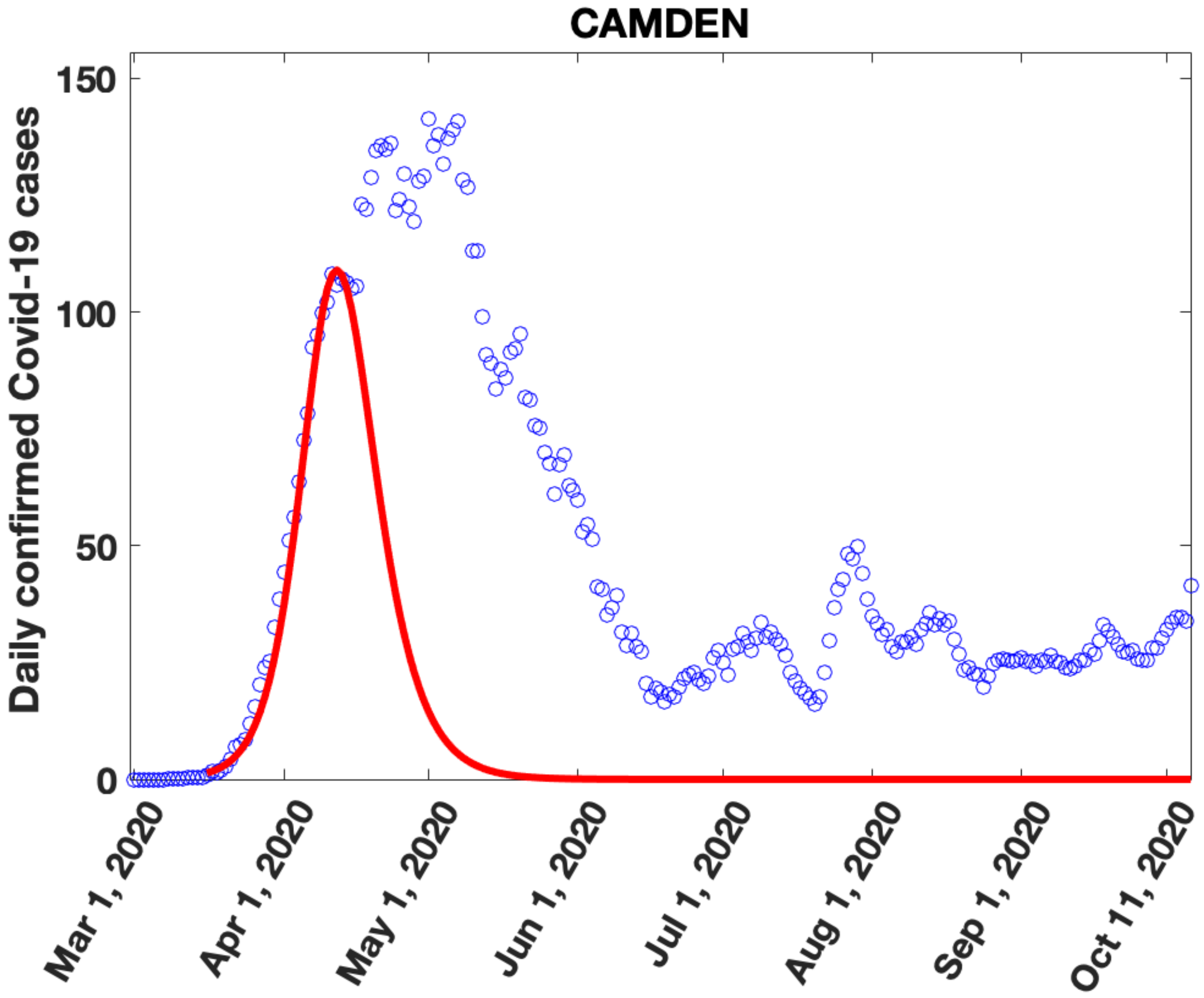

CAMDEN
Daily confirmed Covid-19 cases
150
100
50
0
Mar 1, 2020
Apr 1, 2020
May 1, 2020
Jun 1, 2020
Jul 1, 2020
Aug 1, 2020
Sep 1, 2020
Oct 11, 2020

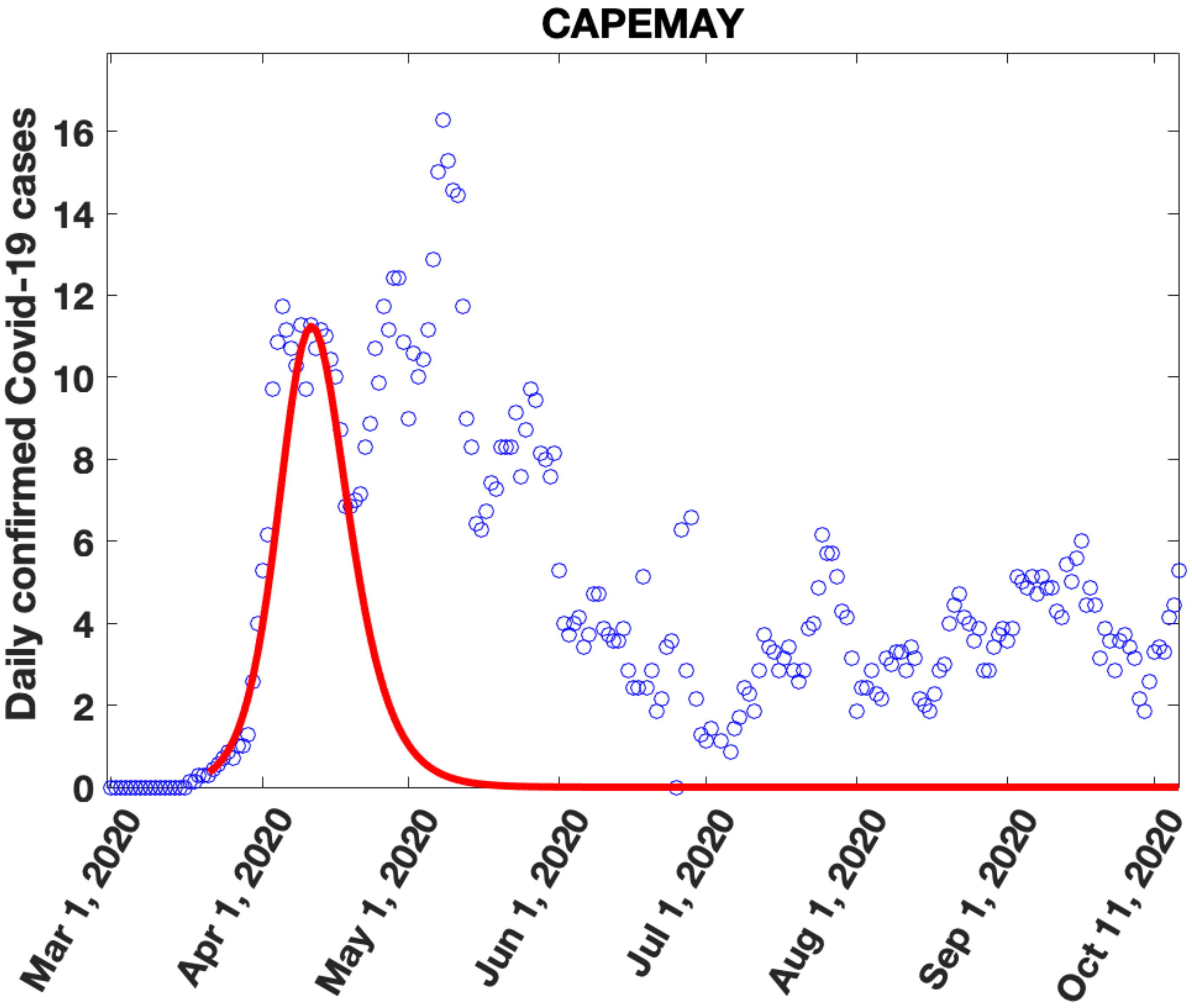

CAPEMAY
Daily confirmed Covid-19 cases
0
2
4
6
8
10
12
14
16
Mar 1, 2020
Apr 1, 2020
May 1, 2020
Jun 1, 2020
Jul 1, 2020
Aug 1, 2020
Sep 1, 2020
Oct 11, 2020

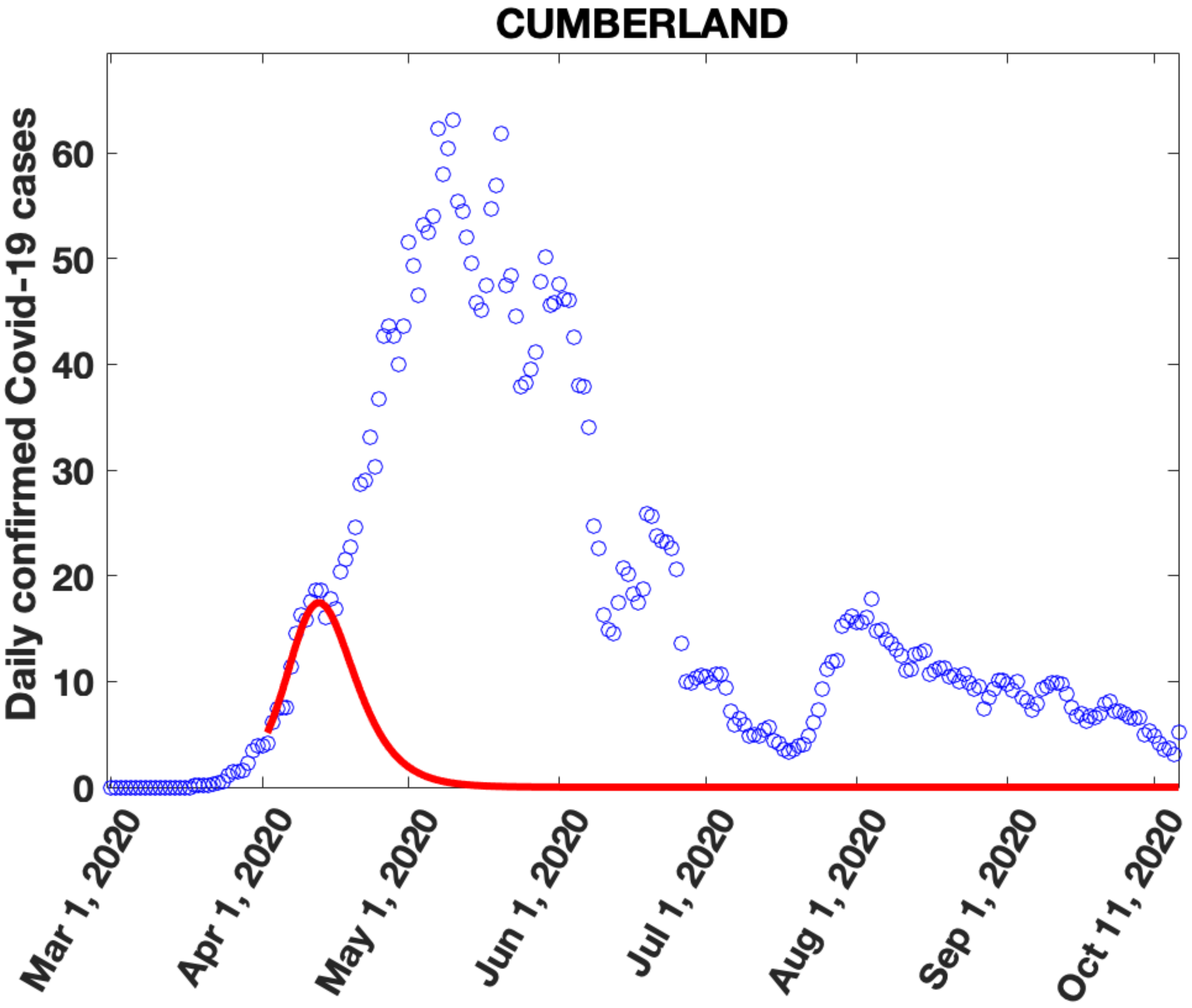

CUMBERLAND
Daily confirmed Covid-19 cases
0
10
20
30
40
50
60
Mar 1, 2020
Apr 1, 2020
May 1, 2020
Jun 1, 2020
Jul 1, 2020
Aug 1, 2020
Sep 1, 2020
Oct 11, 2020

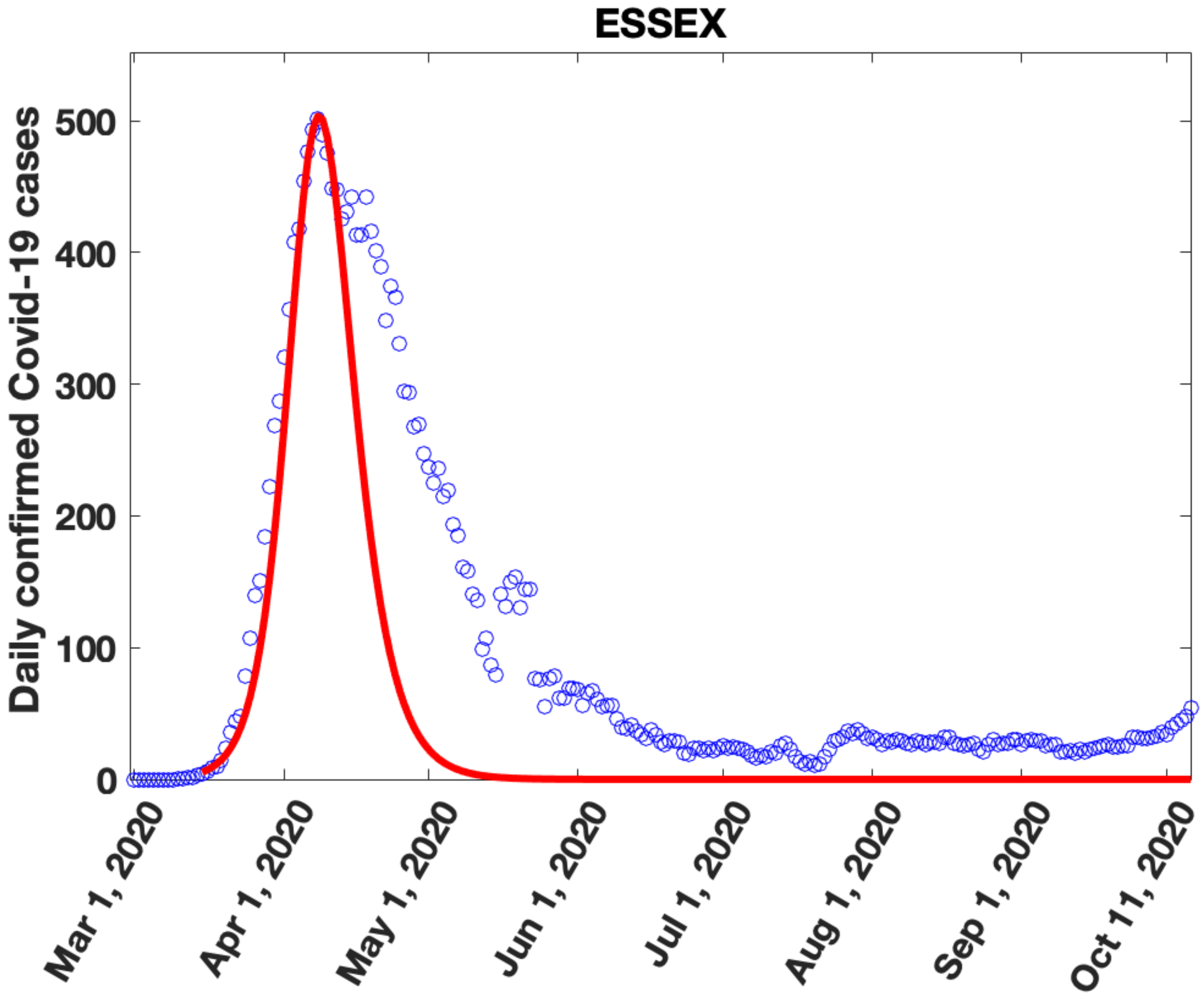
ESSEX
Daily confirmed Covid-19 cases
0
100
200
300
400
500
Mar 1, 2020
Apr 1, 2020
May 1, 2020
Jun 1, 2020
Jul 1, 2020
Aug 1, 2020
Sep 1, 2020
Oct 11, 2020

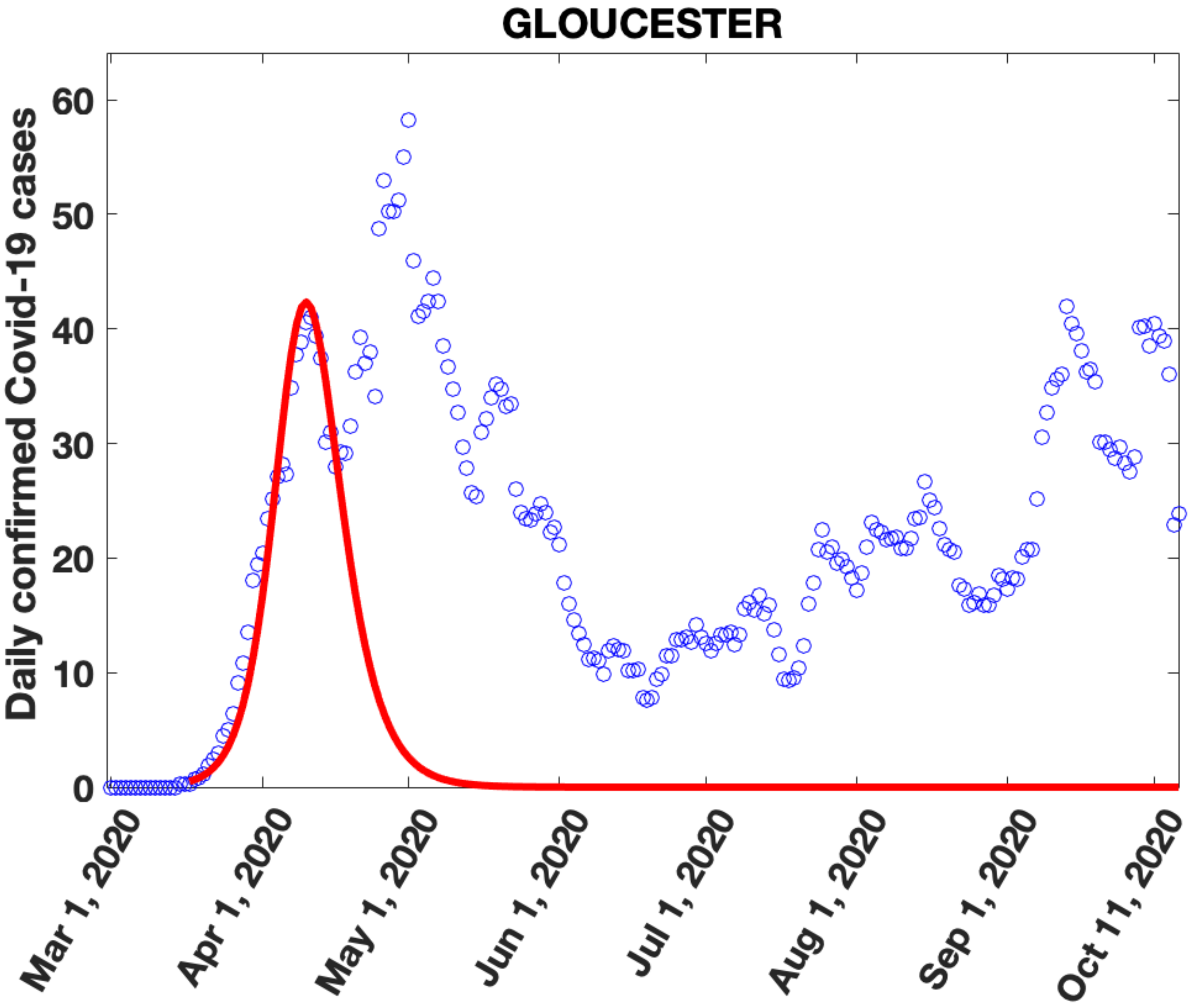
GLOUCESTER
Daily confirmed Covid-19 cases
0
10
20
30
40
50
60
Mar 1, 2020
Apr 1, 2020
May 1, 2020
Jun 1, 2020
Jul 1, 2020
Aug 1, 2020
Sep 1, 2020
Oct 11, 2020

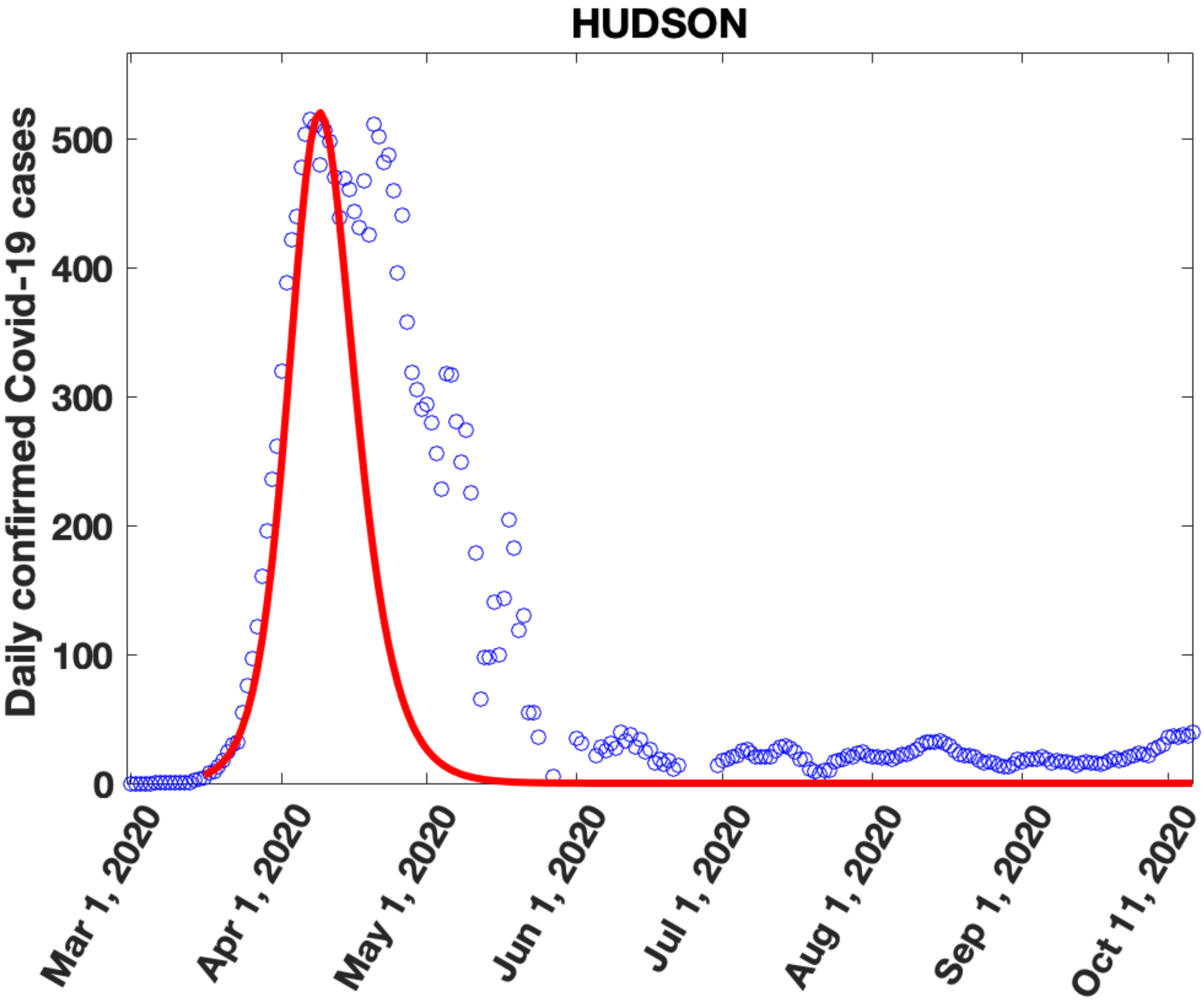

HUDSON
Daily confirmed Covid-19 cases
0
100
200
300
400
500
Mar 1, 2020
Apr 1, 2020
May 1, 2020
Jun 1, 2020
Jul 1, 2020
Aug 1, 2020
Sep 1, 2020
Oct 11, 2020

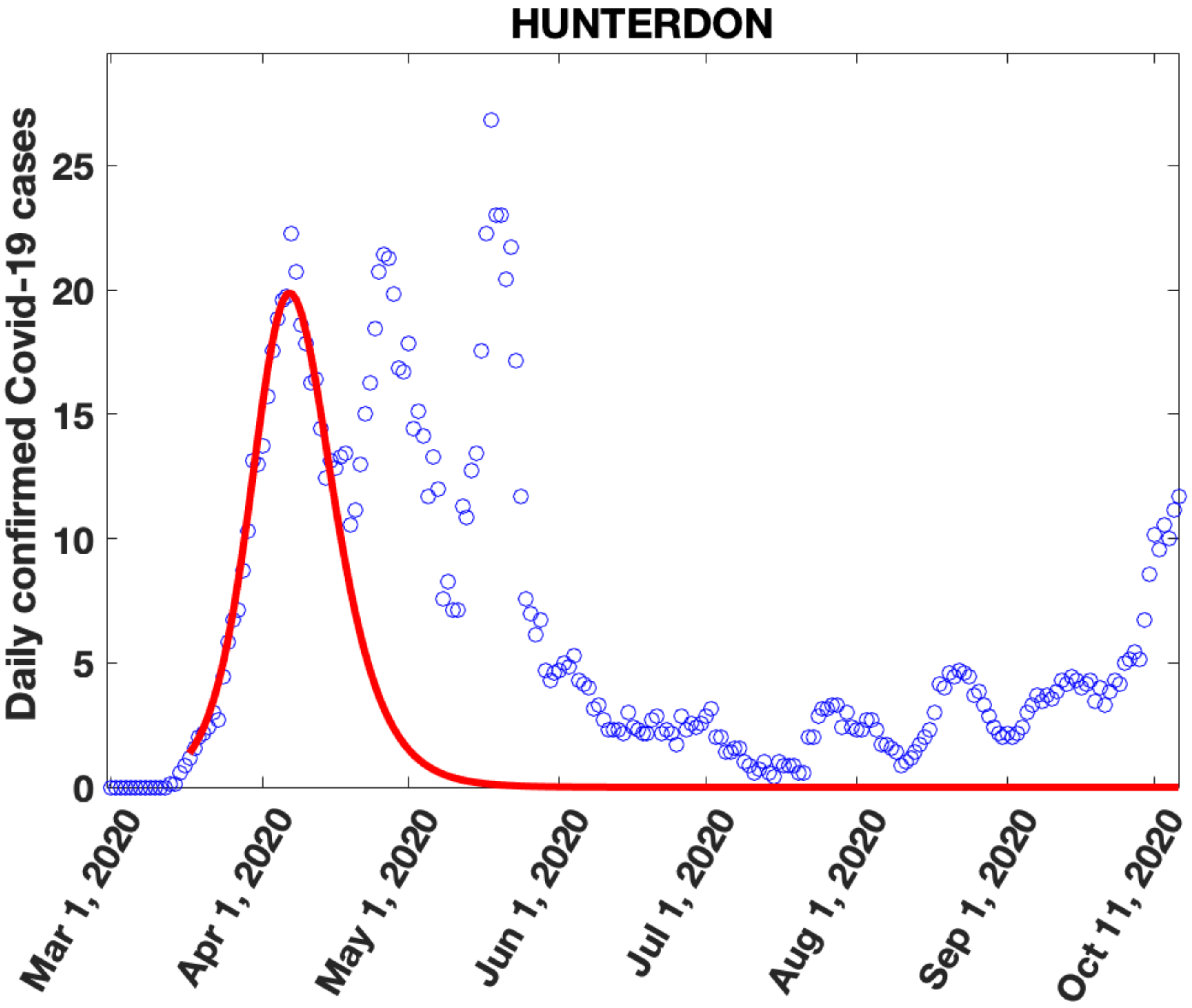
HUNTERDON
Daily confirmed Covid-19 cases
0
5
10
15
20
25
Mar 1, 2020
Apr 1, 2020
May 1, 2020
Jun 1, 2020
Jul 1, 2020
Aug 1, 2020
Sep 1, 2020
Oct 11, 2020

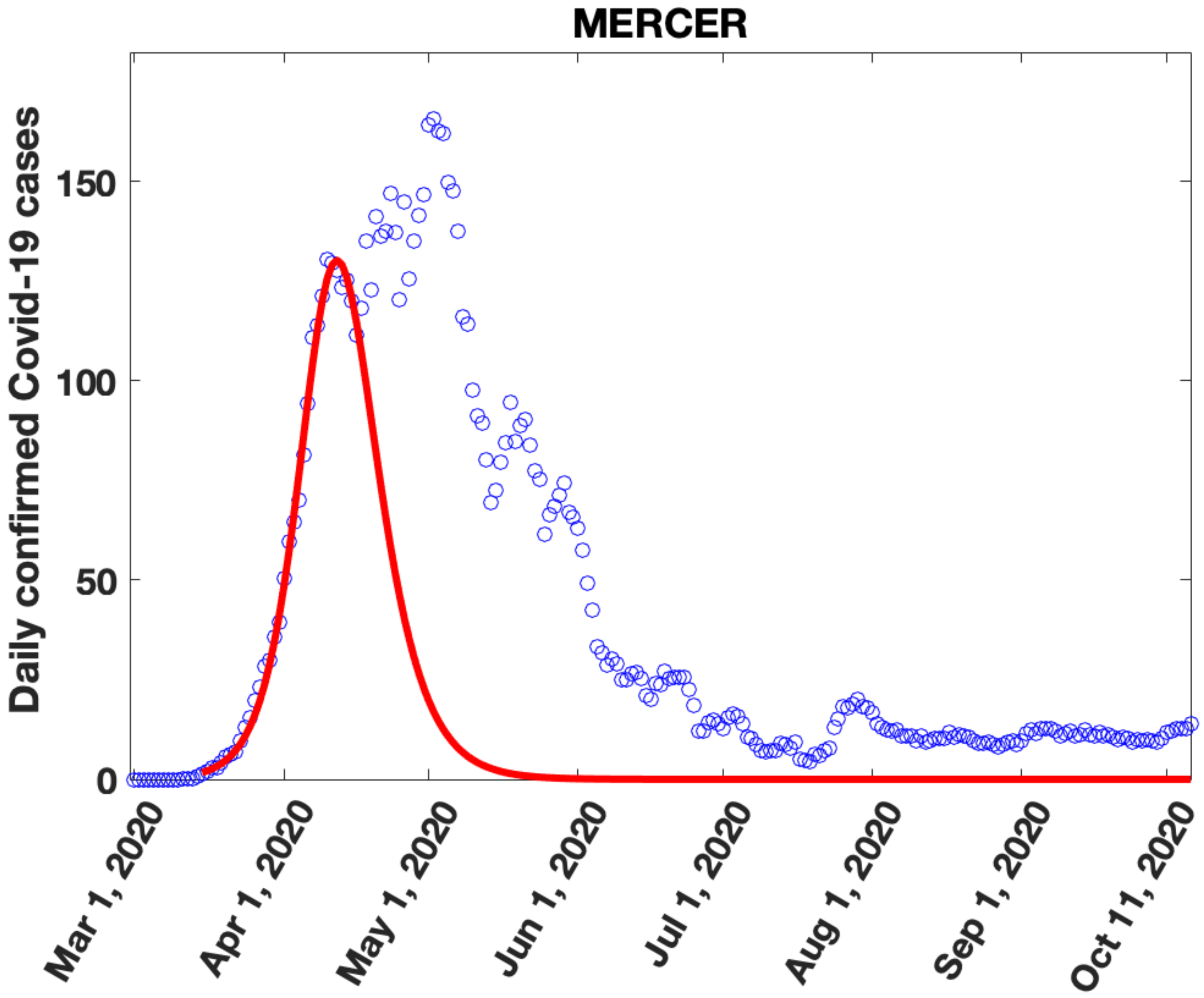
MERCER
Daily confirmed Covid-19 cases
150
100
50
0
Mar 1, 2020
Apr 1, 2020
May 1, 2020
Jun 1, 2020
Jul 1, 2020
Aug 1, 2020
Sep 1, 2020
Oct 11, 2020

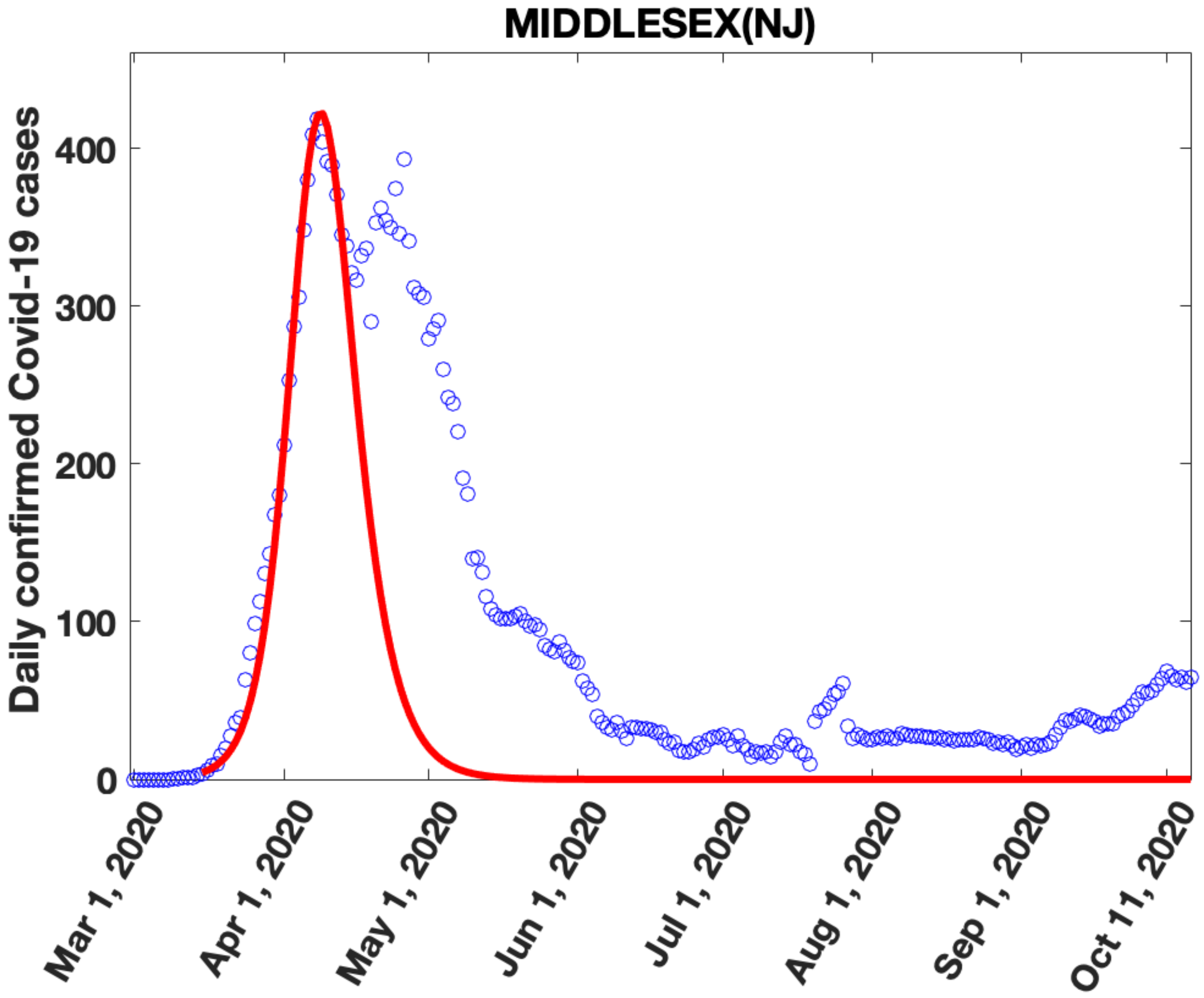
MIDDLESEX(NJ)
Daily confirmed Covid-19 cases
0
100
200
300
400
Mar 1, 2020
Apr 1, 2020
May 1, 2020
Jun 1, 2020
Jul 1, 2020
Aug 1, 2020
Sep 1, 2020
Oct 11, 2020

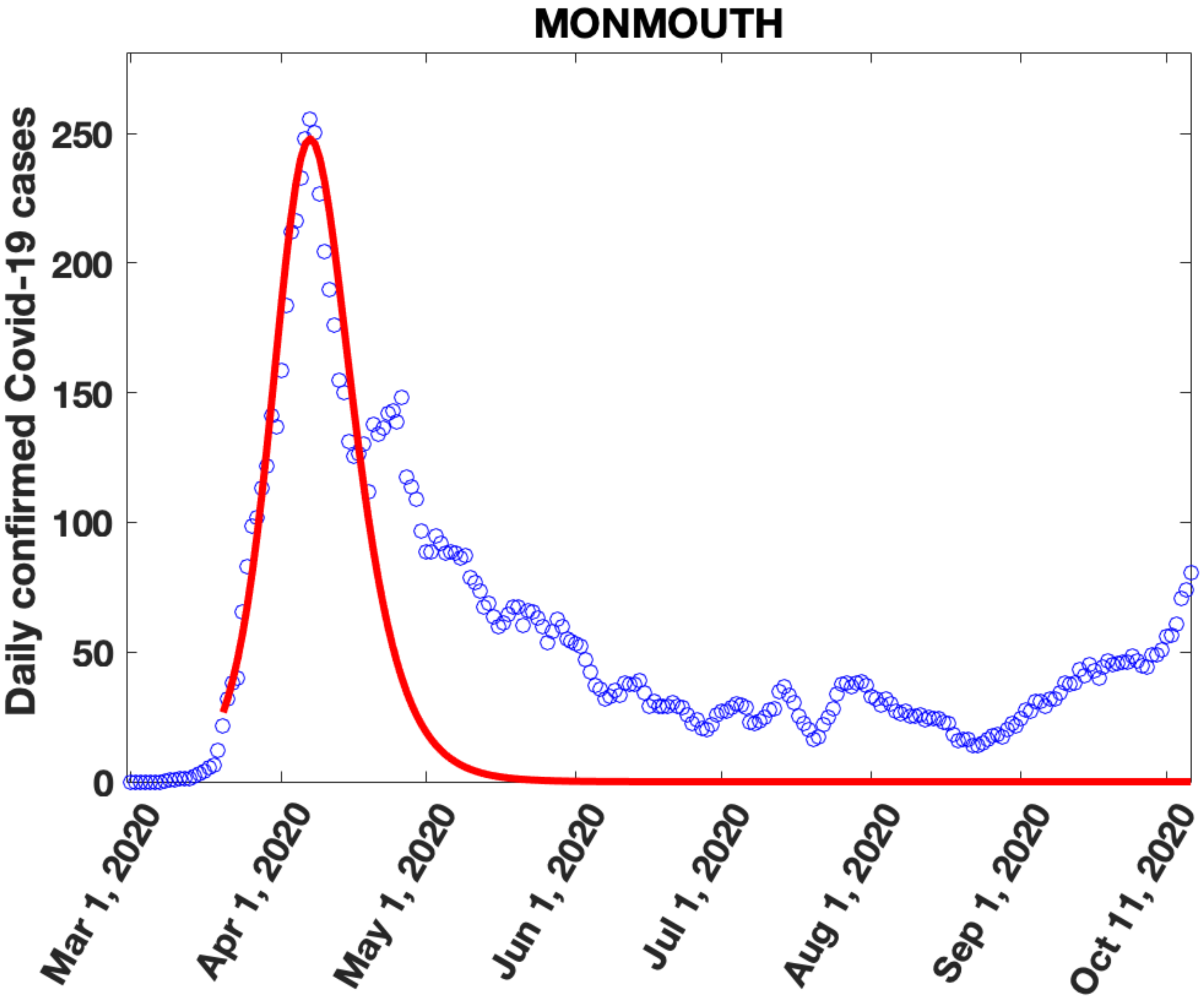

MONMOUTH
Daily confirmed Covid-19 cases
250
200
150
100
50
0
Mar 1, 2020
Apr 1, 2020
May 1, 2020
Jun 1, 2020
Jul 1, 2020
Aug 1, 2020
Sep 1, 2020
Oct 11, 2020

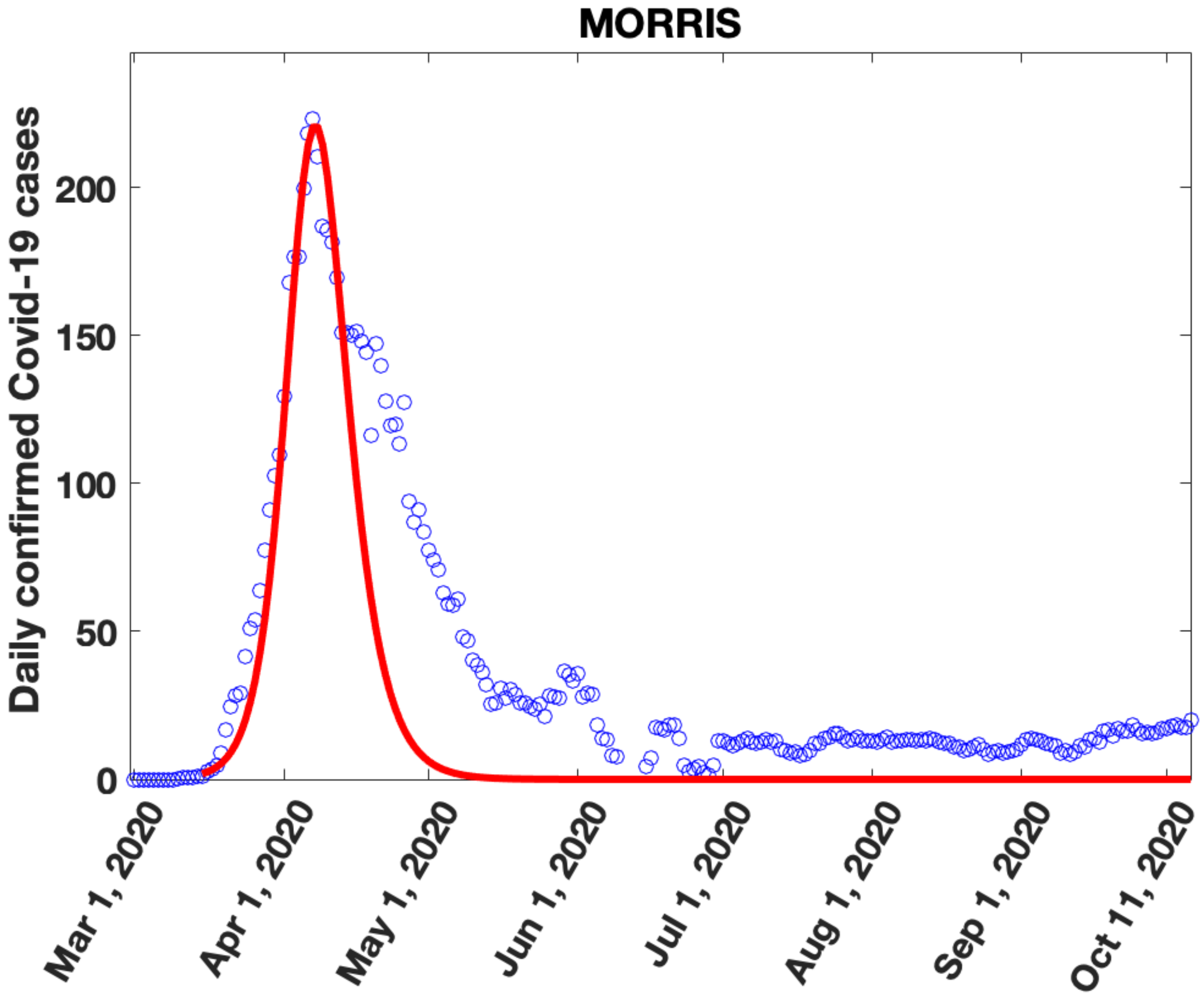
MORRIS
Daily confirmed Covid-19 cases
0
50
100
150
200
Mar 1, 2020
Apr 1, 2020
May 1, 2020
Jun 1, 2020
Jul 1, 2020
Aug 1, 2020
Sep 1, 2020
Oct 11, 2020

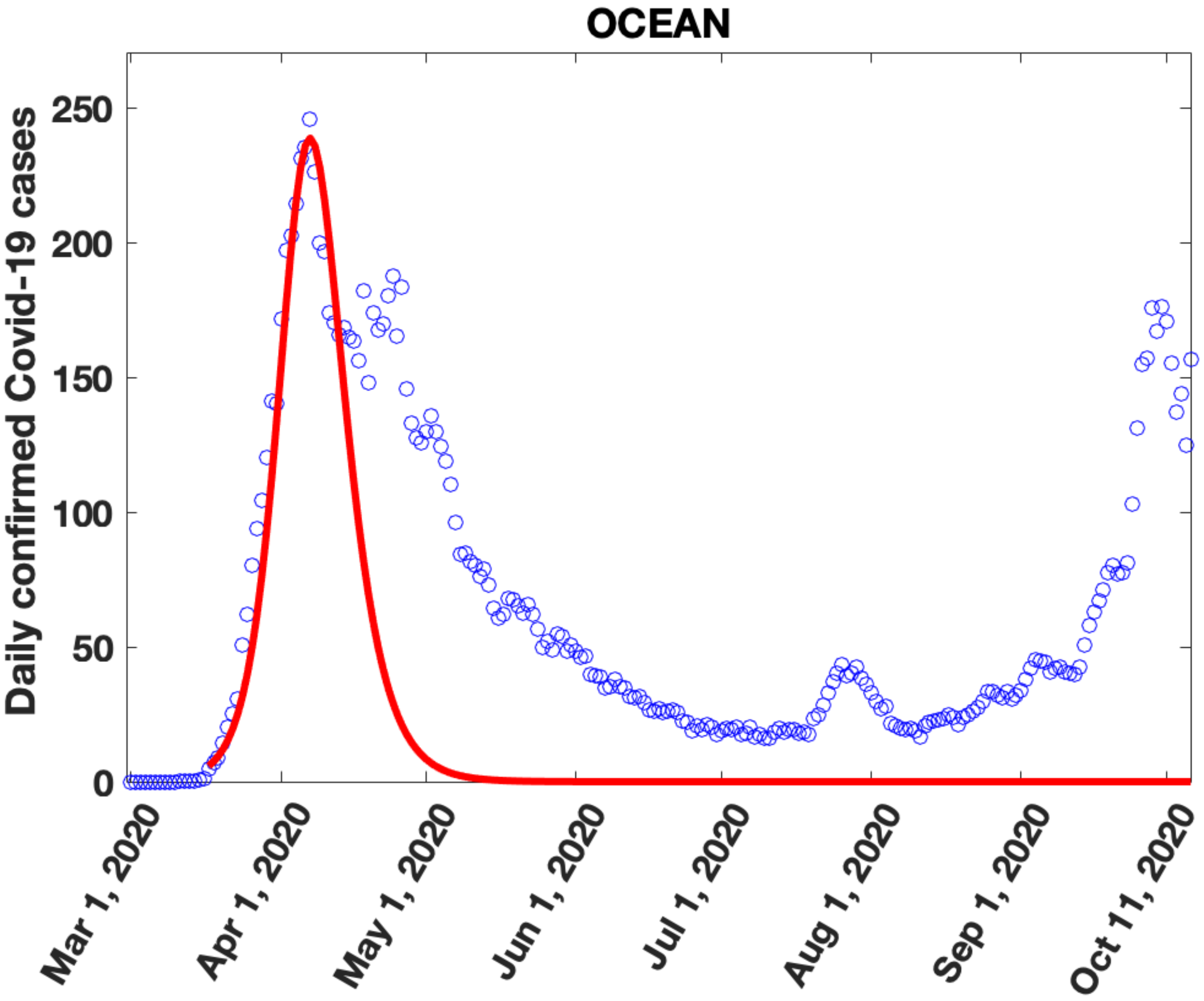
OCEAN
Daily confirmed Covid-19 cases
250
200
150
100
50
0
Mar 1, 2020
Apr 1, 2020
May 1, 2020
Jun 1, 2020
Jul 1, 2020
Aug 1, 2020
Sep 1, 2020
Oct 11, 2020

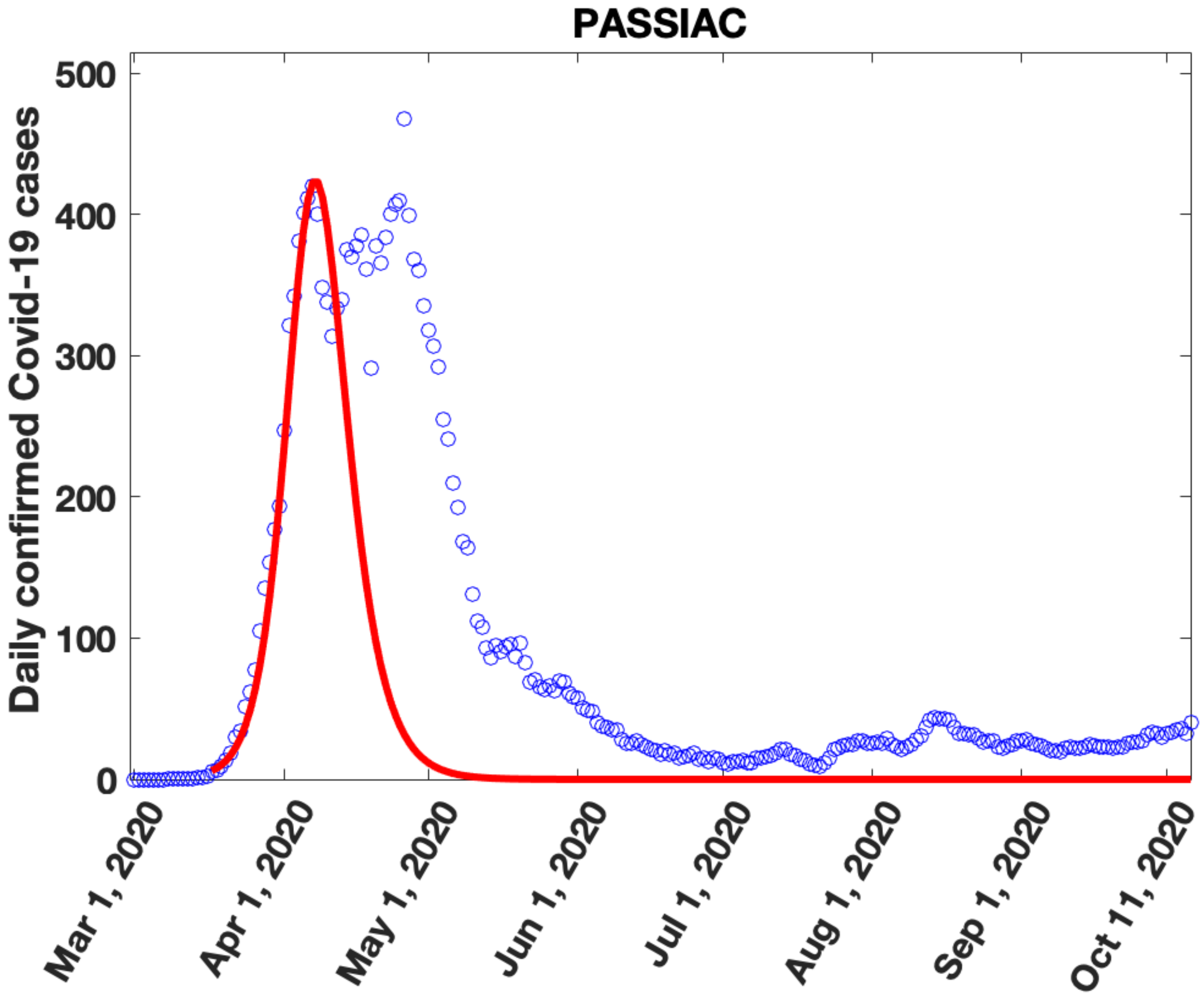
PASSIAC
Daily confirmed Covid-19 cases
0
100
200
300
400
500
Mar 1, 2020
Apr 1, 2020
May 1, 2020
Jun 1, 2020
Jul 1, 2020
Aug 1, 2020
Sep 1, 2020
Oct 11, 2020

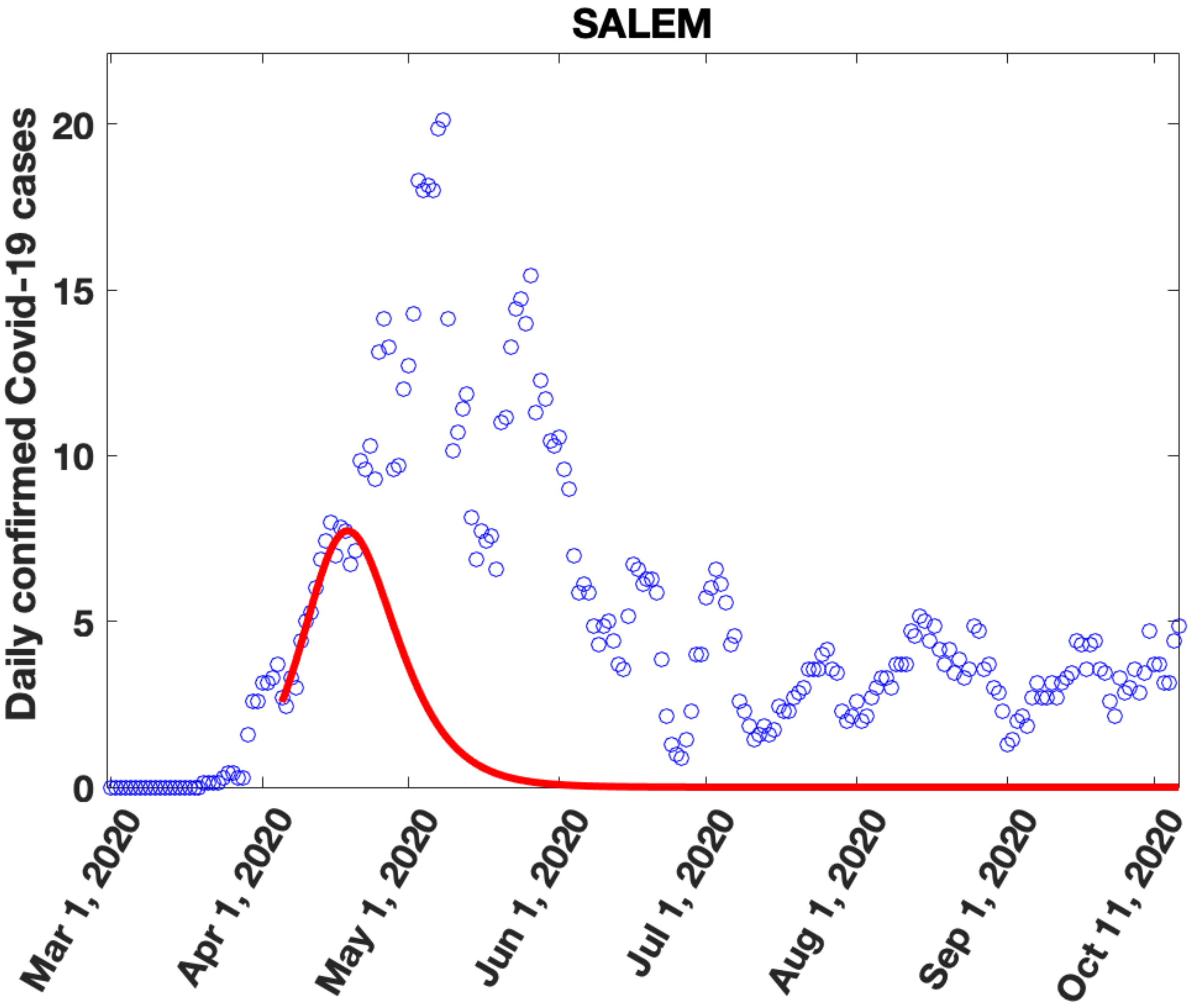
SALEM
Daily confirmed Covid-19 cases
20
15
10
5
0
Mar 1, 2020
Apr 1, 2020
May 1, 2020
Jun 1, 2020
Jul 1, 2020
Aug 1, 2020
Sep 1, 2020
Oct 11, 2020

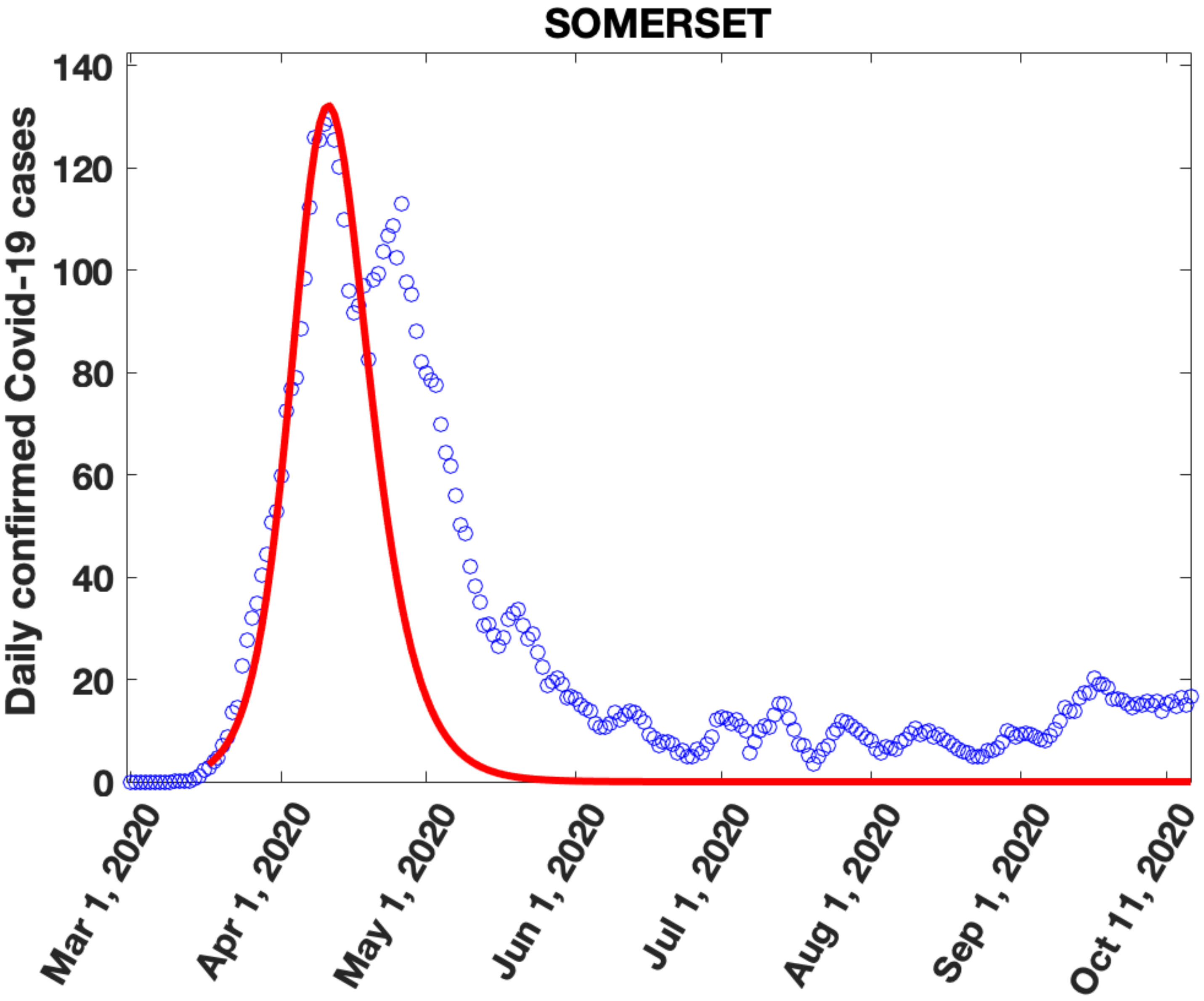

SOMERSET
Daily confirmed Covid-19 cases
140
120
100
80
60
40
20
0
Mar 1, 2020
Apr 1, 2020
May 1, 2020
Jun 1, 2020
Jul 1, 2020
Aug 1, 2020
Sep 1, 2020
Oct 11, 2020

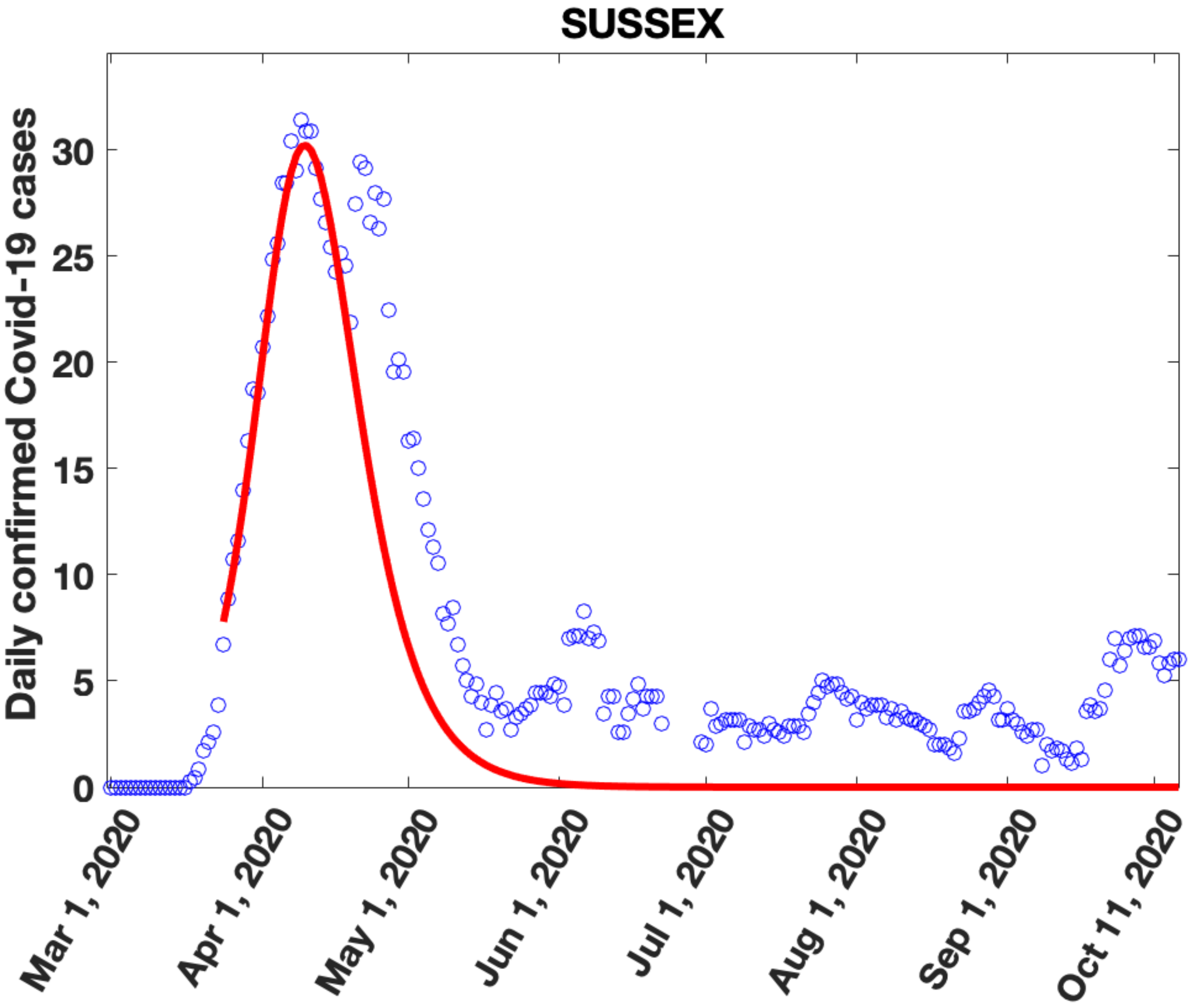

SUSSEX
Daily confirmed Covid-19 cases
0
5
10
15
20
25
30
Mar 1, 2020
Apr 1, 2020
May 1, 2020
Jun 1, 2020
Jul 1, 2020
Aug 1, 2020
Sep 1, 2020
Oct 11, 2020

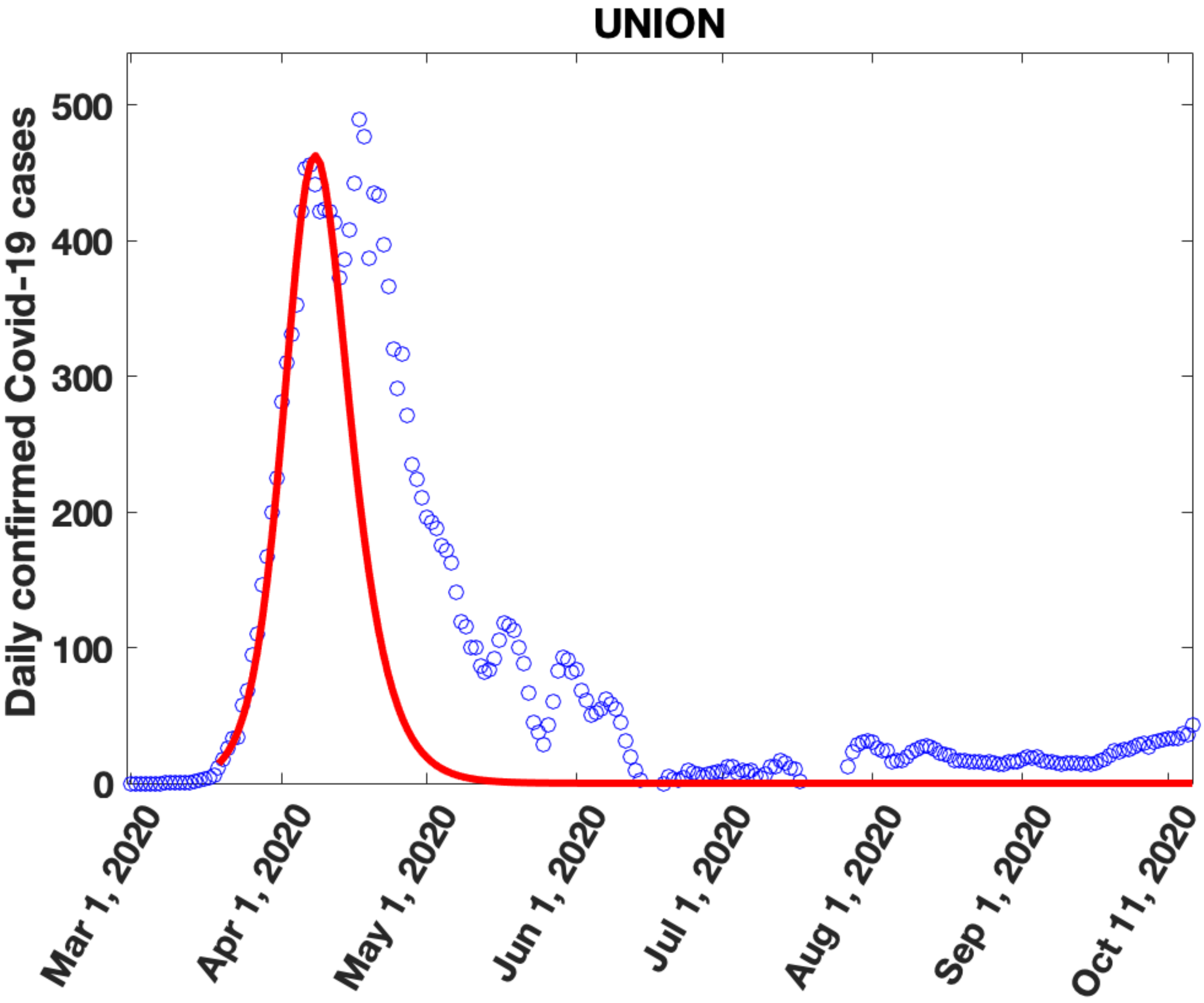
UNION
Daily confirmed Covid-19 cases
500
400
300
200
100
0
Mar 1, 2020
Apr 1, 2020
May 1, 2020
Jun 1, 2020
Jul 1, 2020
Aug 1, 2020
Sep 1, 2020
Oct 11, 2020

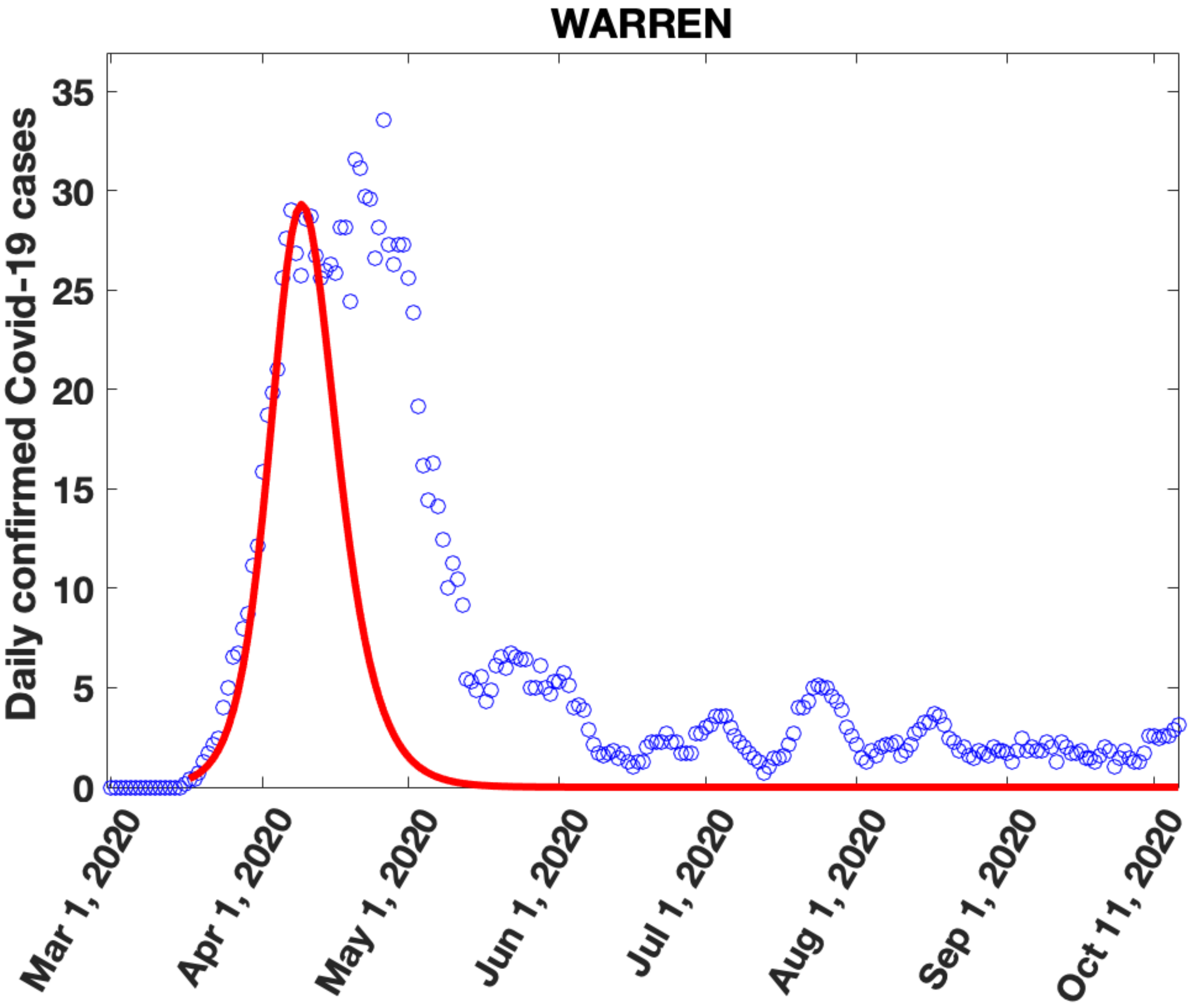

WARREN
Daily confirmed Covid-19 cases
0
5
10
15
20
25
30
35
Mar 1, 2020
Apr 1, 2020
May 1, 2020
Jun 1, 2020
Jul 1, 2020
Aug 1, 2020
Sep 1, 2020
Oct 11, 2020
