## Supplementary Figure 7 for "Migration of households from New York City and the Second Peak in Covid-19 cases in New Jersey, Connecticut and New York Counties"

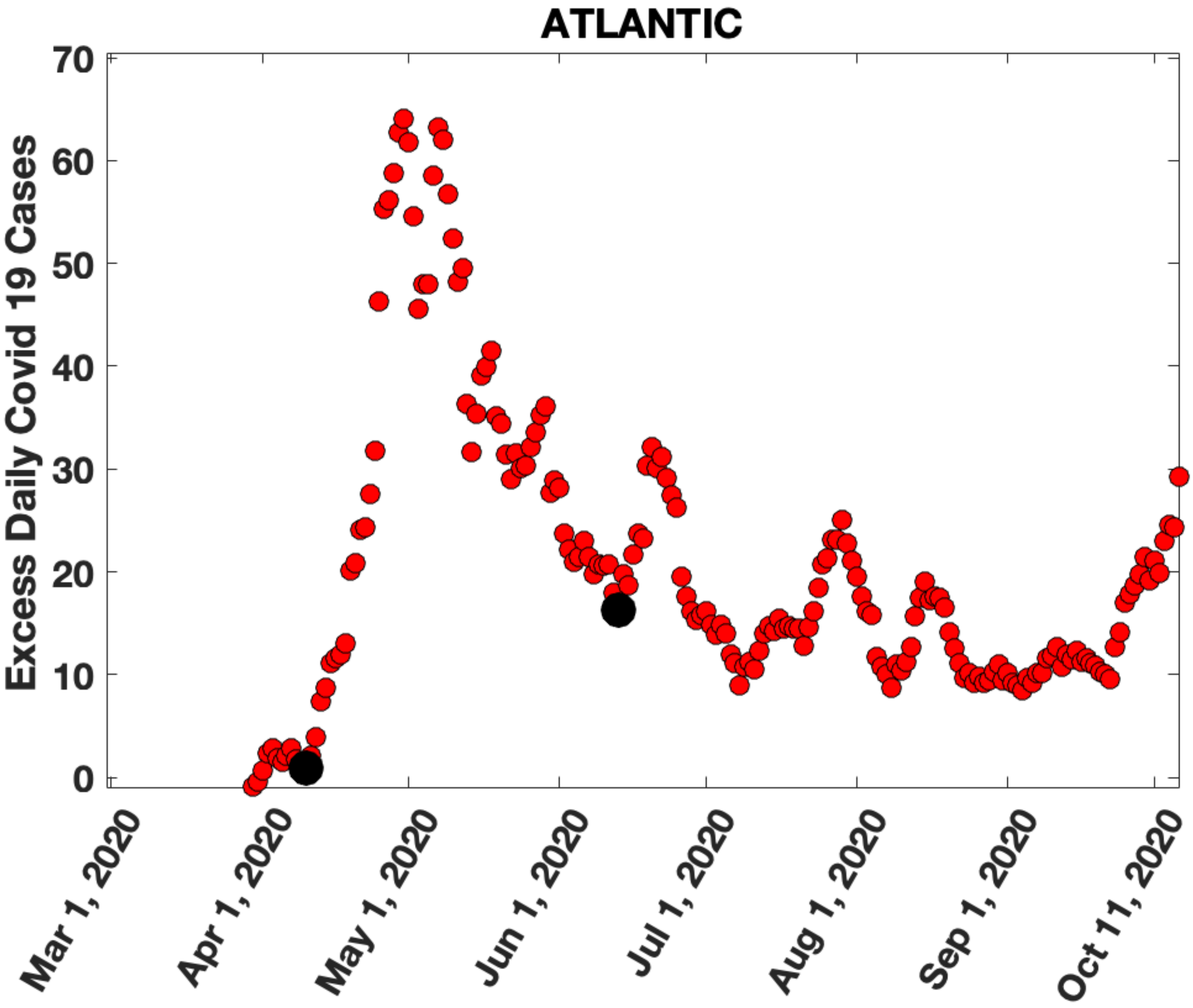
ATLANTIC
Excess Daily Covid 19 Cases
70
60
50
40
30
20
10
0
Mar 1, 2020
Apr 1, 2020
May 1, 2020
Jun 1, 2020
Jul 1, 2020
Aug 1, 2020
Sep 1, 2020
Oct 11, 2020

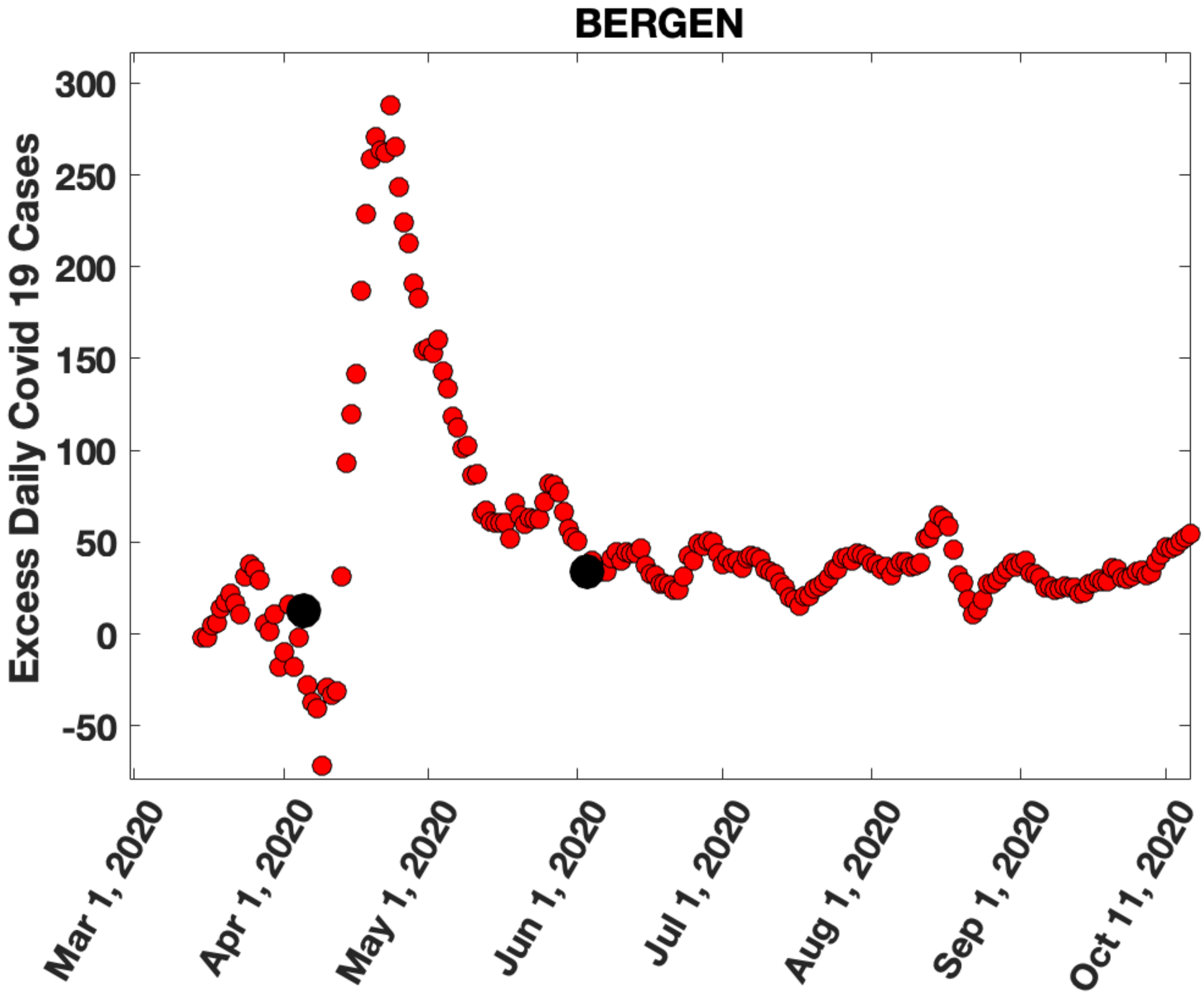

BERGEN
Excess Daily Covid 19 Cases
300
250
200
150
100
50
0
-50
Mar 1, 2020
Apr 1, 2020
May 1, 2020
Jun 1, 2020
Jul 1, 2020
Aug 1, 2020
Sep 1, 2020
Oct 11, 2020

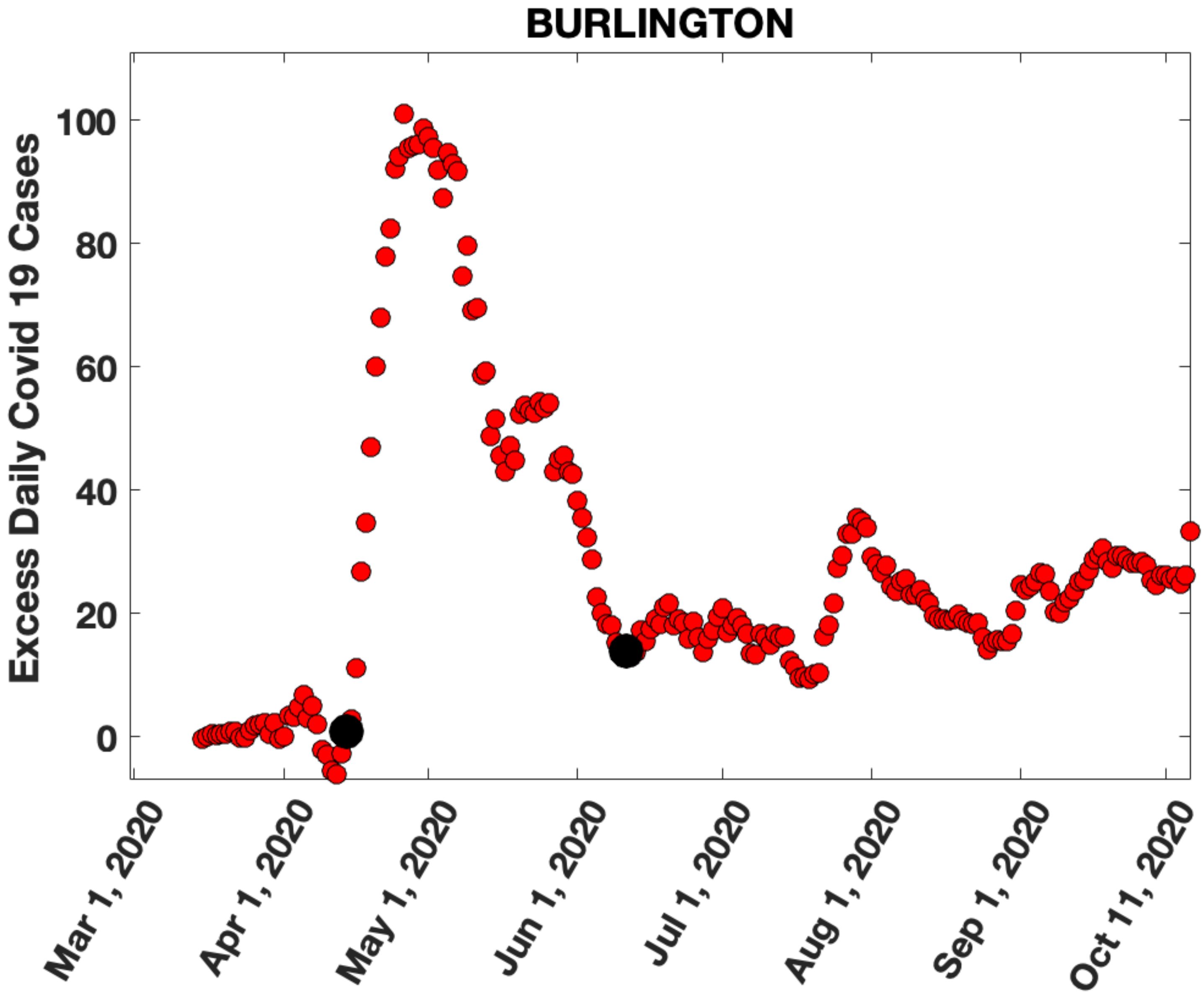

BURLINGTON
Excess Daily Covid 19 Cases
100
80
60
40
20
0
Mar 1, 2020
Apr 1, 2020
May 1, 2020
Jun 1, 2020
Jul 1, 2020
Aug 1, 2020
Sep 1, 2020
Oct 11, 2020

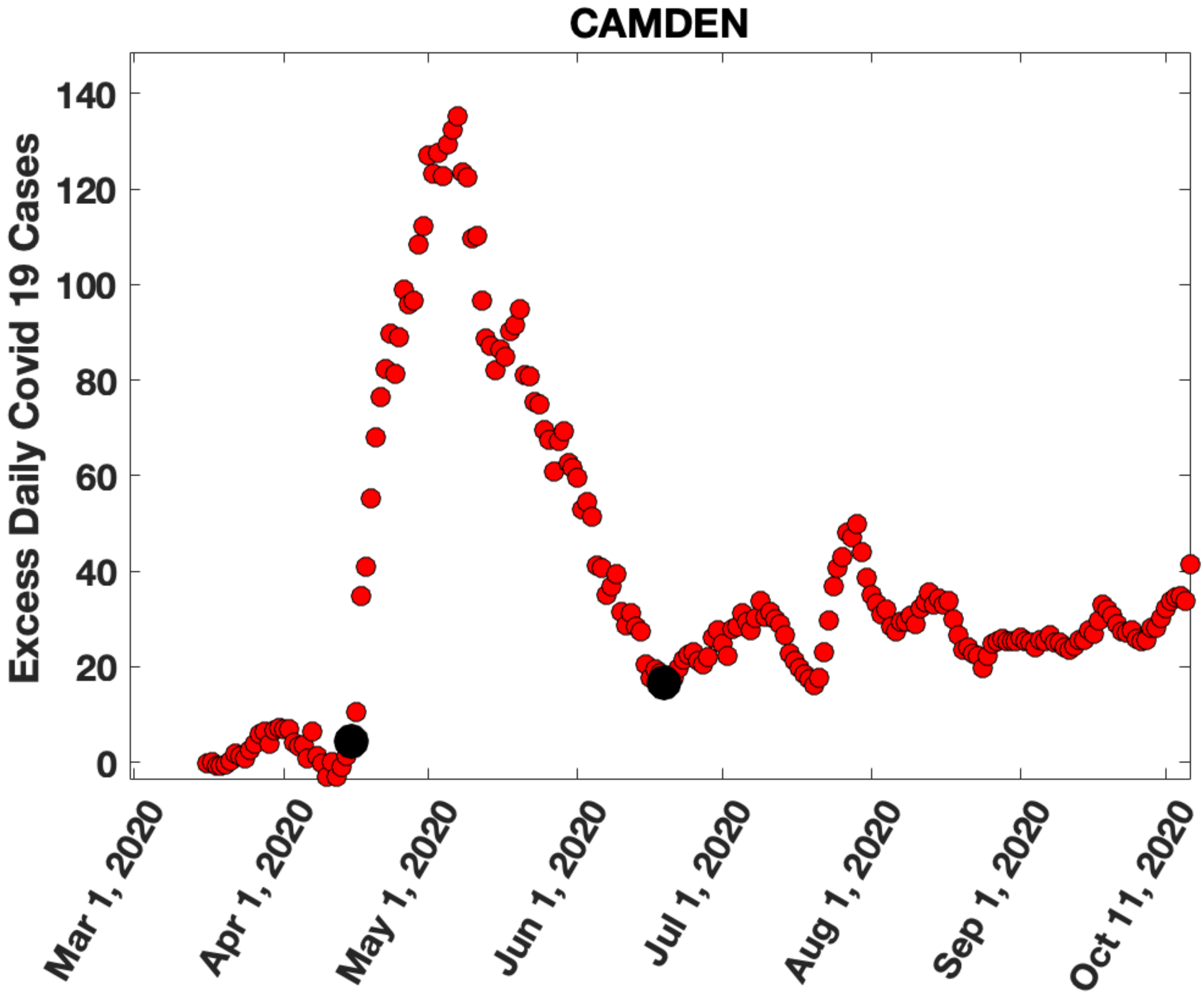

CAMDEN
Excess Daily Covid 19 Cases
140
120
100
80
60
40
20
0
Mar 1, 2020
Apr 1, 2020
May 1, 2020
Jun 1, 2020
Jul 1, 2020
Aug 1, 2020
Sep 1, 2020
Oct 11, 2020

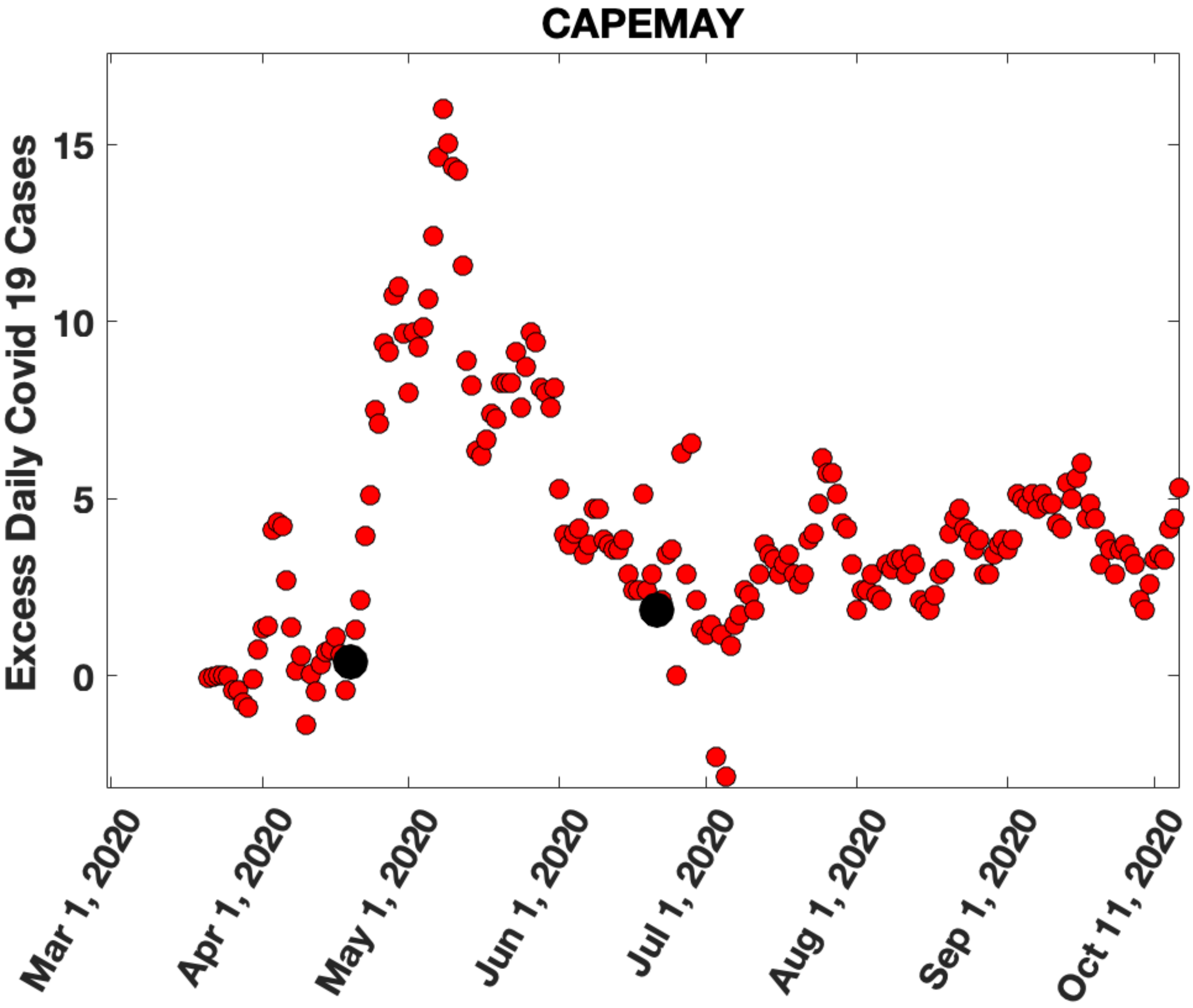
CAPEMAY
Excess Daily Covid 19 Cases
15
10
5
0
Mar 1, 2020
Apr 1, 2020
May 1, 2020
Jun 1, 2020
Jul 1, 2020
Aug 1, 2020
Sep 1, 2020
Oct 11, 2020

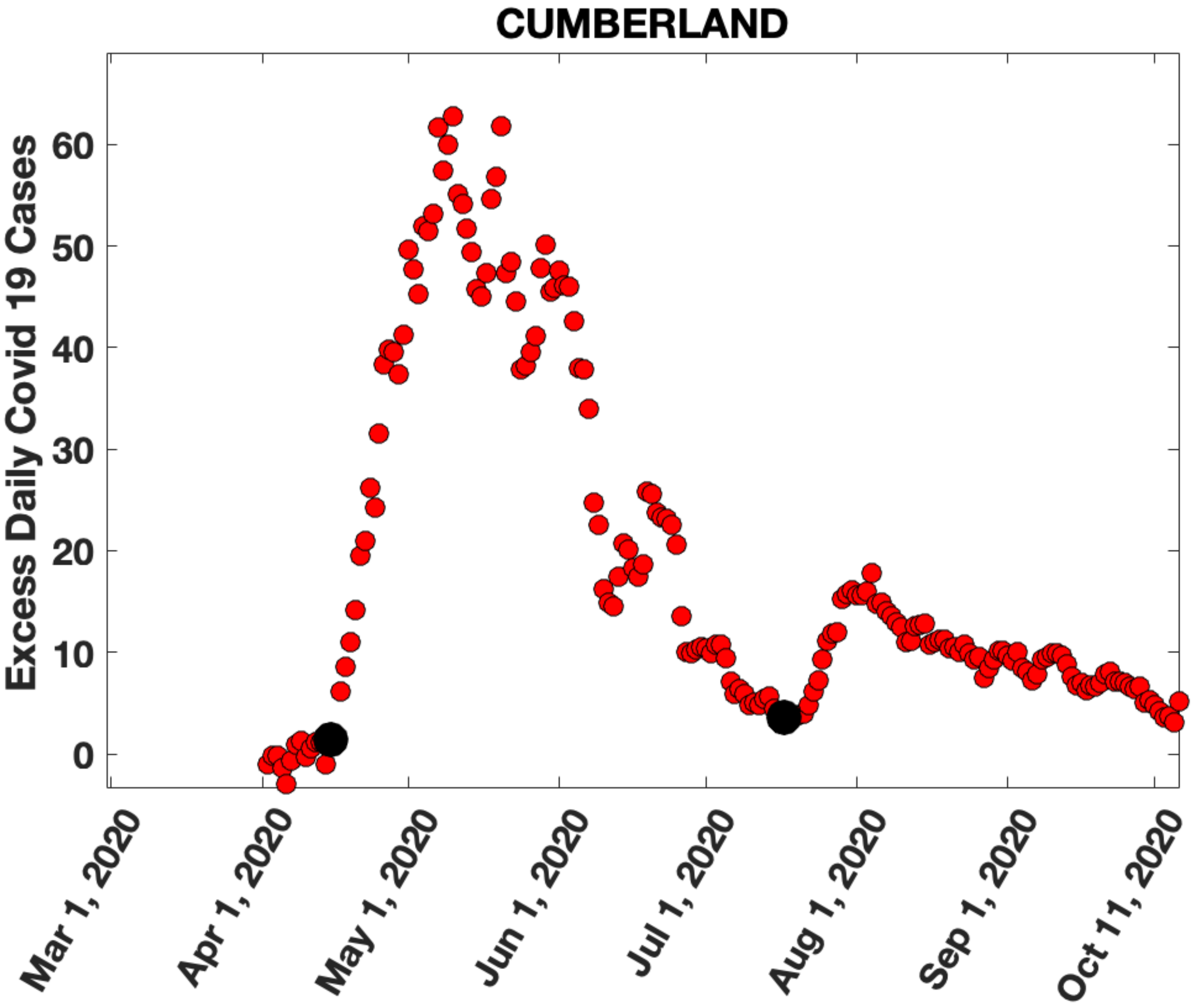

CUMBERLAND
Excess Daily Covid 19 Cases
0
10
20
30
40
50
60
Mar 1, 2020
Apr 1, 2020
May 1, 2020
Jun 1, 2020
Jul 1, 2020
Aug 1, 2020
Sep 1, 2020
Oct 11, 2020

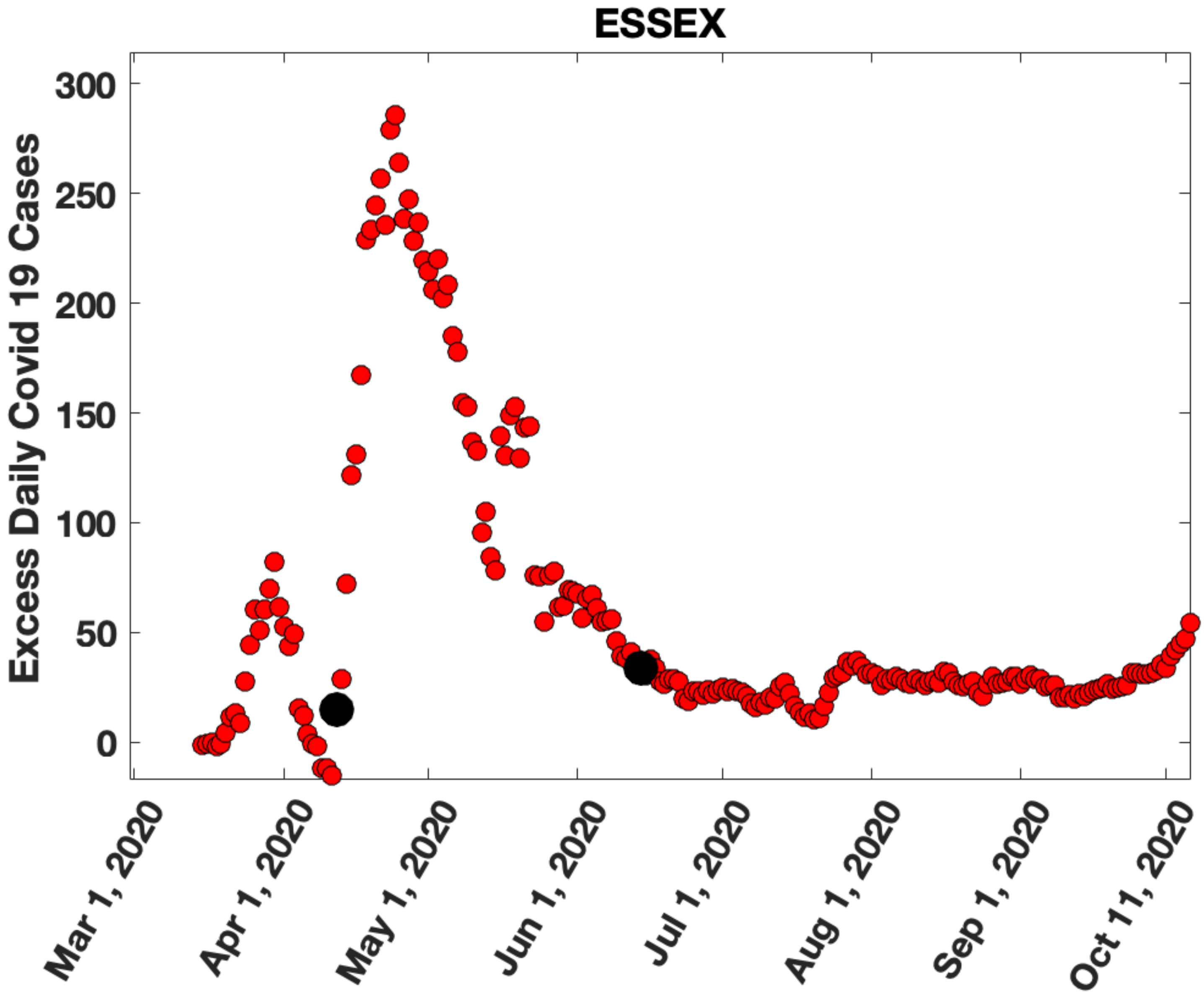

ESSEX
Excess Daily Covid 19 Cases
300
250
200
150
100
50
0
Mar 1, 2020
Apr 1, 2020
May 1, 2020
Jun 1, 2020
Jul 1, 2020
Aug 1, 2020
Sep 1, 2020
Oct 11, 2020

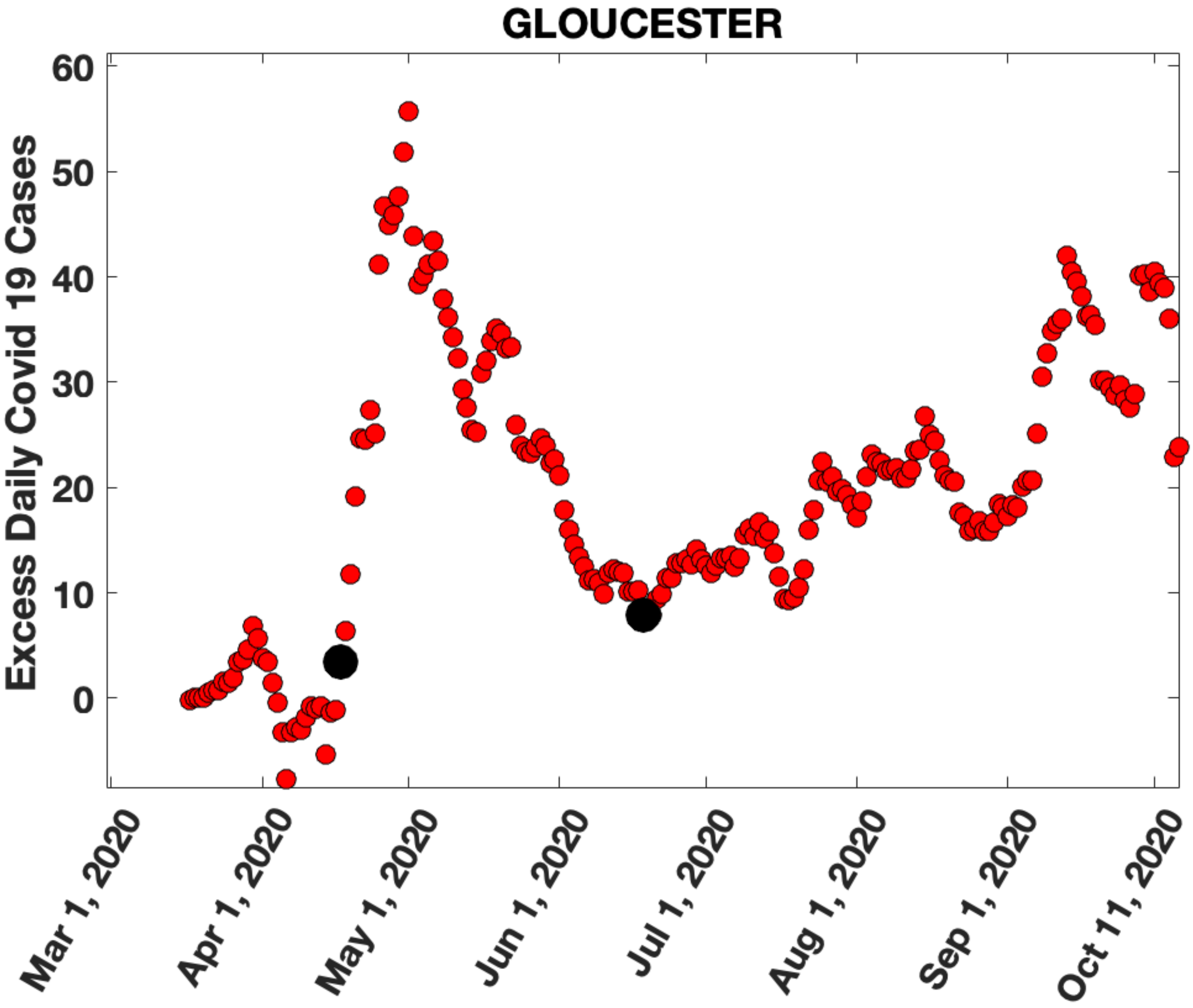
GLOUCESTER
Excess Daily Covid 19 Cases
60
50
40
30
20
10
0
Mar 1, 2020
Apr 1, 2020
May 1, 2020
Jun 1, 2020
Jul 1, 2020
Aug 1, 2020
Sep 1, 2020
Oct 11, 2020

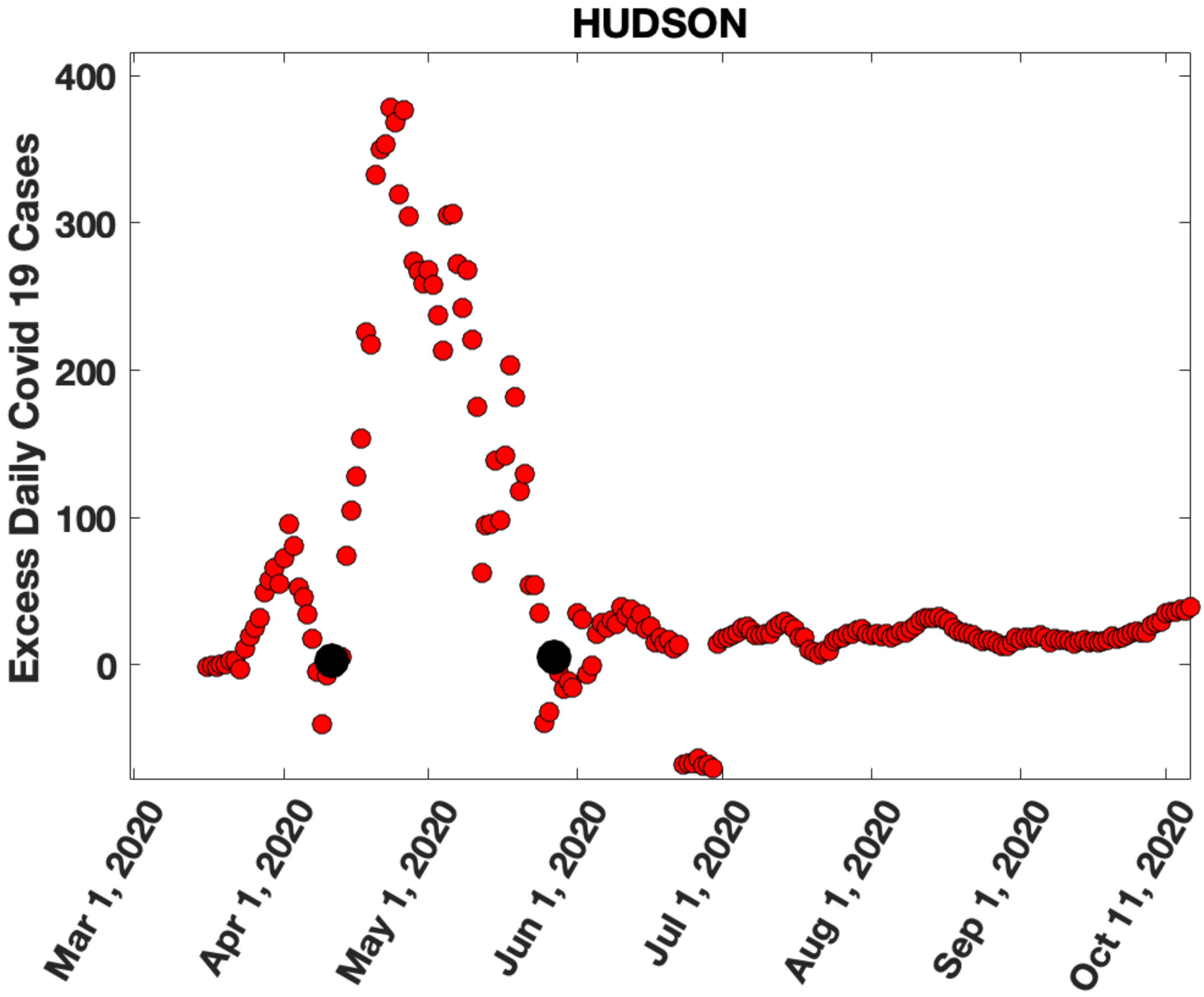
HUDSON
Excess Daily Covid 19 Cases
400
300
200
100
0
Mar 1, 2020
Apr 1, 2020
May 1, 2020
Jun 1, 2020
Jul 1, 2020
Aug 1, 2020
Sep 1, 2020
Oct 11, 2020

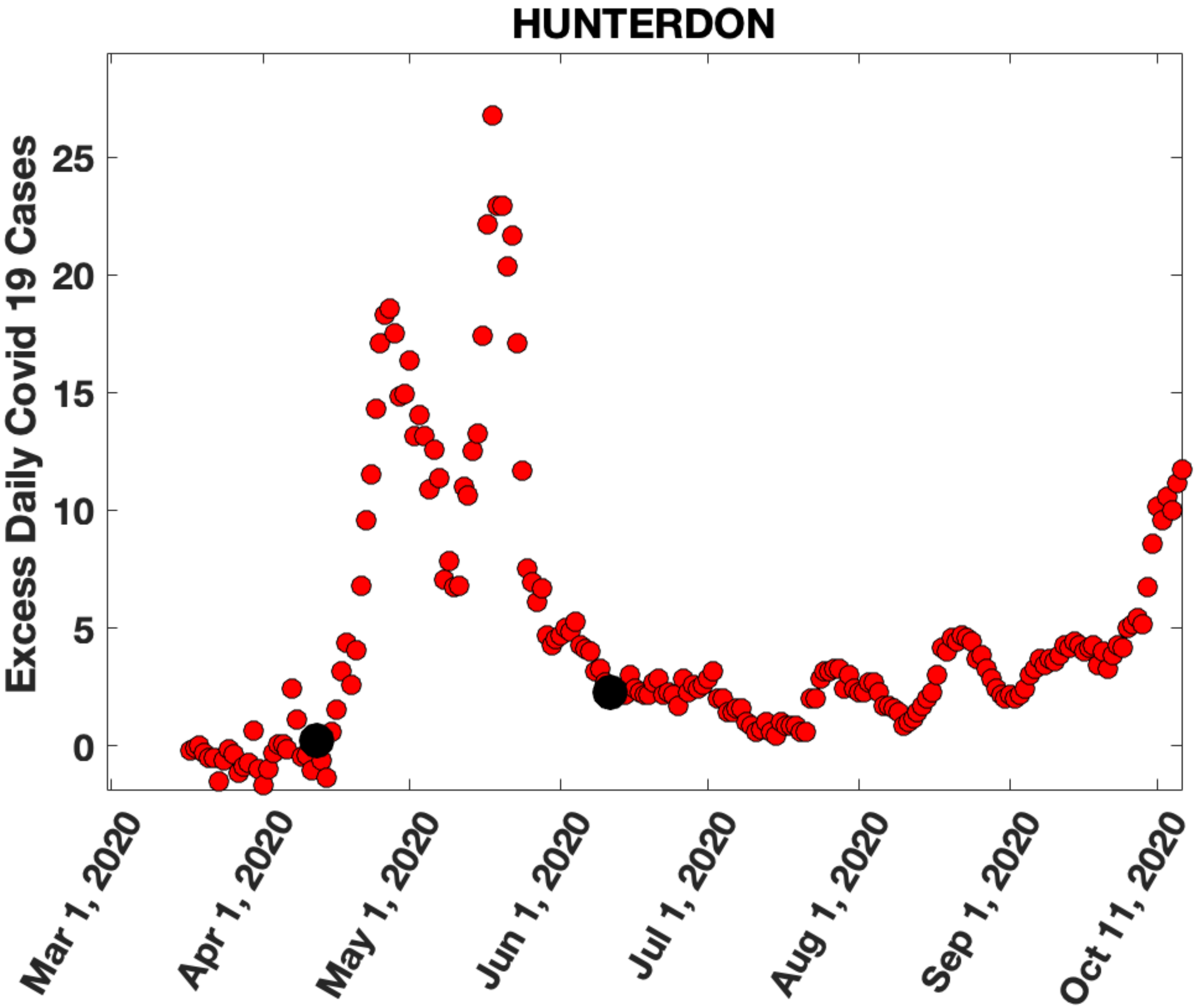

HUNTERDON
Excess Daily Covid 19 Cases
25
20
15
10
5
0
Mar 1, 2020
Apr 1, 2020
May 1, 2020
Jun 1, 2020
Jul 1, 2020
Aug 1, 2020
Sep 1, 2020
Oct 11, 2020

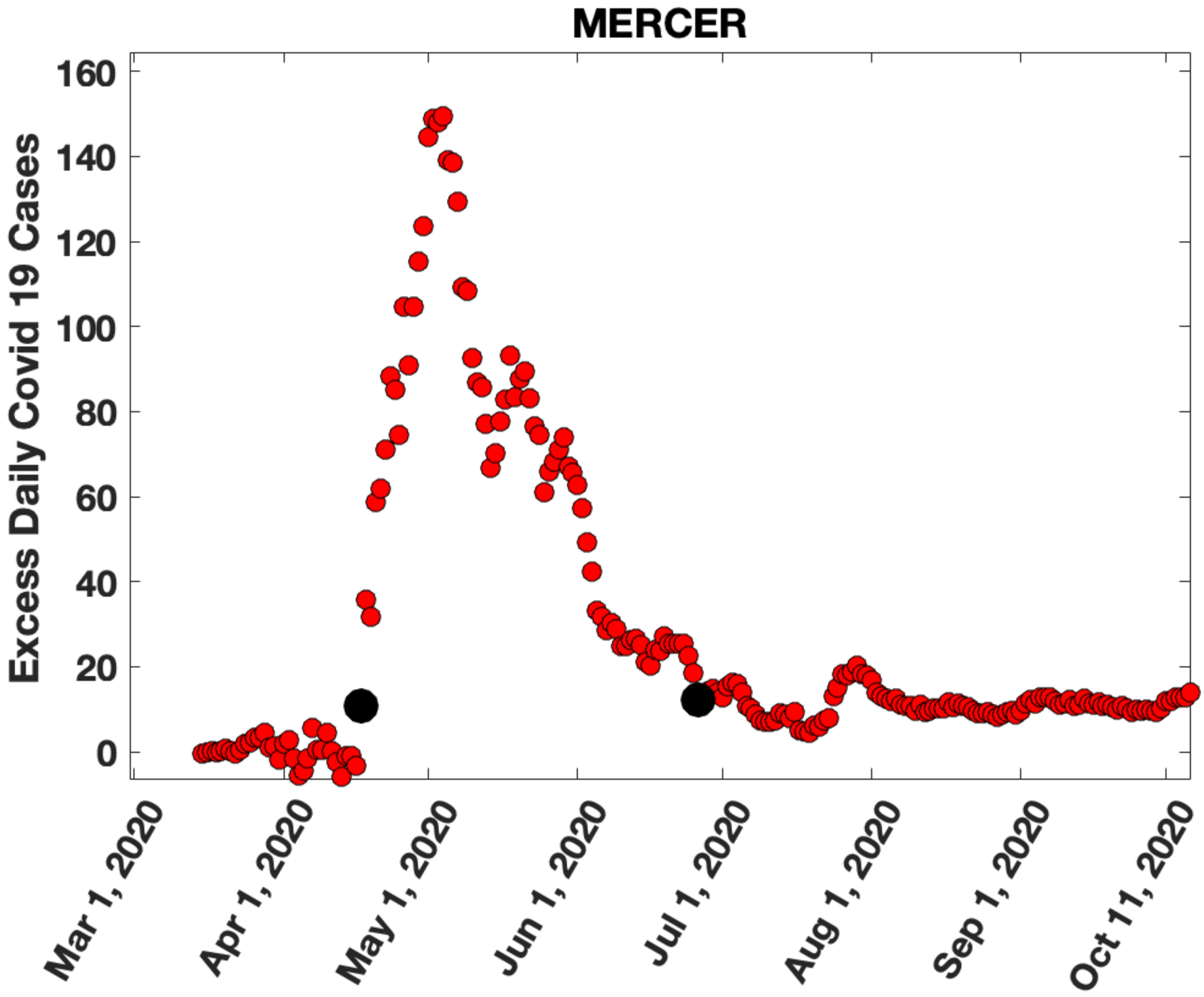
MERCER
Excess Daily Covid 19 Cases
0
20
40
60
80
100
120
140
160
Mar 1, 2020
Apr 1, 2020
May 1, 2020
Jun 1, 2020
Jul 1, 2020
Aug 1, 2020
Sep 1, 2020
Oct 11, 2020

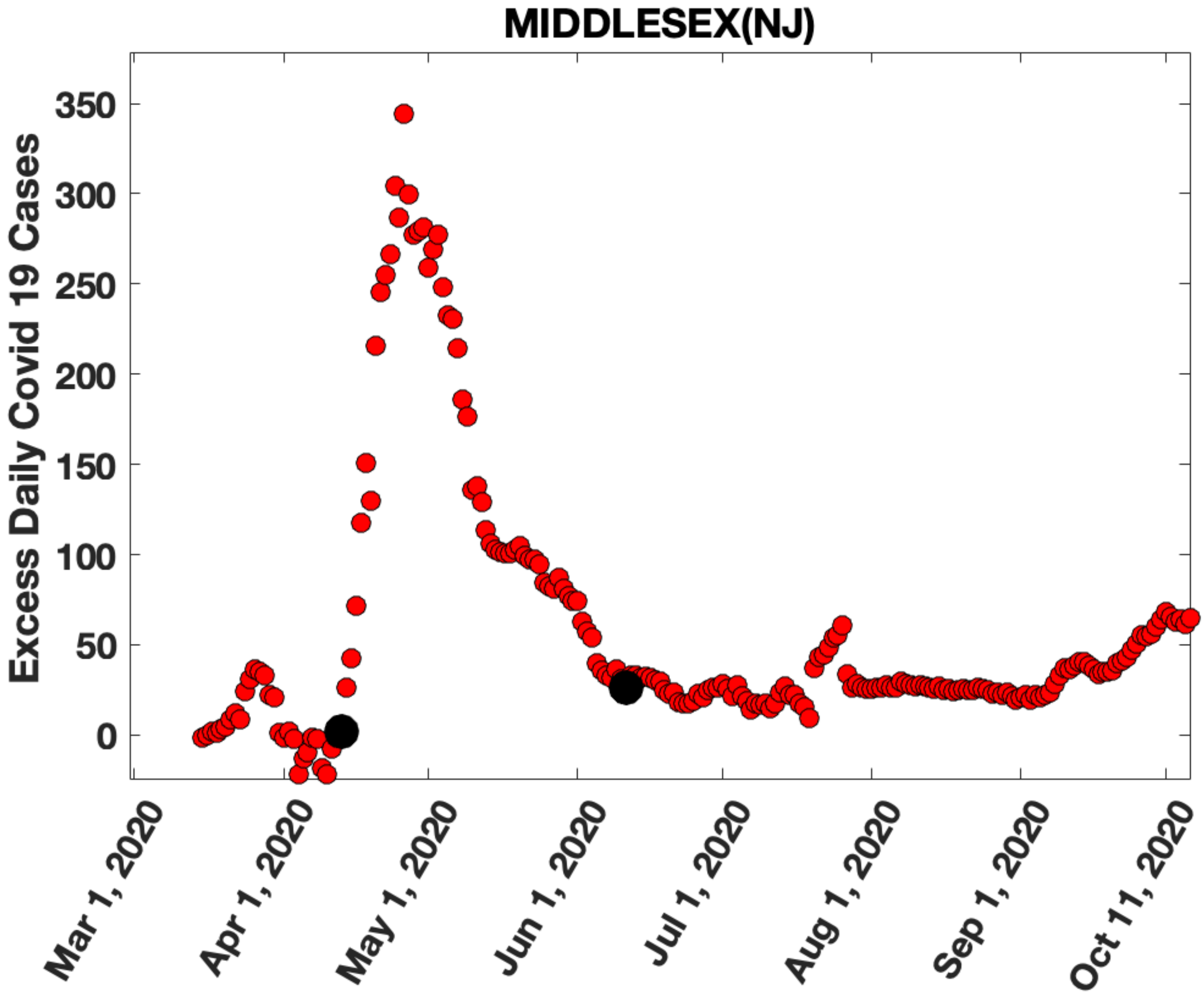

MIDDLESEX(NJ)
Excess Daily Covid 19 Cases
350
300
250
200
150
100
50
0
Mar 1, 2020
Apr 1, 2020
May 1, 2020
Jun 1, 2020
Jul 1, 2020
Aug 1, 2020
Sep 1, 2020
Oct 11, 2020

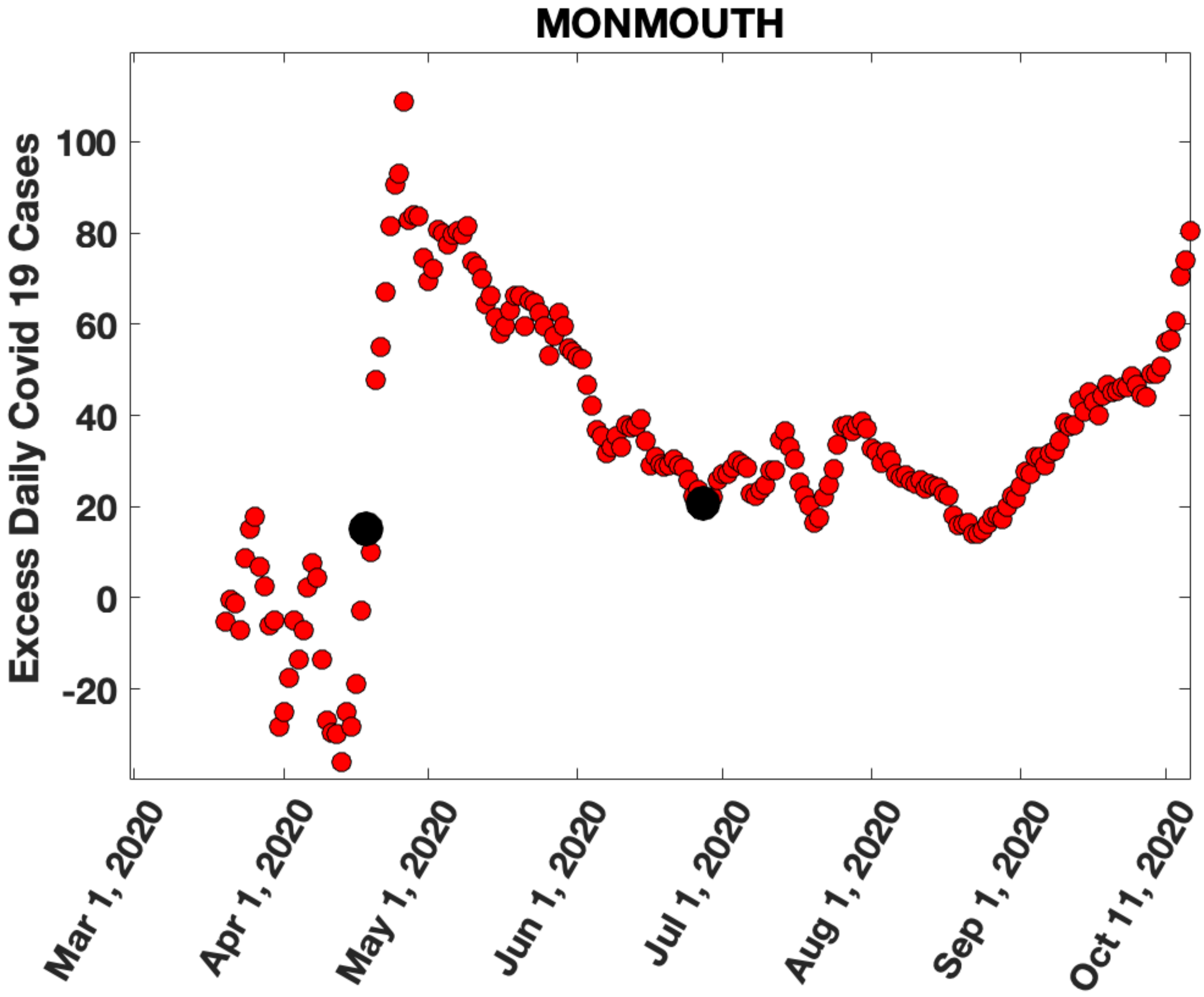
MONMOUTH
Excess Daily Covid 19 Cases
100
80
60
40
20
0
-20
Mar 1, 2020
Apr 1, 2020
May 1, 2020
Jun 1, 2020
Jul 1, 2020
Aug 1, 2020
Sep 1, 2020
Oct 11, 2020

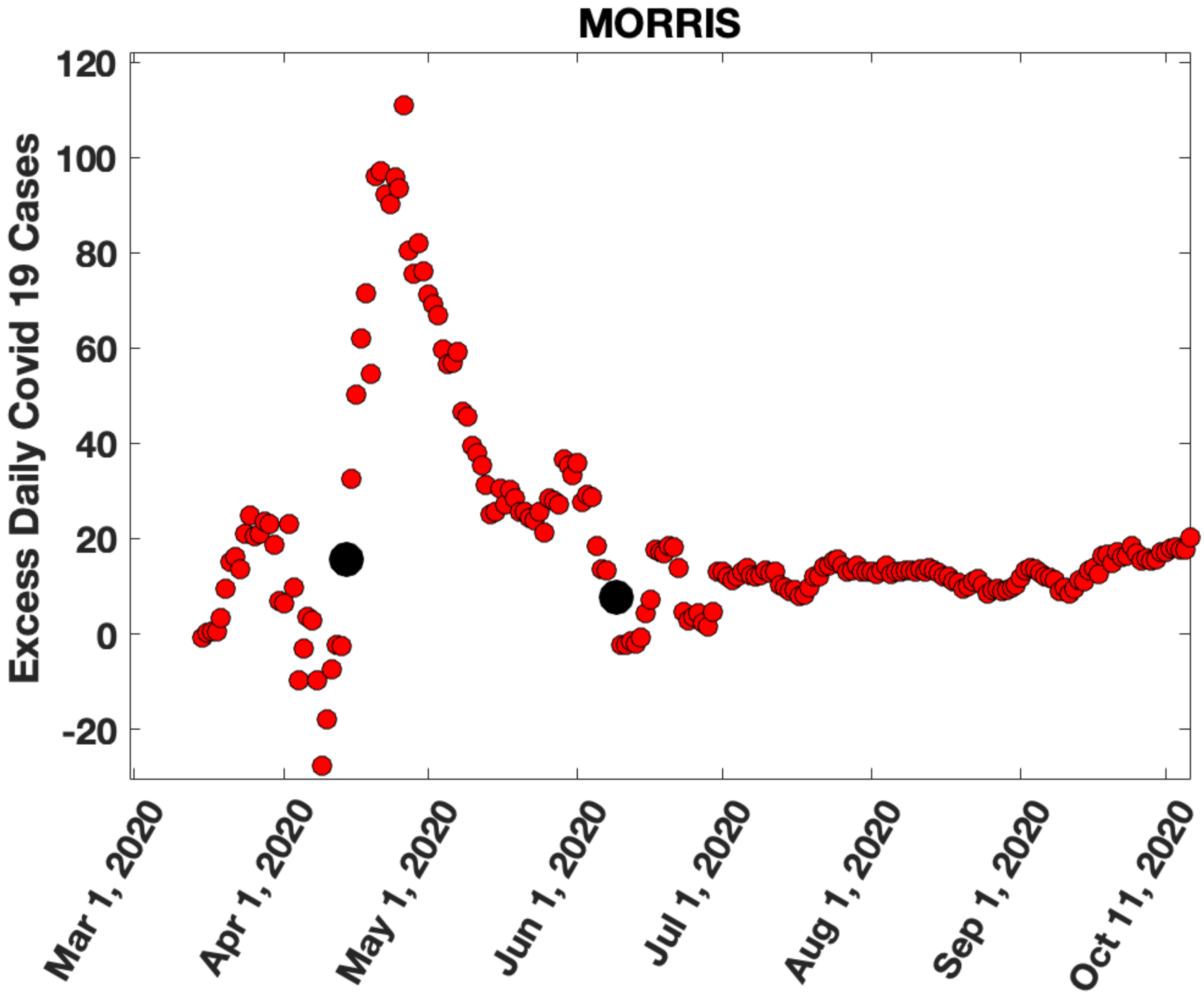
MORRIS
Excess Daily Covid 19 Cases
120
100
80
60
40
20
0
-20
Mar 1, 2020
Apr 1, 2020
May 1, 2020
Jun 1, 2020
Jul 1, 2020
Aug 1, 2020
Sep 1, 2020
Oct 11, 2020

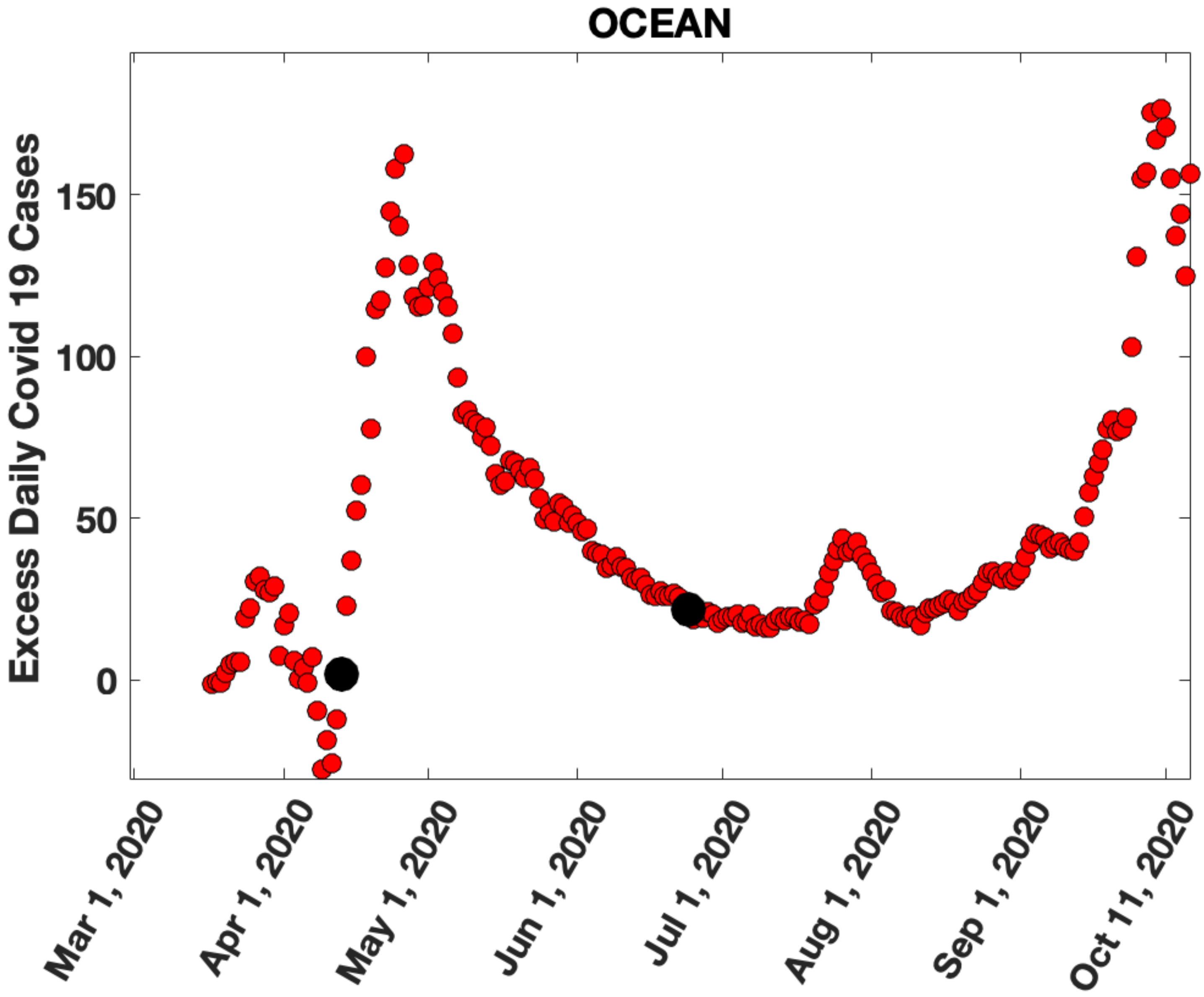
OCEAN
Excess Daily Covid 19 Cases
150
100
50
0
Mar 1, 2020
Apr 1, 2020
May 1, 2020
Jun 1, 2020
Jul 1, 2020
Aug 1, 2020
Sep 1, 2020
Oct 11, 2020

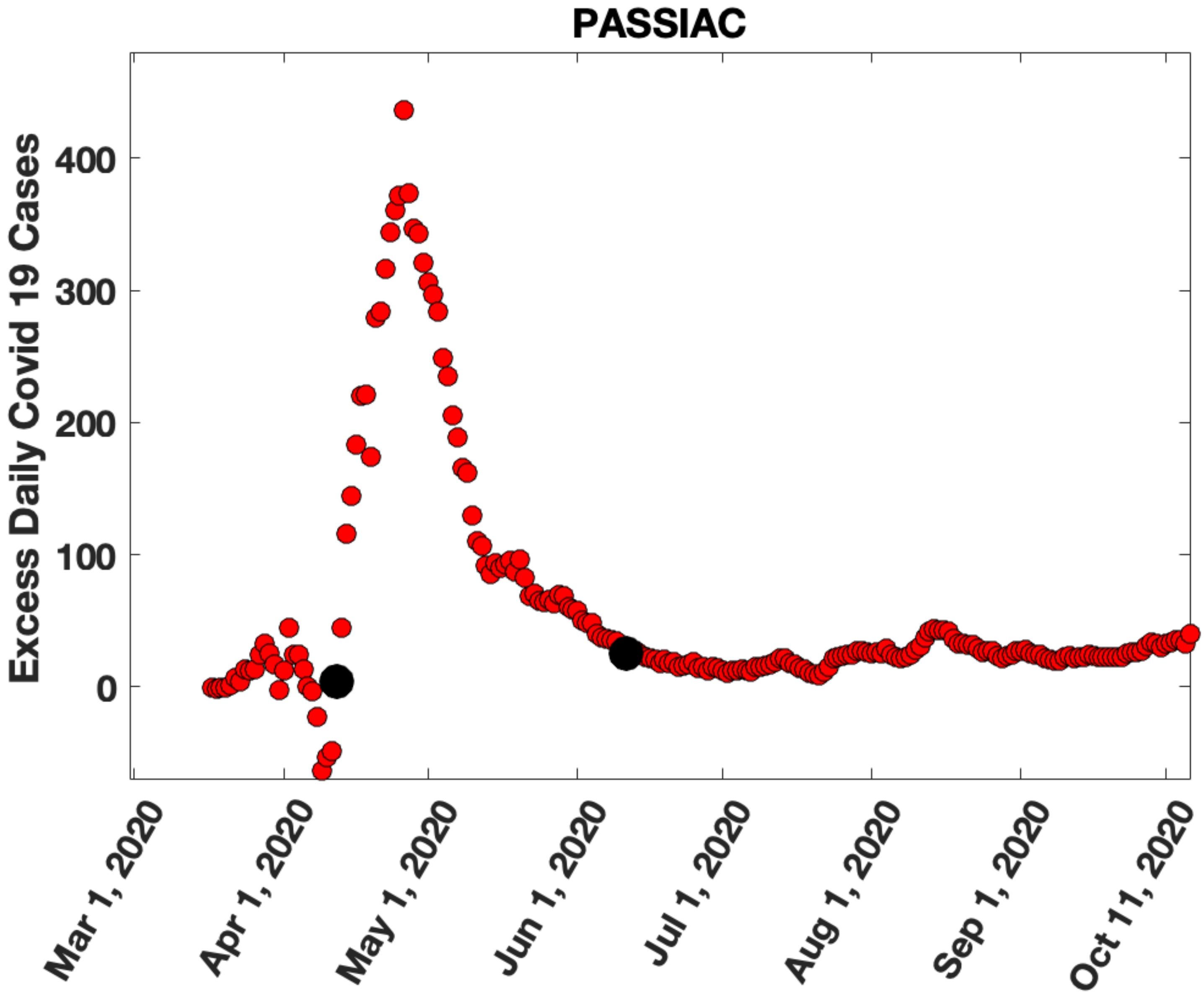
PASSIAC
Excess Daily Covid 19 Cases
400
300
200
100
0
Mar 1, 2020
Apr 1, 2020
May 1, 2020
Jun 1, 2020
Jul 1, 2020
Aug 1, 2020
Sep 1, 2020
Oct 11, 2020

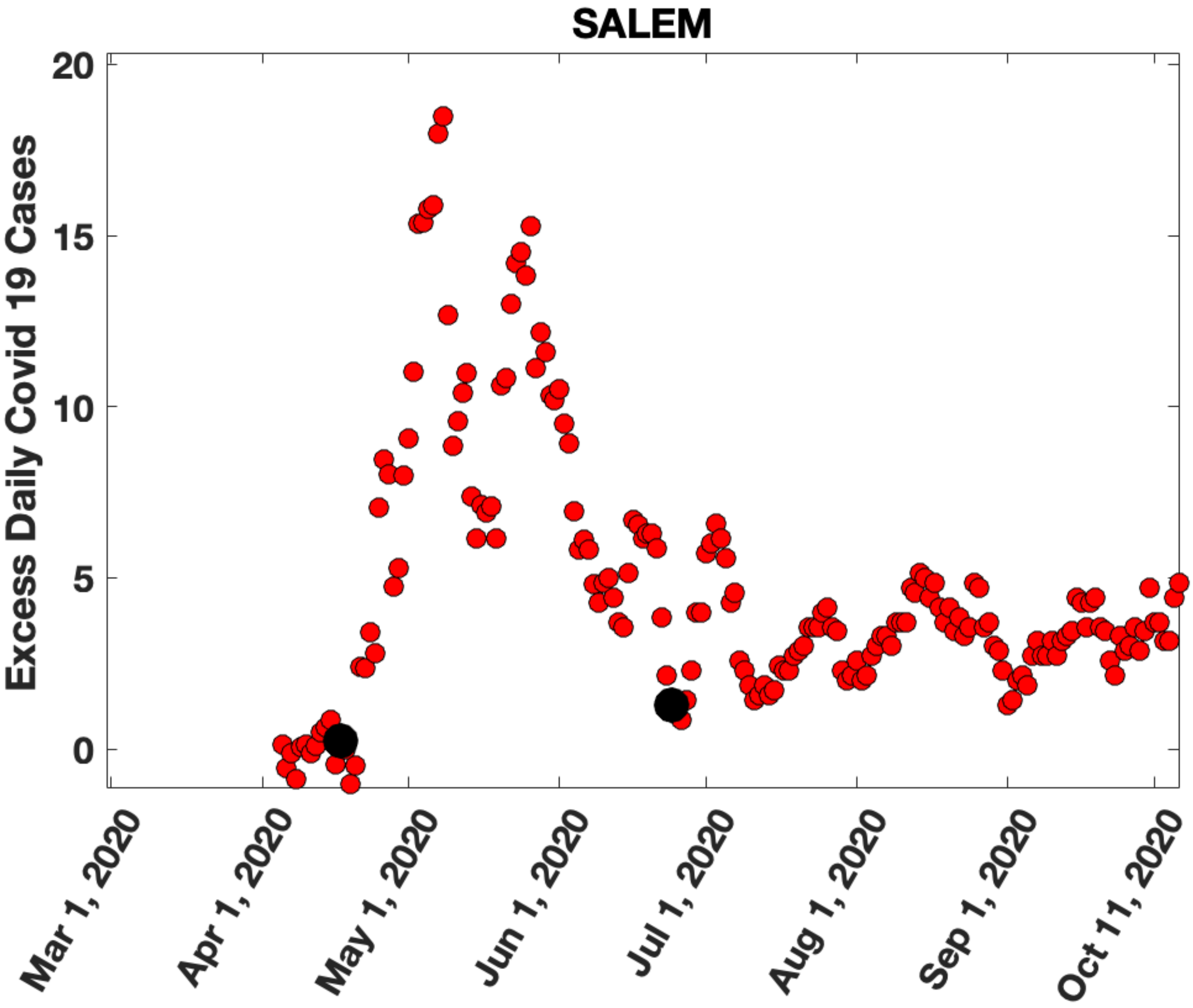
SALEM
Excess Daily Covid 19 Cases
20
15
10
5
0
Mar 1, 2020
Apr 1, 2020
May 1, 2020
Jun 1, 2020
Jul 1, 2020
Aug 1, 2020
Sep 1, 2020
Oct 11, 2020

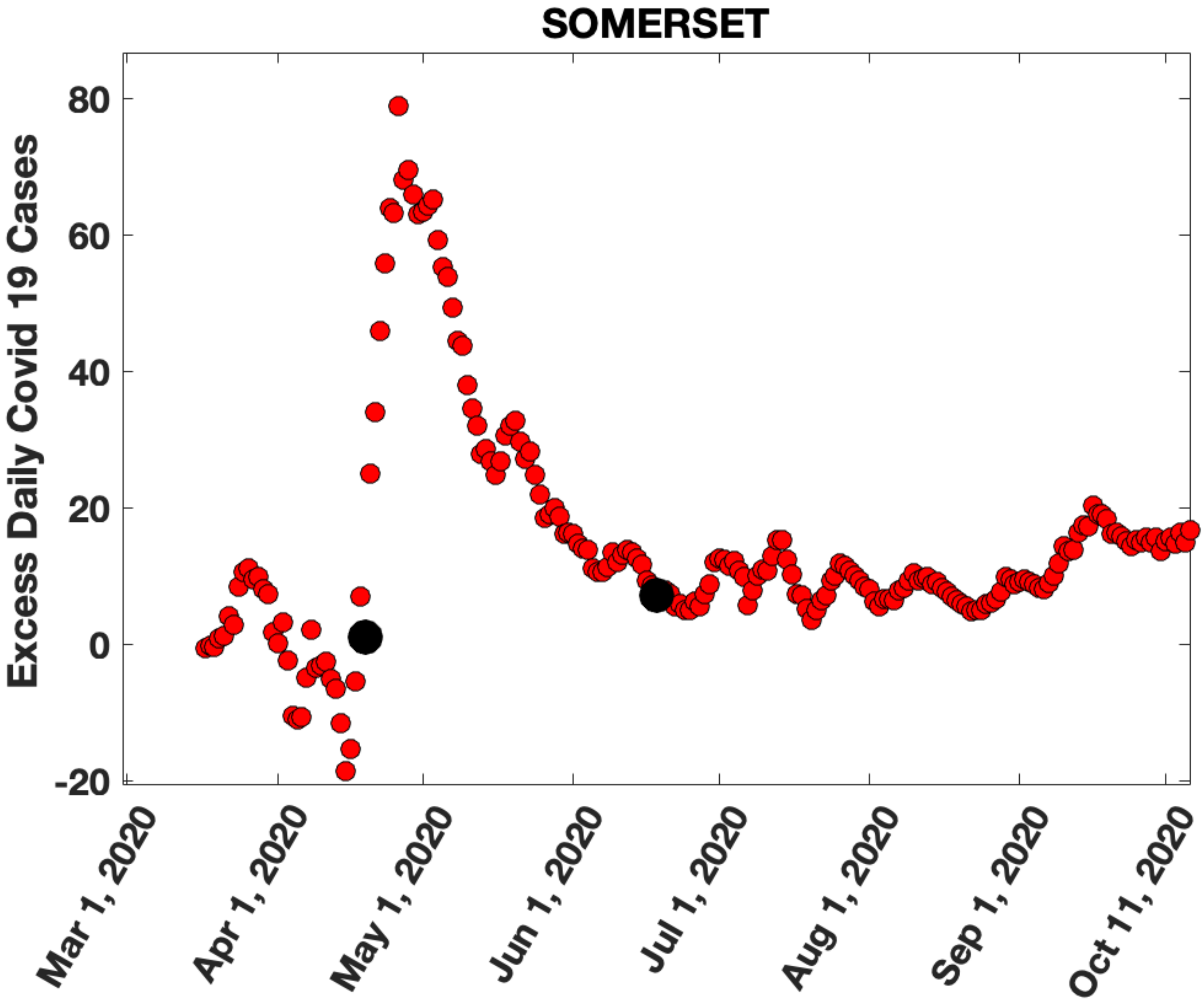
SOMERSET
Excess Daily Covid 19 Cases
80
60
40
20
0
-20
Mar 1, 2020
Apr 1, 2020
May 1, 2020
Jun 1, 2020
Jul 1, 2020
Aug 1, 2020
Sep 1, 2020
Oct 11, 2020

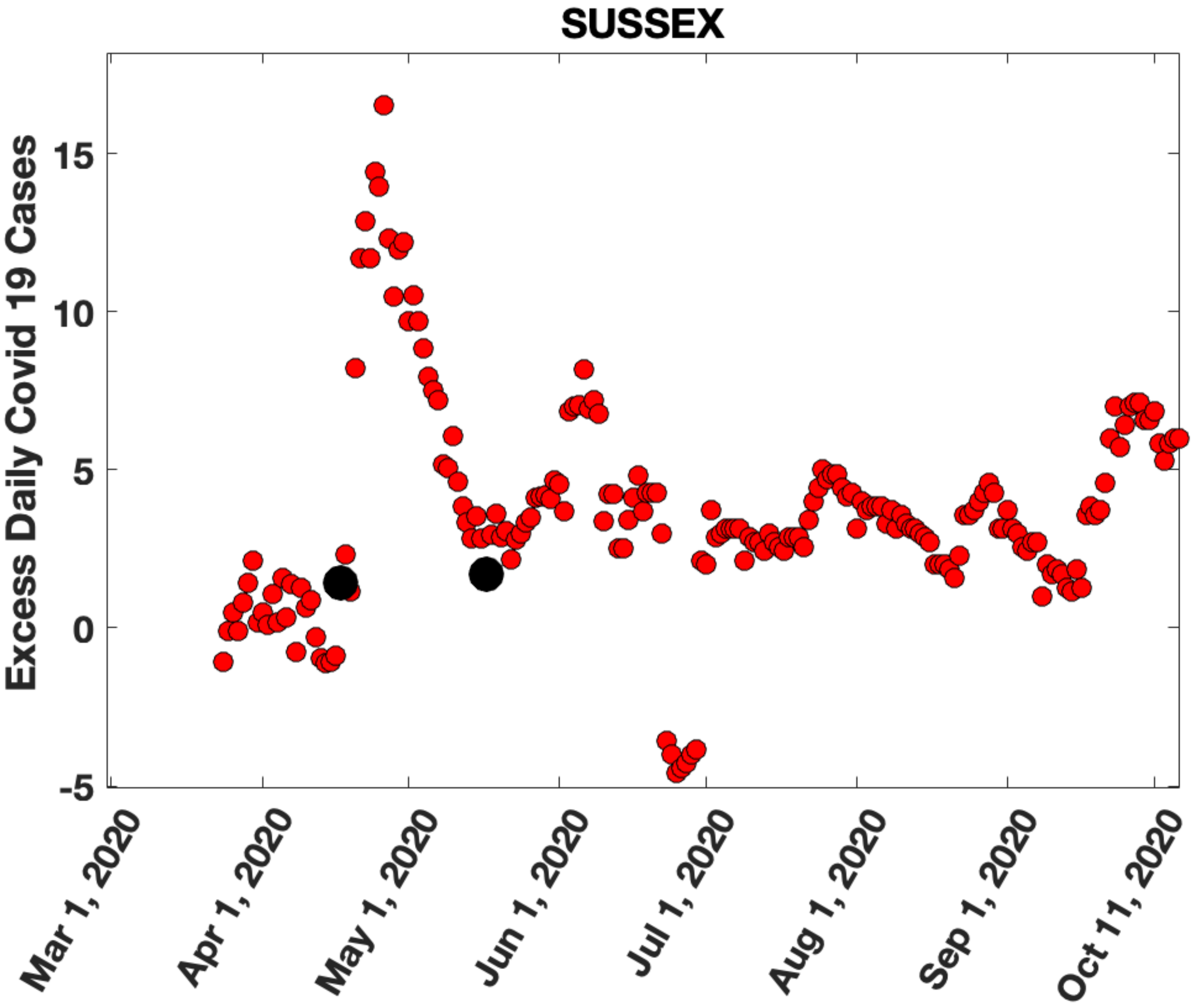

SUSSEX
Excess Daily Covid 19 Cases
15
10
5
0
-5
Mar 1, 2020
Apr 1, 2020
May 1, 2020
Jun 1, 2020
Jul 1, 2020
Aug 1, 2020
Sep 1, 2020
Oct 11, 2020

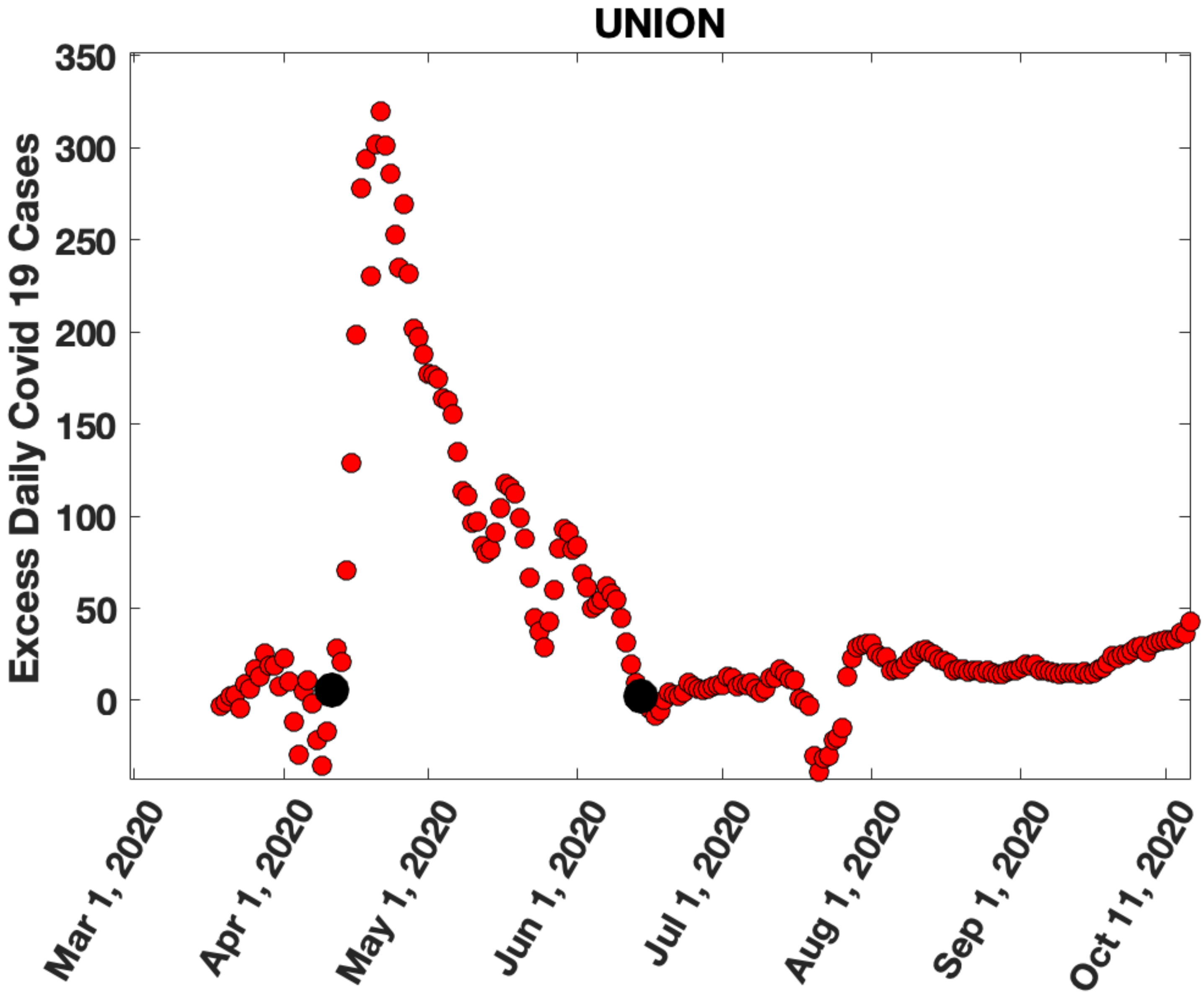
UNION
Excess Daily Covid 19 Cases
350
300
250
200
150
100
50
0
Mar 1, 2020
Apr 1, 2020
May 1, 2020
Jun 1, 2020
Jul 1, 2020
Aug 1, 2020
Sep 1, 2020
Oct 11, 2020

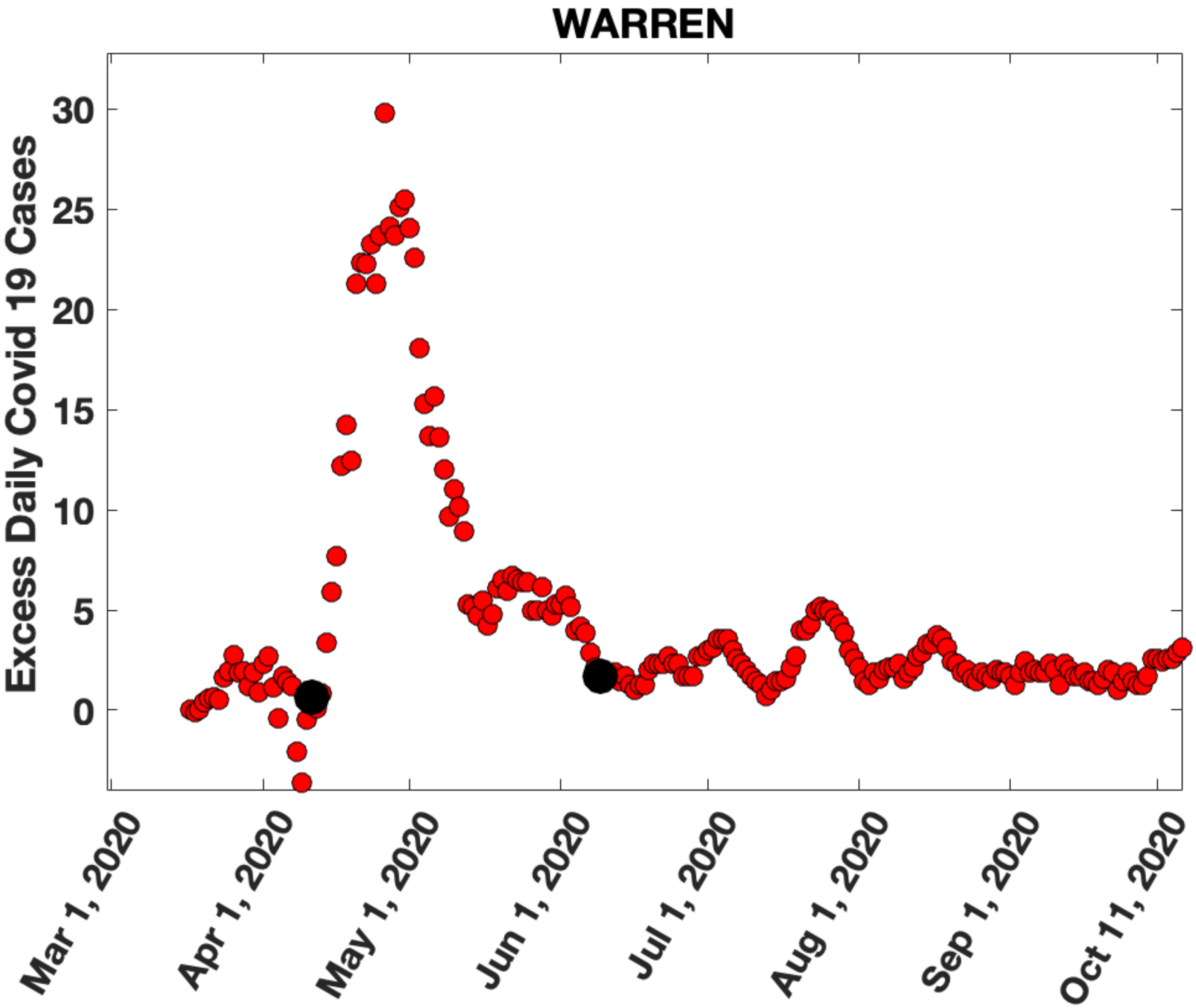

WARREN
Excess Daily Covid 19 Cases
30
25
20
15
10
5
0
Mar 1, 2020
Apr 1, 2020
May 1, 2020
Jun 1, 2020
Jul 1, 2020
Aug 1, 2020
Sep 1, 2020
Oct 11, 2020
