## Supplementary Figure 10 for "Migration of households from New York City and the Second Peak in Covid-19 cases in New Jersey, Connecticut and New York Counties"

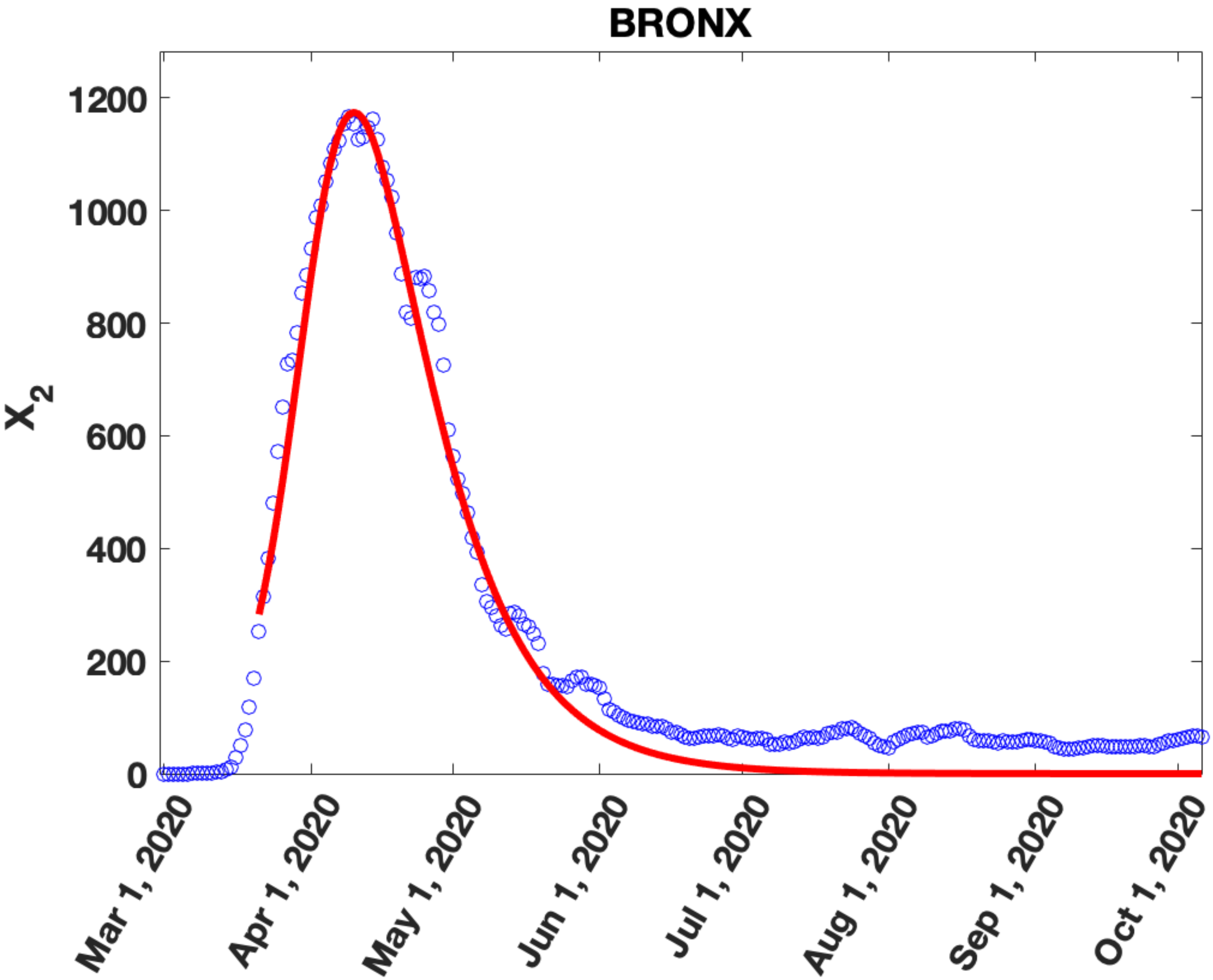
BRONX
X_2
0
200
400
600
800
1000
1200
Mar 1, 2020
Apr 1, 2020
May 1, 2020
Jun 1, 2020
Jul 1, 2020
Aug 1, 2020
Sep 1, 2020
Oct 1, 2020

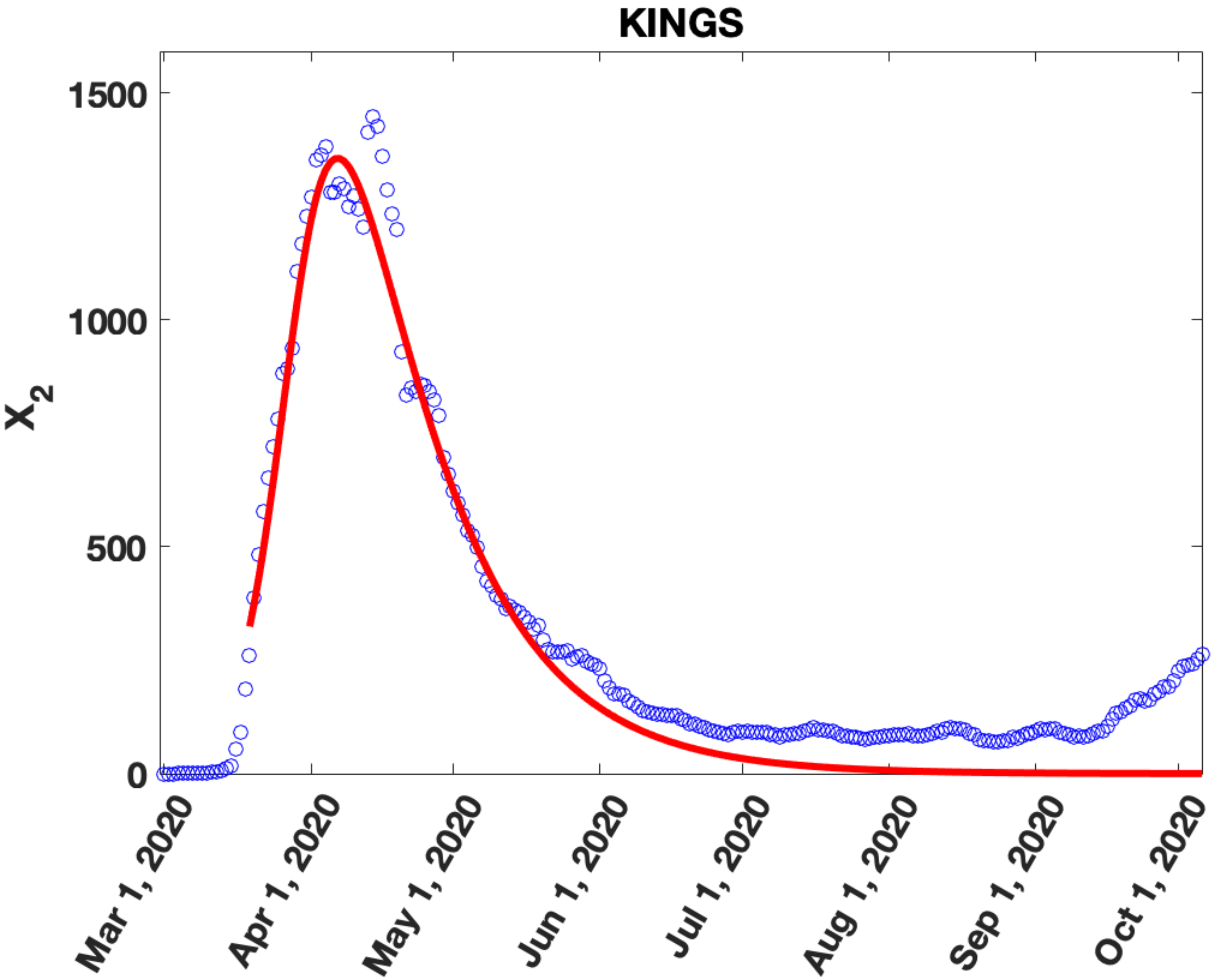

KINGS
1500
1000
500
0
$X_2$
Mar 1, 2020
Apr 1, 2020
May 1, 2020
Jun 1, 2020
Jul 1, 2020
Aug 1, 2020
Sep 1, 2020
Oct 1, 2020

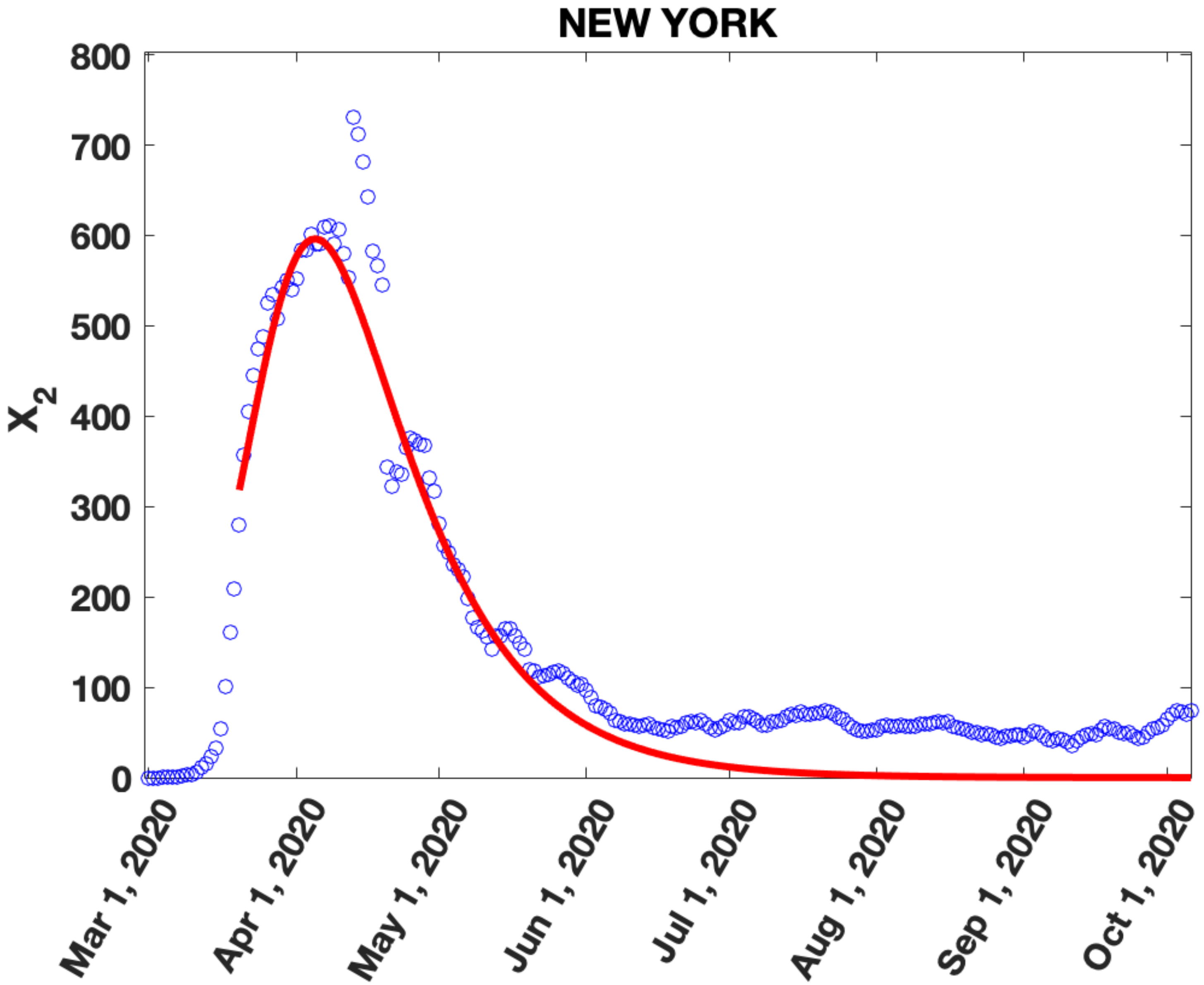
NEW YORK
800
700
600
500
400
300
200
100
0
X2
Mar 1, 2020
Apr 1, 2020
May 1, 2020
Jun 1, 2020
Jul 1, 2020
Aug 1, 2020
Sep 1, 2020
Oct 1, 2020

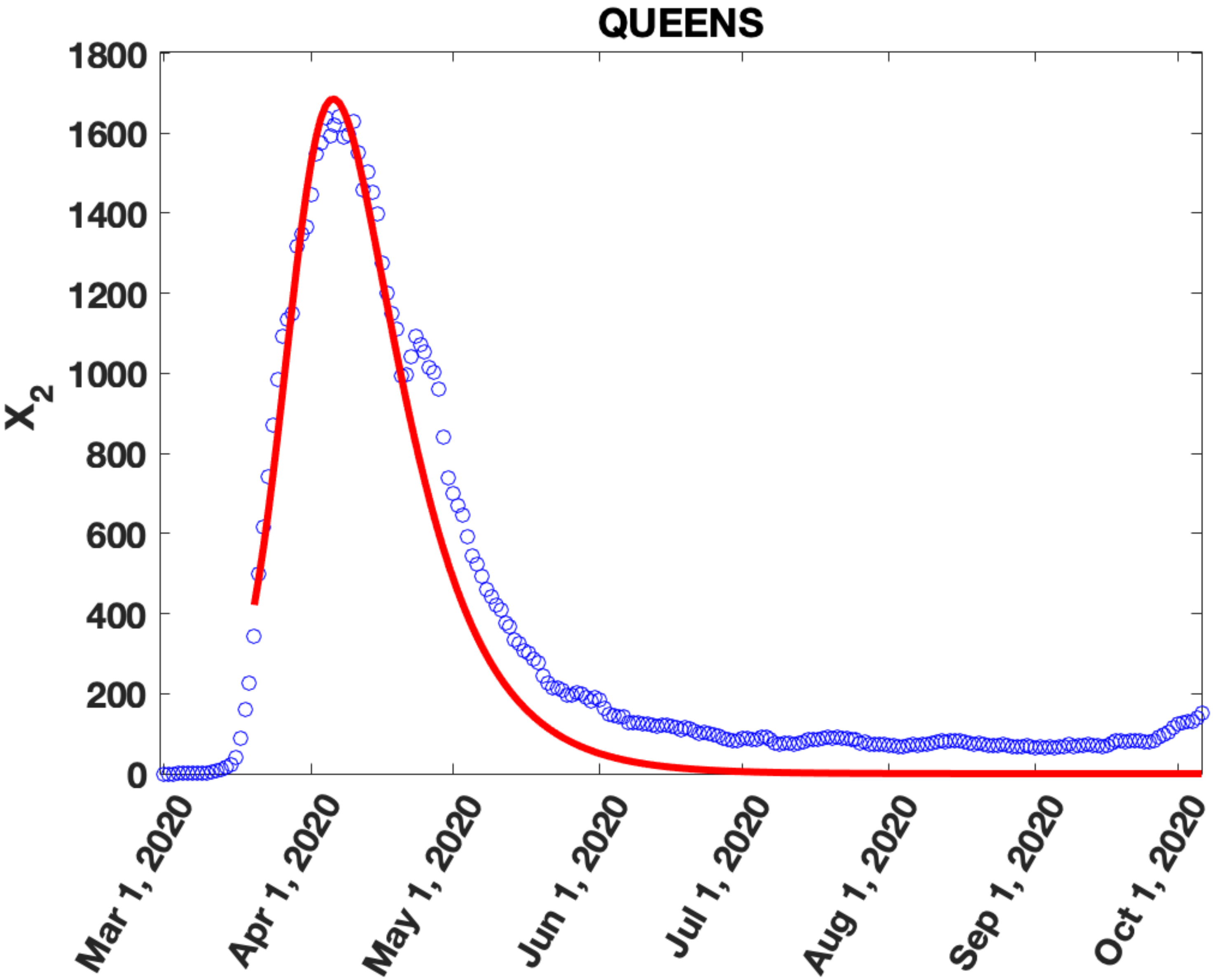

QUEENS
X_2
1800
1600
1400
1200
1000
800
600
400
200
0
Mar 1, 2020
Apr 1, 2020
May 1, 2020
Jun 1, 2020
Jul 1, 2020
Aug 1, 2020
Sep 1, 2020
Oct 1, 2020

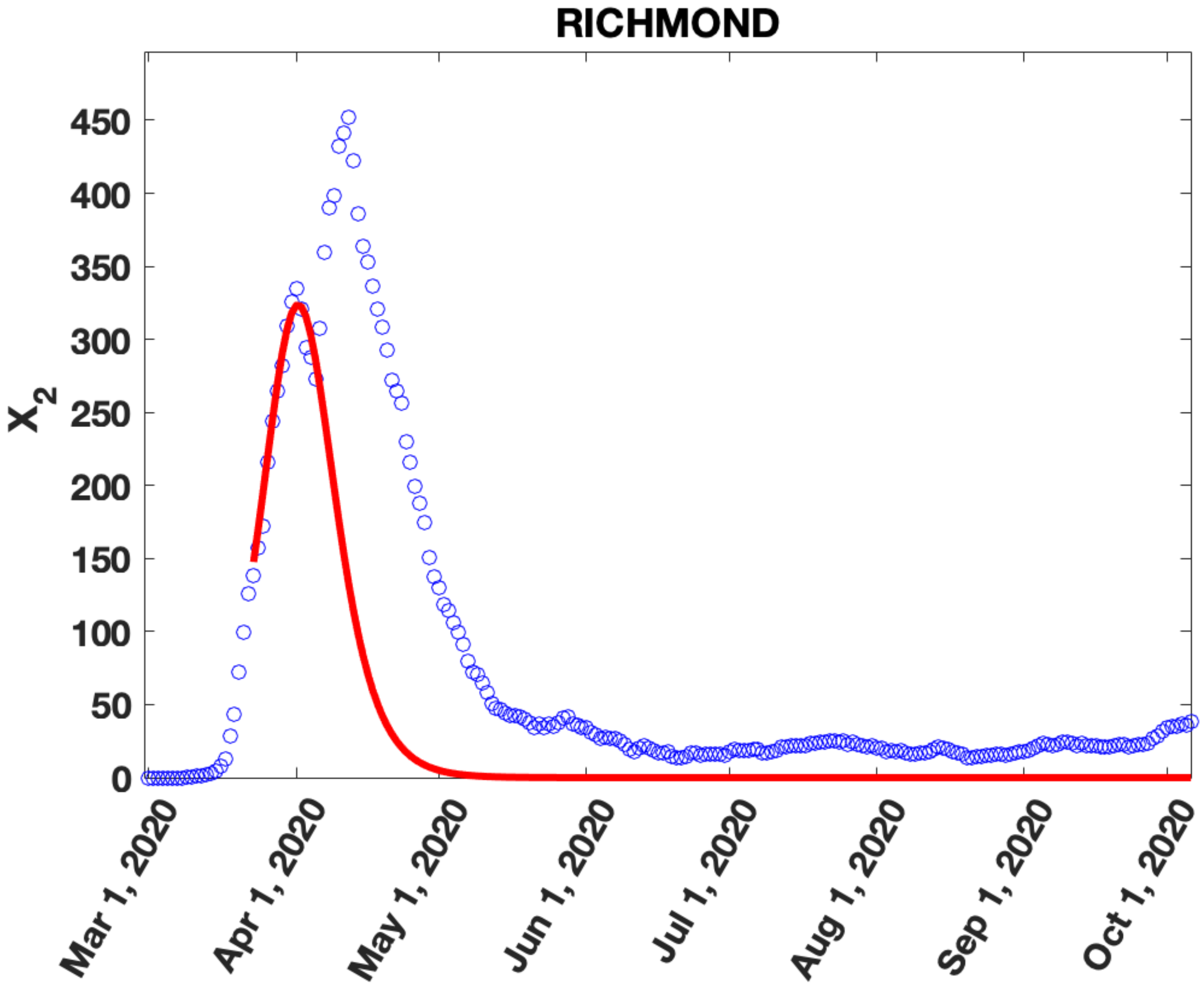
RICHMOND
X2
450
400
350
300
250
200
150
100
50
0
Mar 1, 2020
Apr 1, 2020
May 1, 2020
Jun 1, 2020
Jul 1, 2020
Aug 1, 2020
Sep 1, 2020
Oct 1, 2020

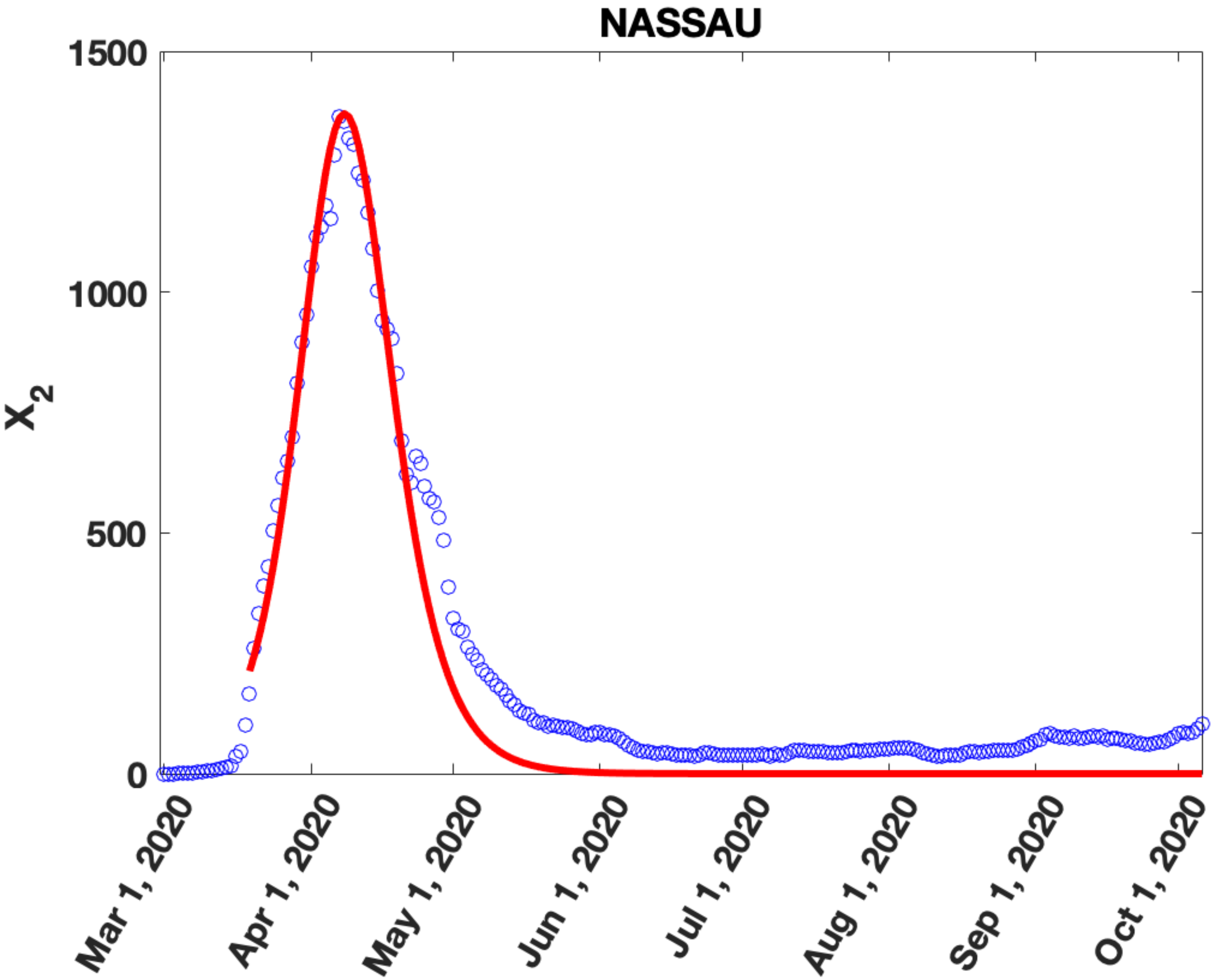
NASSAU
1500
1000
500
0
$X_2$
Mar 1, 2020
Apr 1, 2020
May 1, 2020
Jun 1, 2020
Jul 1, 2020
Aug 1, 2020
Sep 1, 2020
Oct 1, 2020

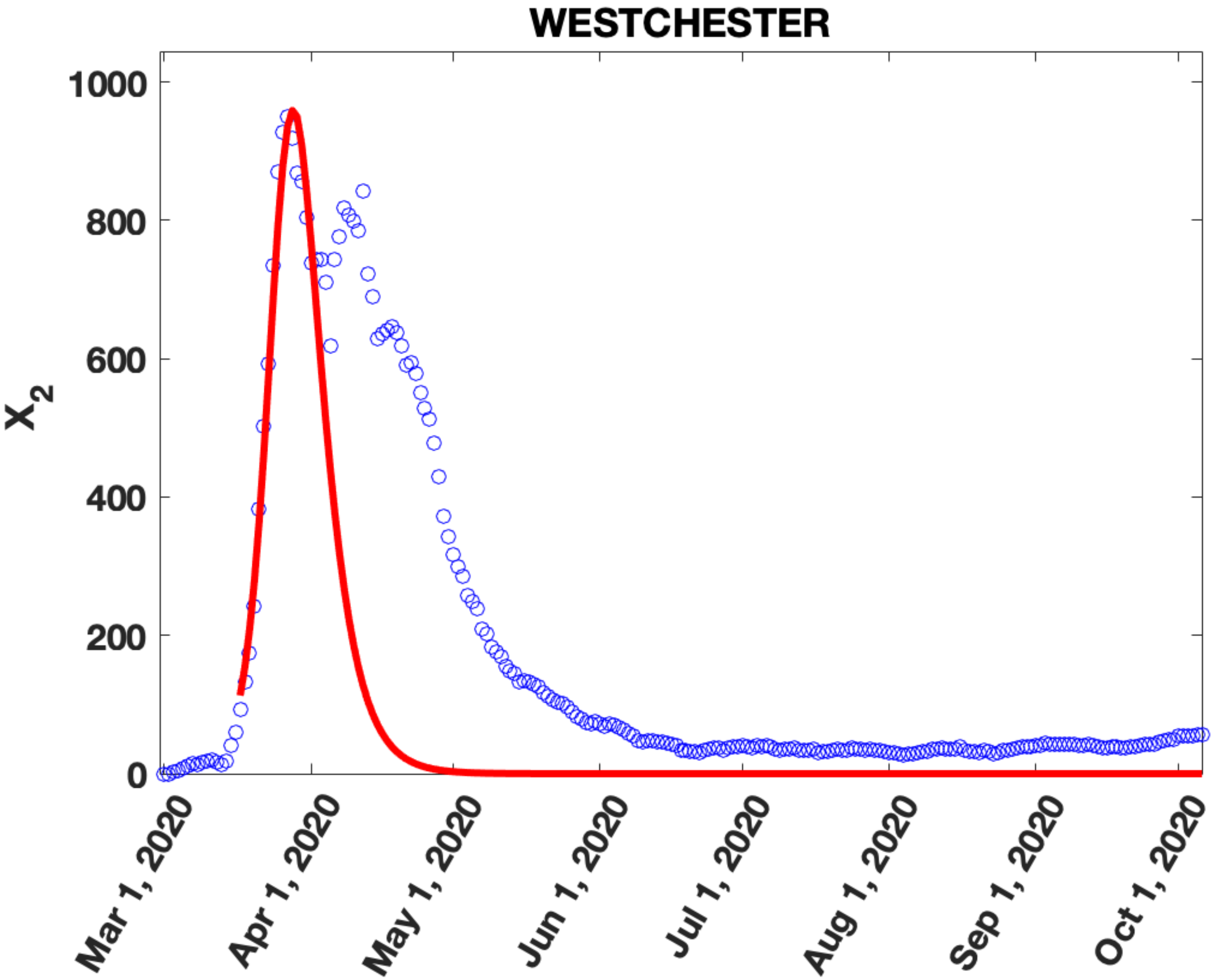
WESTCHESTER
1000
800
600
400
200
0
$X_2$
Mar 1, 2020
Apr 1, 2020
May 1, 2020
Jun 1, 2020
Jul 1, 2020
Aug 1, 2020
Sep 1, 2020
Oct 1, 2020

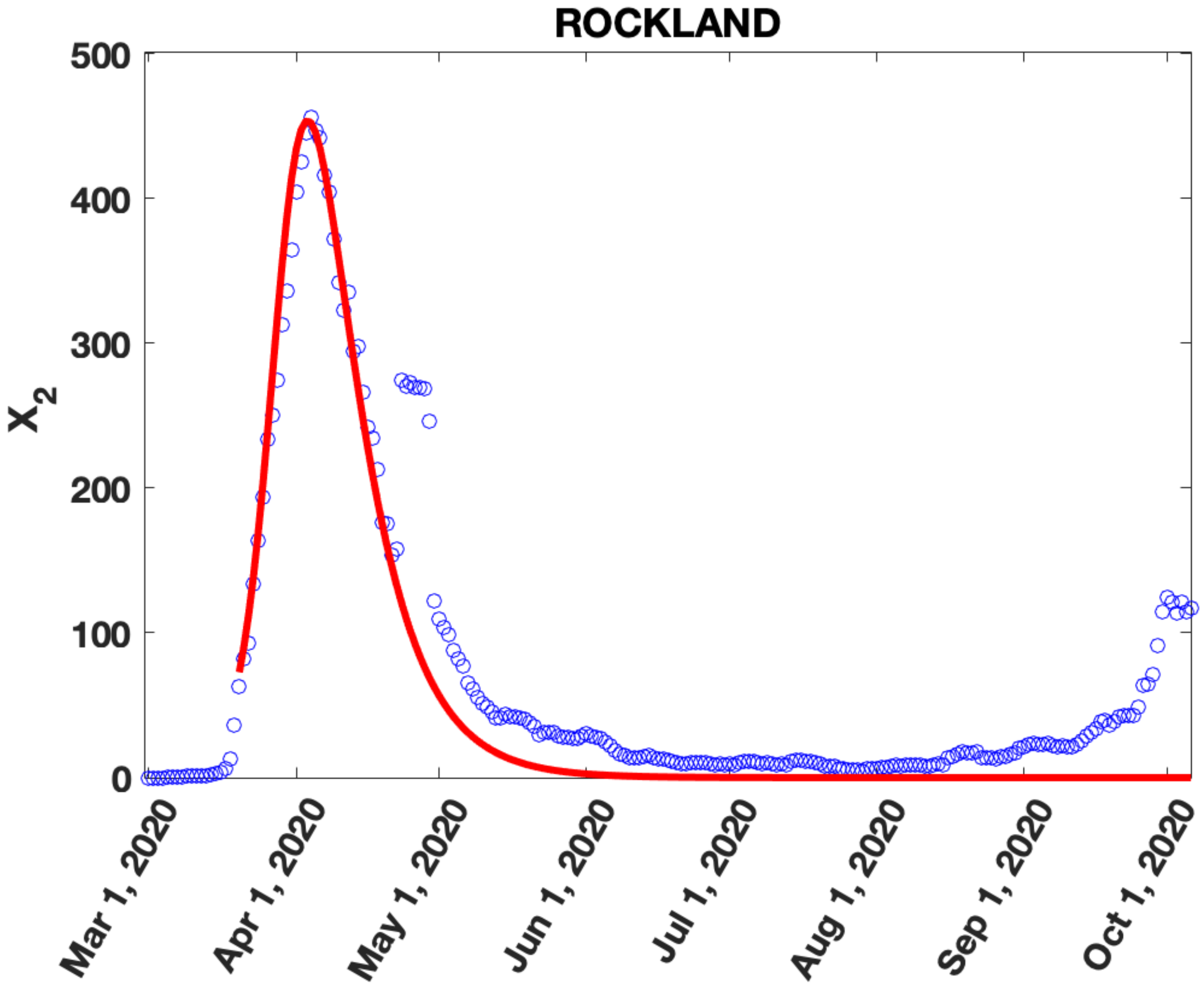
ROCKLAND
500
400
300
200
100
0
$X_2$
Mar 1, 2020
Apr 1, 2020
May 1, 2020
Jun 1, 2020
Jul 1, 2020
Aug 1, 2020
Sep 1, 2020
Oct 1, 2020

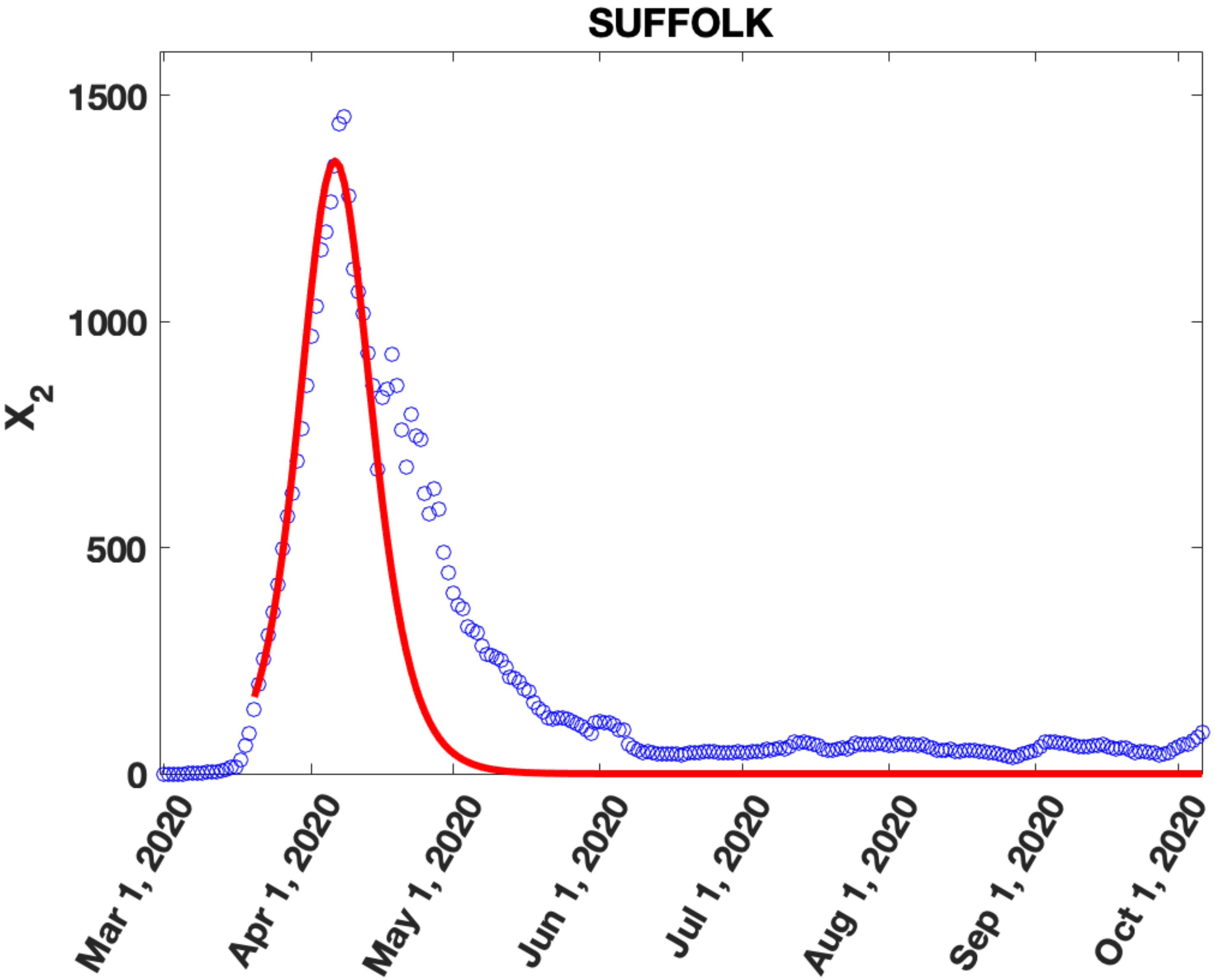

SUFFOLK
X_2
1500
1000
500
0
Mar 1, 2020
Apr 1, 2020
May 1, 2020
Jun 1, 2020
Jul 1, 2020
Aug 1, 2020
Sep 1, 2020
Oct 1, 2020

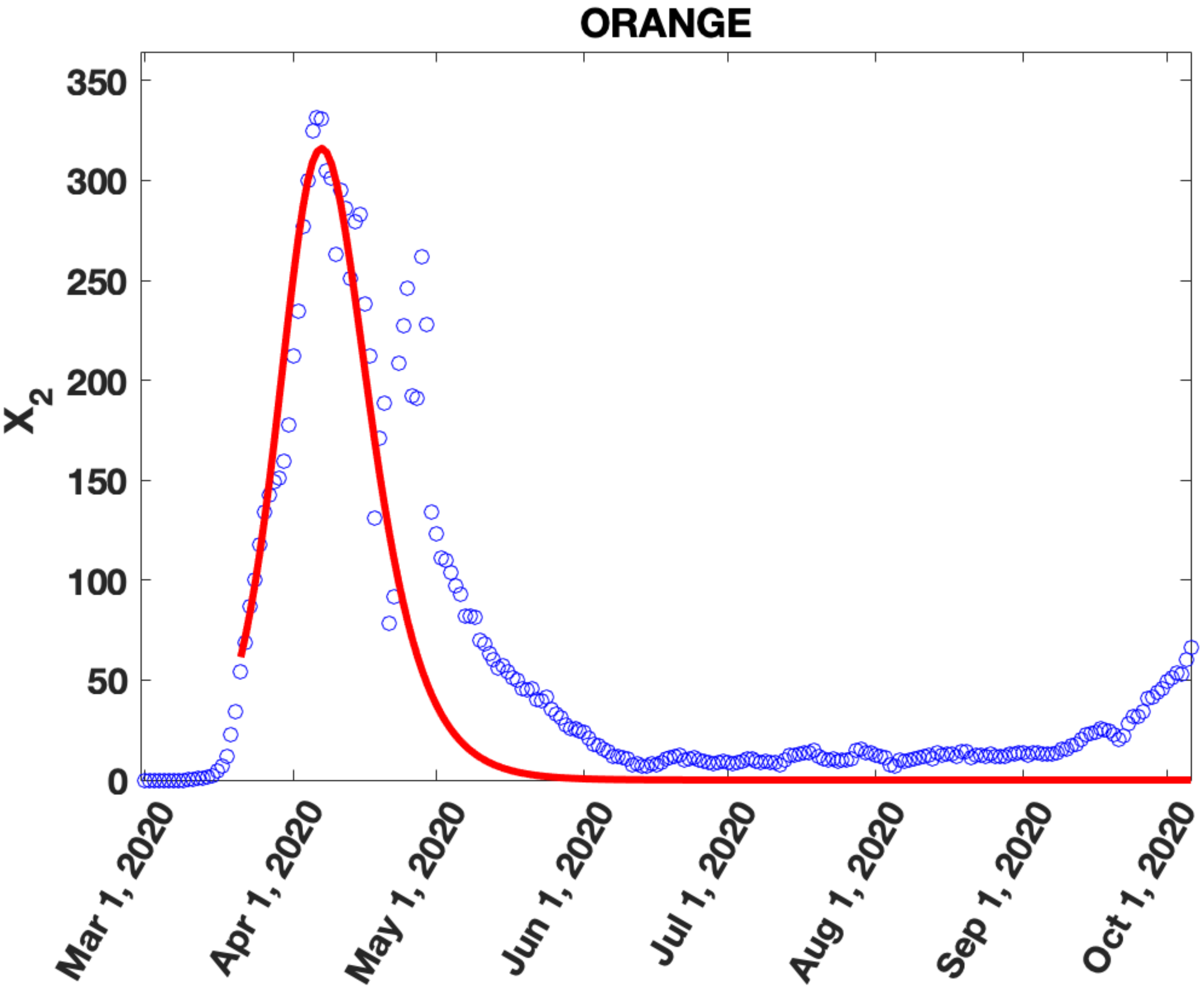

ORANGE
350
300
250
200
150
100
50
0
$X_2$
Mar 1, 2020
Apr 1, 2020
May 1, 2020
Jun 1, 2020
Jul 1, 2020
Aug 1, 2020
Sep 1, 2020
Oct 1, 2020

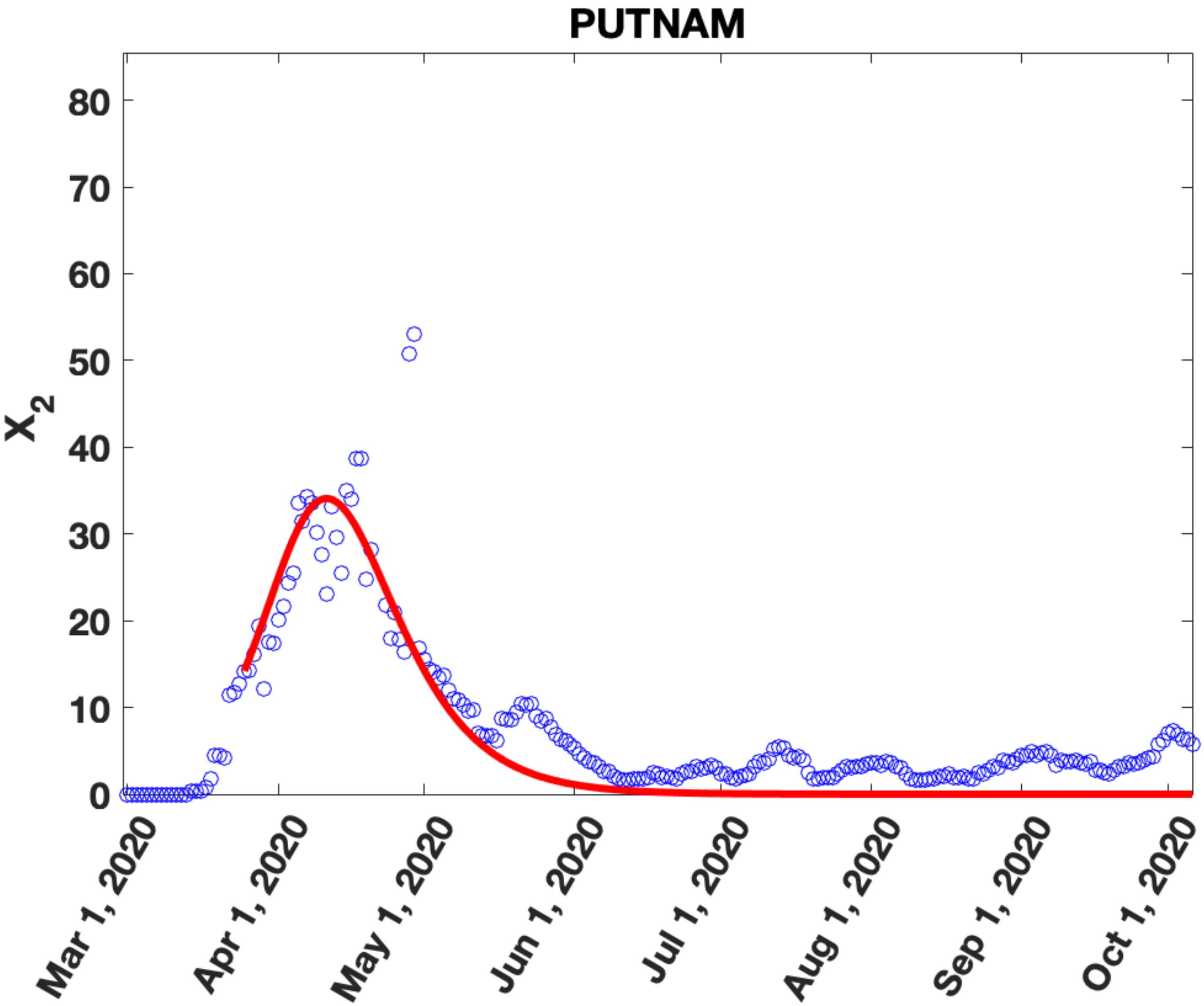

PUTNAM
80
70
60
50
40
30
20
10
0
$X_2$
Mar 1, 2020
Apr 1, 2020
May 1, 2020
Jun 1, 2020
Jul 1, 2020
Aug 1, 2020
Sep 1, 2020
Oct 1, 2020

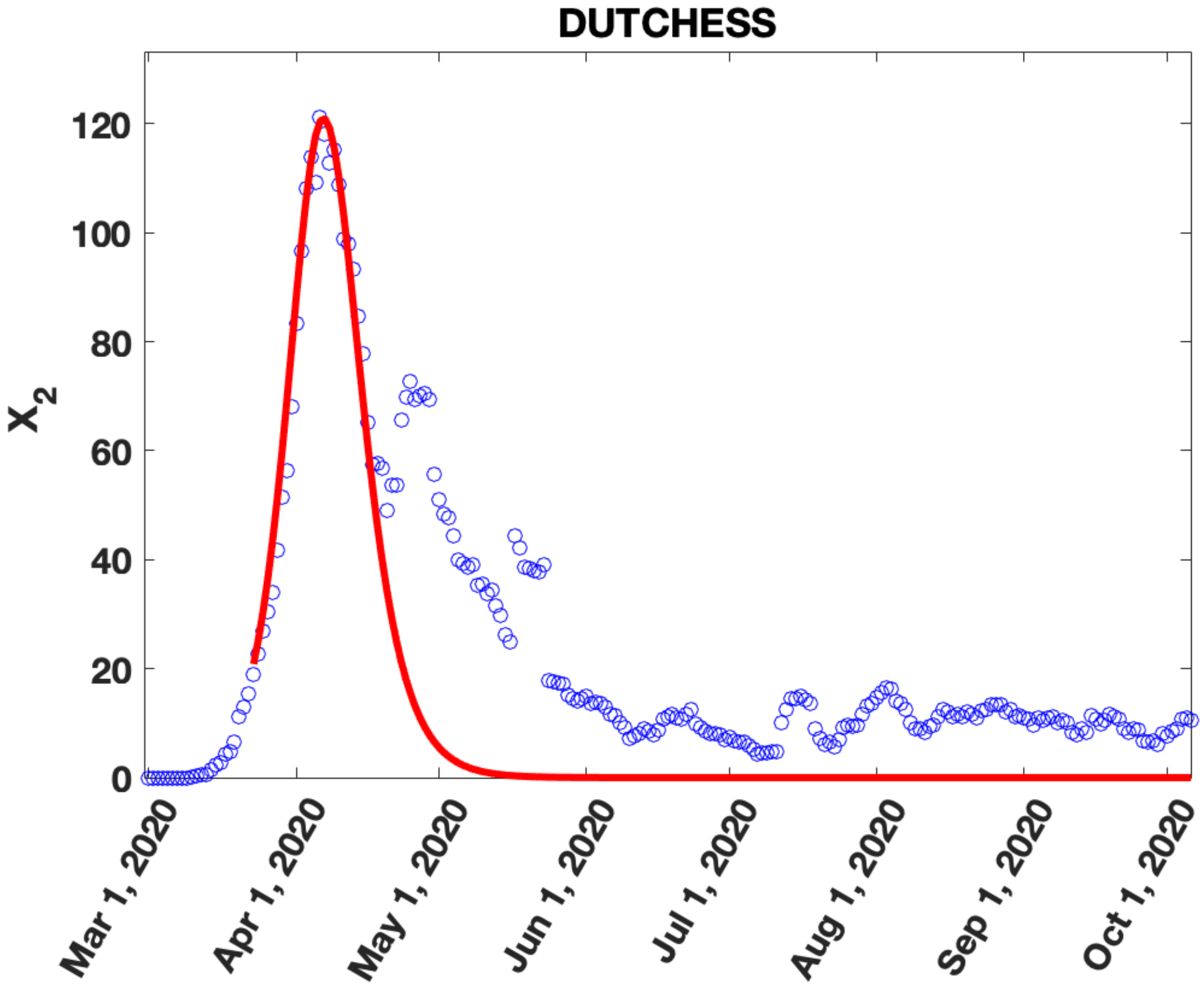

DUTCHESS
$X_2$
0
20
40
60
80
100
120
Mar 1, 2020
Apr 1, 2020
May 1, 2020
Jun 1, 2020
Jul 1, 2020
Aug 1, 2020
Sep 1, 2020
Oct 1, 2020

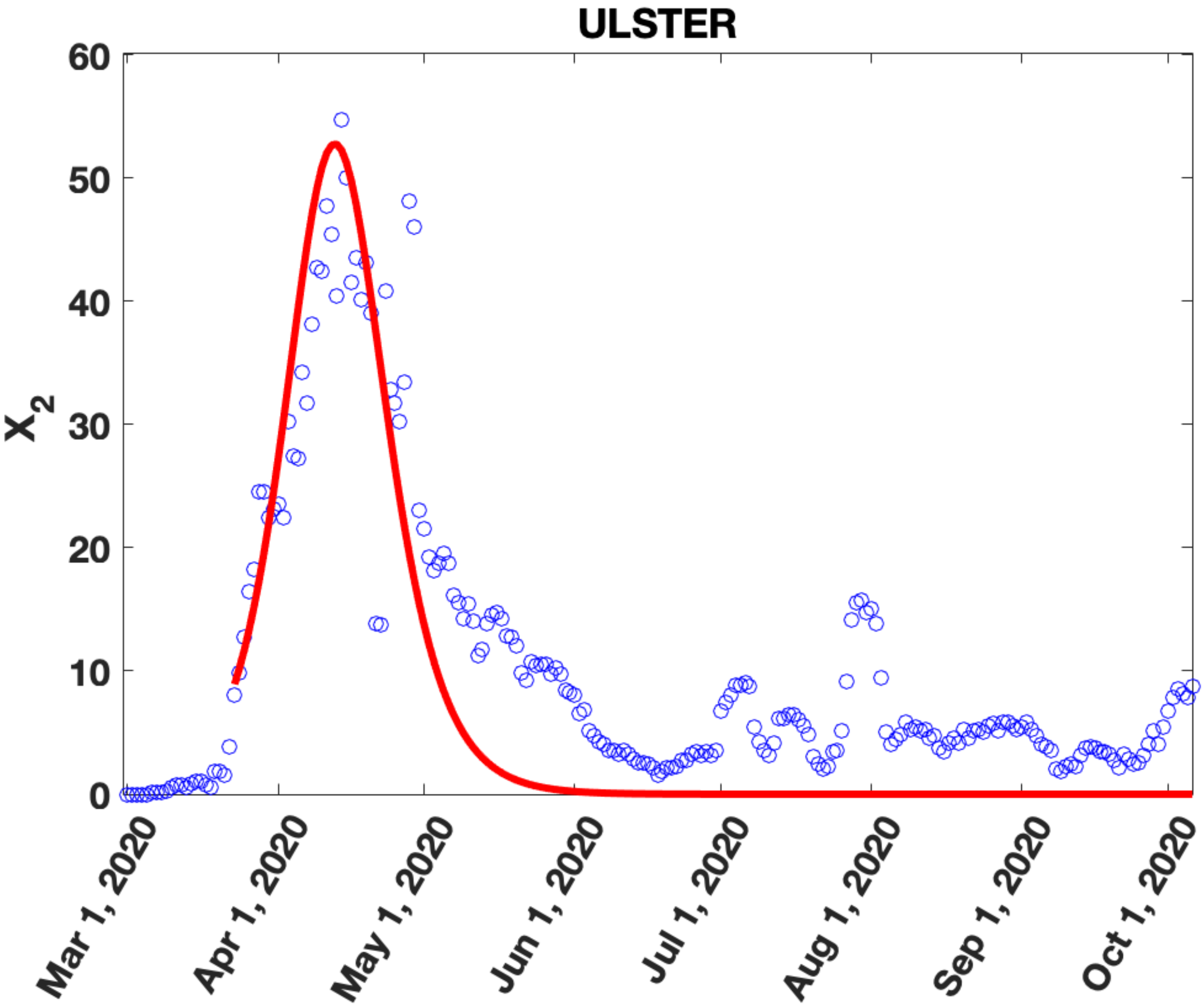

ULSTER
X2
0
10
20
30
40
50
60
Mar 1, 2020
Apr 1, 2020
May 1, 2020
Jun 1, 2020
Jul 1, 2020
Aug 1, 2020
Sep 1, 2020
Oct 1, 2020

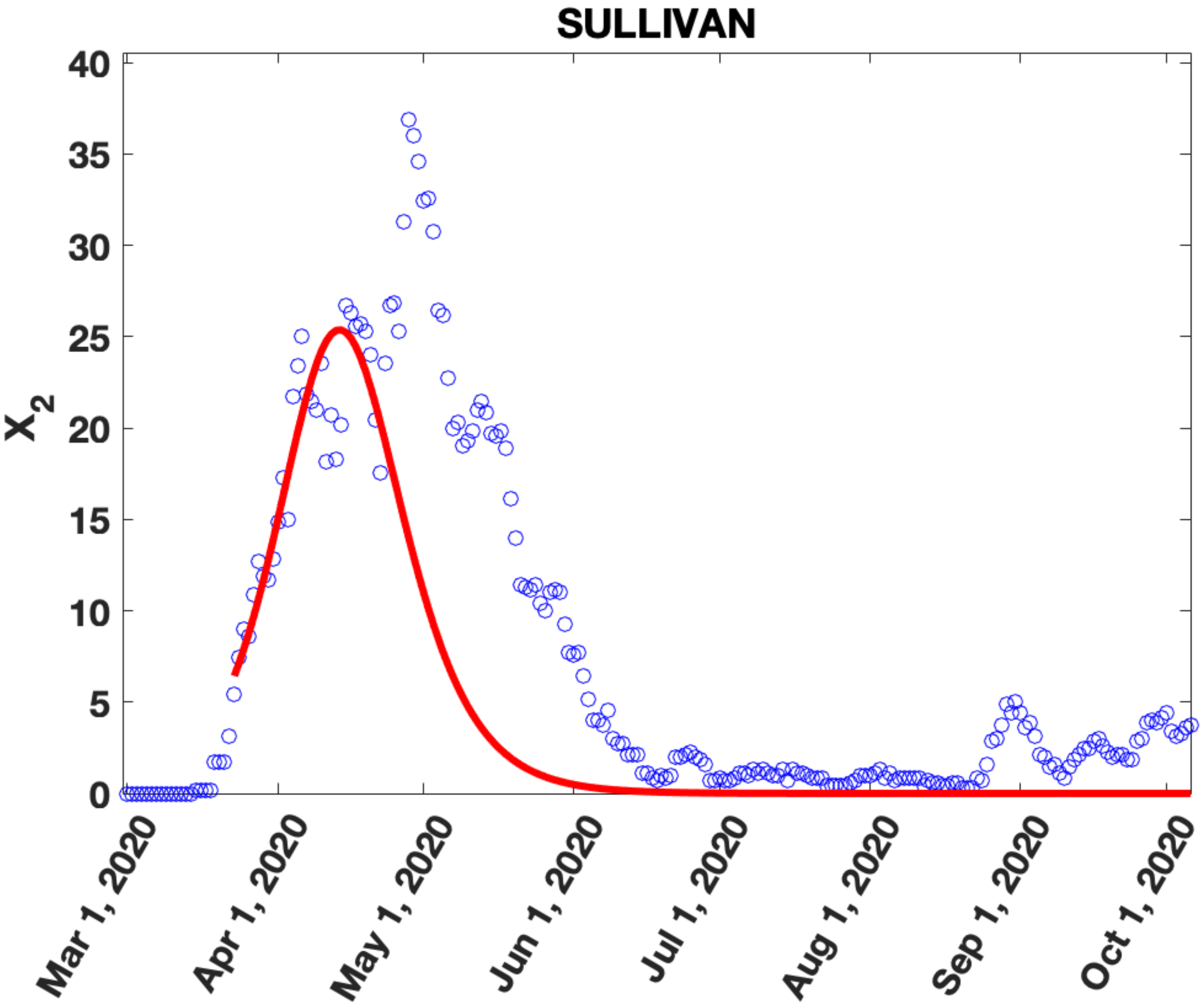

SULLIVAN
$X_2$
40
35
30
25
20
15
10
5
0
Mar 1, 2020
Apr 1, 2020
May 1, 2020
Jun 1, 2020
Jul 1, 2020
Aug 1, 2020
Sep 1, 2020
Oct 1, 2020
