## Supplementary Figure 11 for "Migration of households from New York City and the Second Peak in Covid-19 cases in New Jersey, Connecticut and New York Counties"

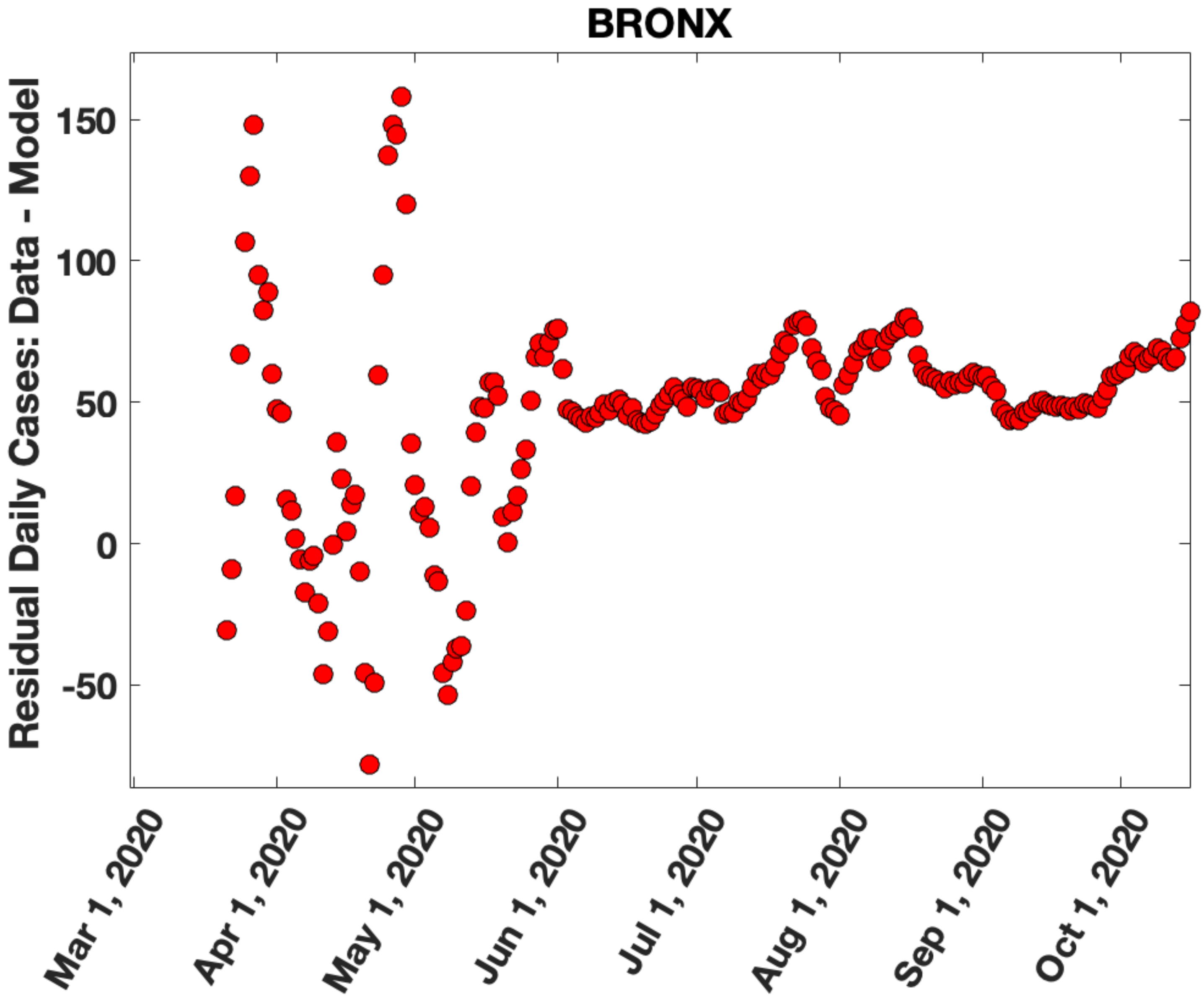

BRONX
Residual Daily Cases: Data - Model
150
100
50
0
-50
Mar 1, 2020
Apr 1, 2020
May 1, 2020
Jun 1, 2020
Jul 1, 2020
Aug 1, 2020
Sep 1, 2020
Oct 1, 2020

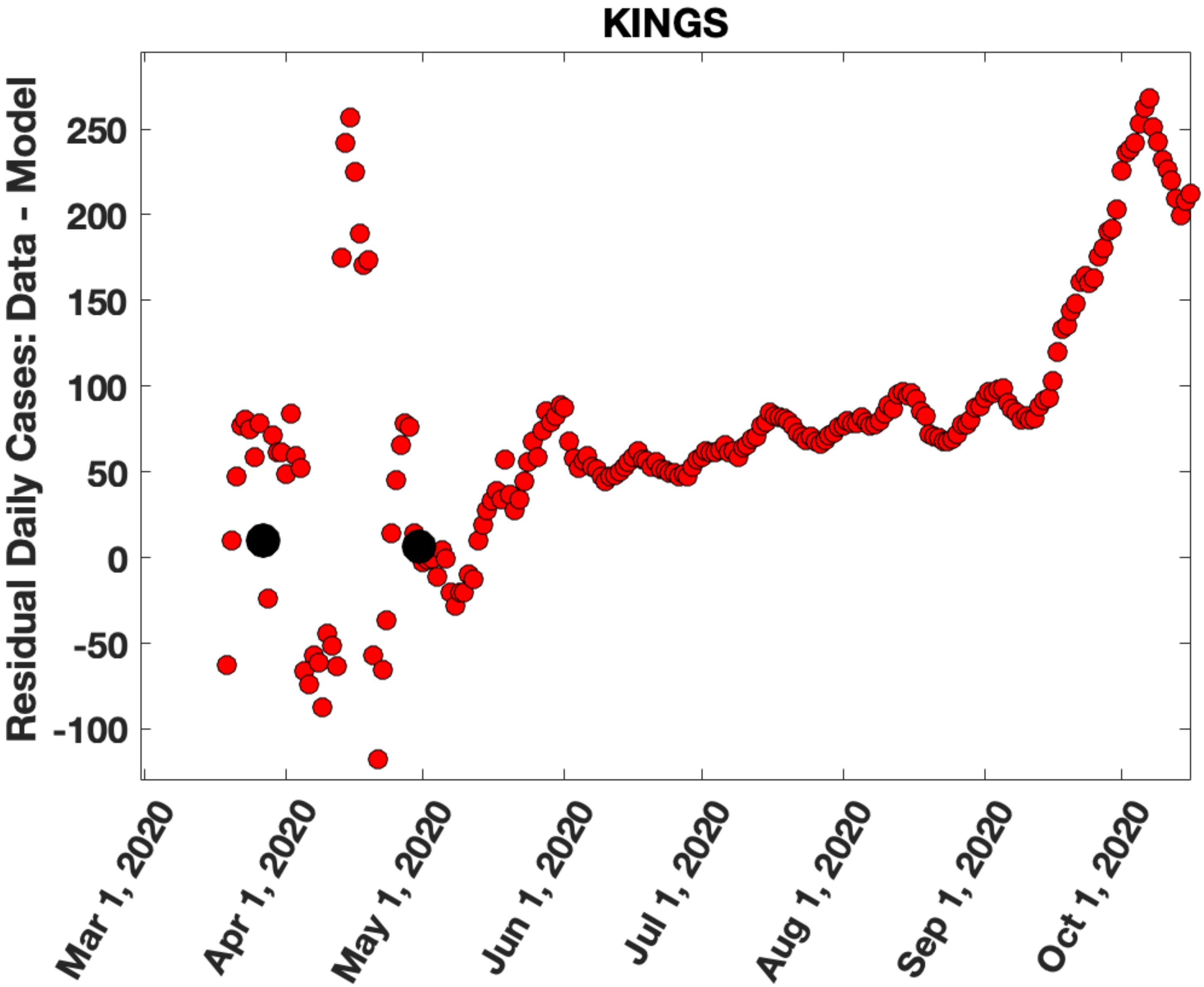
KINGS
Residual Daily Cases: Data - Model
250
200
150
100
50
0
-50
-100
Mar 1, 2020
Apr 1, 2020
May 1, 2020
Jun 1, 2020
Jul 1, 2020
Aug 1, 2020
Sep 1, 2020
Oct 1, 2020

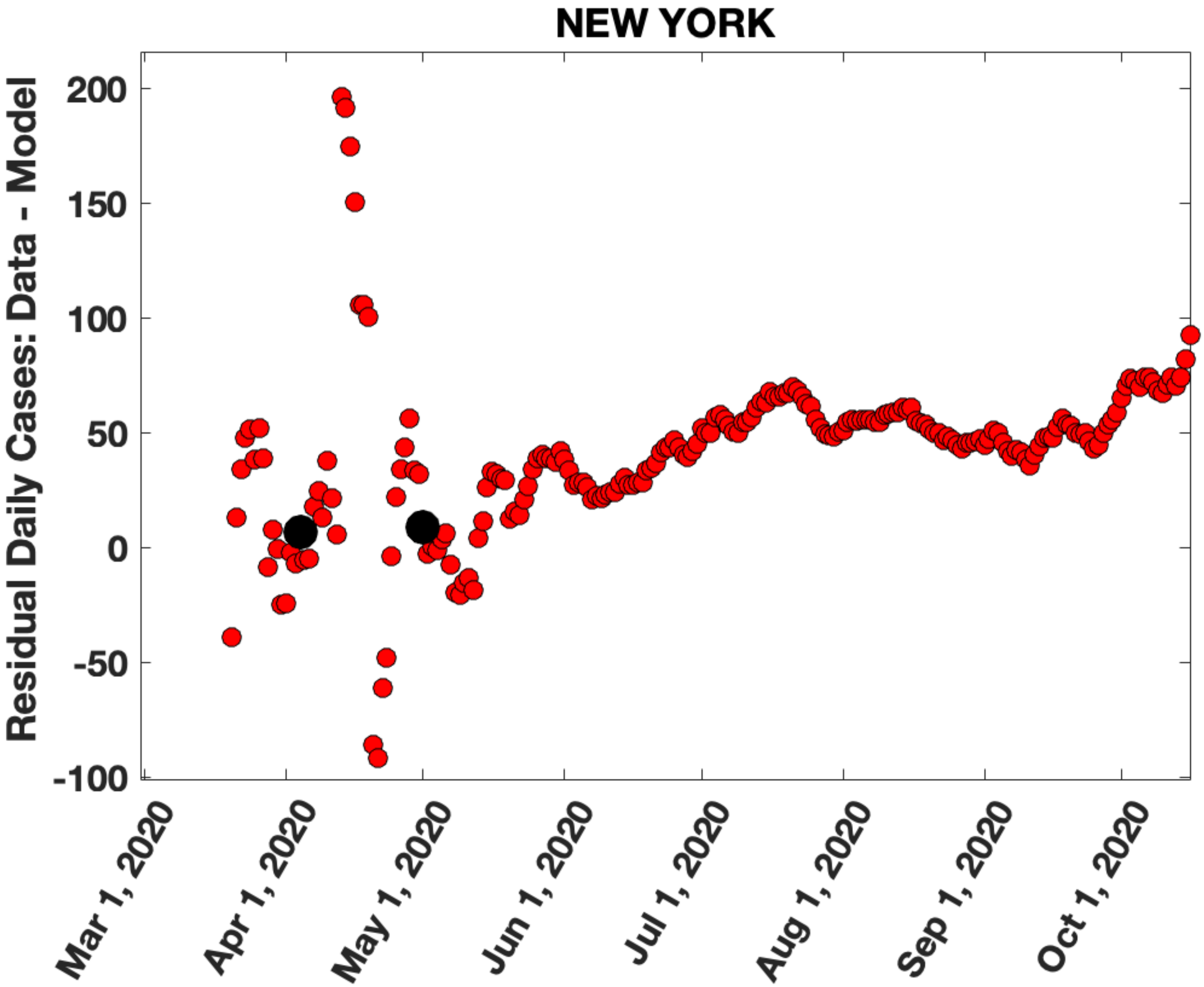
NEW YORK
Residual Daily Cases: Data - Model
200
150
100
50
0
-50
-100
Mar 1, 2020
Apr 1, 2020
May 1, 2020
Jun 1, 2020
Jul 1, 2020
Aug 1, 2020
Sep 1, 2020
Oct 1, 2020

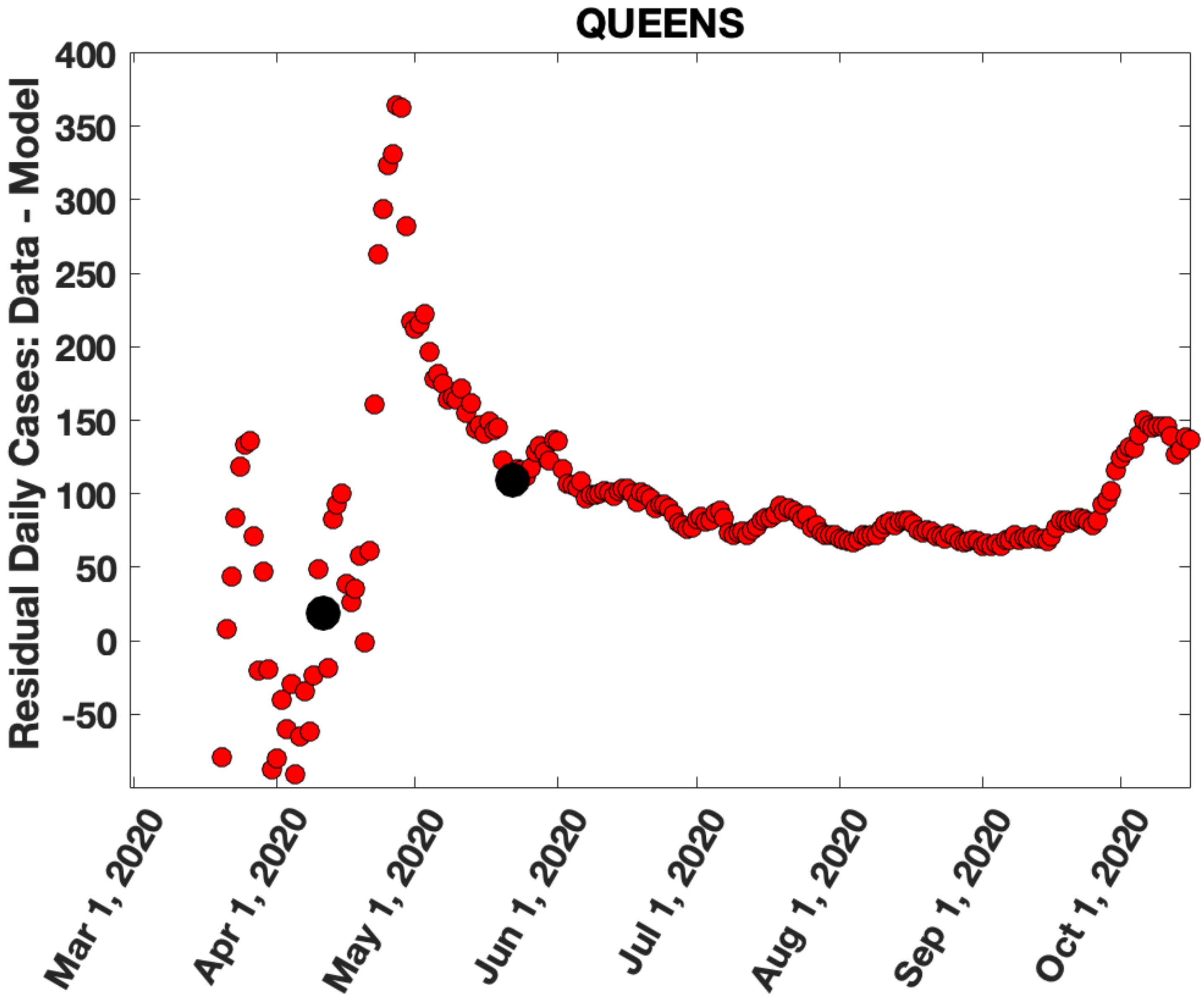
QUEENS
Residual Daily Cases: Data - Model
400
350
300
250
200
150
100
50
0
-50
Mar 1, 2020
Apr 1, 2020
May 1, 2020
Jun 1, 2020
Jul 1, 2020
Aug 1, 2020
Sep 1, 2020
Oct 1, 2020

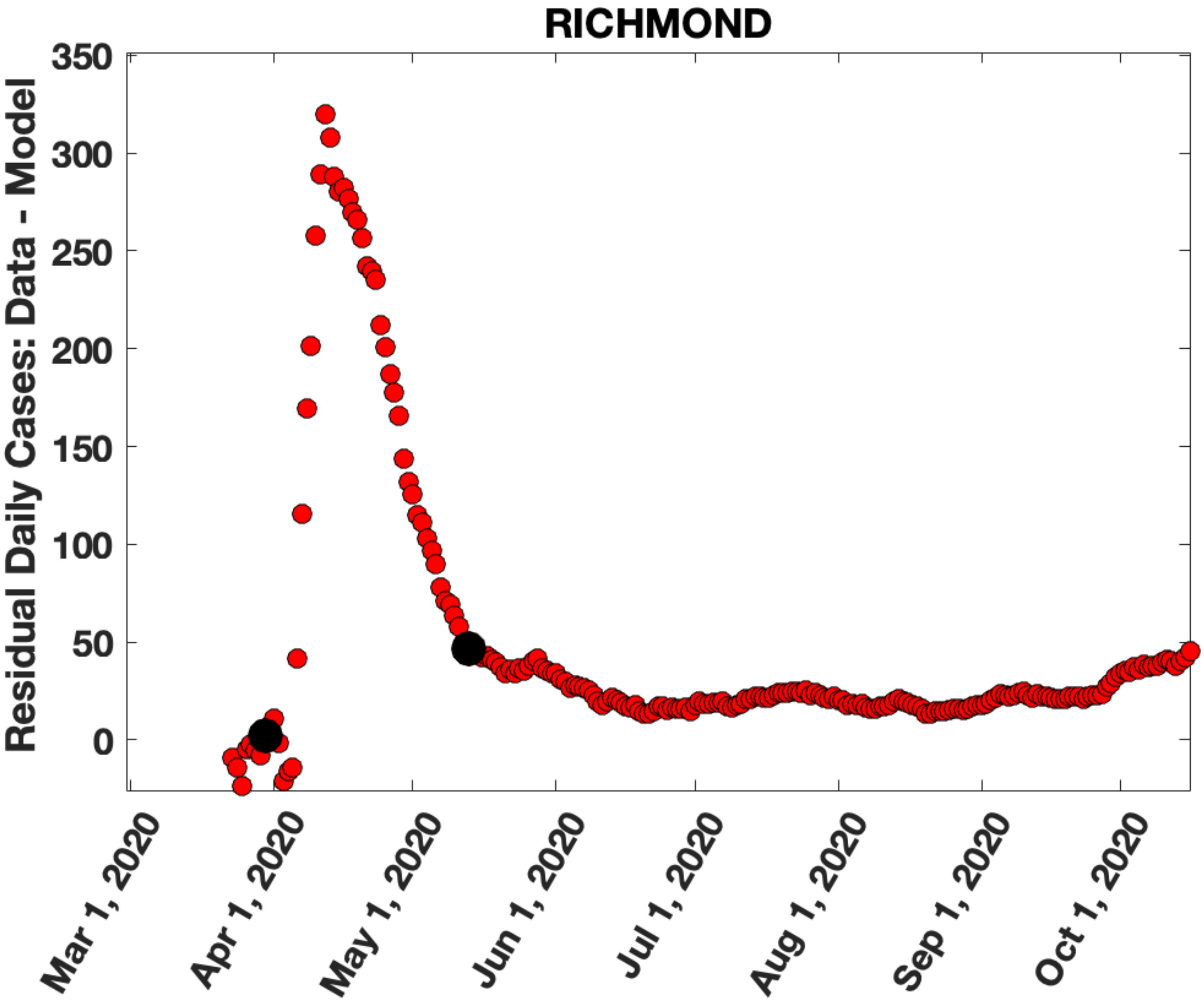

RICHMOND
Residual Daily Cases: Data - Model
350
300
250
200
150
100
50
0
Mar 1, 2020
Apr 1, 2020
May 1, 2020
Jun 1, 2020
Jul 1, 2020
Aug 1, 2020
Sep 1, 2020
Oct 1, 2020

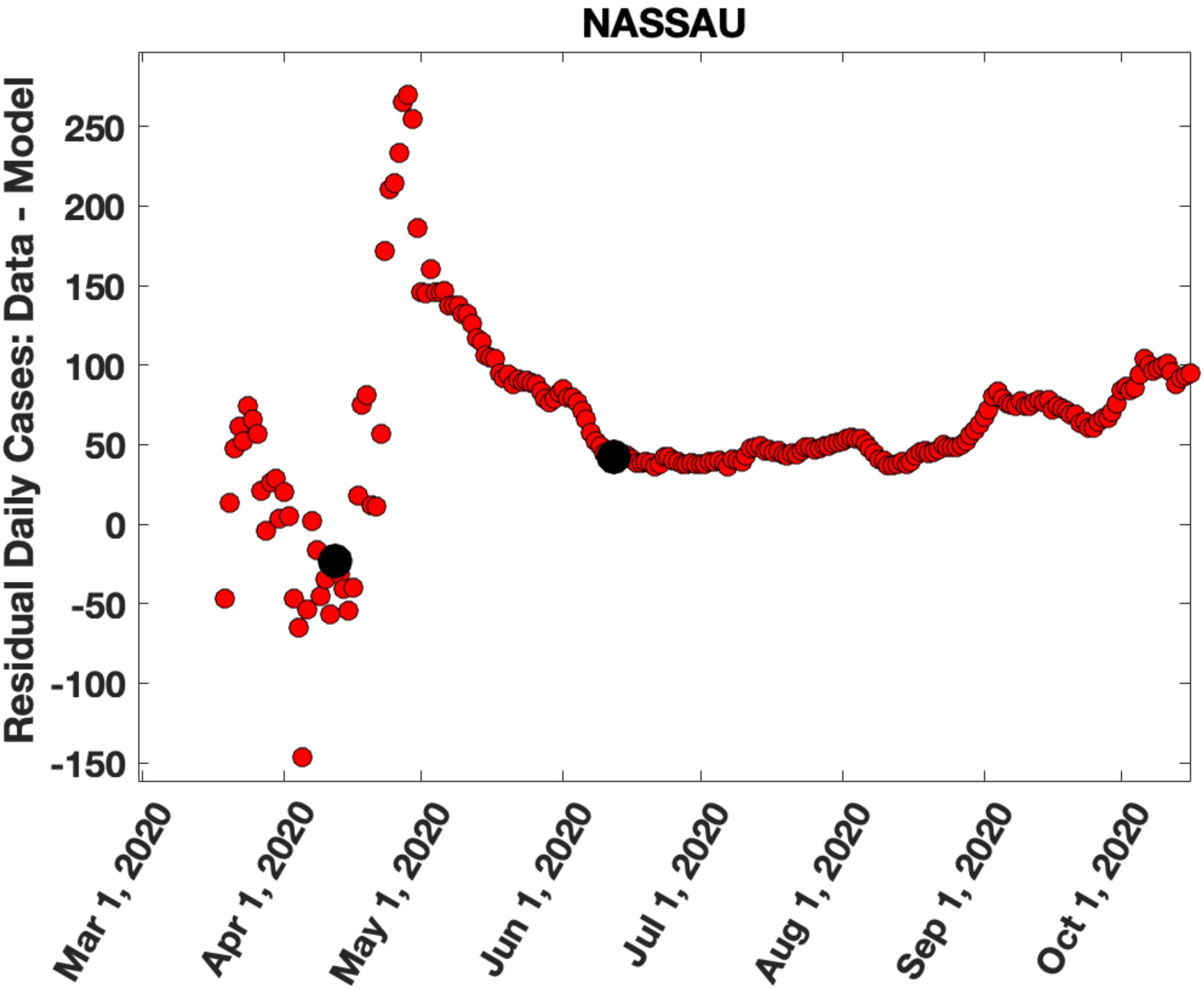
NASSAU
Residual Daily Cases: Data - Model
250
200
150
100
50
0
-50
-100
-150
Mar 1, 2020
Apr 1, 2020
May 1, 2020
Jun 1, 2020
Jul 1, 2020
Aug 1, 2020
Sep 1, 2020
Oct 1, 2020

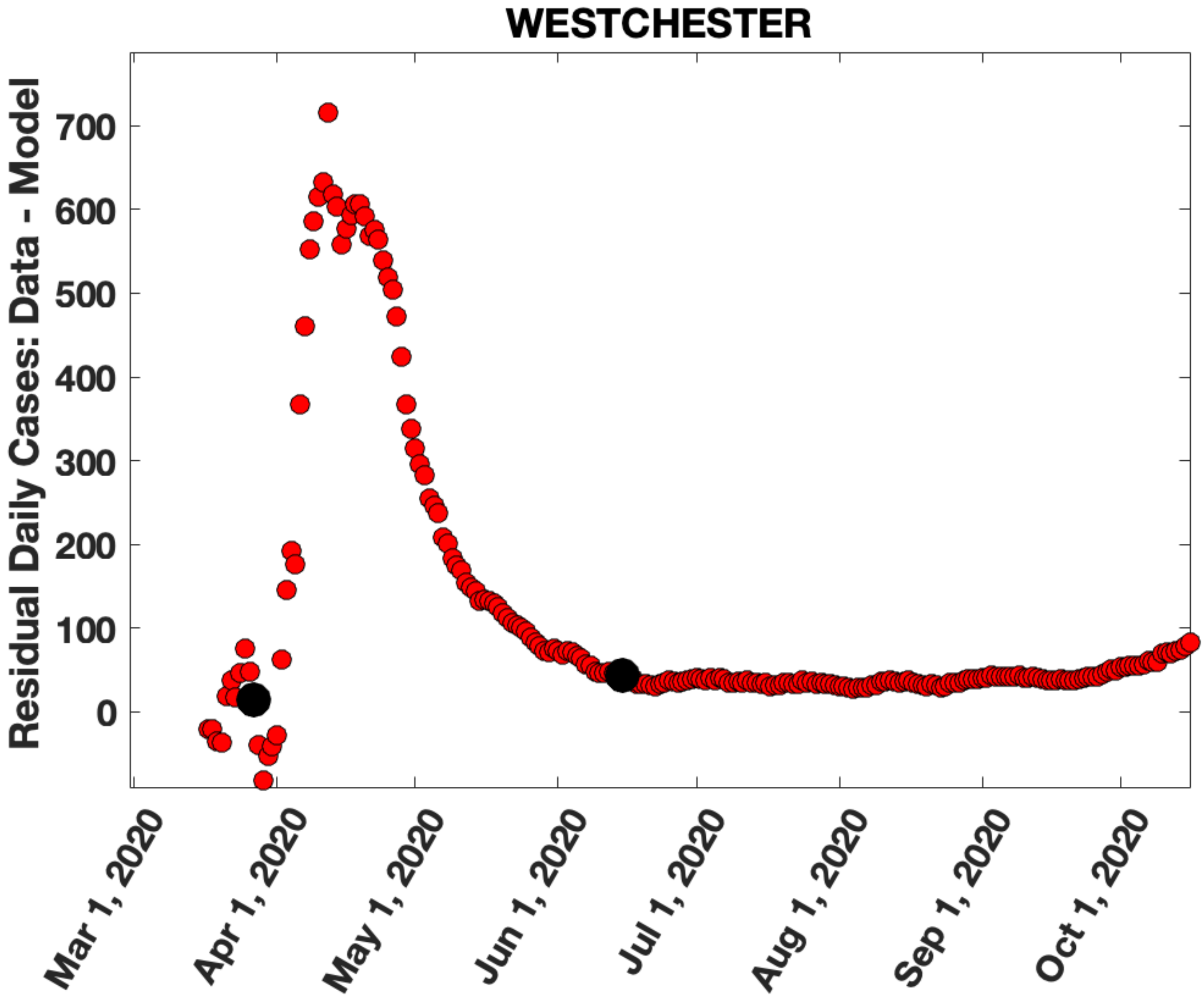

WESTCHESTER
Residual Daily Cases: Data - Model
700
600
500
400
300
200
100
0
Mar 1, 2020
Apr 1, 2020
May 1, 2020
Jun 1, 2020
Jul 1, 2020
Aug 1, 2020
Sep 1, 2020
Oct 1, 2020

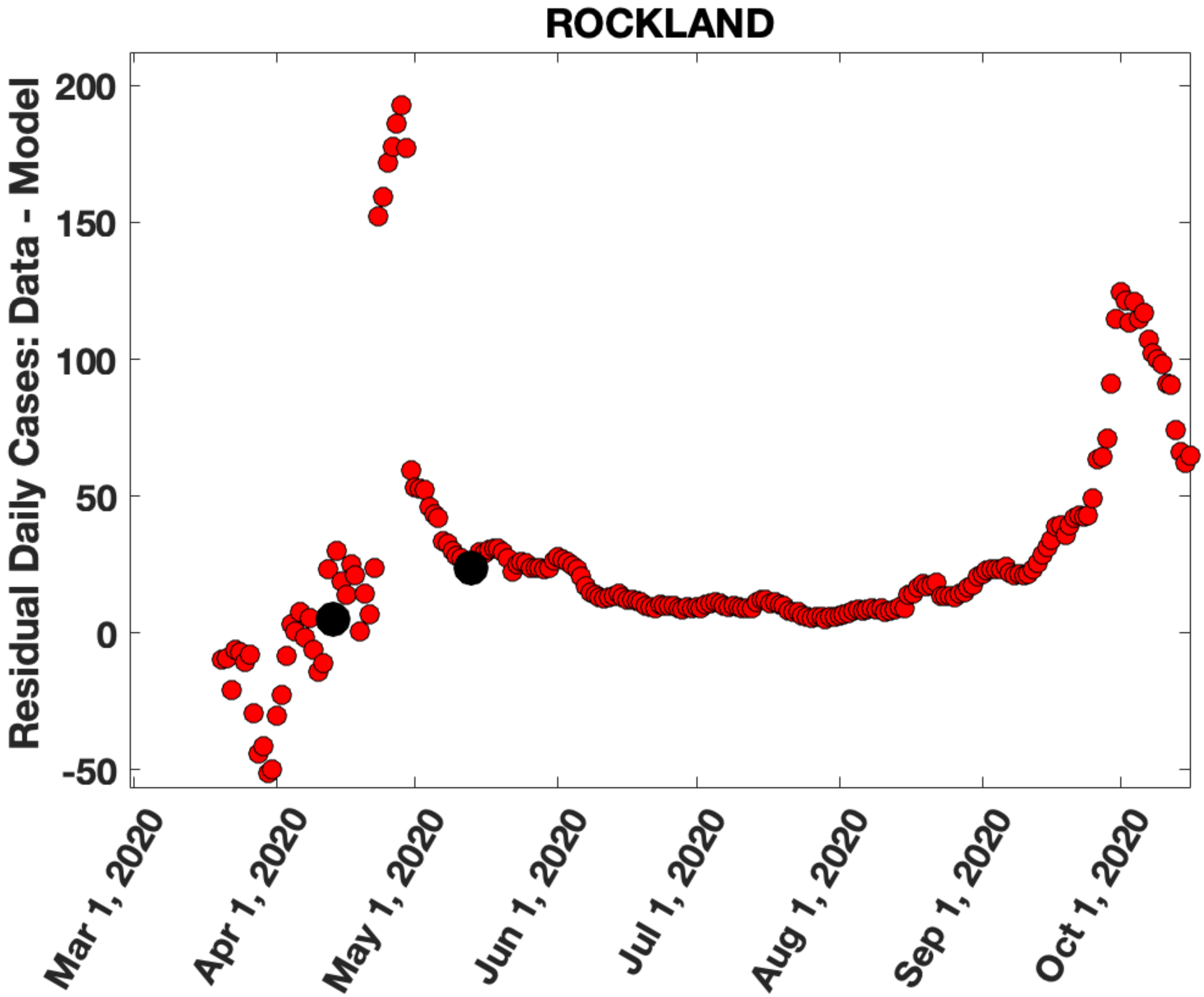
ROCKLAND
Residual Daily Cases: Data - Model
200
150
100
50
0
-50
Mar 1, 2020
Apr 1, 2020
May 1, 2020
Jun 1, 2020
Jul 1, 2020
Aug 1, 2020
Sep 1, 2020
Oct 1, 2020

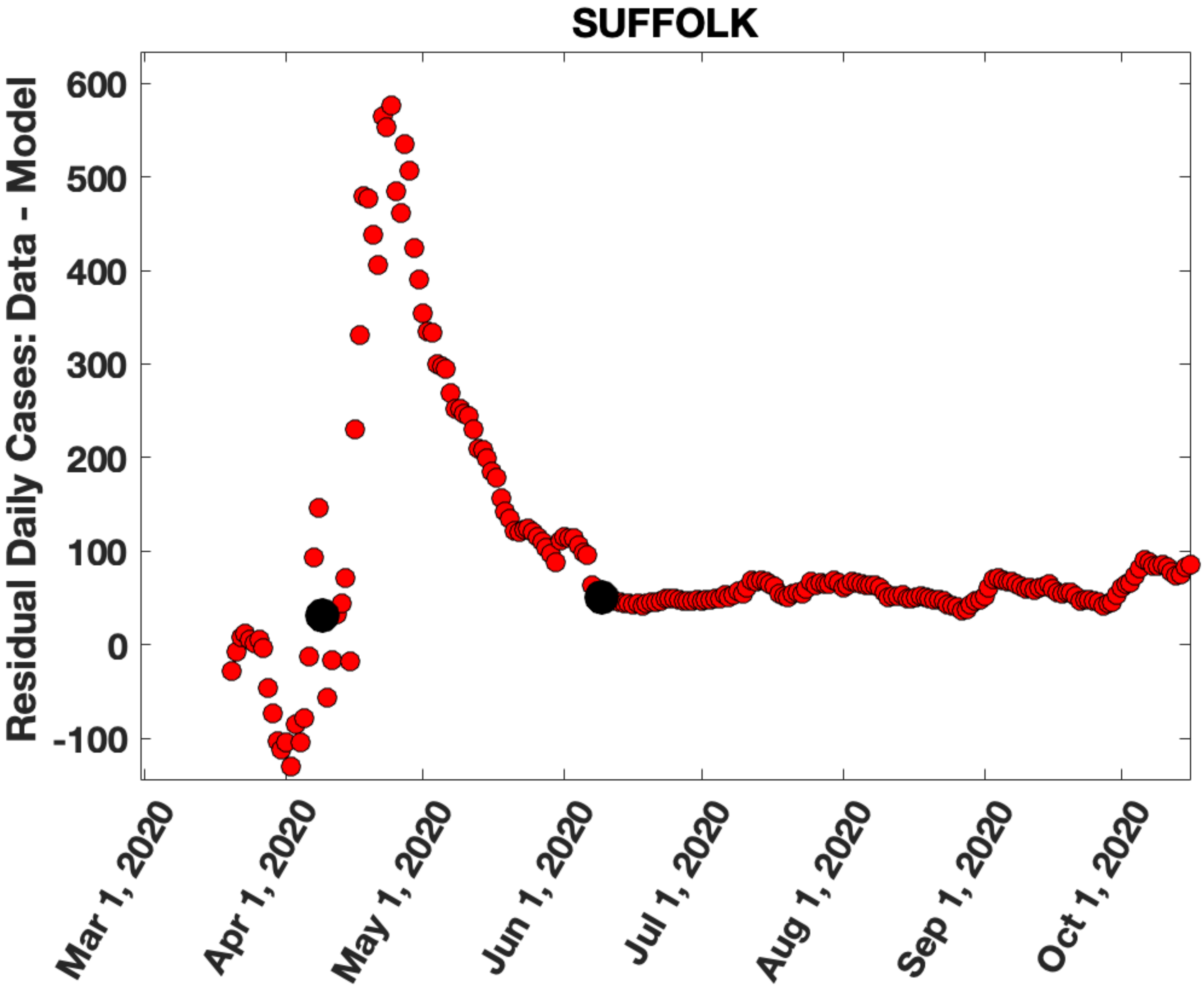
SUFFOLK
Residual Daily Cases: Data - Model
600
500
400
300
200
100
0
-100
Mar 1, 2020
Apr 1, 2020
May 1, 2020
Jun 1, 2020
Jul 1, 2020
Aug 1, 2020
Sep 1, 2020
Oct 1, 2020

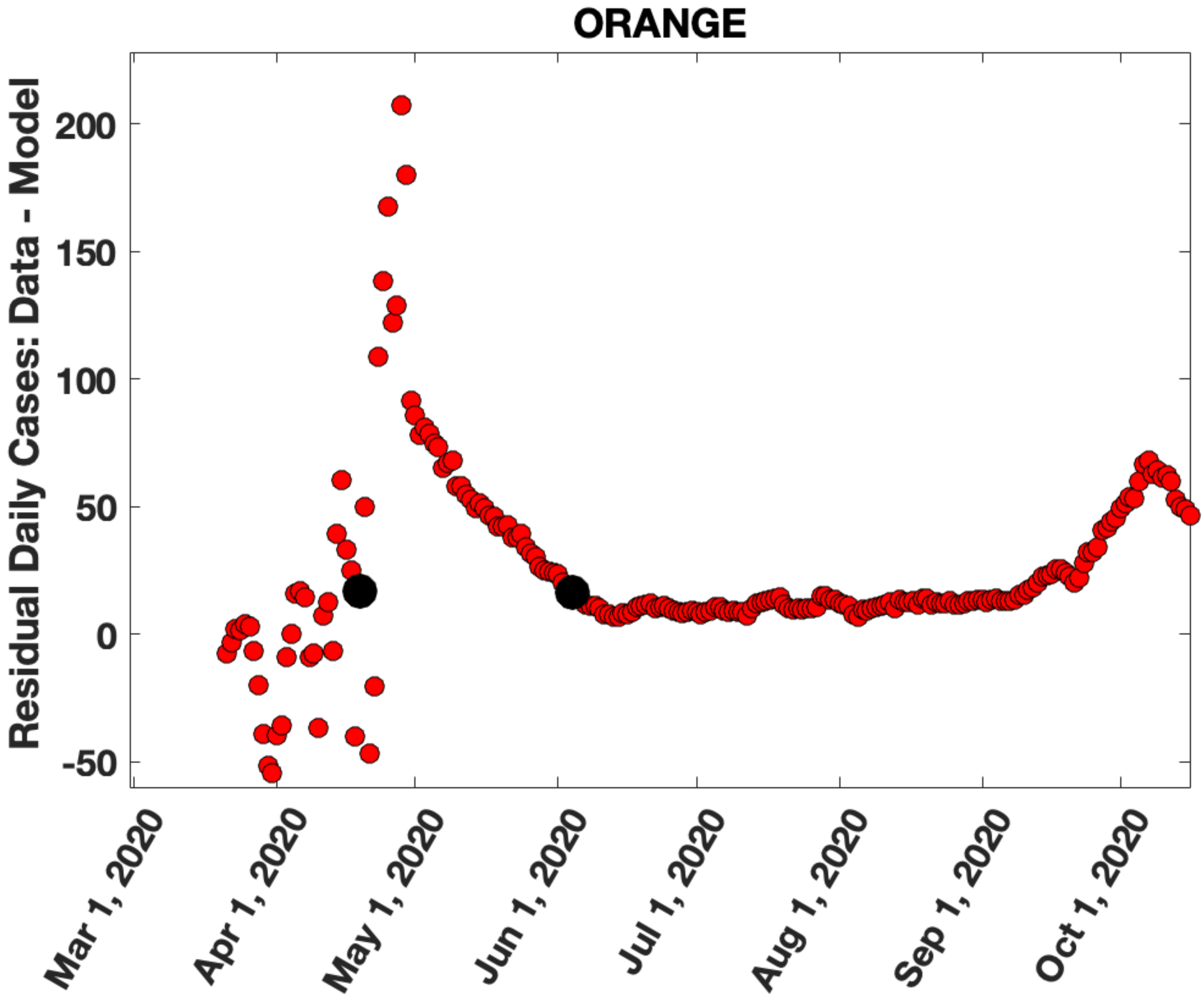

ORANGE
Residual Daily Cases: Data - Model
200
150
100
50
0
-50
Mar 1, 2020
Apr 1, 2020
May 1, 2020
Jun 1, 2020
Jul 1, 2020
Aug 1, 2020
Sep 1, 2020
Oct 1, 2020

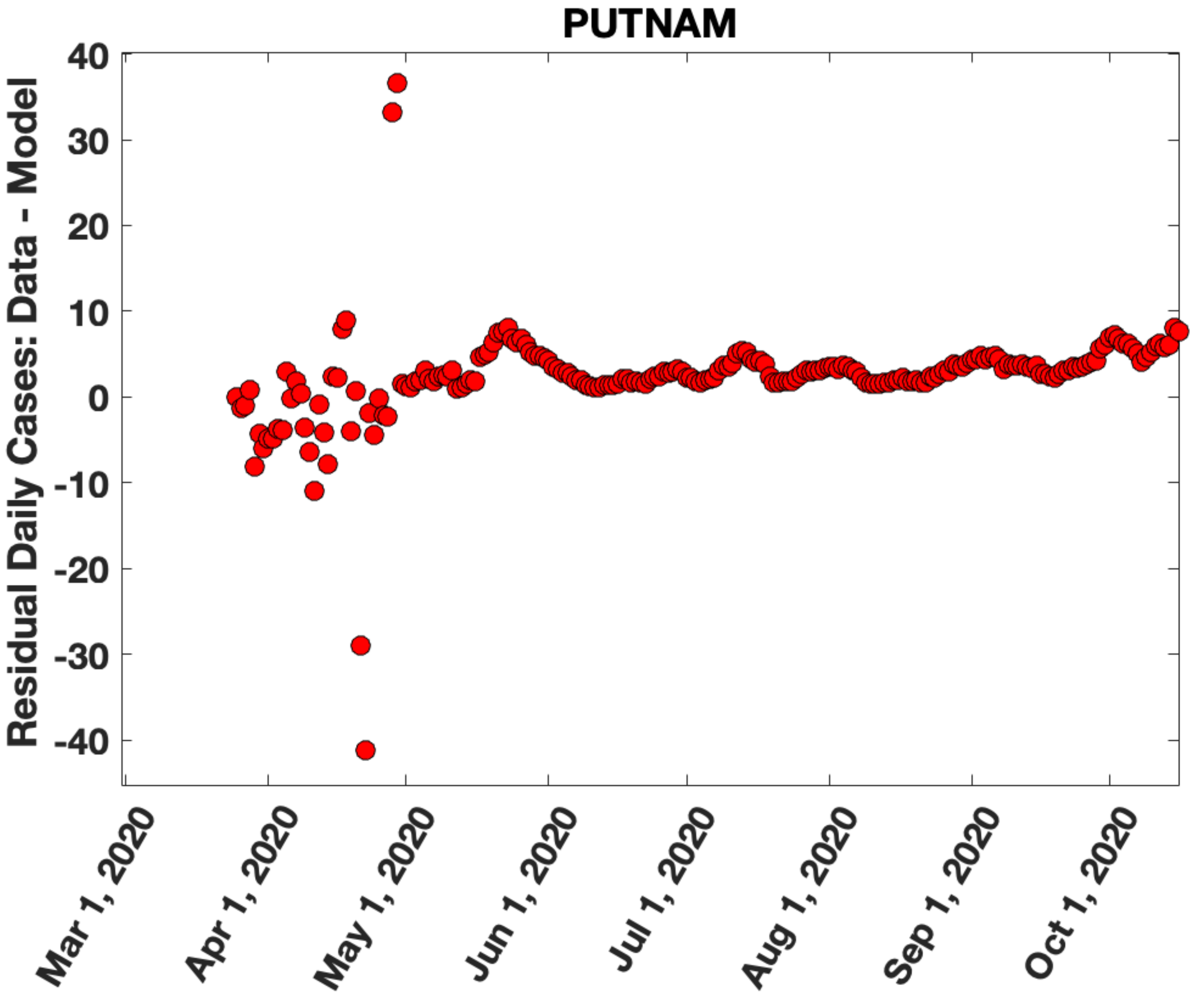
PUTNAM
Residual Daily Cases: Data - Model
40
30
20
10
0
-10
-20
-30
-40
Mar 1, 2020
Apr 1, 2020
May 1, 2020
Jun 1, 2020
Jul 1, 2020
Aug 1, 2020
Sep 1, 2020
Oct 1, 2020

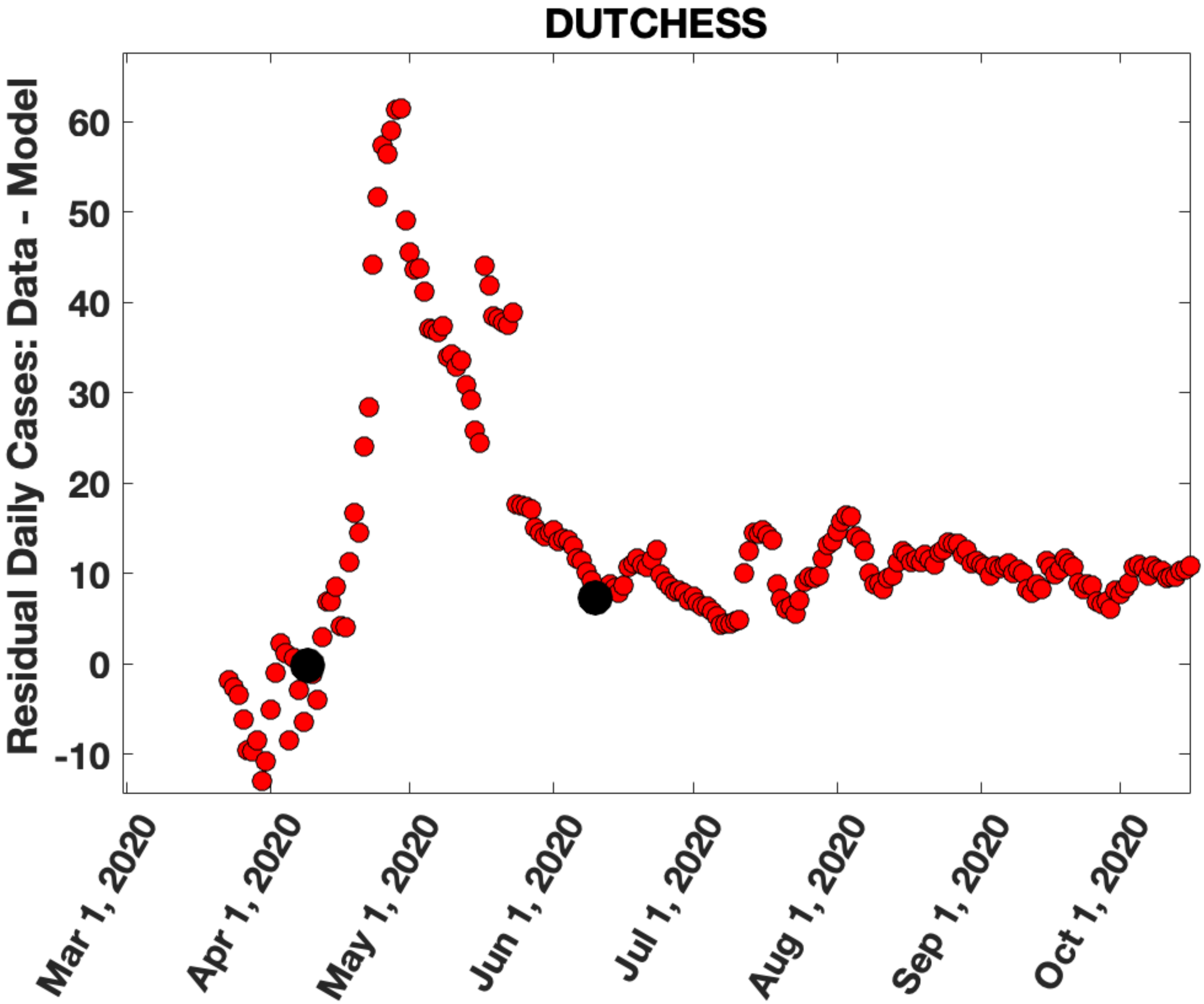

DUTCHESS
Residual Daily Cases: Data - Model
60
50
40
30
20
10
0
-10
Mar 1, 2020
Apr 1, 2020
May 1, 2020
Jun 1, 2020
Jul 1, 2020
Aug 1, 2020
Sep 1, 2020
Oct 1, 2020

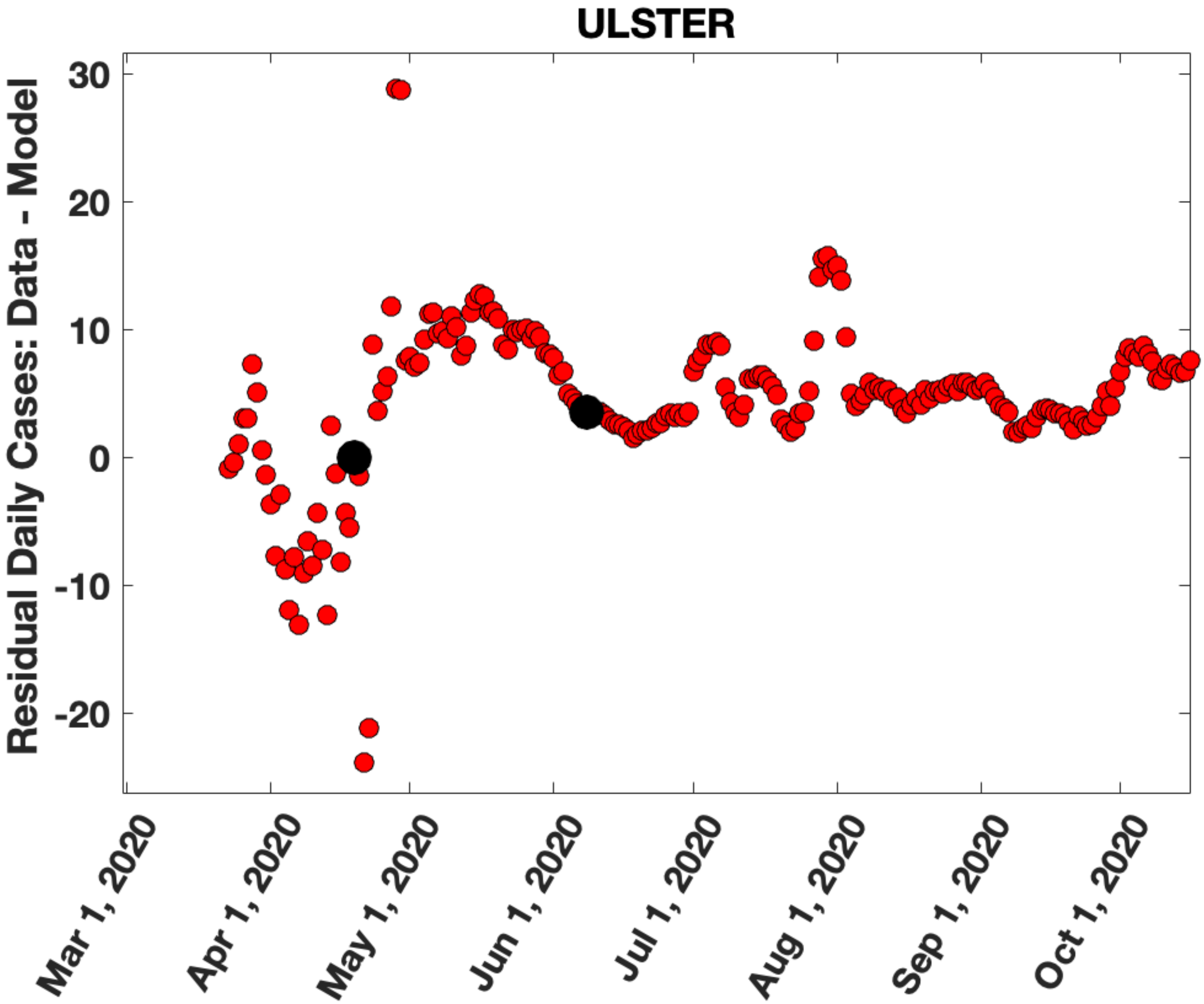
ULSTER
Residual Daily Cases: Data - Model
30
20
10
0
-10
-20
Mar 1, 2020
Apr 1, 2020
May 1, 2020
Jun 1, 2020
Jul 1, 2020
Aug 1, 2020
Sep 1, 2020
Oct 1, 2020

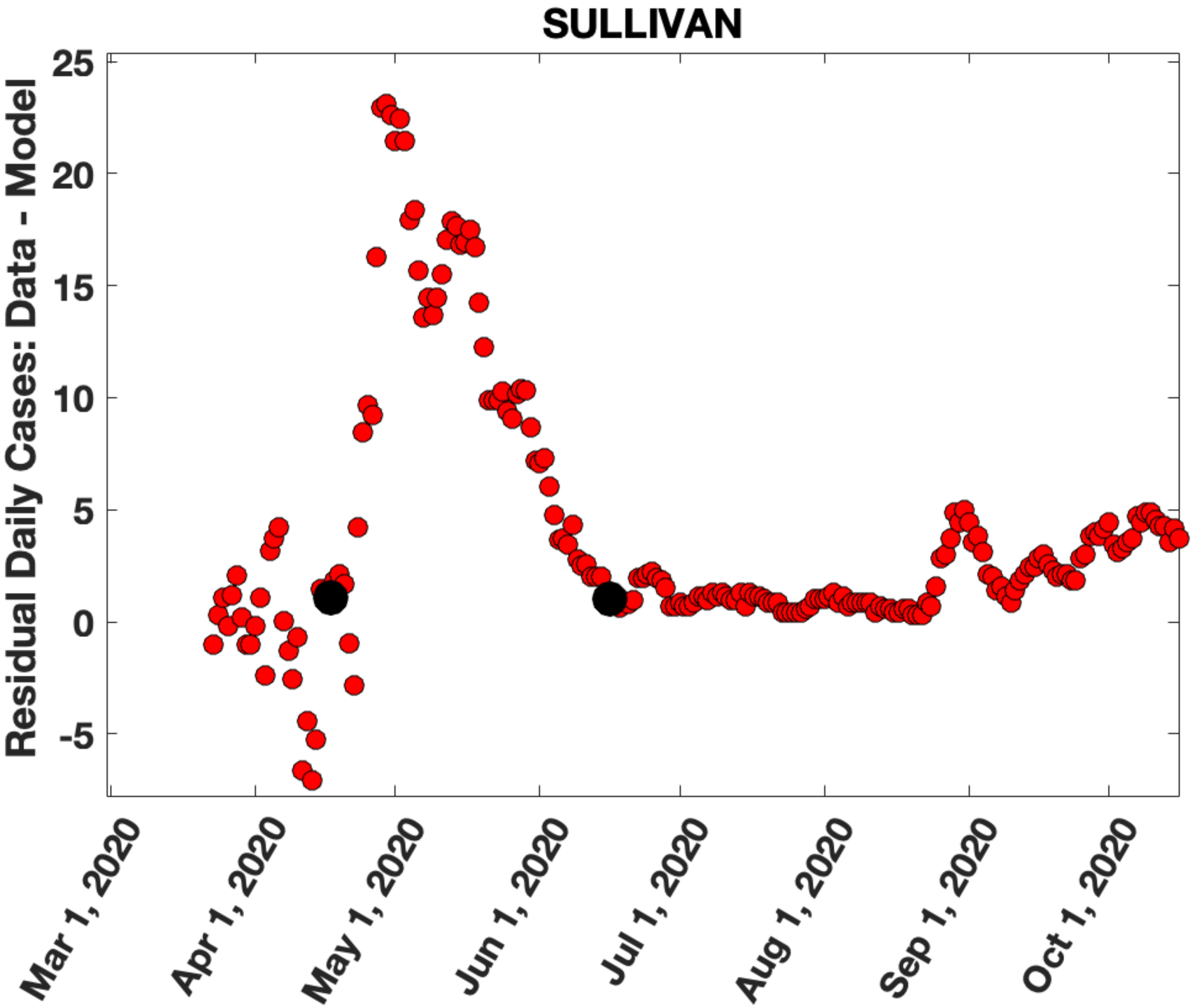
SULLIVAN
Residual Daily Cases: Data - Model
25
20
15
10
5
0
-5
Mar 1, 2020
Apr 1, 2020
May 1, 2020
Jun 1, 2020
Jul 1, 2020
Aug 1, 2020
Sep 1, 2020
Oct 1, 2020
